# Integrated multi-omics data analysis identifies a novel genetics-risk gene of *IRF4* associated with prognosis of oral cavity cancer

**DOI:** 10.1101/2021.11.17.21266500

**Authors:** Yan Lv, Yukuang Huang, Xuejun Xu, Zhiwei Wang, Yanfang Yu, Yunlong Ma, Mengjie Wu

## Abstract

**Background:** Oral cavity cancer (OCC) is one of the most common carcinoma diseases. Recent genome-wide association studies (GWAS) have reported numerous genetic variants associated with OCC susceptibility. However, the regulatory mechanisms of these genetic variants underlying OCC remain largely unclear.

**Objective:** This study aimed to identify OCC-related genetics risk genes contributing to the prognosis of OCC.

**Methods:** By combining GWAS summary statistics (N = 4,151) with expression quantitative trait loci (eQTL) across 49 different tissues from the GTEx database, we performed an integrative genomics analysis to uncover novel risk genes associated with OCC. By leveraging various computational methods based on multi-omics data, risk genes were prioritized as promising candidate genes for drug repurposing in OCC.

**Results:** Using two independent computational algorithms, we found that 14 risk genes whose genetics-modulated expressions showed a notable association with OCC. Among them, nine genes were newly identified, such as *IRF4* (P = 2.5×10^-9^ and P = 1.06×10^-4^)*, TNS3* (P = 1.44×10^-6^ and P = 4.45×10^-3^)*, ZFP90* (P = 2.37×10^-6^ and P = 2.93×10^-4^), and *DRD2* (P = 2.0×10^-5^ and P = 6.12×10^-3^).

These 14 genes were significantly overrepresented in several cancer-related terms, and 10 of 14 genes were enriched in 10 potential druggable gene categories. Based on differential gene expression analysis, the majority of these genes (71.43%) showed remarkable differential expressions between OCC patients and paracancerous controls. Integration of multi-omics-based evidence from genetics, eQTL, and gene expression, we identified that the novel risk gene of *IRF4* exhibited the highest ranked risk score for OCC. Survival analysis showed that dysregulation of *IRF4* expression was significantly associated with cancer patients outcomes (P = 8.1×10^-5^).

**Conclusions:** In summary, we prioritized 14 OCC-associated genes with nine novel risk genes, especially the *IRF4* gene, which provides a drug repurposing resource to develop therapeutic drugs for oral cancer.

## 1. Introduction

Oral cavity cancer (OCC) is one of the most common malignancy diseases [1, 2]. There are approximately 405,000 new OCC patients anticipated each year worldwide [3]. Although the rapid development of cancer diagnoses and therapies, the 5-year survival rate of OCC patients has remained at a dismal 50%[4]. Genetic components have been reported to largely influence OCC, especially variants located in alcohol-relevant genes, including alcohol dehydrogenase 7 (*ADH7*) and alcohol dehydrogenase 1B (*ADH1B*) [5, 6]. Hence, understanding the genetic underpinnings of OCC is of considerable interests, which may promote the advancement of individualized treatment strategies to maximize oncologic control and minimize the adverse influence of therapy.

With the development of high-throughput microarray and sequencing technologies, genome-wide association study (GWAS) as an effective method is widely applied to simultaneously examine the associations of millions of single nucleotide polymorphisms (SNPs) with complex diseases [7, 8]. The GWAS approach leveraged large-scale sample sizes to enhance the statistical power for discovering disease-associated risk SNPs and genes, such as the UK-Biobank project [9, 10] and the COVID-19 Host Genetics Initiative project [11–13]. These large GWAS consortia have identified hundreds of thousands of genetic variants associated with complex diseases [14]. In recent years, numerous GWASs have uncovered many genetic loci associated with OCC [15–19]. Due to GWAS applies a strict genome-wide significant threshold at P < 5×10^-8^, many genetic variants with small or moderate effect sizes were hard to be found in single GWAS analysis. This posed an intractable question that how to effectively pinpoint genuine variants from current existing GWAS summary statistics.

In addition, the vast majority of GWAS-reported SNPs were mapped within non-coding genomic regions[20, 21], indicating these variants potentially have *cis-/trans-*regulatory effects on modulating the expression level of a given gene rather than changing the function of its protein. Mounting genomics analyses [22–25] have been carried out to examine the regulatory mechanisms of GWAS-derived genetic variants, and determine whether GWAS-nominated genes whose aberrant alterations of expression contributing to disease pathogenesis due to genetic pleiotropy. For example, He et al. [22] reported a *Sherlock* integrative genomics tool based on Bayesian inference algorithm, which could incorporate genetic statistical values from GWAS summary data and expression quantitative trait loci (eQTL) data to systematically uncover the regulatory effects of genetic variants on gene expression for complex diseases [23, 26–28]. Recently, Barbeira and coworkers [29] introduced an efficient GWAS summary result-based extension statistical method termed S-MultiXcan, which could utilize the substantial sharing of eQTL across multiple tissues to enhance the capability to pinpoint potential target genes.

The primary goal of the current comprehensive genomics study is to identify genuine risk genes for OCC susceptibility and extend our understanding of genetic architecture implicated in the etiology of OCC. We first combined the converging effects of SNPs around a given gene, and then performed an integrative genomics analysis by incorporating GWAS summary statistics with eQTL to identify novel OCC-relevant genetic variants and susceptible genes. Moreover, by leveraging various computational methods based on multi-omics data, we prioritized risk genes as promising candidates for drug repurposing in OCC.

## 2. Materials and Methods

### 2.1. GWAS summary statistics on OCC

To yield genetic statistical information concerning OCC, we downloaded a GWAS summary statistics on oral cavity cancer from the MRC Integrative Epidemiology Unit (IEU) database (https://gwas.mrcieu.ac.uk/, accession ID: ieu-b-94) [16]. This resource, which is a manually curated collection of complete GWAS summary datasets on a brand of complex phenotypes, could be queried a database of the complete data. This dataset [16] comprised 1,223 OCC patients and 2,928 controls, part of the International Head and Neck Cancer Epidemiology Consortium (INHANCE). Written informed consent was signed for all subjects, and the ethical approval was approved by the respective institutional review boards. The PLINK 1.934 [30] was used to carry out systematic quality controls on genotype calling. After stringent quality control, a total of 7,510,833 common SNPs were included in the current analysis. More detailed clinical information was described in the original paper [16]. The web-access tool of LocusZoom (http://locuszoom.sph.umich.edu/) was used to visualize the regional genetic association signals for SNPs within risk genes. cis-eQTL and cis-sQTL analysis was conducted based on the GTEx online tool (https://gtexportal.org/home/).

### 2.2. Integrative genomics analysis of combining GWAS data with eQTL data

To identify risk genes whose genetically-regulated expression, we applied a comprehensive integrative genomics analysis to integrate GWAS summary statistics on OCC with expression quantitative trait loci (eQTL) data for 49 tissues from the GTEx database (vserion 8) [31]. First, based on the MASHR statistical method, we leveraged a linear regression model in S-PrediXcan to evaluate gene expression weights based on samples who having both genotypes and gene expression data. Through combining the variance and co-variance information of SNPs based on the reference panel of the 1,000 Genomes Project European Phase 3 [32], these estimated expression weights were assessed with a log odds ratio (OR) and standard errors from the GWAS summary data to calculate the levels of gene expression. For effective utilizing the information of significant genes whose expression has similar functions across different tissues, we further used the S-MultiXcan method to meta-analyze these MASHR results from S-PrediXcan to improve the statistical power. By integrating all available genes with convergent evidence across 49 GTEx tissues, the S-MultiXcan tool conducted a multivariate regression model for 22,331 genes, and FDR < 0.05 was considered to be of significance.

### 2.3. Gene-level genetic association analysis

To pinpoint risk genes associated with OCC, we conducted a gene-level genetic association analysis by incorporating the converging genetic signals around a given gene based on the Multi-marker Analysis of GenoMic Annotation (MAGMA) [7, 33]. We first extracted the SNP information including P values and chromosome position from GWAS summary statistics, and defined an extended genomic region for a specific gene containing the analyzed SNPs located within downstream or upstream 20kb of the gene. To calculate the linkage disequilibrium (LD) between SNPs, we used the 1,000 Genome Project Phase3 European Panel for reference [32]. To adjust the raw P values for multiple testing correction, we leveraged the widely-used method of Benjamini-Hochberg false discovery rate (FDR) [34], and FDR < 0.05 was considered to be of significance.

### 2.4. Pathway-based enrichment analysis

To understand the biological functions for these identified risk genes, we conducted a pathway-based enrichment analysis according to the authorized KEGG pathway resource [35] through utilizing the online-access tool of WEB-based Gene SeT AnaLysis Toolkit (WebGestalt; http://www.webgestalt.org) [36]. There were three well-documented methods used for enrichment analysis, including gene set enrichment analysis, network topology-based analysis, and over-representation analysis. By using the over-representation approach, we inputted these identified significant or suggestive gene lists for establishing the functional relationships of these genes with biological pathways. In addition, we also performed a gene-ontology (GO) enrichment analysis using the WebGestalt tool based on three widely-used functional categories: cellular component (CC), molecular function (MF), and biological process (BP). The number of genes in each pathway or GO-term were limited to a range between 5 and 2,000. The hypergeometric test was adopted to assess the significance level of each enrichment analysis, and the Benjamini-Hochberg FDR method was used for multiple testing correction.

### 2.5. Computer-based permutation analysis

To determine whether there exist prominently consistent results between genes from MAGMA analysis (N = 1,324, P < 0.05) and genes from S-MultiXcan analysis (N = 1,366, P < 0.05), we used a permutation method [11] to carry out a *in silico* permutation analysis with 100,000 times of random selections. In the first step, we calculated the overlapped genes between MAGMA and S-MultiXcan (***N*** *_observation_* = 472 genes). In the second step, we leveraged all examined genes from S-MultiXcan analysis as background genes (***N*** *_background_* =22,331 genes). By randomly selecting the same number of genes as those identified from S-MultiXcan analysis from background genes to overlap with genes identified from MAGMA for 100,000 times, we counted the number of overlapped genes in each random time (***N*** *_random_*). The empirically permuted P value = 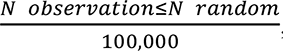 and P value < 0.05 is of significance.

### 2.6. Protein-protein interaction network analysis

To uncover the functional interaction patterns of these identified OCC-risk genes, we performed a protein-protein interaction (PPI) network analysis based on two extensively-used databases of STRING (v11.0, https://string-db.org/) [37] and GeneMANIA (http://www.gene-mania.org)[38]. There were 14 risk genes as an input list for searching interaction relationships based on current existing proteomics and genomics data, including genetic interactions, co-expression links, physical interactions, pathway links, and co-localizations. The Cytoscape platform [39] was adopted for visualizing the subnetworks.

### 2.7. Drug-gene interaction analysis

To find effective gene-targeted drugs for OCC, we submitted these 14 significant OCC-risk genes into the widely-applied Drug Gene Interaction Database (DGIdb v.3.0.2, http://www.dgidb.org/) based on 20 databases with 51 known interaction types. Based on the druggable categories in the DGIdb database, we attempted to search genes with potential drug abilities for these 14 OCC-risk genes. Furthermore, based on the GLAD4U database [40], we applied the hypergeometric test to conduct a drug-based enrichment analysis of these identified common genes between MAGMA analysis and S-MultiXcan analysis (Supplemental Table S5) for obtaining a drug repurposing resource.

### 2.9. Gene expression profiles between OCC and matched controls

To further validate the functionality of these 14 identified OCC-risk genes, we downloaded two independent RNA expression datasets from NCBI GEO database (Accession Nos. GSE160042 and GSE139869). The first analyzed dataset, GSE160042, contained 10 fresh human oral squamous cell carcinoma and their paired adjacent non-cancerous counterparts. All these enrolled patients received radical surgery without any form of pre-surgical adjuvant therapy. The RNA expression profiles were detected by the Agilent-045142 Human LncRNA v4 4X180K Microarray. The second RNA expression dataset (i.e., GSE139869) contained five paired tongue squamous cell carcinoma and paracancerous tissues, detected the transcriptomes with the use of the Agilent-062918 OE Human lncRNA Microarray. The paired Student’s t-test was used to calculate the significance between cancer patients and controls, and the *Corrplot* R package was used to visualizing the co-expression patterns among these 14 risk genes. Based on the expression dataset on head and neck squamous cell carcinoma (N = 518) from Cancer Genome Atlas (TCGA) database, we applied the GEPIA v2 online tool (http://gepia2.cancer-pku.cn/#survival) to perform survival analysis for identified genes and plot Kaplan-Meier curves.

## 3. Results

### 3.1. The workflow of current integrative genomics analysis

Current study is a two-stage designed integrative genomics analysis to identify novel risk

variants and genes for OCC in the discovery stage, and replicate these results in the validation stage (Figure 1). In the discovery stage, we collected a GWAS summary dataset on OCC with 1,223 patients and 2,928 matched controls [16], and collected eQTL data across 49 tissues from the GTEx database [31]. Among these eQTL datasets, RNA sequencing expression samples with < 10 million mapped reads were excluded, and normalized across samples using the *edgeR* R package [41]. There were 838 donors having RNA sequencing data and genotype data. Combining the GWAS summary data with eQTL data, we highlighted risk genes whose genetically-regulated abnormal expression associated with OCC. In the validation stage, we further conducted subsequent comprehensive computational analyses by using multiple bioinformatics tools for replication.

**Figure 1.**
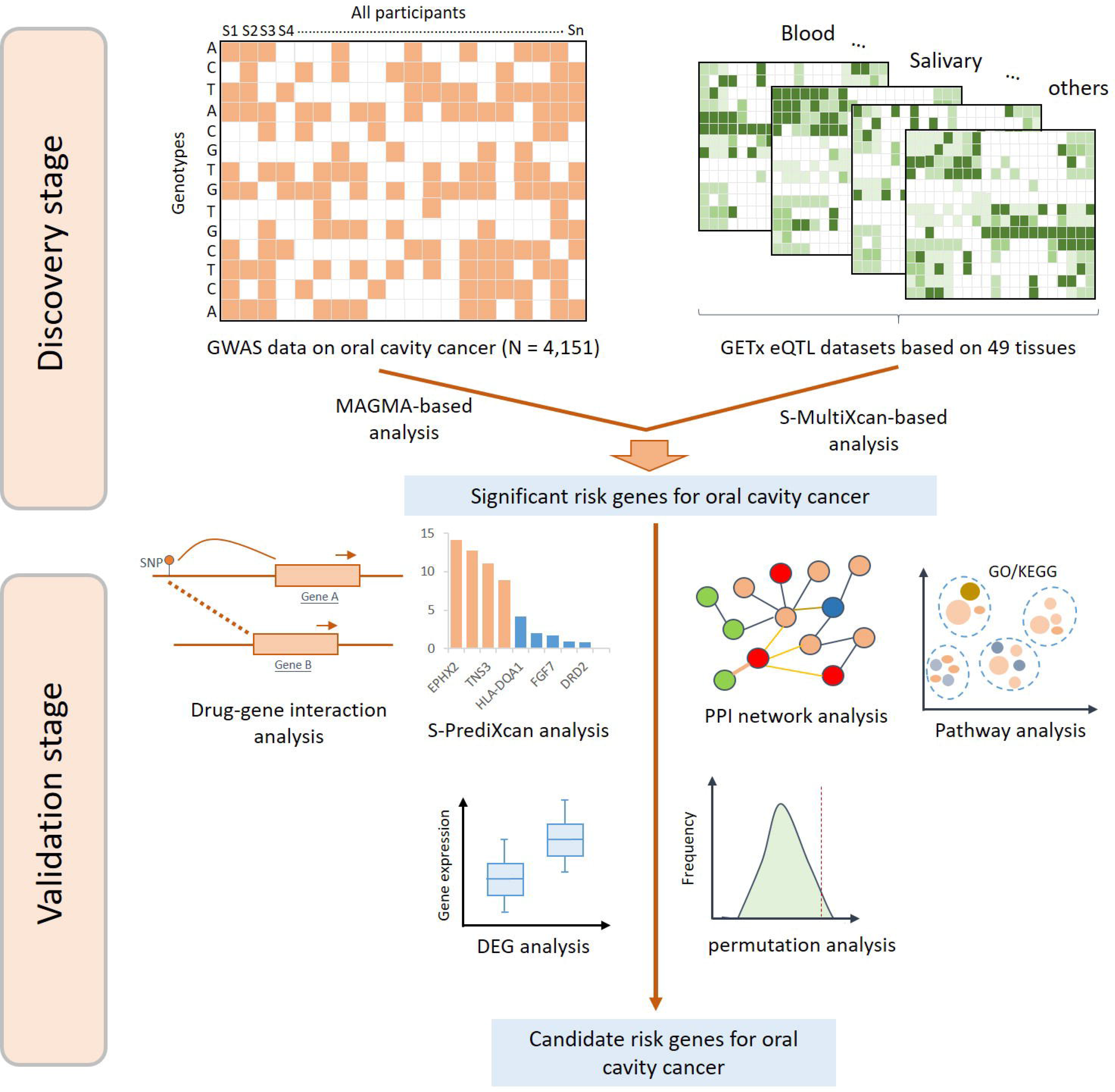
The whole process of the current integrative genomics analysis.

### 3.2. Gene-based genetic association analysis for OCC

In view of the original study [16] did not perform a gene-level genetic association analysis, we conducted a MAGMA gene-based association analysis to incorporate the converging effects of SNPs in a given gene. There were 14 genes showing notable associations with OCC susceptibility (FDR < 0.05, Figure 2 and Supplemental Table S1); including *IRF4* (P = 2.5×10^-9^), *TNS3* (P = 1.44×10^-6^), *NOTCH1* (P = 2.13×10^-6^), *ZFP90* (P = 2.37×10^-6^), *ATRN* (P = 3.28×10^-6^), *HLA-DQA1* (P = 6.45×10^-6^), *DRD2* (P = 2.0×10^-5^), *EPHX2* (P = 2.03×10^-5^), *CLPTM1L* (P = 2.13×10^-5^), *SCL6A2* (P = 2.76×10^-5^), *FGF7* (P = 2.78×10^-5^), *GTF2IRD1* (P = 3.15×10^-5^), *NLRP12* (P = 3.26×10^-5^), and *CDH16* (P = 3.46×10^-5^). Quantile-quantile plot exhibited a low-level genomic inflation (λ = 1.02) in the gene-based association analysis (Supplemental Figure S1). There were two loci of 5p15.33 and 9q34 mapped by significant genes of *CLPTM1L* and *NOTCH1* were consistent with the previous study [16].

**Figure 2.**
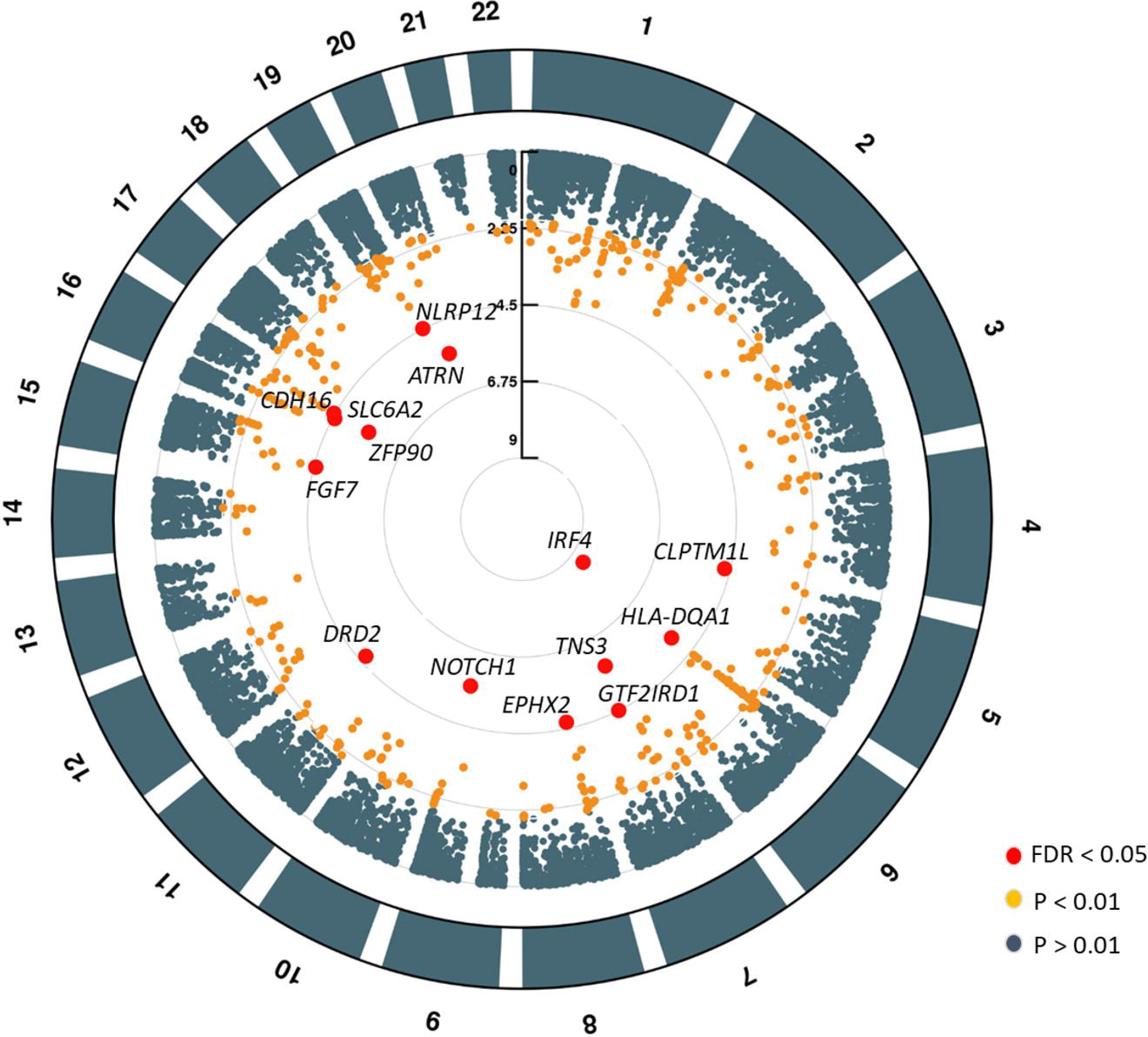
Results of gene-based association analysis using MAGMA tool. A) Circus plot showing the results of MAGMA-identified risk genes associated with OCC. Note: The outer ring shows the 22 autosomal human chromosomes (Chr1-22). A circular symbol in the inner ring stands for a specific gene. Color for each circular symbol represents the statistical significant level of a gene (red color represents FDR < 0.05 and orange color represent P < 0.01).

Among these 14 genes with multiple converging genetic association signals, there were 15 index SNPs showing notable associations with OCC (Supplemental Figures S2-S16), such as rs10775305 in *ZFP90*, rs6499103 in *CDH16*, rs4586205 in *DRD2*, rs10283378 in *EPHX2*, and rs12916839 in *FGF7*. For the most significantly MAGMA-identified gene of *IRF4*, the top lead SNP rs7773324 (P = 7.8×10^-7^) has high CADD score and regulomeDB score with strong activating chromatin state (Figure 3 and Supplemental Figure S17). In addition, both top lead SNPs of rs3125011 in *NOTCH1* (P = 7.9×10^-7^) and rs12916839 (P = 6.28×10^-7^) in *FGF7* have high CADD score and regulomeDB score with strong activating chromatin state (Supplemental Figures S18-S19). We observed that rs12916839 exhibited prominently allelic specific associations with *FGF7* among multiple tissues, and the T allele of rs1296839 showed down-regulated effects on the expression of *FGF7* (Supplemental Figure S20). Furthermore, the rs12916839 represents a splicing quantitative trait locus (sQTL) for *FGF7* (Supplemental Figures S21-S22).

**Figure 3.**
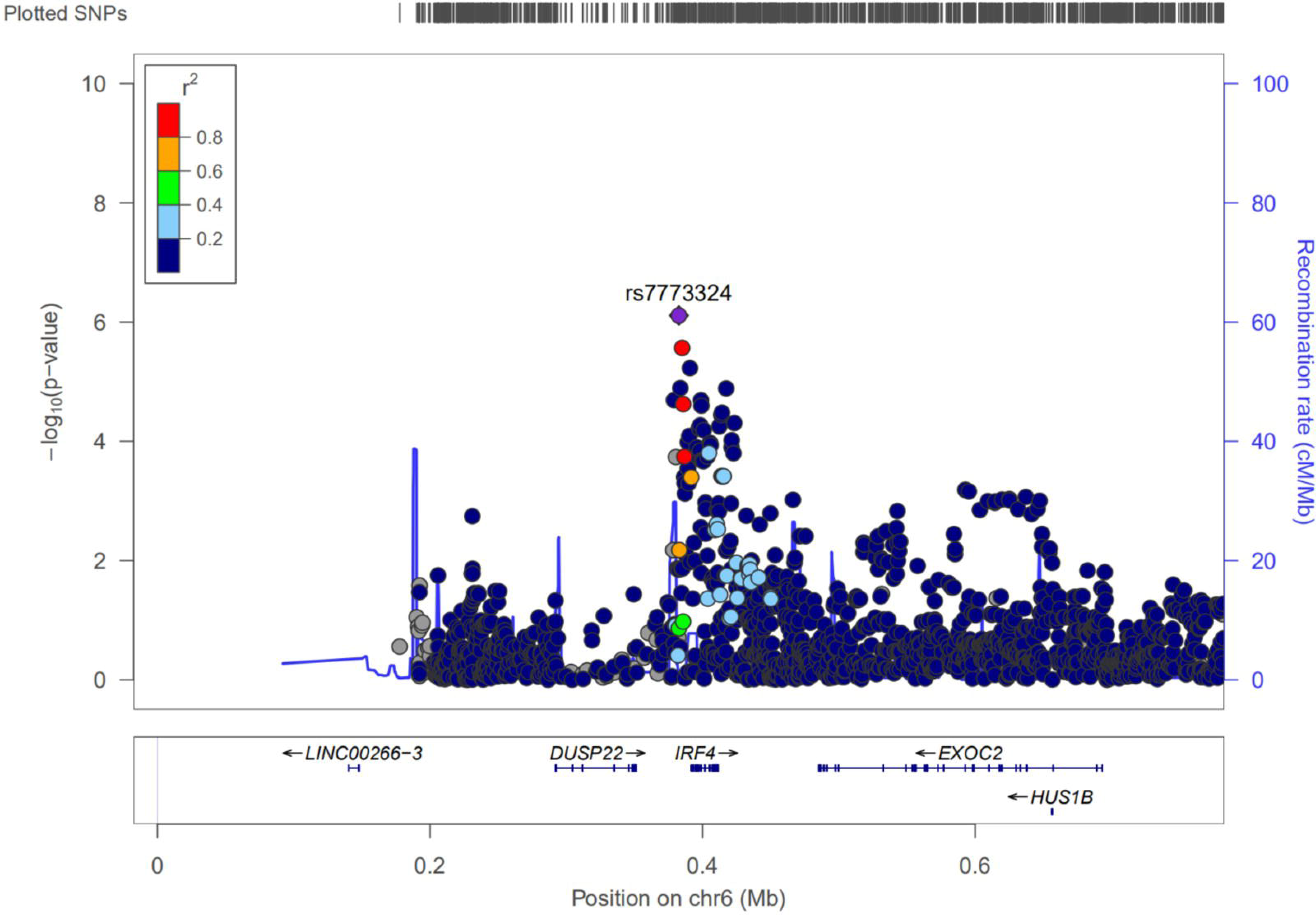
Regional plot for the top-ranked OCC-associated risk gene of *IRF4* identified from MAGMA analysis. The purple diamond marks the most strongly associated SNP of rs7773324 in each gene with OCC. The color illustrates LD information with the given SNP, as shown in the color legend.

Moreover, a number of 1,310 genes showed suggestive associations with OCC susceptibility from MAGMA analysis (P < 0.05, Supplemental Table S1). Among these suggestive genes, 191 genes have been documented to be implicated in other cancers in the GWAS Catalog database (Supplemental Table S1), and 11 genes of *HLA-DQB1*, *ZNF512*, *CCDC121*, *C2orf16*, *GPN1*, *GALNT14*, *AIF1, CTLA4, MICB, FAM175A,* and *ADH7* have been reported to be associated with OCC in previous studies (Supplemental Table S1). To examine their biological functions, we performed a pathway-based enrichment analysis of these suggestive genes, and found that six biological pathways were significantly associated with OCC (FDR < 0.05, Figure 4A-B, Supplemental Table S2). For example, staphylococcus aureus infection (P = 8.04×10^-6^), antigen processing and presentation (P = 9.18×10^-5^), graft-versus-host disease (P = 2.01×10^-4^), and cell adhesion molecules (P = 8.40×10^-4^). By performing a GO enrichment analysis, we found that there were 4 cellular component (CC) terms and 11 molecular function (MF) terms significantly enriched, including MHC protein complex, synaptic membrane, endonuclease complex, and GABA receptor activity (Figure 4C-D, and Supplemental Tables S3-S4). Overall, these results suggest that these observed genes may be a good resource for pinpointing causal risk genes for OCC.

**Figure 4.**
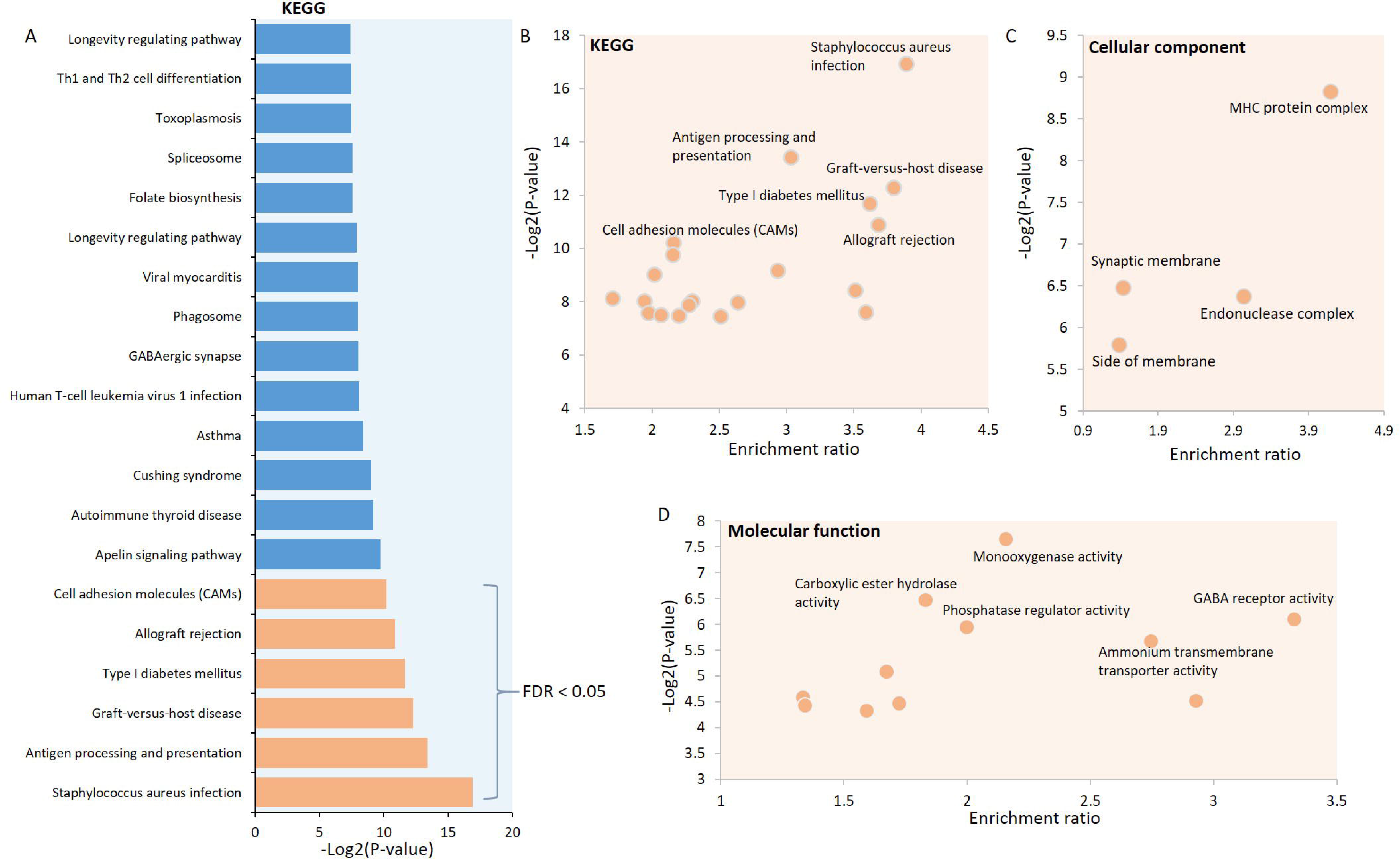
Functional enrichment analysis for MAGMA-identified significant or suggestive genes. A) Pathway enrichment analysis based on the KEGG pathway database for 1,324 suggestive OCC-risk genes (P < 0.05). Orange bar in the plot stands for significant enriched pathway with FDR < 0.05. B) Scatter plot showing the results of pathway enrichment analysis based on the KEGG pathway database. C) Scatter plot showing the results of GO-terms enrichment analysis on the category of cellular component. D) Scatter plot showing the results of GO-terms enrichment analysis on the category of molecular function.

### 3.3. Integrative genomics analysis identifies OCC-risk genes

By integrating GWAS summary statistics with eQTL data across 49 GTEx tissues using the S-MultiXcan method, we identified that 1,366 genes whose aberrant gene expressions were significantly or suggestively associated with OCC (P < 0.05, Supplemental Table S5). Such as *ATRN* (P = 5.9×10^-6^), *VWA7* (P = 1.64×10^-5^), *FGD4* (P = 3.75×10^-5^), *TERT* (P = 6.61×10^-5^), *HLA-DRB1* (P = 8.51×10^-5^), *CDH3* (P = 8.84×10^-5^), and *BTNL2* (P = 8.89×10^-5^). Among them, 33 genes have been published to be significantly associated with OCC, such as *HLA-DQA1*, *CLPTM1L*, *ZNF512*, *HLA-DQB1*, *GALNT14*, *CCDC121*, *CTLA4*, *GPN1*, *C2orf16*, *TENM3*, *LAMC3*, *ADH7*, *MICB*, *CCDC192*, *AIF1*, *RERGL*, *OR52N2*, *METAP1*, *WDFY3*, *ALDH2*, *MUC21*, *IL1A*, *KCNC4*, *STK31*, *BTBD11*, and *ADCY2* [6, 15, 16, 42]. By comparing these identified genes between S-MultiXcan and MAGMA, we observed that 472 genes have shared significant evidence between two methods (Figure 5A and Supplemental Table S5). Based on 100,000 times of *in silico* permutation analysis, we found that there existed a prominent concordance of results from between S-MultiXcan and MAGMA analysis (Empirical P < 1.0×10^-5^, Figure 5B), suggesting that these findings were relatively reliable and could be used for subsequent analyses.

**Figure 5.**
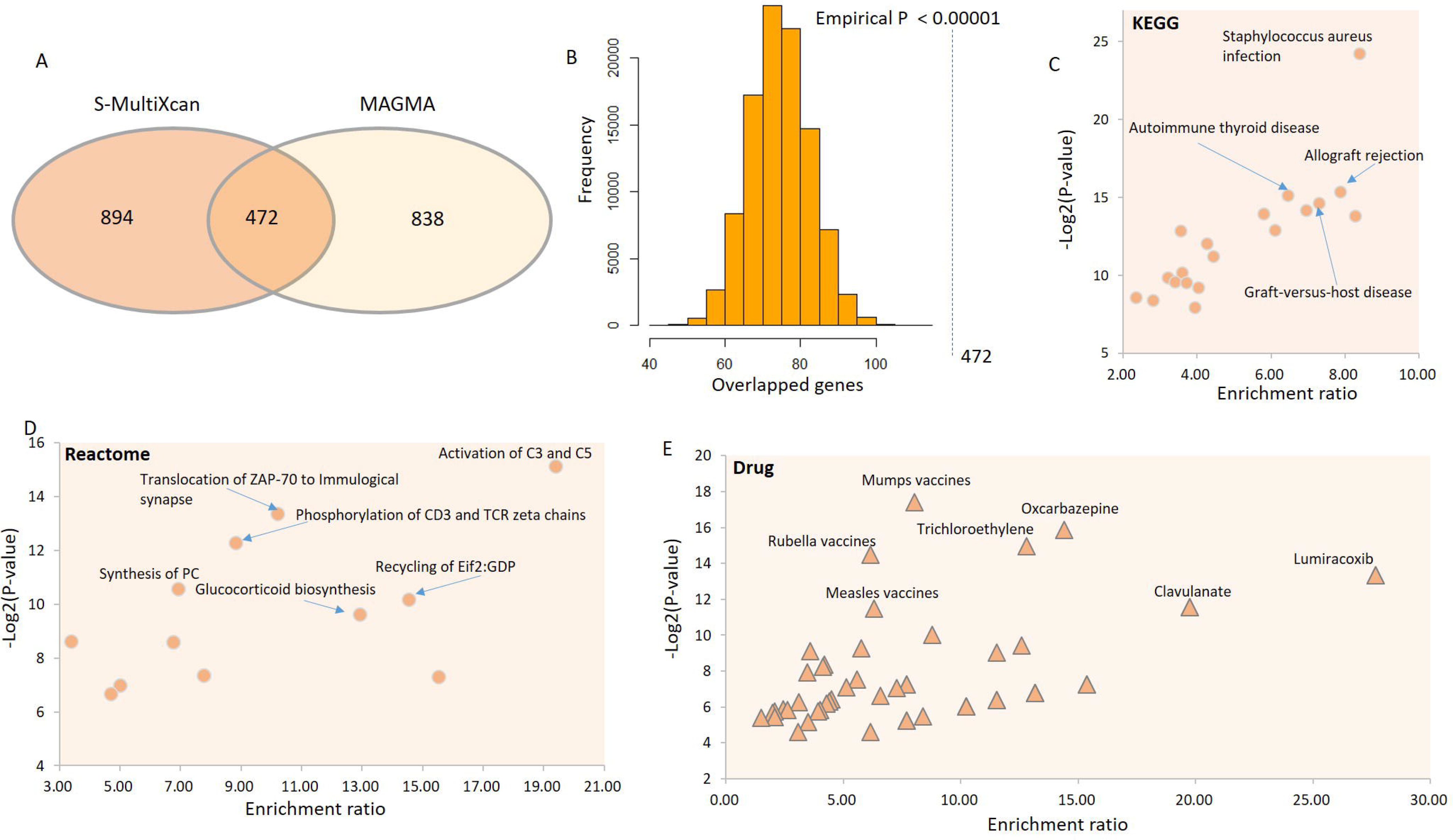
Consistent results between MAGMA analysis and S-MultiXcan analysis. A) Common genes identified from MAGMA analysis and S-MultiXcan analysis. There were 472 genes in common between independent methods. B) *In silico* permutation analysis for the results between MAGMA and S-MultiXcan analysis. There were 100,000 times random selections for this analysis, and empirically permuted P value < 0.00001. C) Scatter plot showing the results of pathway enrichment analysis based on the KEGG pathway database for these 472 common genes. D) Scatter plot showing the results of pathway enrichment analysis based on the Reactome pathway database for these 472 common genes. E) Scatter plot showing the results of drug-term enrichment analysis based on the GLAD4U drug database.

Furthermore, we performed two pathway enrichment analyses for these 472 common genes based on two independent pathways resources of KEGG and Reactome. We observed that 18 biological pathways were significantly overrepresented (FDR < 0.05, Figure 5C-D and Supplemental Tables S6-S7). For example, staphylococcus aureus infection (P = 5.09×10^-8^), allograft rejection (P = 2.35×10^-5^), autoimmune thyroid disease (P = 2.79×10^-5^), graft-versus-host disease (P = 3.94×10^-5^), type I diabetes mellitus (P = 5.43×10^-5^), and activation of C3 and C5 (P = 2.78×10^-5^), reminiscing that these pathways showed significant enrichments in aforementioned pathway analysis.

To explore the druggable actions of these 472 common genes, we carried out a drug-based enrichment analysis based on the GLAD4U database. We found that seven drug-terms were significantly enriched (FDR < 0.05, Figure 5E and Supplemental Table S8), including mumps vaccines (P = 5.73×10^-6^), oxcarbazepine (P = 1.67×10^-5^), trichloroethylene (P = 3.16 ×10^-5^), rubella vaccines (P = 4.32×10^-5^), desflurane (P = 9.79×10^-5^), and lumiracoxib (P = 9.79×10^-5^). In addition, there were 45 suggestively-enriched drug-terms by these OCC-related common genes (P < 0.05, Supplemental Table S8). These observed drug terms and risk genes may provide a repurposing resource for searching drug targets for OCC.

### 3.4. Prioritization of OCC-risk genes

Among the 14 MAGMA-identified significant genes (FDR < 0.05), 13 genes (13/14=92.86%) were significantly replicated by using S-MultiXcan integrative analysis (Figure 6A). There exist a moderate correlation of these top-ranked significant genes from two independent tools (r = 0.386, Figure 6B). Five genes of *HLA-DQA1, NOTCH1, FGF7, SLC6A2*, and *CLPTM1L* have been reported to be associated with oral and pharynx cancer [15–19]. By performing the S-PrediXcan integrative analysis, we found that several genes showed significant associations with oral cancer for each specific tissue (Supplemental Figures S23-S26). By using the DisGeNET algorithm [43] in the Metascape tool [44], these 14 risk genes were significantly enriched in cancer-related categories, including malignant carcinoid syndrome, small lymphocytic lymphoma, basal cell neoplasm, and basal cell cancer (FDR < 0.05, Supplemental Figure S27 and Table S9).

**Figure 6.**
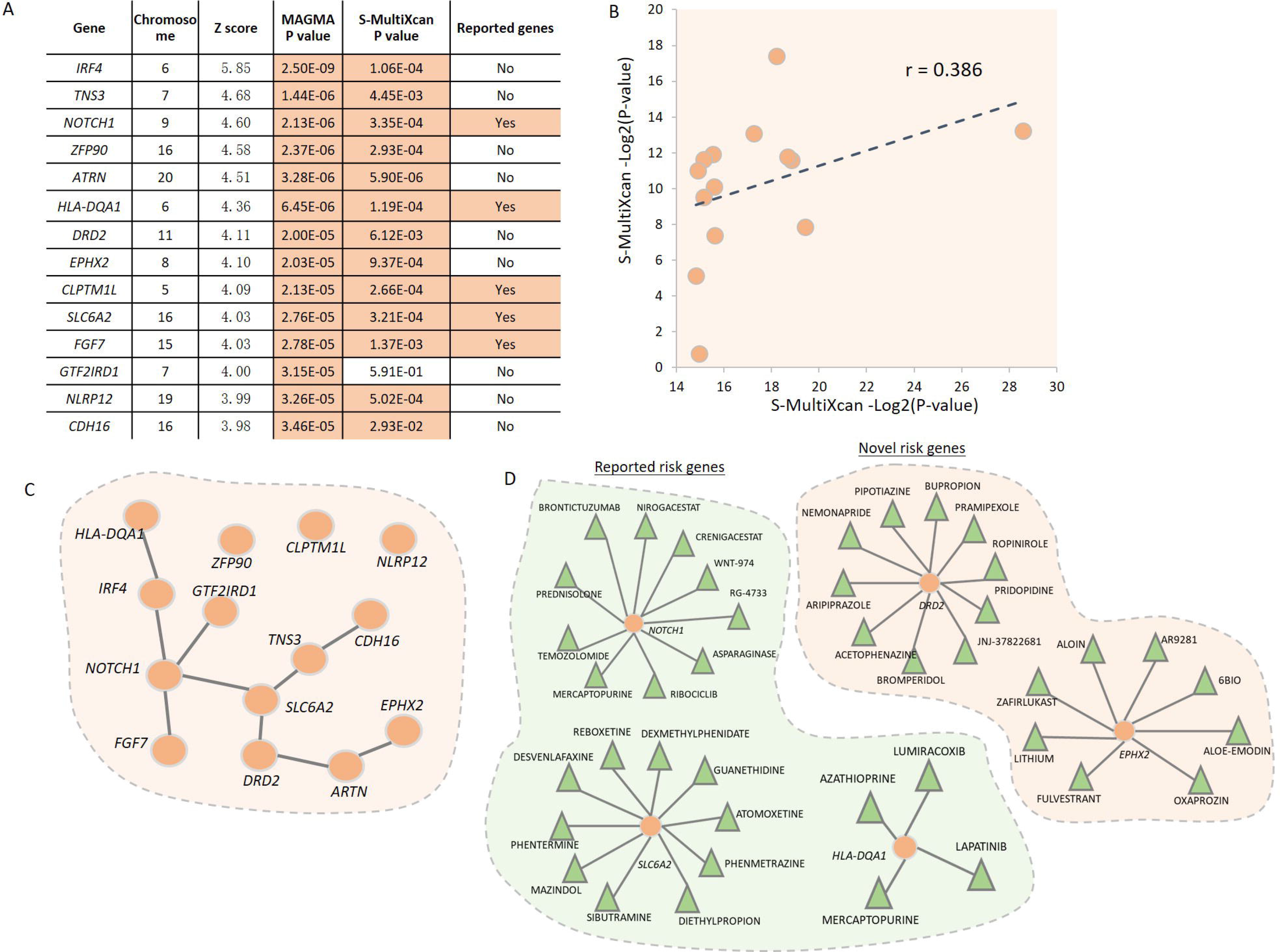
Identification of 14 risk genes associated with OCC susceptibility. A) Summary of these 14 significant OCC-associated genes identified from MAGMA and S-MultiXcan analysis. B) Scatter plot showing the correlation of results on these 14 risk genes from MAGMA and S-MultiXcan analysis. C) Protein-protein interaction network analysis of these 14 genes. D) Plot showing the results of the drug-gene interactions on five druggable genes. The orange circular symbol represents a given gene (i.e., *NOTCH1, HLA-DQA1, DRD2, SLC6A2*, and *EPHX2*). The green triangle represents a specific drug that is targeted with each gene.

To explore whether there exist collective functions of these 14 genes on OCC, we conducted a PPI network enrichment analysis based on two databases of STRING and GeneMAINA, and these genes were remarkably enriched in a functional subnetwork (P < 0.05, Figure 6C), suggesting that these risks have highly biological interactions implicated in oral cancer. Compared with patients with non-oral squamous cell carcinoma or other squamous cell carcinomas, mutations in *NOTCH1* have been frequently identified in patients with oral squamous cell carcinomas [17]. The genes of *IRF4*, *GTF2IRD1*, *FGF7*, and *SLC6A2* showed functional connections with *NOTCH1*. An earlier study [45] showed that there existed a genetic interaction of *NOTCH1* with *SLC6A2* and *GTF2IRD1*. By performing an interlaboratory comparability study, Dobbin et al. [46] reported that the cancer-related gene of *ARTN* was highly co-expressed with *EPHX2*.

Based on a drug-gene interaction analysis, we observed that 10 of 14 OCC-associated genes (71.43%) were overrepresented in 10 potential “druggable” gene categories, such as druggable genome, cell surface, drug resistance, transcription factor, growth factor, and G-protein coupled receptor (Supplemental Figure S28). There were five genes, namely *NOTCH1*, *SLC6A2*, *DRD2*, *EPHX2*, and *HLA-DQA1*, found to be targeted at least four drugs (Figure 6D). For example, the gene of *NOTCH1* is targeted by 10 drugs, including brontictuzumab, nirogacestat, crenigacestat, wnt-974, rg-4733, asparaginase, ribociclib, mercaptopurine, temozolomide, and prednisolone. *DRD2* gene shows molecular interactions with 10 drugs, including nemonapride, pipotiazine, bupropion, pramipexole, ropinirole, pridopidine, bromperidol, acetophenazine, and aripiprazole. These results suggest that these 14 risk genes predisposed to have collective functions on OCC.

### 3.5. Differential expression of 14 risk genes between OCC patients and control samples

By using the expression dataset of GSE139869, we conducted a DGE analysis of these 14 risk genes. Three genes not expressed in this dataset. We found that six of 11 genes were prominently expressed between OCC patients and paracancerous controls (Figure 7A). *ATRN* (P = 0.027), *CLPTM1L* (P = 0.007), *IFR4* (P = 0.0054), and *TNS3* (P = 7.6×10^-4^) showed significantly higher expressions among OCC patients than that among controls, and *EPHX2* (P = 4.0×10^-5^) and *GTF2IRD1* (P = 0.017) exhibited significantly lower expressions among OCC patients than that among controls (P < 0.05, Figure 7A). Consistently, by analyzing the expression dataset of GSE160042, we observed that 10 of 14 genes were significantly differentially expressed between OCC patients and matched controls (Figure 7B). *ATRN* (P = 0.014), *CLPTM1L* (P = 8.7×10^-5^), *HLA-DQA1* (P = 0.003), *IRF4* (P = 4.6×10^-4^), *TNS3* (P = 1.2×10^-6^), and *ZFP90* (P = 0.043) were prominently highly expressed in OCC patients, and *CDH16* (P = 9.6×10^-5^), *EPHX2* (P = 2.1×10^-5^), *GTF2IRD1* (P = 3.3×10^-4^), *NOTCH1* (P = 4.6×10^-4^) were remarkably down-expressed in OCC patients (Figure 7B).

**Figure 7.**
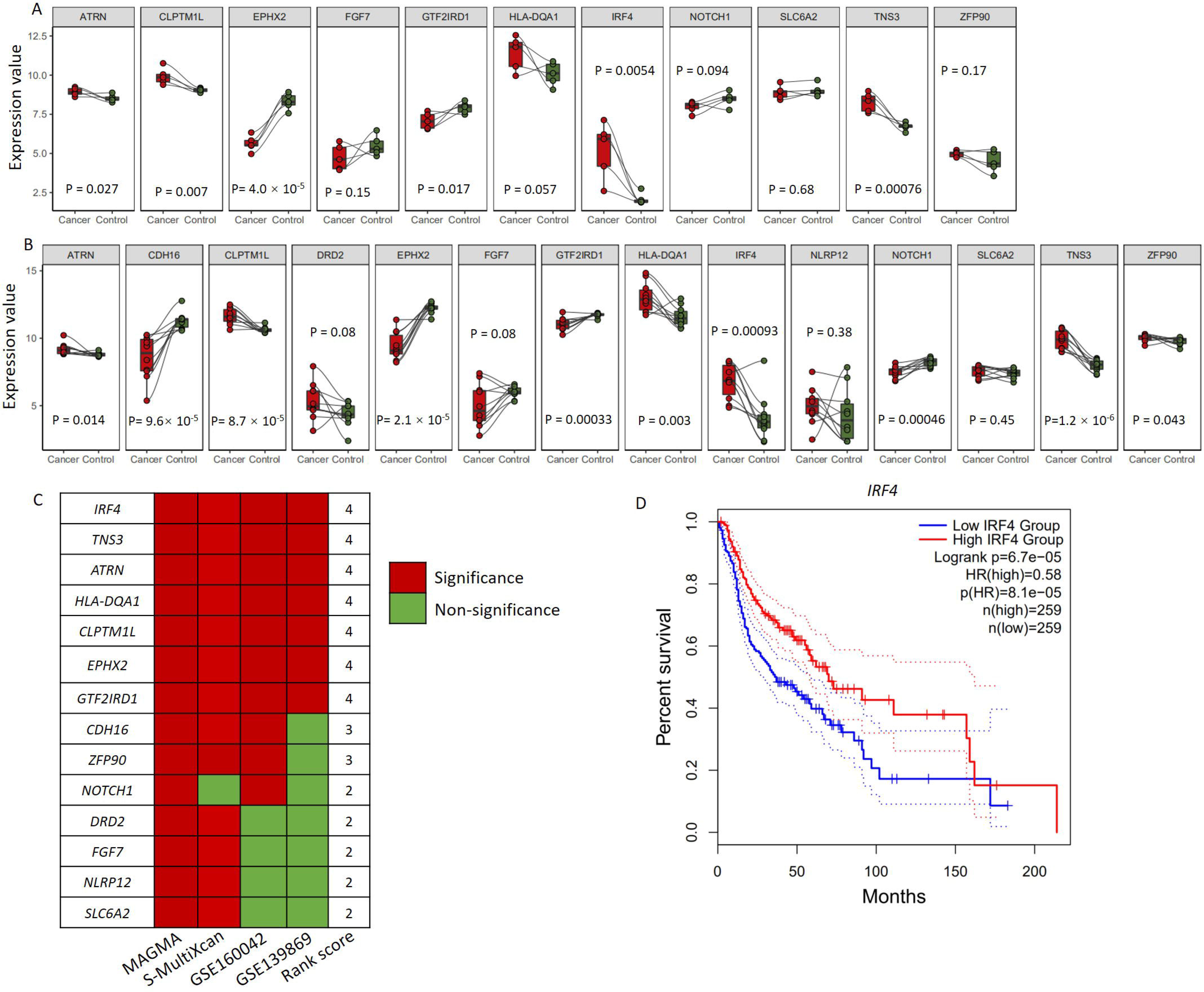
Differential gene expression analyses of 14 risk genes across three independent datasets. A) Boxplots showing the differential expression patterns of 11 genes between OCC patients and paracancerous controls among the GSE139869 dataset. There were three genes that are *CDH16*, *DRD2*, and *NLRP12* not expressed in this dataset. B) Boxplots showing the differential expression patterns of 14 genes between OCC patients and paracancerous controls among the GSE160042 dataset. C) Summing rank score for these 14 risk genes based on multiple lines of evidence from MAGMA, S-MultiXcan, GSE160042, and GSE139869. D) Kaplan-Meier curve exhibiting the result of survive analysis for *IRF4* based on the expression dataset on head and neck squamous cell carcinoma from the TCGA database.

Furthermore, to determine whether the co-expression patterns among these 14 genetics-risk genes were changed by OCC disease status, we conducted a Pearson correlation analysis for the expression data from three datasets. Interestingly, we observed that an obvious change of the co-expression patterns among 14 genes categorized by OCC status (Supplemental Figures S29-S30). For example, the correlation coefficient between *ATRN* and *CLPTM1L* was fundamentally increased from -0.356 in controls to 0.779 in OCC patients among the GSE160042 dataset.

Integrating multi-omic evidence across different datasets, we calculated the ranked scores of these 14 risk genes by summing five pieces of evidence from genetic, eQTL, and gene expression, and found that the novel risk gene of *IRF4* exhibited the highest rank score (Figure 7C). Based on a large-scale TCGA dataset on head and neck squamous cell carcinoma (N = 518), we found the expression level of *IRF4* gene was significantly associated with patients’ survive (P = 8.1×10^-5^, Figure 7D).

## 4. Discussion

Previous genetic studies have indicated that there existed a crucial role of genetic susceptibility in the etiology of OCC [47, 48]. Many genes have been reported to be associated with OCC, such as *ADH1B* and *ADH7* [5, 6, 49]. Lessuer and coworkers [16] found four genetic loci, including 2p23.3, 5p15.33, 9q15.3 and 9q34.12, were prominently associated with OCC. Another study [50] identified six loci including 5q14.3, 6p21.33, 6q16.1, 11q12.2, 12q24.21 and 16p13.2 were remarkably associated with the laryngeal cancer risk. Recently, Shete et al. [15] conducted a two-phase GWAS based on non-Hispanic white for identifying oral cancer-associated genetic loci, and replicated four known cancer loci (namely 2p23.1, 5p15.33, 6p21.32, and 6p21.33) associated with both oral cancer and oropharyngeal cancer risk. However, these reported genetic risk loci are still limited and only explain a small proportion of heritability. More comprehensive genomic studies are warranted.

In the current study, by performing an integrative genomics analysis, we found that 14 risk genes, including *IRF4, TNS3, NOTCH1, ZFP90, ATRN, HLA-DQA1, DRD2, EPHX2, CLPTM1L, SLC6A2, FGF7, GTF2IRD1, NLRP12,* and *CDH16,* contribute to OCC susceptibility. By collecting cancer-related genes from the GWAS Catalog database, seven genes that are *IRF4, TNS3, HLA-DQA1, EPHX2, CLPTM1L, FGF7,* and *NLRP12* have been documented to be associated with the susceptibility of other cancer types [51–58], and five genes that are *NOTCH1, FGF7, SLC6A2, HLA-DQA1,* and *CLPTM1L* have been reported to be associated with oral and pharynx cancer [15–19]. Based on the DisGeNET method, we observed that 14 genes were significantly enriched in cancer-related terms, including malignant carcinoid syndrome, small lymphocytic lymphoma, basal cell neoplasm, and basal cell cancer. These results suggest the 14 identified risk genes may have important functions in OCC. Additionally, we also found that 71.43% of these genes were significantly enriched in 10 druggable gene categories, and five genes that are *NOTCH1, SLC6A2, DRD2, HLA-DQA1,* and *EPHX2* were targeted by at least one known drug, indicating that these identified genes have druggable functions and may as good candidates for drug repurposing in OCC.

The novel risk gene *IRF4*, which is a tumor suppressor gene, encodes a protein that belongs to the interferon regulatory factor family of transcription factors, which have important roles in modulating interferon-inducible genes and activating the innate and adaptive immune systems. The *IRF4* gene was initially found as the transcription factors responsible for the generation of regulatory CD4+T cells specifically controlling Th2 response [59, 60]. Zhang and coworkers [60] demonstrated that mice with specific ablation of *Irf4* in T regulatory cells develop multi-organ autoimmunity because of exacerbated Th1, Tfh, and Th17 responses and plasma cell infiltration. Multiple lines of evidence have demonstrated that *IRF4* gene was significantly associated with numerous human cancers, including myeloma [61], non-small-cell lung cancer [62], breast cancer [63], and large B-cell lymphomas [64]. By performing a large-scale bisulfite genomic sequencing analysis, Vernier et al. [63] reported that reversal of promoter hyper-methylation and consequently de-repression of *IRF4* gene is a factor contributing to the suppression of breast cancer.

The *NOTCH1* gene, which encodes a member of the NOTCH family of proteins, has an important role in the development of numerous cells and tissue types. Mutations in the *NOTCH1* gene have been reported to be associated with chronic lymphocytic leukemia [65–68], pancreatic cancer metastasis [69], head and neck squamous cell carcinoma [70–73], T-cell acute lymphoblastic leukemia [74], and oral cavity or tongue cancer [17, 75, 76]. For example, based on whole exome sequencing and gene copy number analyses, Agrawal et al. [73] reported approximate 40% of the 28 mutation in the *NOTCH1* gene were predicted to truncate the gene product, which indicated that *NOTCH1* may play as a tumor suppressor gene instead of an oncogene. Meanwhile, Stransky and colleagues [70] performed whole exome-wide sequencing analysis and found at least 30% of patients harbored mutations in genes including *NOTCH1*, which is a major driver of head and neck squamous cell carcinoma for modulating squamous differentiation. Using the literature-mining analysis by combining 26 studies with 17,675 samples, Ma et al. [77] found 48 smoking-related methylation genes including *NOTCH1* in the 11 oncogenic pathways, which were associated with lung, oral, and other cancers. Furthermore, we identified that there were 10 drugs targeted with *NOTCH1*. For example, previous studies have demonstrated that brontictuzumab is a humanized monoclonal antibody that targets *NOTCH1* and inhibits pathway activation and is designed to treat cancer [78], and nirogacestat, which has an inhibitory interaction with *NOTCH1*, is an oral small-molecule gamma secretase inhibitor in phase 3 clinical development for patients with cancer [79].

The druggable gene of *SLC6A2*, which encodes a member of the sodium neurotransmitter symporter family that is responsible for the reuptake of norepinephrine into presynaptic nerve terminals, was identified to be targeted by ten drugs, including reboxetine, dexmethylphenidate, and guanethidine. Previous studies have demonstrated that *SLC6A2* is associated with neuroblastoma [80], colorectal cancer [81], and head and neck cancer [18]. Multiple lines of evidence have shown that a number of neurotransmitters have roles in the development and maintenance of fatigue and energy levels in breast cancer [82]. With regard to the *HLA-DQA1* druggable gene, it belongs to the HLA class II alpha chain paralogues and plays a crucial role in the immune system by presenting extracellular peptides. Multiple earlier studies have demonstrated that mutations in *HLA-DQA1* are significantly associated with oral cancer [16, 83], hepatocellular carcinoma [84], cervical cancer[85], and gastric cancer [86].

With regard to the druggable gene of *DRD2*, it encodes the D2 subtype of the dopamine receptor, which belongs to the G-protein coupled receptor that inhibits adenylylcyclase activity. DRD2 as a G-protein coupled receptor showed over-expressions in glioblastoma and controls growth factor signaling via cross-talk mechanisms involving beta-arrestin and scaffold protein [87]. By blockading DRD2 in a preclinical model of glioblastoma, it enables to inactivate growth factor signaling and induce tumor cell death [88]. Based on results from epidemiological statistical analysis, patients with schizophrenia, who inherently have elevated DRD2 signaling, have an elevated risk of cancer, whereas this risk could be recovered to normal by treating with DRD2 antagonists [89]. Furthermore, DRD2 has been reported to be involved in other cancer types, including breast cancer, colorectal cancer, and prostate cancer [90–92]. Mounting evidence has shown that *DRD2* was significantly associated with smoking behaviors [93–98] and alcohol consumption [99–101] among various ethnicities. For example, by performing a meta-analysis based on 11,075 participants, Ma et al. [93] reported that the Taq1A A1/* genotypes were remarkably associated with smoking cessation under both the fixed- and random-effects models. Based on a GWAS of self-reported alcohol consumption in 112,117 subjects from the UKBiobank database, Clarke et al. [100] demonstrated that *DRD2* was found to implicate in the neurobiology of alcohol use. Taken together, these identified risk genes potentially have important roles in the carcinogenesis of oral cavity cancer.

There exist some limitations of the present study needed to be cautious. Although we collected multiple layers of omics data, there were other useful omics data not adopted in the current investigation, such as histone marks [102–105] and DNAase I-hypersensitive sites (DHS) [106, 107]. It should be noted that the sample size used in the GWAS summary statistics was not very large. Furthermore, due to the heterogeneity across different datasets, we applied distinct correction methods for multiple testing for each omics dataset. The current study was based on the European population, and we did not validate these OCC-risk genes in other ethnicities. Further studies are needed to evaluate the inference by using genotype and gene expression data from other ancestries.

## Conclusion

In summary, our integrative genomics analysis provides an effective method for identifying risk genes that convey susceptibility to OCC, and highlights 14 important OCC-risk genes with 9 novel risk genes, such as *IRF4*, *TNS3*, and *DRD2*. These identified genes targeted with drugs may be promising candidates for drug repurposing in the future pharmaceutical studies. Furthermore, we connected risk variants with susceptible genes and functional processes, providing a good clue for explaining the effects of genetic variants on OCC. More relevant studies are warranted to study the molecular functions and therapeutic efficacy of these identified risk genes.

## Data Availability

All the GWAS summary statistics on oral cancer applied in the present analysis can be obtained from the MRC Integrative Epidemiology Unit (IEU) database (https://gwas.mrcieu.ac.uk/, accession ID: ieu-b-94). The GTEx eQTL data (version 8) were gained from the Zenodo repository (https://zenodo.org/record/3518299#.Xv6Z6igzbgl). The 1,000 Genomes Project European Phase 3 reference panel was downloaded from the IGSR: The International Genome Sample Resource (https://www.internationalgenome.org/category/phase-3/).

## Abbreviations

OCC: Oral cavity cancer
GWAS: Genome-wide association study
eQTL: expression quantitative loci trait
sQTL: splicing quantitative loci trait
SNP: Single nucleotide polymorphism
CC: Cellular component
MF: Molecular function
BP: Biological process
IEU: Integrative Epidemiology Unit
INHANCE: International Head and Neck Cancer Epidemiology Consortium
OR: Odds ratio
MAGMA: the Multi-marker Analysis of GenoMic Annotation
LD: Linkage disequilibrium
FDR: False discovery rate
WebGestalt: WEB-based Gene SeT AnaLysis Toolkit
GO: Gene Ontology
PPI: Protein-protein interaction
GEO: the database of Gene Expression Omnibus
TCGA: The Cancer Genome Atlas.

## Authors’ contributions

Y.L., X.X., Z.W., M.W., Y.M., and Y.H. assimilated the data, performed integrative genomics analysis, and wrote the manuscript. Y.L., Y.M., Y.Y., and M.W. were involved in data collection and reviewed the manuscript. Y.M. and M.W. conceived the current study and reviewed the manuscript. All authors have reviewed the current version, and approved the finial manuscript.

## Acknowledgements

Not applicable.

## Supplemental Figures

**Supplemental Figure S1. Quantile-quantile (QQ) plot of the MAGMA-based gene-level association analysis on GWAS summary statistics.**

**Supplemental Figure S2. The 15 lead SNPs with 14 genomic loci associated with OCC**. Left panel shows the ID, P value, risk allele, and chromosome of each index SNP, and right panel shows the odds ratio with 95% confidence interval.

**Supplemental Figure S3. Regional plot for the OCC-associated risk gene of ZFP90 identified from MAGMA analysis.** The purple diamond marks the most strongly associated SNP of rs10775305 in each gene with OCC. The color illustrates LD information with the given SNP, as shown in the color legend.

**Supplemental Figure S4. Regional plot for the OCC-associated risk gene of *CLPTM1L* identified from MAGMA analysis.** The purple diamond marks the most strongly associated SNP of rs10462706 in each gene with OCC. The color illustrates LD information with the given SNP, as shown in the color legend.

**Supplemental Figure S5. Regional plot for the OCC-associated risk gene of *CDH16* identified from MAGMA analysis.** The purple diamond marks the most strongly associated SNP of rs6499103 in each gene with OCC. The color illustrates LD information with the given SNP, as shown in the color legend.

**Supplemental Figure S6. Regional plot for the OCC-associated risk gene of *DRD2* identified from MAGMA analysis.** The purple diamond marks the most strongly associated SNP of rs4586205 in each gene with OCC. The color illustrates LD information with the given SNP, as shown in the color legend.

**Supplemental Figure S7. Regional plot for the OCC-associated risk gene of *EPHX2* identified from MAGMA analysis.** The purple diamond marks the most strongly associated SNP of rs10283378 in each gene with OCC. The color illustrates LD information with the given SNP, as shown in the color legend.

**Supplemental Figure S8. Regional plot for the OCC-associated risk gene of *HLA-DQA1* identified from MAGMA analysis.** The purple diamond marks the most strongly associated SNP of rs9271378 in each gene with OCC. The color illustrates LD information with the given SNP, as shown in the color legend.

**Supplemental Figure S9. Regional plot for the OCC-associated risk gene of *TNS3* identified from MAGMA analysis.** The purple diamond marks the most strongly associated SNP of rs35894636 in each gene with OCC. The color illustrates LD information with the given SNP, as shown in the color legend.

**Supplemental Figure S10. Regional plot for the OCC-associated risk gene of *GTF2IRD1* identified from MAGMA analysis.** The purple diamond marks the most strongly associated SNP of rs4717897 in each gene with OCC. The color illustrates LD information with the given SNP, as shown in the color legend.

**Supplemental Figure S11. Regional plot for the OCC-associated risk gene of *NOTCH1* identified from MAGMA analysis.** The purple diamond marks the most strongly associated SNP of rs3125011 in each gene with OCC. The color illustrates LD information with the given SNP, as shown in the color legend.

**Supplemental Figure S12. Regional plot for the OCC-associated risk gene of *FGF7* identified from MAGMA analysis.** The purple diamond marks the most strongly associated SNP of rs12916839 in each gene with OCC. The color illustrates LD information with the given SNP, as shown in the color legend.

**Supplemental Figure S13. Regional plot for the OCC-associated risk gene of *SLC6A2* identified from MAGMA analysis.** The purple diamond marks the most strongly associated SNP of rs2397776 in each gene with OCC. The color illustrates LD information with the given SNP, as shown in the color legend.

**Supplemental Figure S14. Regional plot for the OCC-associated risk gene of *NLRP12* identified from MAGMA analysis.** The purple diamond marks the most strongly associated SNP of rs4806514 in each gene with OCC. The color illustrates LD information with the given SNP, as shown in the color legend.

**Supplemental Figure S15. Regional plot for the OCC-associated risk gene of *ATRN* identified from MAGMA analysis.** The purple diamond marks the most strongly associated SNP of rs6051873 in each gene with OCC. The color illustrates LD information with the given SNP, as shown in the color legend.

**Supplemental Figure S16. Regional plot for the OCC-associated risk gene of *ATRN* identified from MAGMA analysis.** The purple diamond marks the most strongly associated SNP of rs17711842 in each gene with OCC. The color illustrates LD information with the given SNP, as shown in the color legend.

**Supplemental Figure S17. Multiple layers of evidence supporting the functional role of rs7773324 in *IRF4*.** There were three distinct data used for assessing the variant functionality, including CADD score, regulomeDB, and chromatin state. This plot was generated by using the FUMA online tool (https://fuma.ctglab.nl/).

**Supplemental Figure S18. Multiple layers of evidence supporting the functional role of rs3125011 in *NOTCH1*.** There were three distinct data used for assessing the variant functionality, including CADD score, regulomeDB, and chromatin state. This plot was generated by using the FUMA online tool (https://fuma.ctglab.nl/).

**Supplemental Figure S19. Multiple layers of evidence supporting the functional role of rs12916839 in *FGF7.*** There were three distinct data used for assessing the variant functionality, including CADD score, regulomeDB, and chromatin state. This plot was generated by using the FUMA online tool (https://fuma.ctglab.nl/).

**Supplemental Figure S20. Violin plots showing rs12916839 influences the expression of *FGF7* across multiple tissues.** A)-B) Violin plots showing rs12916839 represents an eQTL quantitative trait locus for *FGF7* across multiple GTEx tissues. C)-I) Violin plots showing rs12916839 represents a splicing quantitative trait locus for *FGF7* across multiple GTEx tissues.

**Supplemental Figure S21. Violin plots showing rs12916839 influences the expression of *FGF7* across multiple tissues.** For A)-I), there were 9 tissues showing a significant sQTL for rs12916839.

**Supplemental Figure S22. Violin plots showing rs12916839 represents a splicing quantitative trait locus for *FGF7* across multiple GTEx tissues.** For A)-L), there were 12 tissues showing a significant sQTL for rs12916839.

**Supplemental Figure S23. S-PrediXcan-based integrative genomics analysis identified risk genes associated with oral cancer in each tissue.** A) Whole blood, B) Thyroid, C) Skin sun exposed lower leg, D) Skin not sun exposed suprapubic, E) Muscle skeletal, and F) Minor salivary gland.

**Supplemental Figure S24. S-PrediXcan-based integrative genomics analysis identified risk genes associated with oral cancer in each tissue.** A) Liver, B) Lung, C) Esophagus Mucosa, D) Adrenal Gland, E) Adipose Visceral Omentum, and F) Adipose Subcutaneous.

**Supplemental Figure S25. S-PrediXcan-based integrative genomics analysis identified risk genes associated with oral cancer in each tissue.** A) Esophagus_Gastroesophageal_Junction, B) Esophagus Muscularis, C) Artery Aorta, D) Artery Coronary, E) Cells- EBV-transformed lymphocytes, and F) Cells-Cultured fibroblasts.

**Supplemental Figure S26. S-PrediXcan-based integrative genomics analysis identified risk genes associated with oral cancer in each tissue.** A) Colon Transverse , B) Pancreas, C) Colon Sigmoid, D) Pituitary, E) Small intestine terminal ileum, and F) Stomach.

**Supplemental Figure S27. Bar plot shows the enrichment results of these 14 common risk genes identified from MAGMA and S-MultiXcan by using the DisGeNET algorithm in the Metascape.**

**Supplemental Figure S28. Drug-gene enrichment analysis shows 10 of 14 risk genes enriched in 10 potential druggable gene categories based on the DGIdb database.**

**Supplemental Figure S29. Co-expression patterns of 14 genes between OCC patients and paracancerous controls based on the GSE139869 dataset.**

**Supplemental Figure S30. Co-expression patterns of 14 genes between OCC patients and paracancerous controls based on the GSE160042 dataset.**

## References

1. Torre LA, Bray F, Siegel RL, Ferlay J, Lortet-Tieulent J, Jemal A: Global cancer statistics, 2012. CA Cancer J Clin 2015, 65:87–108.

2. Siegel RL, Miller KD, Jemal A: Cancer statistics, 2020. CA Cancer J Clin 2020, 70:7–30.

3. Montero PH, Patel SG: Cancer of the oral cavity. Surg Oncol Clin N Am 2015, 24:491–508.

4. Kumar M, Nanavati R, Modi TG, Dobariya C: Oral cancer: Etiology and risk factors: A review. J Cancer Res Ther 2016, 12:458–463.

5. Hashibe M, McKay JD, Curado MP, Oliveira JC, Koifman S, Koifman R, Zaridze D, Shangina O, Wünsch-Filho V, Eluf-Neto J, et al: Multiple ADH genes are associated with upper aerodigestive cancers. Nat Genet 2008, 40:707–709.

6. McKay JD, Truong T, Gaborieau V, Chabrier A, Chuang SC, Byrnes G, Zaridze D, Shangina O, Szeszenia-Dabrowska N, Lissowska J, et al: A genome-wide association study of upper aerodigestive tract cancers conducted within the INHANCE consortium. PLoS Genet 2011, 7:e1001333.

7. Xu M, Li J, Xiao Z, Lou J, Pan X, Ma Y: Integrative genomics analysis identifies promising SNPs and genes implicated in tuberculosis risk based on multiple omics datasets. Aging (Albany NY) 2020, 12:19173–19220.

8. Visscher PM, Brown MA, McCarthy MI, Yang J: Five years of GWAS discovery. Am J Hum Genet 2012, 90:7–24.

9. Canela-Xandri O, Rawlik K, Tenesa A: An atlas of genetic associations in UK Biobank. Nat Genet 2018, 50:1593–1599.

10. Sudlow C, Gallacher J, Allen N, Beral V, Burton P, Danesh J, Downey P, Elliott P, Green J, Landray M, et al: UK biobank: an open access resource for identifying the causes of a wide range of complex diseases of middle and old age. PLoS Med 2015, 12:e1001779.

11. Ma Y, Huang Y, Zhao S, Yao Y, Zhang Y, Qu J, Wu N, Su J: Integrative Genomics Analysis Reveals a 21q22.11 Locus Contributing Risk to COVID-19. Hum Mol Genet 2021.

12. The COVID-19 Host Genetics Initiative, a global initiative to elucidate the role of host genetic factors in susceptibility and severity of the SARS-CoV-2 virus pandemic. Eur J Hum Genet 2020, 28:715–718.

13. Pairo-Castineira E, Clohisey S, Klaric L, Bretherick AD, Rawlik K, Pasko D, Walker S, Parkinson N, Fourman MH, Russell CD, et al: Genetic mechanisms of critical illness in COVID-19. Nature 2021, 591:92–98.

14. Buniello A, MacArthur JAL, Cerezo M, Harris LW, Hayhurst J, Malangone C, McMahon A, Morales J, Mountjoy E, Sollis E, et al: The NHGRI-EBI GWAS Catalog of published genome-wide association studies, targeted arrays and summary statistics 2019. Nucleic Acids Res 2019, 47:D1005–d1012.

15. Shete S, Liu H, Wang J, Yu R, Sturgis EM, Li G, Dahlstrom KR, Liu Z, Amos CI, Wei Q: A Genome-Wide Association Study Identifies Two Novel Susceptible Regions for Squamous Cell Carcinoma of the Head and Neck. Cancer Res 2020, 80:2451–2460.

16. Lesseur C, Diergaarde B, Olshan AF, Wünsch-Filho V, Ness AR, Liu G, Lacko M, Eluf-Neto J, Franceschi S, Lagiou P, et al: Genome-wide association analyses identify new susceptibility loci for oral cavity and pharyngeal cancer. Nat Genet 2016, 48:1544–1550.

17. Chai AWY, Lim KP, Cheong SC: Translational genomics and recent advances in oral squamous cell carcinoma. Semin Cancer Biol 2020, 61:71–83.

18. Lopes-Santos G, Bernabé DG, Miyahara GI, Tjioe KC: Beta-adrenergic pathway activation enhances aggressiveness and inhibits stemness in head and neck cancer. Transl Oncol 2021, 14:101117.

19. Maruyama S, Cheng J, Yamazaki M, Zhou XJ, Zhang ZY, He RG, Saku T: Metastasis-associated genes in oral squamous cell carcinoma and salivary adenoid cystic carcinoma: a differential DNA chip analysis between metastatic and nonmetastatic cell systems. Cancer Genet Cytogenet 2010, 196:14–22.

20. Hindorff LA, Sethupathy P, Junkins HA, Ramos EM, Mehta JP, Collins FS, Manolio TA: Potential etiologic and functional implications of genome-wide association loci for human diseases and traits. Proc Natl Acad Sci U S A 2009, 106:9362–9367.

21. Li MJ, Liu Z, Wang P, Wong MP, Nelson MR, Kocher JP, Yeager M, Sham PC, Chanock SJ, Xia Z, Wang J: GWASdb v2: an update database for human genetic variants identified by genome-wide association studies. Nucleic Acids Res 2016, 44:D869–876.

22. He X, Fuller CK, Song Y, Meng Q, Zhang B, Yang X, Li H: Sherlock: detecting gene-disease associations by matching patterns of expression QTL and GWAS. Am J Hum Genet 2013, 92:667–680.

23. Dong Z, Ma Y, Zhou H, Shi L, Ye G, Yang L, Liu P, Zhou L: Integrated genomics analysis highlights important SNPs and genes implicated in moderate-to-severe asthma based on GWAS and eQTL datasets. BMC Pulm Med 2020, 20:270.

24. Schadt EE, Lamb J, Yang X, Zhu J, Edwards S, Guhathakurta D, Sieberts SK, Monks S, Reitman M, Zhang C, et al: An integrative genomics approach to infer causal associations between gene expression and disease. Nat Genet 2005, 37:710–717.

25. Zhu Z, Zhang F, Hu H, Bakshi A, Robinson MR, Powell JE, Montgomery GW, Goddard ME, Wray NR, Visscher PM, Yang J: Integration of summary data from GWAS and eQTL studies predicts complex trait gene targets. Nat Genet 2016, 48:481–487.

26. Sun H, Zhang J, Ma Y, Liu J: Integrative genomics analysis identifies five promising genes implicated in insomnia risk based on multiple omics datasets. Biosci Rep 2020, 40.

27. Luo XJ, Mattheisen M, Li M, Huang L, Rietschel M, Børglum AD, Als TD, van den Oord EJ, Aberg KA, Mors O, et al: Systematic Integration of Brain eQTL and GWAS Identifies ZNF323 as a Novel Schizophrenia Risk Gene and Suggests Recent Positive Selection Based on Compensatory Advantage on Pulmonary Function. Schizophr Bull 2015, 41:1294–1308.

28. Ma C, Gu C, Huo Y, Li X, Luo XJ: The integrated landscape of causal genes and pathways in schizophrenia. Transl Psychiatry 2018, 8:67.

29. Barbeira AN, Pividori M, Zheng J, Wheeler HE, Nicolae DL, Im HK: Integrating predicted transcriptome from multiple tissues improves association detection. PLoS Genet 2019, 15:e1007889.

30. Chang CC, Chow CC, Tellier LC, Vattikuti S, Purcell SM, Lee JJ: Second-generation PLINK: rising to the challenge of larger and richer datasets. Gigascience 2015, 4:7.

31. . The Genotype-Tissue Expression (GTEx) project. Nat Genet 2013, 45:580–585.

32. Auton A, Brooks LD, Durbin RM, Garrison EP, Kang HM, Korbel JO, Marchini JL, McCarthy S, McVean GA, Abecasis GR: A global reference for human genetic variation. Nature 2015, 526:68–74.

33. de Leeuw CA, Mooij JM, Heskes T, Posthuma D: MAGMA: generalized gene-set analysis of GWAS data. PLoS Comput Biol 2015, 11:e1004219.

34. Ma X, Wang P, Xu G, Yu F, Ma Y: Integrative genomics analysis of various omics data and networks identify risk genes and variants vulnerable to childhood-onset asthma. BMC Med Genomics 2020, 13:123.

35. Kanehisa M, Goto S: KEGG: kyoto encyclopedia of genes and genomes. Nucleic Acids Res 2000, 28:27–30.

36. Wang J, Duncan D, Shi Z, Zhang B: WEB-based GEne SeT AnaLysis Toolkit (WebGestalt): update 2013. Nucleic Acids Res 2013, 41:W77–83.

37. von Mering C, Huynen M, Jaeggi D, Schmidt S, Bork P, Snel B: STRING: a database of predicted functional associations between proteins. Nucleic Acids Res 2003, 31:258–261.

38. Warde-Farley D, Donaldson SL, Comes O, Zuberi K, Badrawi R, Chao P, Franz M, Grouios C, Kazi F, Lopes CT, et al: The GeneMANIA prediction server: biological network integration for gene prioritization and predicting gene function. Nucleic Acids Res 2010, 38:W214–220.

39. Shannon P, Markiel A, Ozier O, Baliga NS, Wang JT, Ramage D, Amin N, Schwikowski B, Ideker T: Cytoscape: a software environment for integrated models of biomolecular interaction networks. Genome Res 2003, 13:2498–2504.

40. Jourquin J, Duncan D, Shi Z, Zhang B: GLAD4U: deriving and prioritizing gene lists from PubMed literature. BMC Genomics 2012, 13 Suppl 8:S20.

41. Robinson MD, Oshlack A: A scaling normalization method for differential expression analysis of RNA-seq data. Genome Biol 2010, 11:R25.

42. Rashkin SR, Graff RE, Kachuri L, Thai KK, Alexeeff SE, Blatchins MA, Cavazos TB, Corley DA, Emami NC, Hoffman JD, et al: Pan-cancer study detects genetic risk variants and shared genetic basis in two large cohorts. Nat Commun 2020, 11:4423.

43. Piñero J, Bravo À, Queralt-Rosinach N, Gutiérrez-Sacristán A, Deu-Pons J, Centeno E, García-García J, Sanz F, Furlong LI: DisGeNET: a comprehensive platform integrating information on human disease-associated genes and variants. Nucleic Acids Res 2017, 45:D833–d839.

44. Zhou Y, Zhou B, Pache L, Chang M, Khodabakhshi AH, Tanaseichuk O, Benner C, Chanda SK: Metascape provides a biologist-oriented resource for the analysis of systems-level datasets. Nat Commun 2019, 10:1523.

45. Lin A, Wang RT, Ahn S, Park CC, Smith DJ: A genome-wide map of human genetic interactions inferred from radiation hybrid genotypes. Genome Res 2010, 20:1122–1132.

46. Dobbin KK, Beer DG, Meyerson M, Yeatman TJ, Gerald WL, Jacobson JW, Conley B, Buetow KH, Heiskanen M, Simon RM, et al: Interlaboratory comparability study of cancer gene expression analysis using oligonucleotide microarrays. Clin Cancer Res 2005, 11:565–572.

47. Ho T, Wei Q, Sturgis EM: Epidemiology of carcinogen metabolism genes and risk of squamous cell carcinoma of the head and neck. Head Neck 2007, 29:682–699.

48. Neumann AS, Sturgis EM, Wei Q: Nucleotide excision repair as a marker for susceptibility to tobacco-related cancers: a review of molecular epidemiological studies. Mol Carcinog 2005, 42:65–92.

49. Wei S, Liu Z, Zhao H, Niu J, Wang LE, El-Naggar AK, Sturgis EM, Wei Q: A single nucleotide polymorphism in the alcohol dehydrogenase 7 gene (alanine to glycine substitution at amino acid 92) is associated with the risk of squamous cell carcinoma of the head and neck. Cancer 2010, 116:2984–2992.

50. Wei Q, Yu D, Liu M, Wang M, Zhao M, Liu M, Jia W, Ma H, Fang J, Xu W, et al: Genome-wide association study identifies three susceptibility loci for laryngeal squamous cell carcinoma in the Chinese population. Nat Genet 2014, 46:1110–1114.

51. Liyanage UE, Law MH, Han X, An J, Ong JS, Gharahkhani P, Gordon S, Neale RE, Olsen CM, MacGregor S, Whiteman DC: Combined analysis of keratinocyte cancers identifies novel genome-wide loci. Hum Mol Genet 2019, 28:3148–3160.

52. McKay JD, Hung RJ, Han Y, Zong X, Carreras-Torres R, Christiani DC, Caporaso NE, Johansson M, Xiao X, Li Y, et al: Large-scale association analysis identifies new lung cancer susceptibility loci and heterogeneity in genetic susceptibility across histological subtypes. Nat Genet 2017, 49:1126–1132.

53. Sarin KY, Lin Y, Daneshjou R, Ziyatdinov A, Thorleifsson G, Rubin A, Pardo LM, Wu W, Khavari PA, Uitterlinden A, et al: Genome-wide meta-analysis identifies eight new susceptibility loci for cutaneous squamous cell carcinoma. Nat Commun 2020, 11:820.

54. Visconti A, Duffy DL, Liu F, Zhu G, Wu W, Chen Y, Hysi PG, Zeng C, Sanna M, Iles MM, et al: Genome-wide association study in 176,678 Europeans reveals genetic loci for tanning response to sun exposure. Nat Commun 2018, 9:1684.

55. Figueiredo JC, Hsu L, Hutter CM, Lin Y, Campbell PT, Baron JA, Berndt SI, Jiao S, Casey G, Fortini B, et al: Genome-wide diet-gene interaction analyses for risk of colorectal cancer. PLoS Genet 2014, 10:e1004228.

56. Eeles RA, Olama AA, Benlloch S, Saunders EJ, Leongamornlert DA, Tymrakiewicz M, Ghoussaini M, Luccarini C, Dennis J, Jugurnauth-Little S, et al: Identification of 23 new prostate cancer susceptibility loci using the iCOGS custom genotyping array. Nat Genet 2013, 45:385–391, 391e381-382.

57. Huyghe JR, Bien SA, Harrison TA, Kang HM, Chen S, Schmit SL, Conti DV, Qu C, Jeon J, Edlund CK, et al: Discovery of common and rare genetic risk variants for colorectal cancer. Nat Genet 2019, 51:76–87.

58. Litchfield K, Levy M, Orlando G, Loveday C, Law PJ, Migliorini G, Holroyd A, Broderick P, Karlsson R, Haugen TB, et al: Identification of 19 new risk loci and potential regulatory mechanisms influencing susceptibility to testicular germ cell tumor. Nat Genet 2017, 49:1133–1140.

59. Alvisi G, Brummelman J, Puccio S, Mazza EM, Tomada EP, Losurdo A, Zanon V, Peano C, Colombo FS, Scarpa A, et al: IRF4 instructs effector Treg differentiation and immune suppression in human cancer. J Clin Invest 2020, 130:3137–3150.

60. Zheng Y, Chaudhry A, Kas A, deRoos P, Kim JM, Chu TT, Corcoran L, Treuting P, Klein U, Rudensky AY: Regulatory T-cell suppressor program co-opts transcription factor IRF4 to control T(H)2 responses. Nature 2009, 458:351–356.

61. Agnarelli A, Chevassut T, Mancini EJ: IRF4 in multiple myeloma-Biology, disease and therapeutic target. Leuk Res 2018, 72:52–58.

62. Akimova T, Zhang T, Negorev D, Singhal S, Stadanlick J, Rao A, Annunziata M, Levine MH, Beier UH, Diamond JM, et al: Human lung tumor FOXP3+ Tregs upregulate four “Treg-locking” transcription factors. JCI Insight 2017, 2.

63. Vernier M, McGuirk S, Dufour CR, Wan L, Audet-Walsh E, St-Pierre J, Giguère V: Inhibition of DNMT1 and ERRα crosstalk suppresses breast cancer via derepression of IRF4. Oncogene 2020, 39:6406–6420.

64. Ramis-Zaldivar JE, Gonzalez-Farré B, Balagué O, Celis V, Nadeu F, Salmerón-Villalobos J, Andrés M, Martin-Guerrero I, Garrido-Pontnou M, Gaafar A, et al: Distinct molecular profile of IRF4-rearranged large B-cell lymphoma. Blood 2020, 135:274–286.

65. Edelmann J, Holzmann K, Tausch E, Saunderson EA, Jebaraj BMC, Steinbrecher D, Dolnik A, Blätte TJ, Landau DA, Saub J, et al: Genomic alterations in high-risk chronic lymphocytic leukemia frequently affect cell cycle key regulators and NOTCH1-regulated transcription. Haematologica 2020, 105:1379–1390.

66. Arruga F, Bracciamà V, Vitale N, Vaisitti T, Gizzi K, Yeomans A, Coscia M, D’Arena G, Gaidano G, Allan JN, et al: Bidirectional linkage between the B-cell receptor and NOTCH1 in chronic lymphocytic leukemia and in Richter’s syndrome: therapeutic implications. Leukemia 2020, 34:462–477.

67. Rosati E, Baldoni S, De Falco F, Del Papa B, Dorillo E, Rompietti C, Albi E, Falzetti F, Di Ianni M, Sportoletti P: NOTCH1 Aberrations in Chronic Lymphocytic Leukemia. Front Oncol 2018, 8:229.

68. Nadeu F, Delgado J, Royo C, Baumann T, Stankovic T, Pinyol M, Jares P, Navarro A, Martín-García D, Beà S, et al: Clinical impact of clonal and subclonal TP53, SF3B1, BIRC3, NOTCH1, and ATM mutations in chronic lymphocytic leukemia. Blood 2016, 127:2122–2130.

69. Geng Y, Fan J, Chen L, Zhang C, Qu C, Qian L, Chen K, Meng Z, Chen Z, Wang P: A Notch-Dependent Inflammatory Feedback Circuit between Macrophages and Cancer Cells Regulates Pancreatic Cancer Metastasis. Cancer Res 2021, 81:64–76.

70. Stransky N, Egloff AM, Tward AD, Kostic AD, Cibulskis K, Sivachenko A, Kryukov GV, Lawrence MS, Sougnez C, McKenna A, et al: The mutational landscape of head and neck squamous cell carcinoma. Science 2011, 333:1157–1160.

71. Shah PA, Huang C, Li Q, Kazi SA, Byers LA, Wang J, Johnson FM, Frederick MJ: NOTCH1 Signaling in Head and Neck Squamous Cell Carcinoma. Cells 2020, 9.

72. Comprehensive genomic characterization of head and neck squamous cell carcinomas. Nature 2015, 517:576–582.

73. Agrawal N, Frederick MJ, Pickering CR, Bettegowda C, Chang K, Li RJ, Fakhry C, Xie TX, Zhang J, Wang J, et al: Exome sequencing of head and neck squamous cell carcinoma reveals inactivating mutations in NOTCH1. Science 2011, 333:1154–1157.

74. Aref S, Rizk R, El Agder M, Fakhry W, El Zafarany M, Sabry M: NOTCH-1 Gene Mutations Influence Survival in Acute Myeloid Leukemia Patients. Asian Pac J Cancer Prev 2020, 21:1987–1992.

75. Gan RH, Wei H, Xie J, Zheng DP, Luo EL, Huang XY, Xie J, Zhao Y, Ding LC, Su BH, et al: Notch1 regulates tongue cancer cells proliferation, apoptosis and invasion. Cell Cycle 2018, 17:216–224.

76. Kujan O, Huang G, Ravindran A, Vijayan M, Farah CS: CDK4, CDK6, cyclin D1 and Notch1 immunocytochemical expression of oral brush liquid-based cytology for the diagnosis of oral leukoplakia and oral cancer. J Oral Pathol Med 2019, 48:566–573.

77. Ma Y, Li MD: Establishment of a Strong Link Between Smoking and Cancer Pathogenesis through DNA Methylation Analysis. Sci Rep 2017, 7:1811.

78. Ferrarotto R, Eckhardt G, Patnaik A, LoRusso P, Faoro L, Heymach JV, Kapoun AM, Xu L, Munster P: A phase I dose-escalation and dose-expansion study of brontictuzumab in subjects with selected solid tumors. Ann Oncol 2018, 29:1561–1568.

79. Moore G, Annett S, McClements L, Robson T: Top Notch Targeting Strategies in Cancer: A Detailed Overview of Recent Insights and Current Perspectives. Cells 2020, 9.

80. Wilzén A, Nilsson S, Sjöberg RM, Kogner P, Martinsson T, Abel F: The Phox2 pathway is differentially expressed in neuroblastoma tumors, but no mutations were found in the candidate tumor suppressor gene PHOX2A. Int J Oncol 2009, 34:697–705.

81. Ashktorab H, Washington K, Zarnogi S, Shakoori A, Varma S, Lee E, Shokrani B, Laiyemo A, Brim H: Determination of distinctive hypomethylated genes in African American colorectal neoplastic lesions. Therap Adv Gastroenterol 2020, 13:1756284820905482.

82. Eshragh J, Dhruva A, Paul SM, Cooper BA, Mastick J, Hamolsky D, Levine JD, Miaskowski C, Kober KM: Associations Between Neurotransmitter Genes and Fatigue and Energy Levels in Women After Breast Cancer Surgery. J Pain Symptom Manage 2017, 53:67–84.e67.

83. Tsai SC, Sheen MC, Chen BH: Association between HLA-DQA1, HLA-DQB1 and oral cancer. Kaohsiung J Med Sci 2011, 27:441–445.

84. Kozuka R, Enomoto M, Sato-Matsubara M, Yoshida K, Motoyama H, Hagihara A, Fujii H, Uchida-Kobayashi S, Morikawa H, Tamori A, et al: Association between HLA-DQA1/DRB1 polymorphism and development of hepatocellular carcinoma during entecavir treatment. J Gastroenterol Hepatol 2019, 34:937–946.

85. Shim H, Park B, Shin HJ, Joo J, Yoon KA, Kim YM, Hayashi T, Tokunaga K, Kong SY, Kim JY: Protective association of HLA-DRB1*13:02, HLA-DRB1*04:06, and HLA-DQB1*06:04 alleles with cervical cancer in a Korean population. Hum Immunol 2019, 80:107–111.

86. Garza-González E, Bosques-Padilla FJ, Pérez-Pérez GI, Flores-Gutiérrez JP, Tijerina-Menchaca R: Association of gastric cancer, HLA-DQA1, and infection with Helicobacter pylori CagA+ and VacA+ in a Mexican population. J Gastroenterol 2004, 39:1138–1142.

87. Arrillaga-Romany I, Chi AS, Allen JE, Oster W, Wen PY, Batchelor TT: A phase 2 study of the first imipridone ONC201, a selective DRD2 antagonist for oncology, administered every three weeks in recurrent glioblastoma. Oncotarget 2017, 8:79298–79304.

88. Li J, Zhu S, Kozono D, Ng K, Futalan D, Shen Y, Akers JC, Steed T, Kushwaha D, Schlabach M, et al: Genome-wide shRNA screen revealed integrated mitogenic signaling between dopamine receptor D2 (DRD2) and epidermal growth factor receptor (EGFR) in glioblastoma. Oncotarget 2014, 5:882–893.

89. Lin GM, Chen YJ, Kuo DJ, Jaiteh LE, Wu YC, Lo TS, Li YH: Cancer incidence in patients with schizophrenia or bipolar disorder: a nationwide population-based study in Taiwan, 1997-2009. Schizophr Bull 2013, 39:407–416.

90. Morton LM, Wang SS, Bergen AW, Chatterjee N, Kvale P, Welch R, Yeager M, Hayes RB, Chanock SJ, Caporaso NE: DRD2 genetic variation in relation to smoking and obesity in the Prostate, Lung, Colorectal, and Ovarian Cancer Screening Trial. Pharmacogenet Genomics 2006, 16:901–910.

91. Tan Y, Sun R, Liu L, Yang D, Xiang Q, Li L, Tang J, Qiu Z, Peng W, Wang Y, et al: Tumor suppressor DRD2 facilitates M1 macrophages and restricts NF-κB signaling to trigger pyroptosis in breast cancer. Theranostics 2021, 11:5214–5231.

92. Madhukar NS, Khade PK, Huang L, Gayvert K, Galletti G, Stogniew M, Allen JE, Giannakakou P, Elemento O: A Bayesian machine learning approach for drug target identification using diverse data types. Nat Commun 2019, 10:5221.

93. Ma Y, Wang M, Yuan W, Su K, Li MD: The significant association of Taq1A genotypes in DRD2/ANKK1 with smoking cessation in a large-scale meta-analysis of Caucasian populations. Transl Psychiatry 2015, 5:e686.

94. Ma Y, Yuan W, Jiang X, Cui WY, Li MD: Updated findings of the association and functional studies of DRD2/ANKK1 variants with addictions. Mol Neurobiol 2015, 51:281–299.

95. Munafò MR, Timpson NJ, David SP, Ebrahim S, Lawlor DA: Association of the DRD2 gene Taq1A polymorphism and smoking behavior: a meta-analysis and new data. Nicotine Tob Res 2009, 11:64–76.

96. Ma Y, Yuan W, Cui W, Li MD: Meta-analysis reveals significant association of 3’-UTR VNTR in SLC6A3 with smoking cessation in Caucasian populations. Pharmacogenomics J 2016, 16:10–17.

97. Ma Y, Wen L, Cui W, Yuan W, Yang Z, Jiang K, Jiang X, Huo M, Sun Z, Han H, et al: Prevalence of Cigarette Smoking and Nicotine Dependence in Men and Women Residing in Two Provinces in China. Front Psychiatry 2017, 8:254.

98. Liu Q, Xu Y, Mao Y, Ma Y, Wang M, Han H, Cui W, Yuan W, Payne TJ, Xu Y, et al: Genetic and Epigenetic Analysis Revealing Variants in the NCAM1-TTC12-ANKK1-DRD2 Cluster Associated Significantly With Nicotine Dependence in Chinese Han Smokers. Nicotine Tob Res 2020, 22:1301–1309.

99. Bidwell LC, Karoly HC, Thayer RE, Claus ED, Bryan AD, Weiland BJ, YorkWilliams S, Hutchison KE: DRD2 promoter methylation and measures of alcohol reward: functional activation of reward circuits and clinical severity. Addict Biol 2019, 24:539–548.

100. Clarke TK, Adams MJ, Davies G, Howard DM, Hall LS, Padmanabhan S, Murray AD, Smith BH, Campbell A, Hayward C, et al: Genome-wide association study of alcohol consumption and genetic overlap with other health-related traits in UK Biobank (N=112 117). Mol Psychiatry 2017, 22:1376–1384.

101. Ma Y, Fan R, Li MD: Meta-Analysis Reveals Significant Association of the 3’-UTR VNTR in SLC6A3 with Alcohol Dependence. Alcohol Clin Exp Res 2016, 40:1443–1453.

102. Ernst J, Kheradpour P, Mikkelsen TS, Shoresh N, Ward LD, Epstein CB, Zhang X, Wang L, Issner R, Coyne M, et al: Mapping and analysis of chromatin state dynamics in nine human cell types. Nature 2011, 473:43–49.

103. Trynka G, Sandor C, Han B, Xu H, Stranger BE, Liu XS, Raychaudhuri S: Chromatin marks identify critical cell types for fine mapping complex trait variants. Nat Genet 2013, 45:124–130.

104. Farh KK, Marson A, Zhu J, Kleinewietfeld M, Housley WJ, Beik S, Shoresh N, Whitton H, Ryan RJ, Shishkin AA, et al: Genetic and epigenetic fine mapping of causal autoimmune disease variants. Nature 2015, 518:337–343.

105. Finucane HK, Bulik-Sullivan B, Gusev A, Trynka G, Reshef Y, Loh PR, Anttila V, Xu H, Zang C, Farh K, et al: Partitioning heritability by functional annotation using genome-wide association summary statistics. Nat Genet 2015, 47:1228–1235.

106. Maurano MT, Humbert R, Rynes E, Thurman RE, Haugen E, Wang H, Reynolds AP, Sandstrom R, Qu H, Brody J, et al: Systematic localization of common disease-associated variation in regulatory DNA. Science 2012, 337:1190–1195.

107. . Pickrell JK: Joint analysis of functional genomic data and genome-wide association studies of 18 human traits. Am J Hum Genet 2014, 94:559–573.

