## Supplemental Figures for "Integrated multi-omics data analysis identifies a novel genetics-risk gene of *IRF4* associated with prognosis of oral cavity cancer"

**Supplemental Figure S1.** Quantile-quantile (QQ) plot of the MAGMA-based gene-level association analysis on GWAS summary statistics.

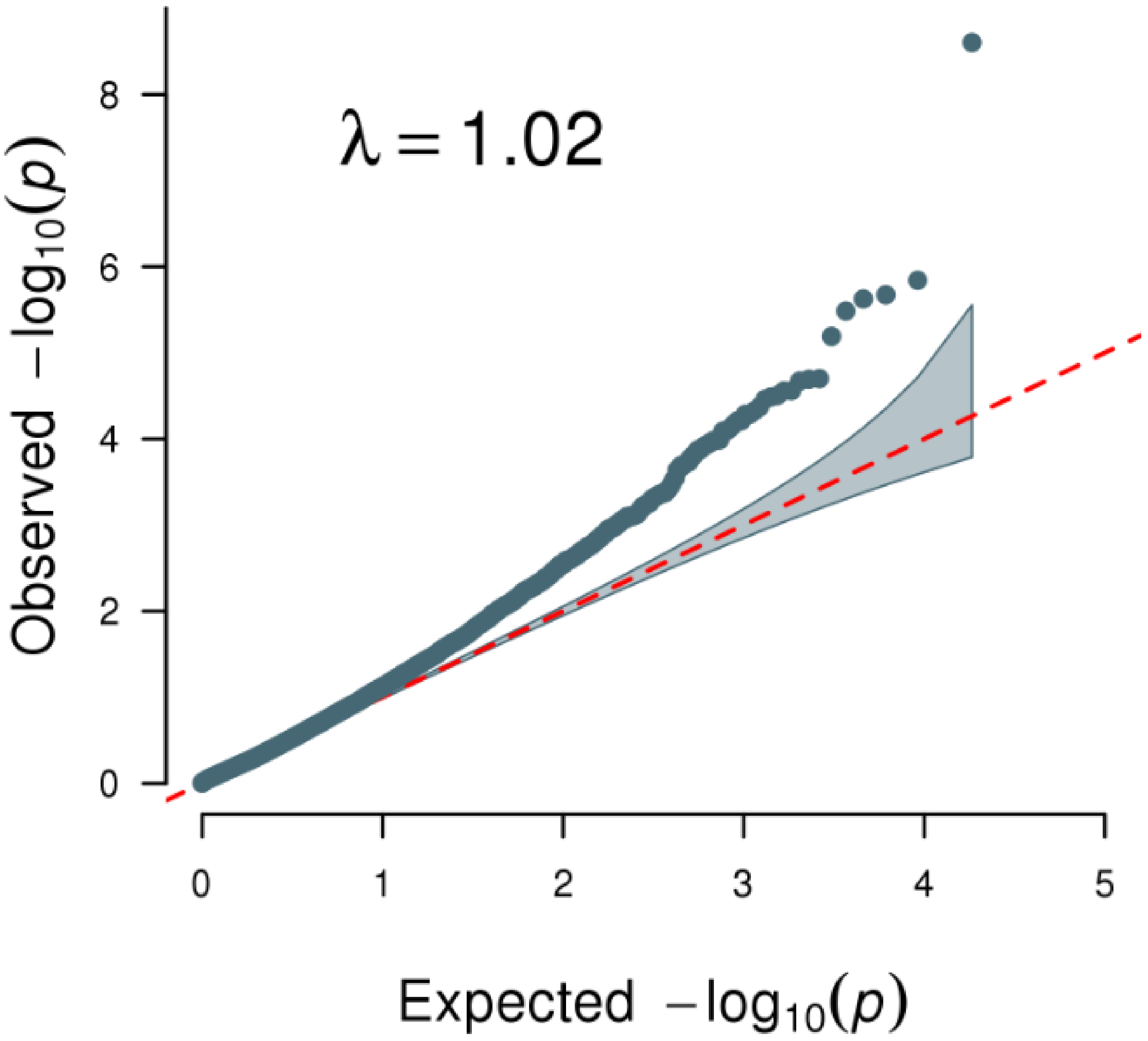

**Supplemental Figure S2.** The 15 lead SNPs with 14 genomic loci associated with OCC. Left panel shows the ID, P value, risk allele, and chromosome of each index SNP, and right panel shows the odds ratio with 95% confidence interval.

| CHR | SNP | Allele | P value |
|---|---|---|---|
| 5 | rs10462706 | C | 0.000187 |
| 6 | rs7773324 | G | 7.75e-07 |
| 6 | rs9271378 | A | 4.96e-06 |
| 7 | rs35894636 | G | 1.89e-05 |
| 7 | rs4717897 | A | 0.000147 |
| 8 | rs10283378 | T | 1.24e-05 |
| 9 | rs3125011 | G | 7.92e-07 |
| 11 | rs4586205 | G | 1.21e-05 |
| 15 | rs12916839 | C | 6.28e-07 |
| 16 | rs2397776 | T | 3.37e-05 |
| 16 | rs6499103 | C | 2.83e-05 |
| 16 | rs10775305 | A | 3.97e-06 |
| 19 | rs4806514 | C | 2.47e-05 |
| 20 | rs6051873 | T | 7.15e-06 |
| 20 | rs17711842 | C | 1.89e-05 |

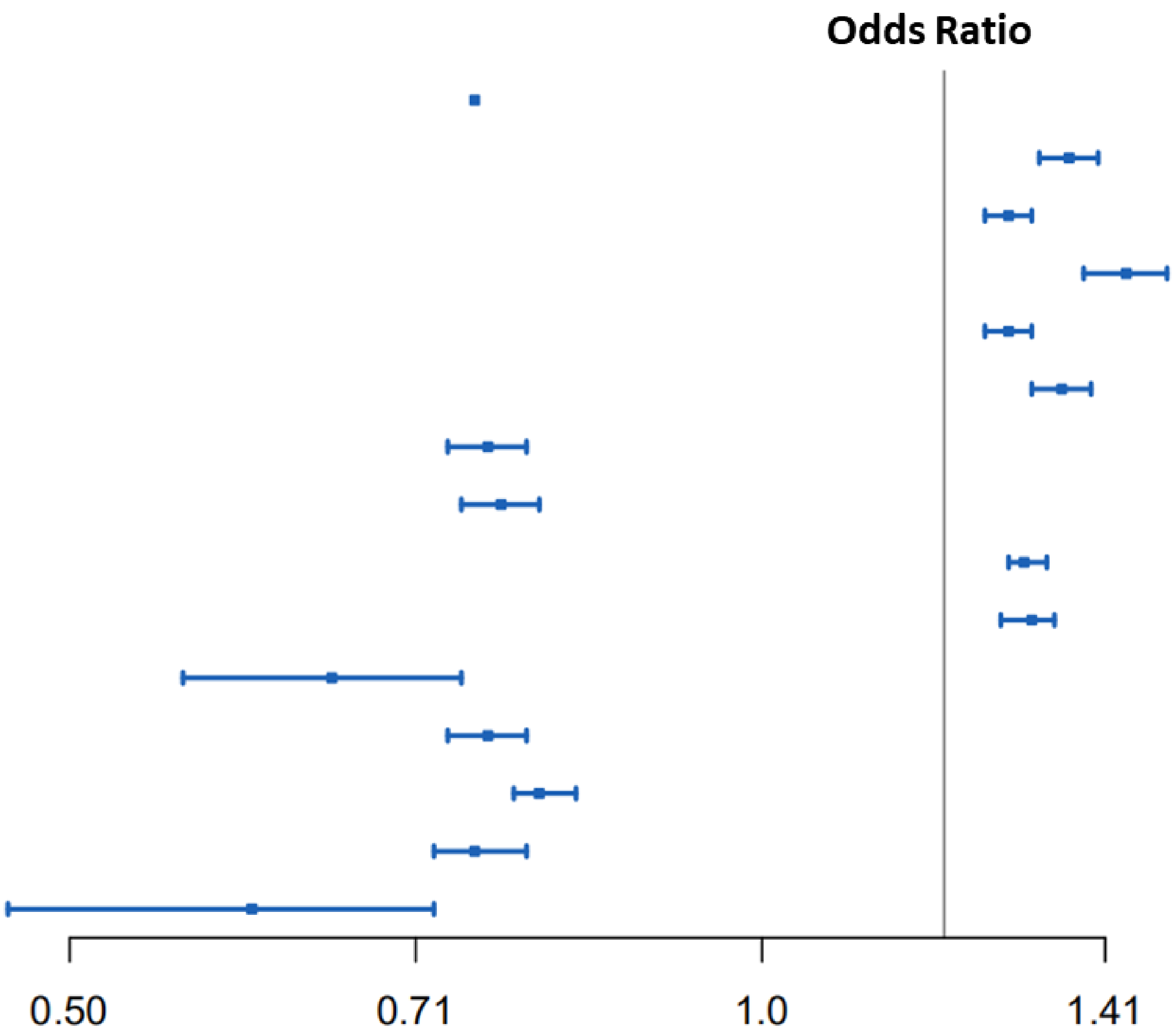


Supplemental Figure S2

**Supplemental Figure S3.** Regional plot for the OCC-associated risk gene of ZFP90 identified from MAGMA analysis. The purple diamond marks the most strongly associated SNP of rs10775305 in each gene with OCC. The color illustrates LD information with the given SNP, as shown in the color legend.

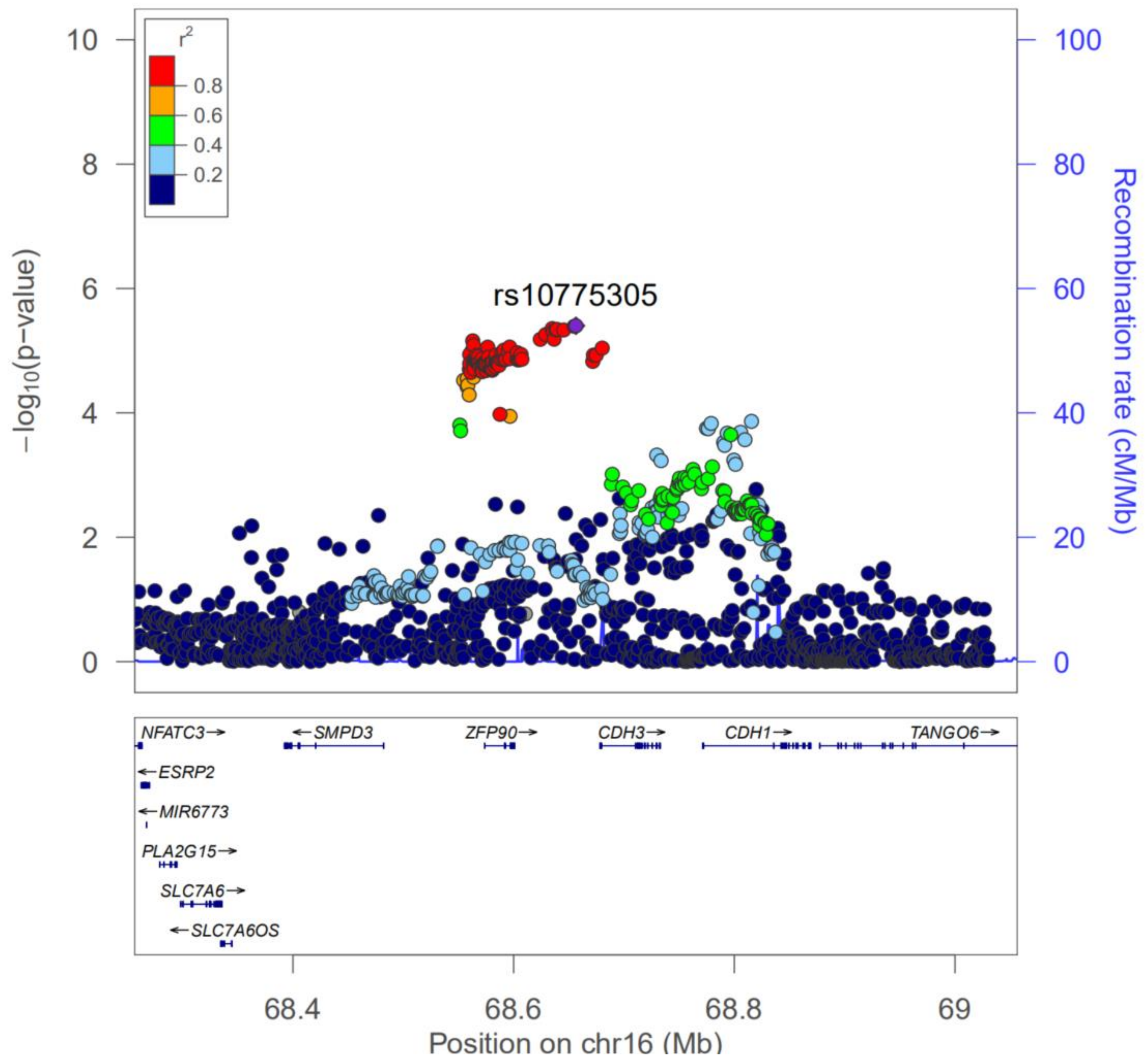

**Supplemental Figure S4.** Regional plot for the OCC-associated risk gene of *CLPTM1L* identified from MAGMA analysis. The purple diamond marks the most strongly associated SNP of rs10462706 in each gene with OCC. The color illustrates LD information with the given SNP, as shown in the color legend.

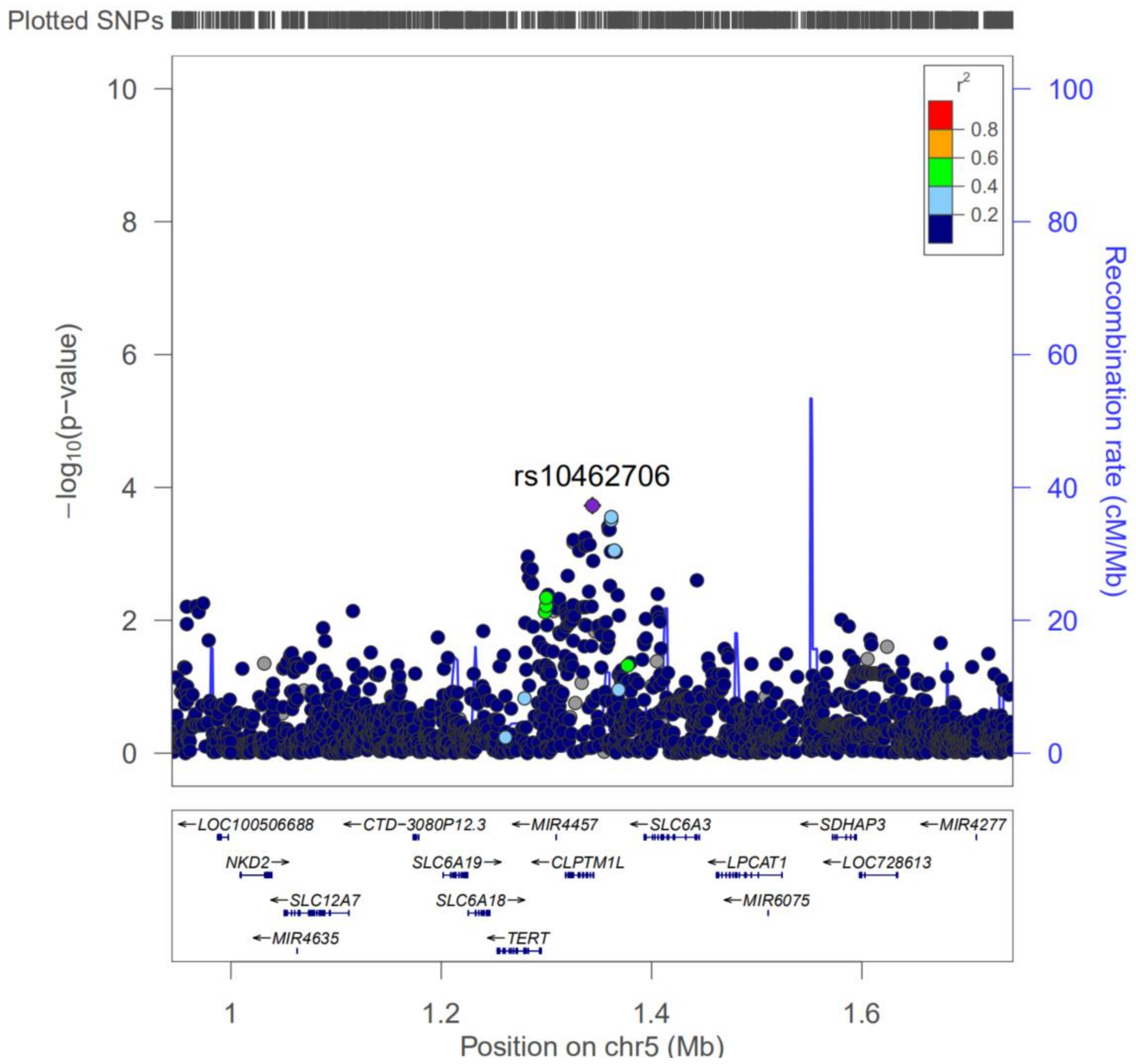


Supplemental Figure S4

**Supplemental Figure S5.** Regional plot for the OCC-associated risk gene of CDH16 identified from MAGMA analysis. The purple diamond marks the most strongly associated SNP of rs6499103 in each gene with OCC. The color illustrates LD information with the given SNP, as shown in the color legend.

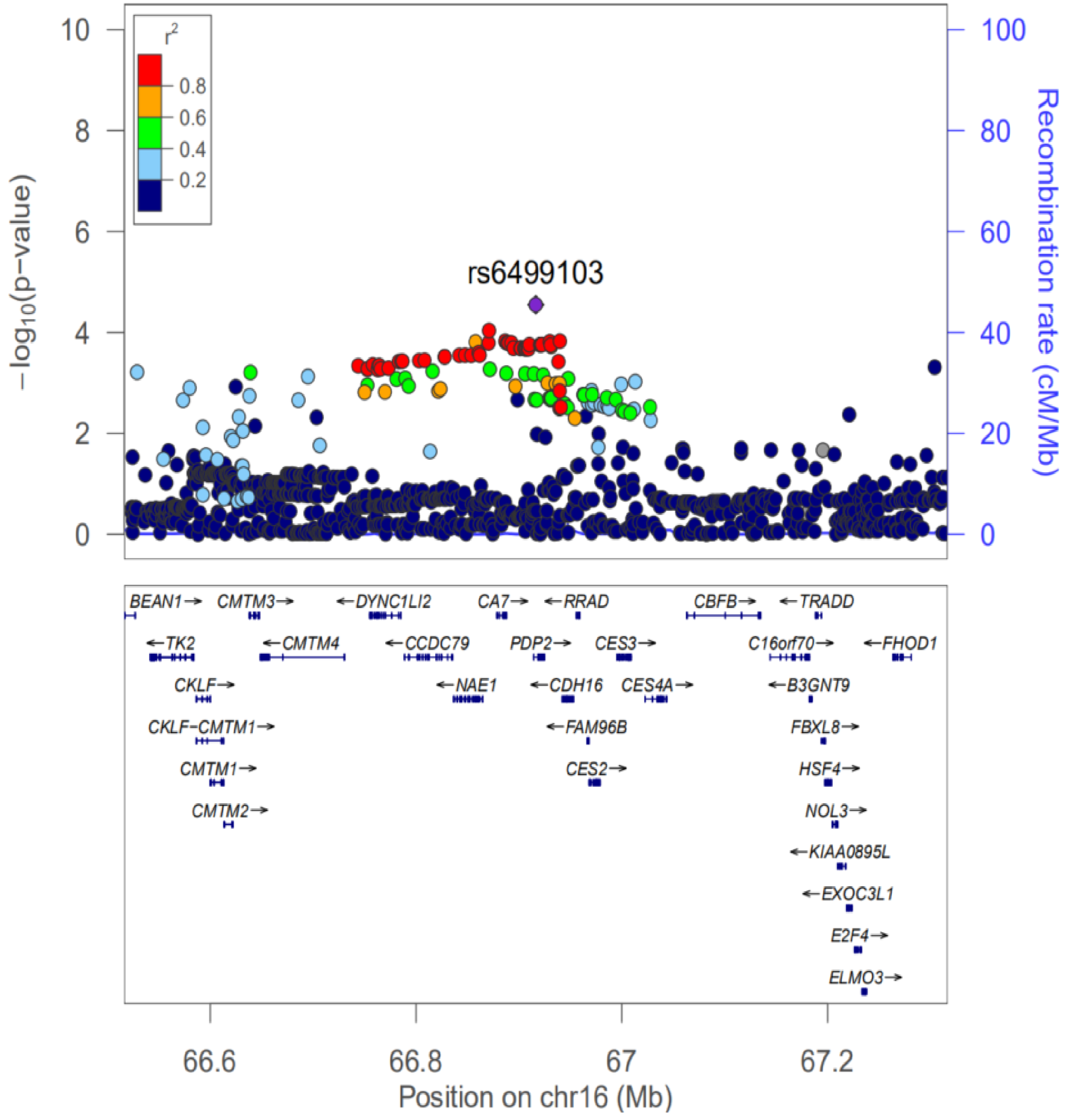

**Supplemental Figure S6**. Regional plot for the OCC-associated risk gene of DRD2 identified from MAGMA analysis. The purple diamond marks the most strongly associated SNP of rs4586205 in each gene with OCC. The color illustrates LD information with the given SNP, as shown in the color legend.

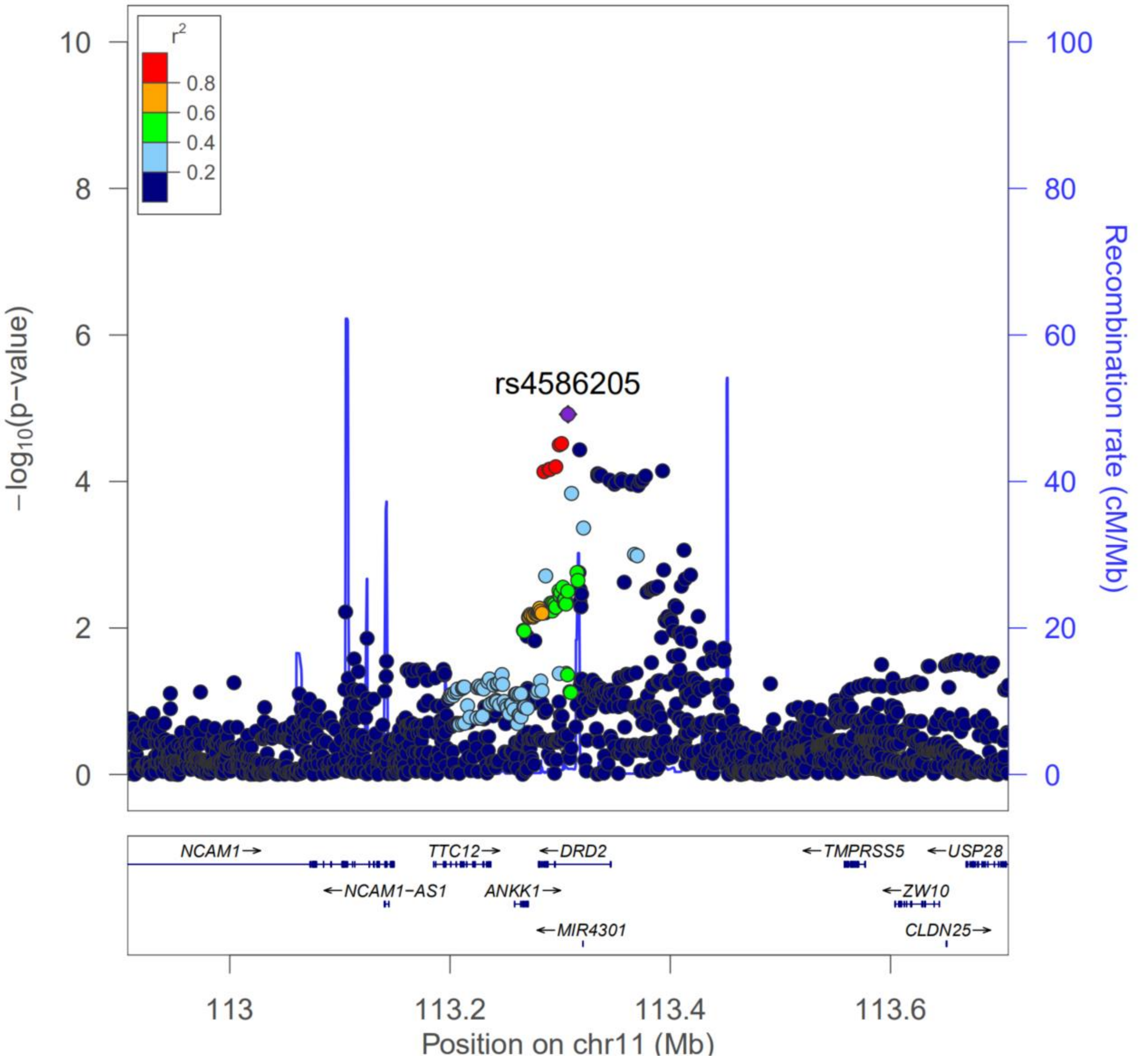

**Supplemental Figure S7.** Regional plot for the OCC-associated risk gene of EPHX2 identified from MAGMA analysis. The purple diamond marks the most strongly associated SNP of rs10283378 in each gene with OCC. The color illustrates LD information with the given SNP, as shown in the color legend.

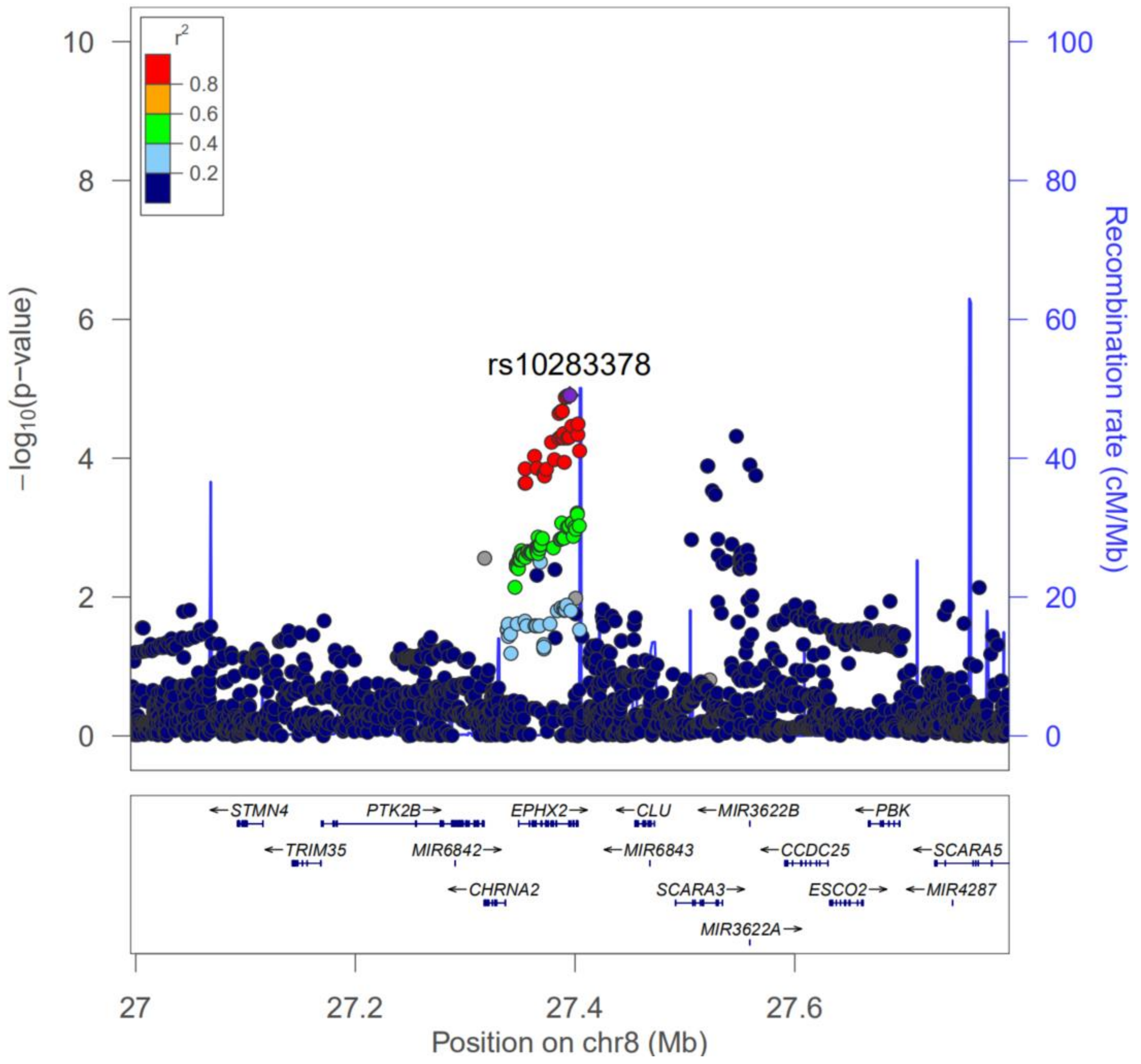

**Supplemental Figure S8.** Regional plot for the OCC-associated risk gene of HLA-DQA1 identified from MAGMA analysis. The purple diamond marks the most strongly associated SNP of rs9271378 in each gene with OCC. The color illustrates LD information with the given SNP, as shown in the color legend.

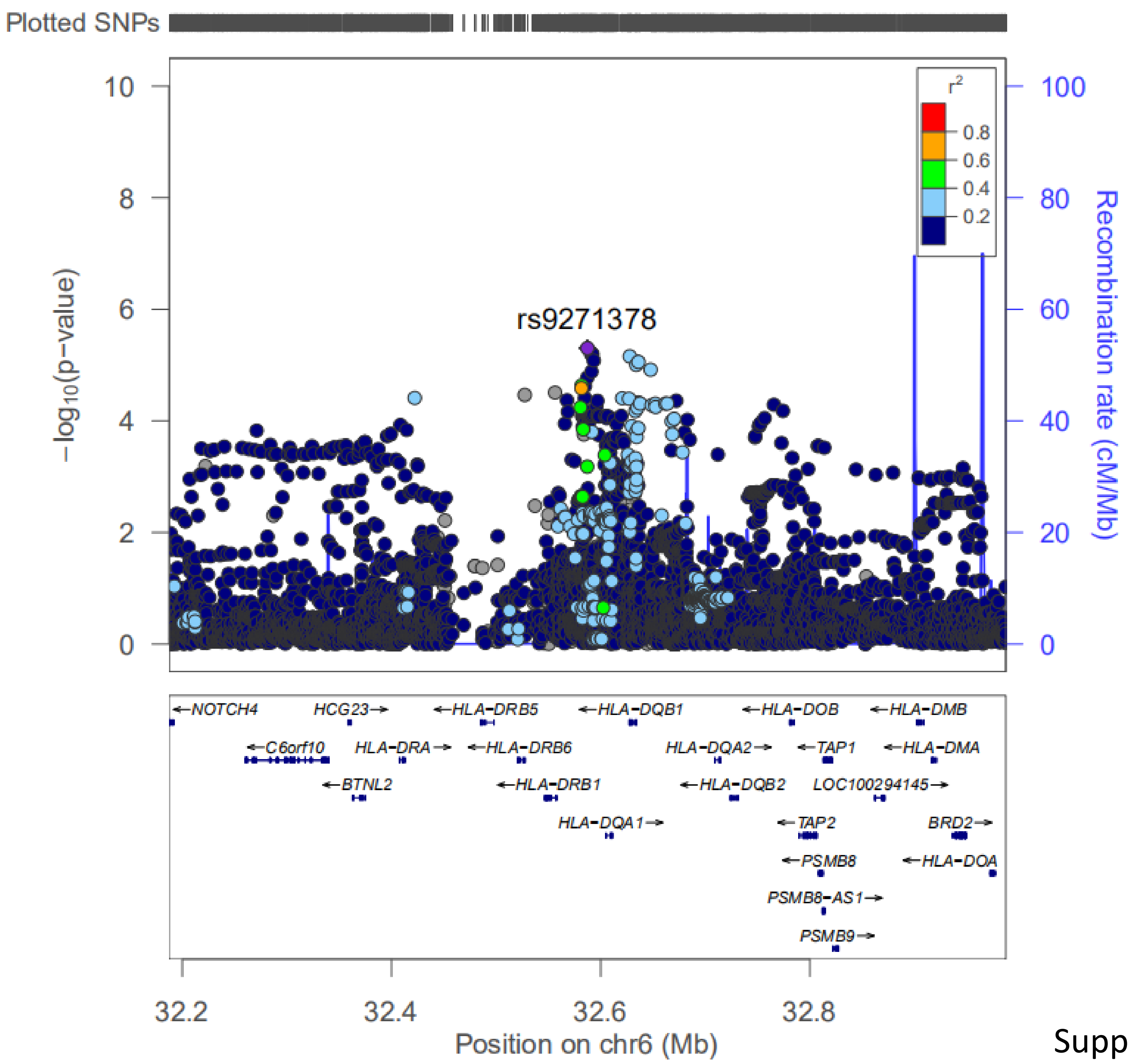


Supplemental Figure S8

**Supplemental Figure S9**. Regional plot for the OCC-associated risk gene of TNS3 identified from MAGMA analysis. The purple diamond marks the most strongly associated SNP of rs35894636 in each gene with OCC. The color illustrates LD information with the given SNP, as shown in the color legend.

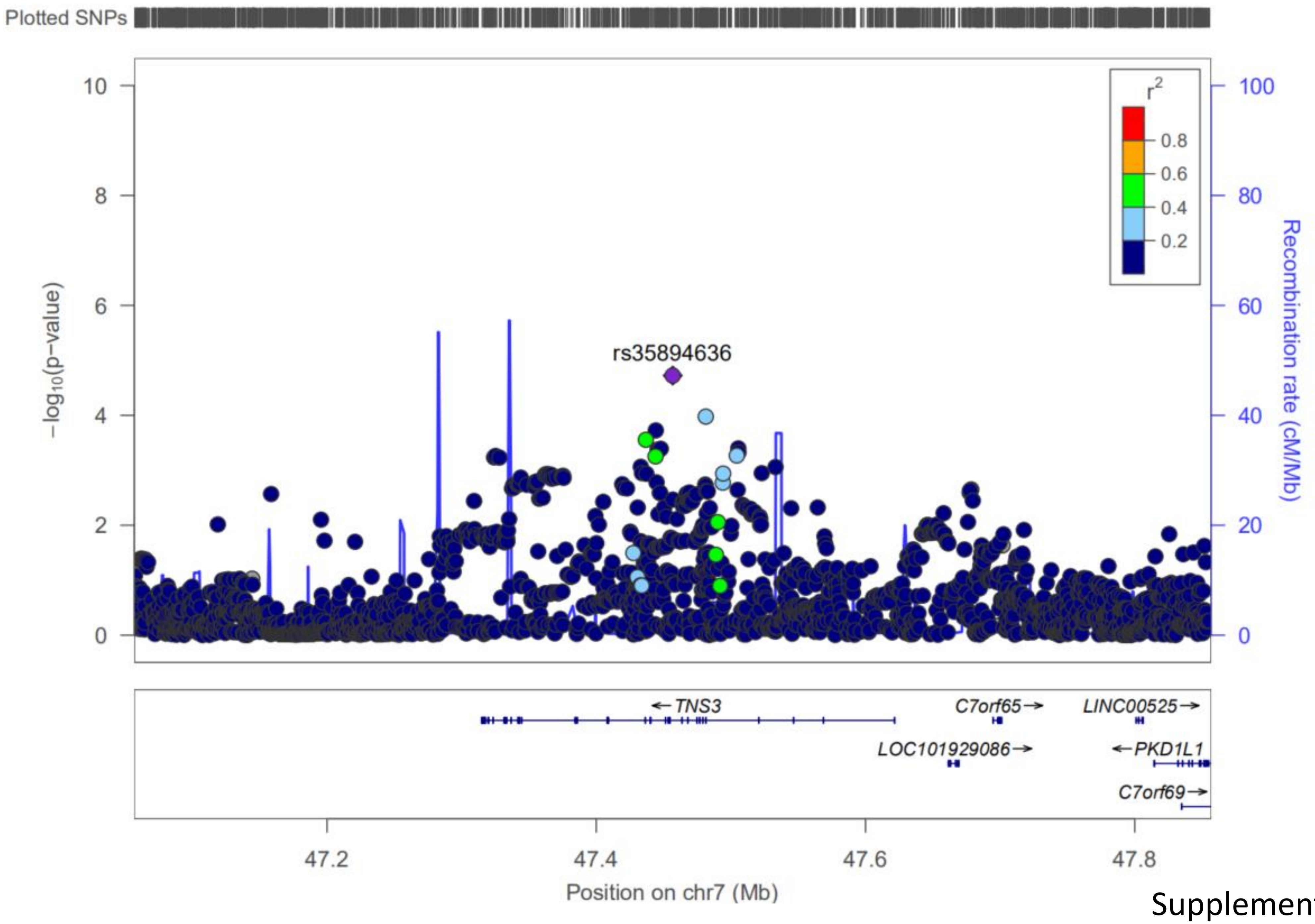


Supplemental Figure S9

**Supplemental Figure S10.** Regional plot for the OCC-associated risk gene of GTF2IRD1 identified from MAGMA analysis. The purple diamond marks the most strongly associated SNP of rs4717897 in each gene with OCC. The color illustrates LD information with the given SNP, as shown in the color legend.

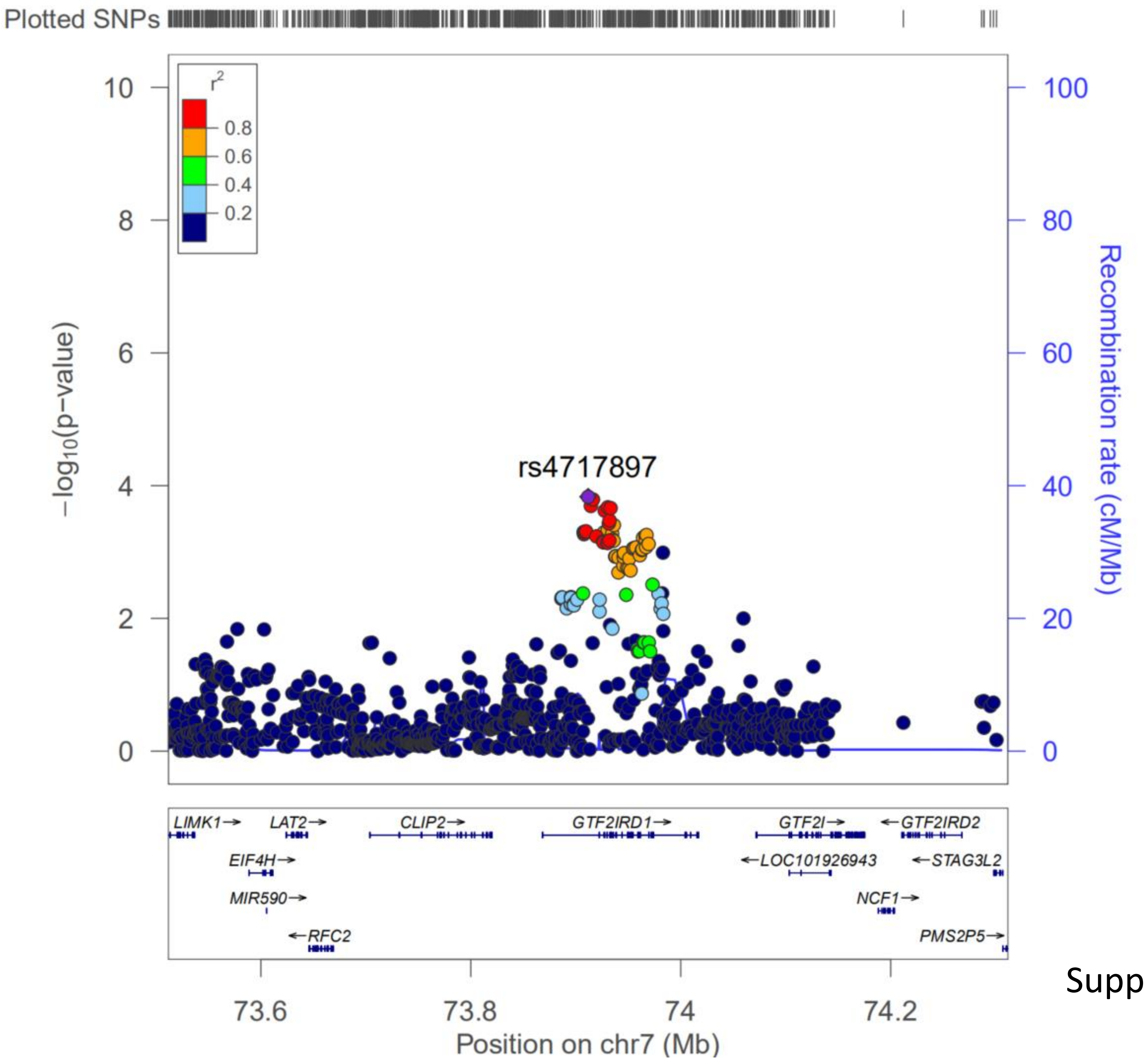


Supplemental Figure S10

**Supplemental Figure S11.** Regional plot for the OCC-associated risk gene of NOTCH1 identified from MAGMA analysis. The purple diamond marks the most strongly associated SNP of rs3125011 in each gene with OCC. The color illustrates LD information with the given SNP, as shown in the color legend.

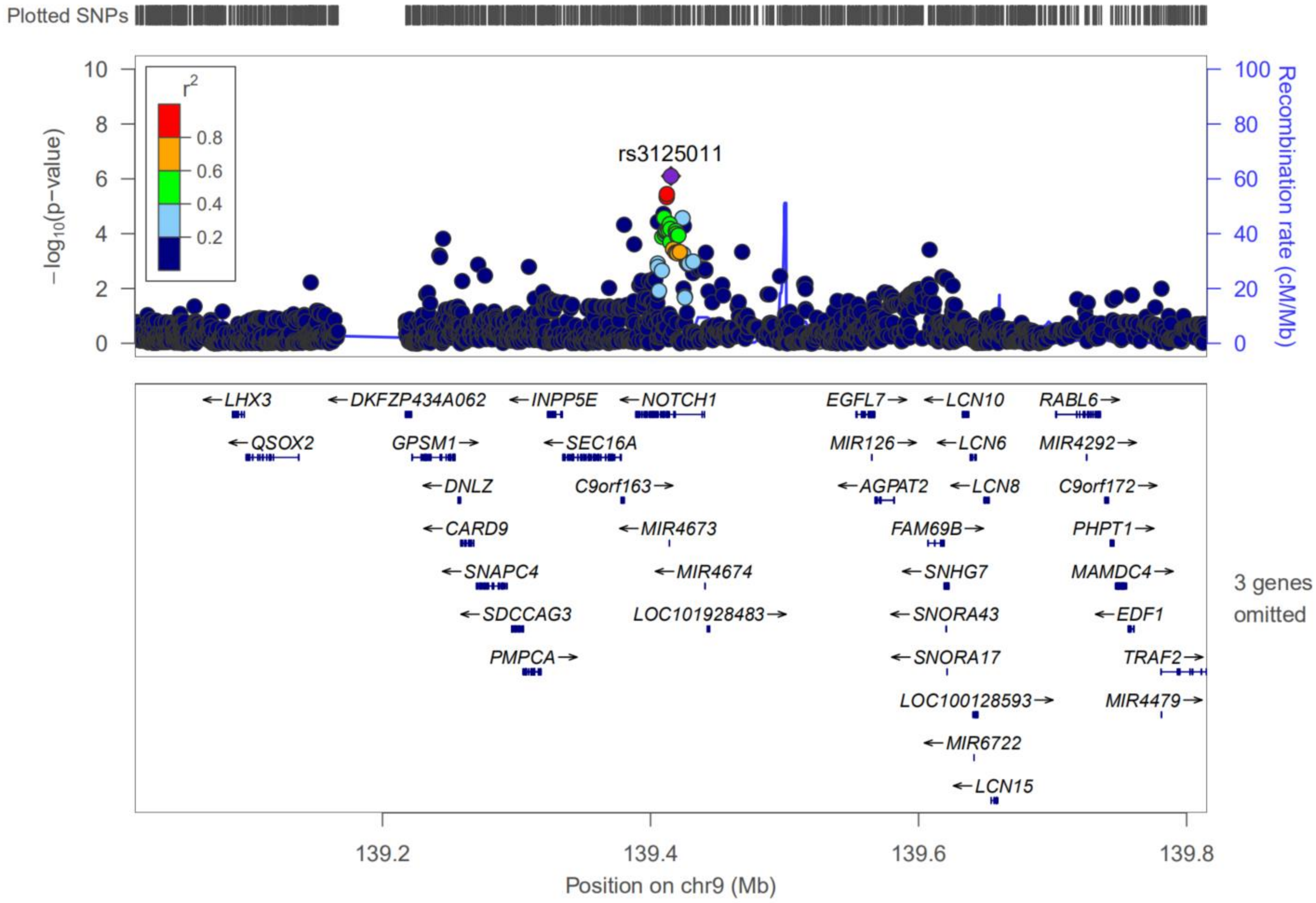


Supplemental Figure S11

**Supplemental Figure S12.** Regional plot for the OCC-associated risk gene of FGF7 identified from MAGMA analysis. The purple diamond marks the most strongly associated SNP of rs12916839 in each gene with OCC. The color illustrates LD information with the given SNP, as shown in the color legend.

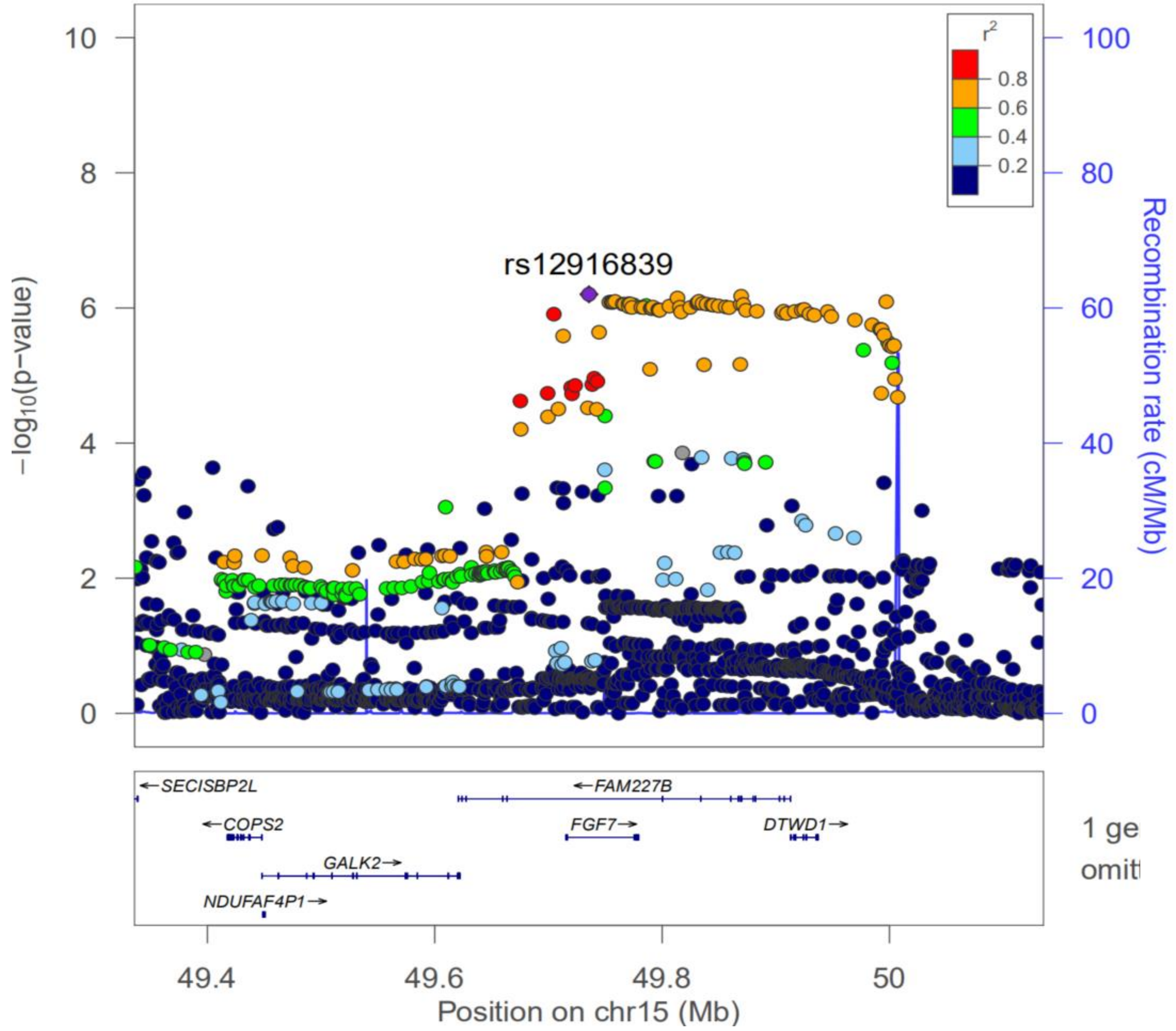


Supplemental Figure S12

**Supplemental Figure S13.** Regional plot for the OCC-associated risk gene of SLC6A2 identified from MAGMA analysis. The purple diamond marks the most strongly associated SNP of rs2397776 in each gene with OCC. The color illustrates LD information with the given SNP, as shown in the color legend.

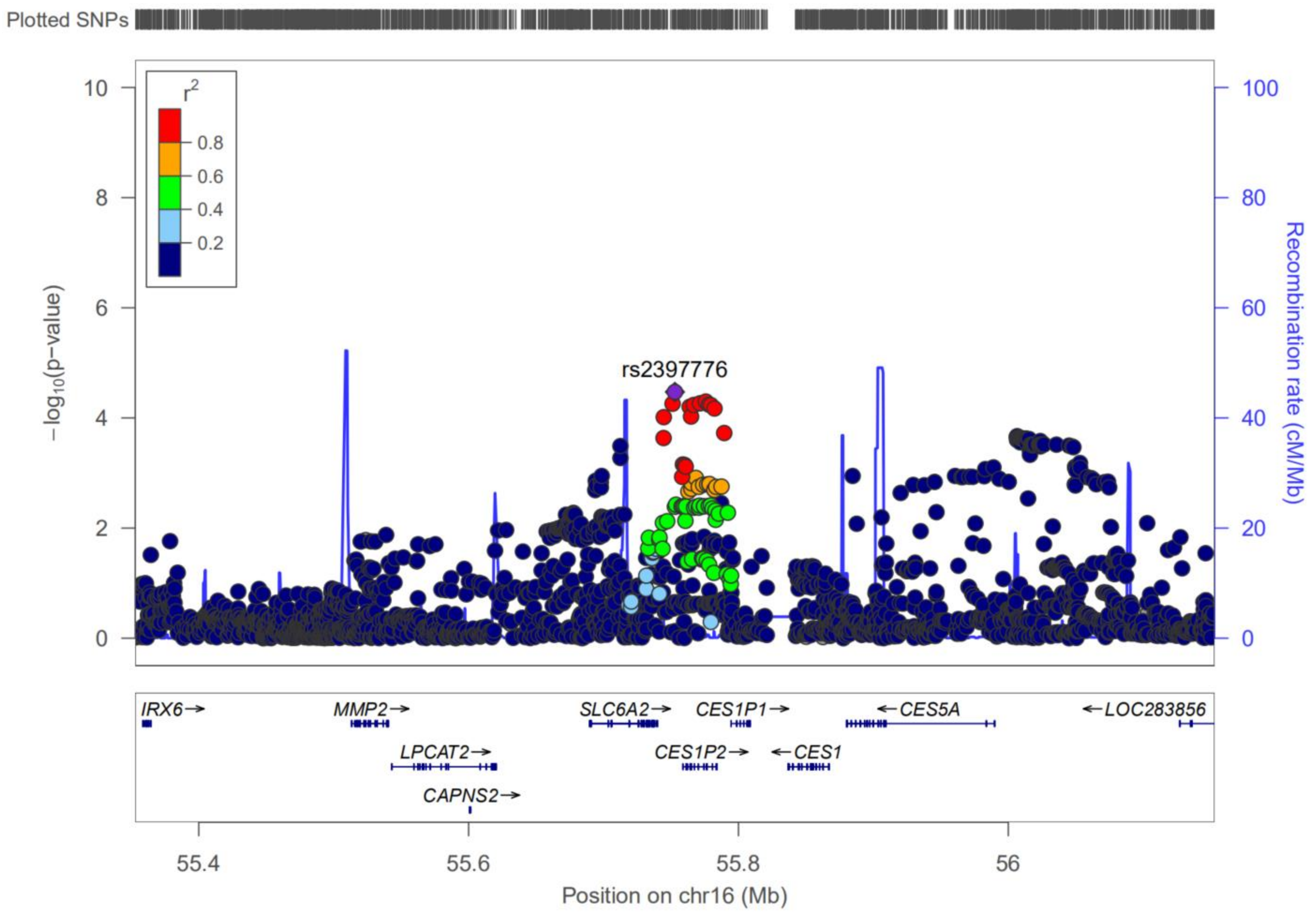


Supplemental Figure S13

**Supplemental Figure S14.** Regional plot for the OCC-associated risk gene of NLRP12 identified from MAGMA analysis. The purple diamond marks the most strongly associated SNP of rs4806514 in each gene with OCC. The color illustrates LD information with the given SNP, as shown in the color legend.

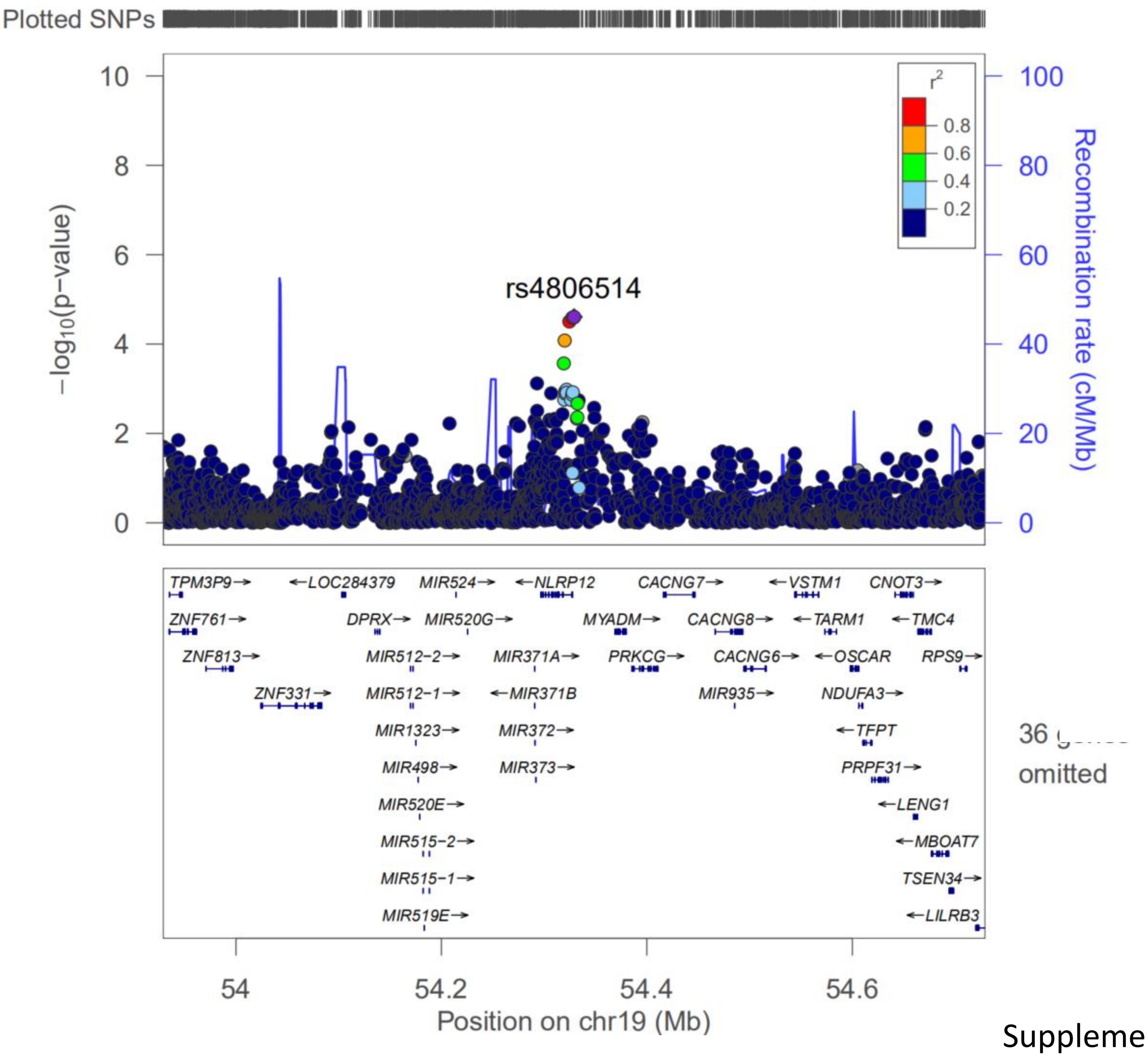


Supplemental Figure S14

**Supplemental Figure S15.** Regional plot for the OCC-associated risk gene of ATRN identified from MAGMA analysis. The purple diamond marks the most strongly associated SNP of rs6051873 in each gene with OCC. The color illustrates LD information with the given SNP, as shown in the color legend.

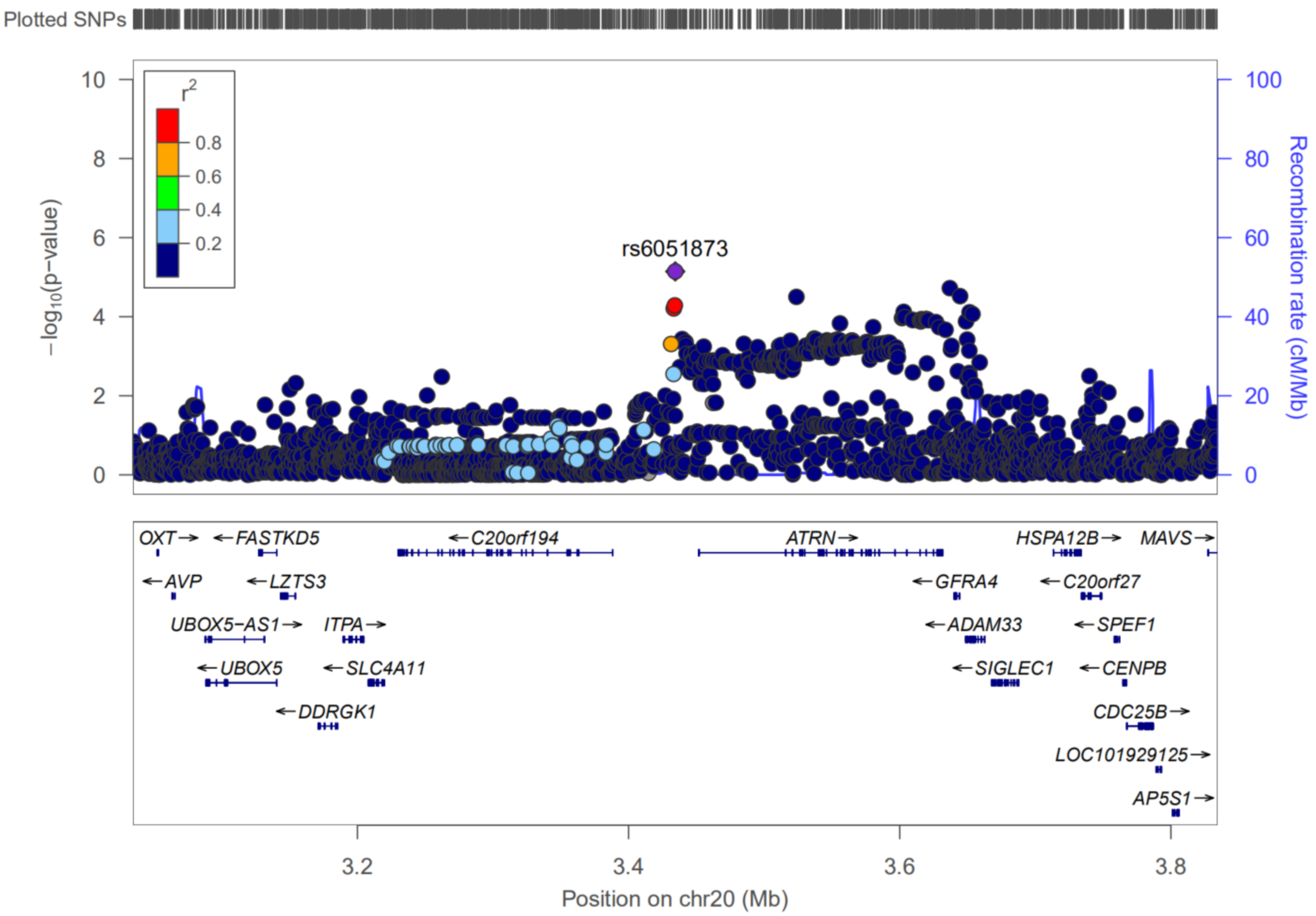


Supplemental Figure S15

**Supplemental Figure S16.** Regional plot for the OCC-associated risk gene of ATRN identified from MAGMA analysis. The purple diamond marks the most strongly associated SNP of rs17711842 in each gene with OCC. The color illustrates LD information with the given SNP, as shown in the color legend.

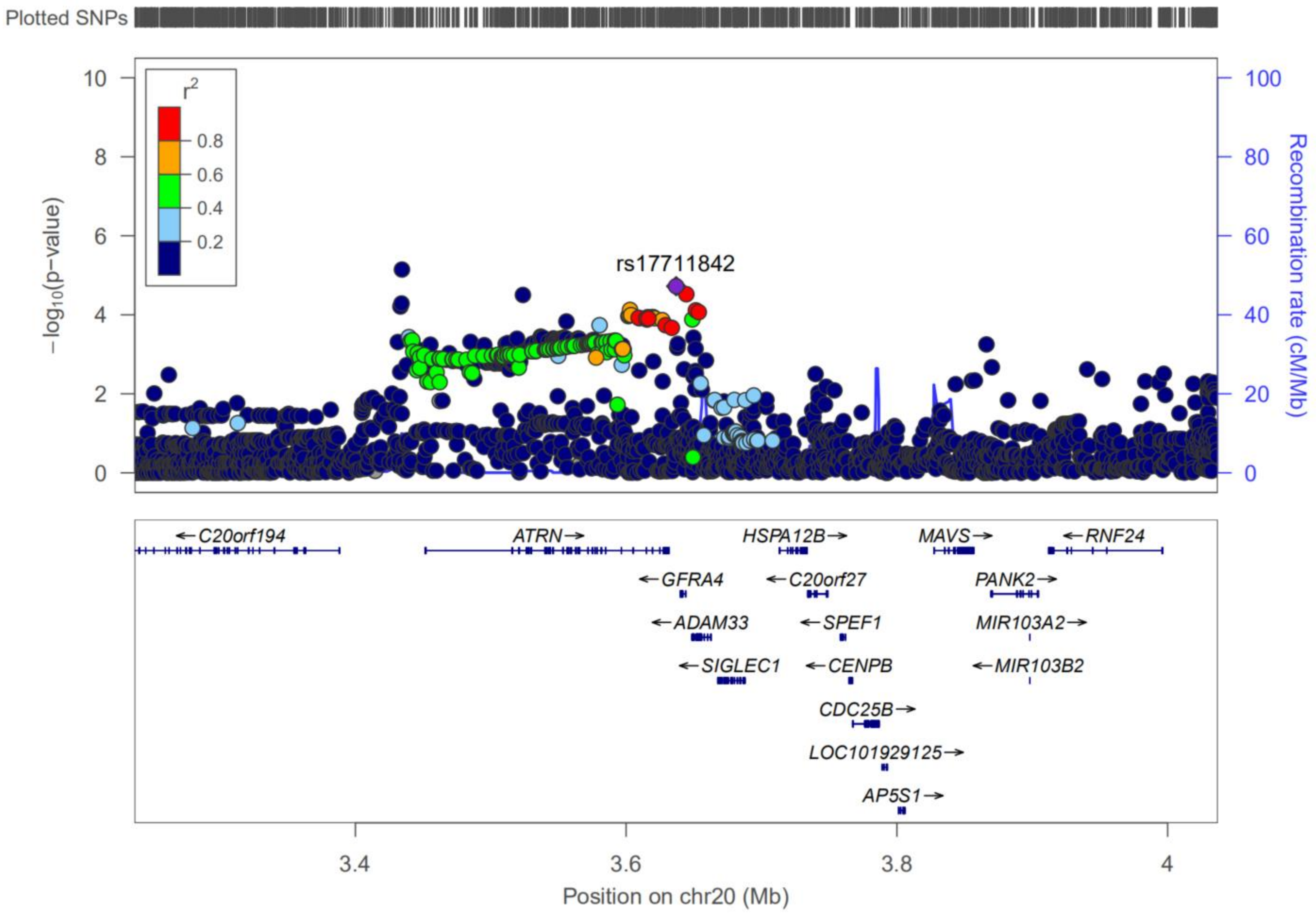

**Supplemental Figure S17.** Multiple layers of evidence supporting the functional role of rs7773324 in IRF4. There were three distinct data used for assessing the variant functionality, including CADD score, regulomeDB, and chromatin state. This plot was generated by using the FUMA online tool (https://fuma.ctglab.nl/).

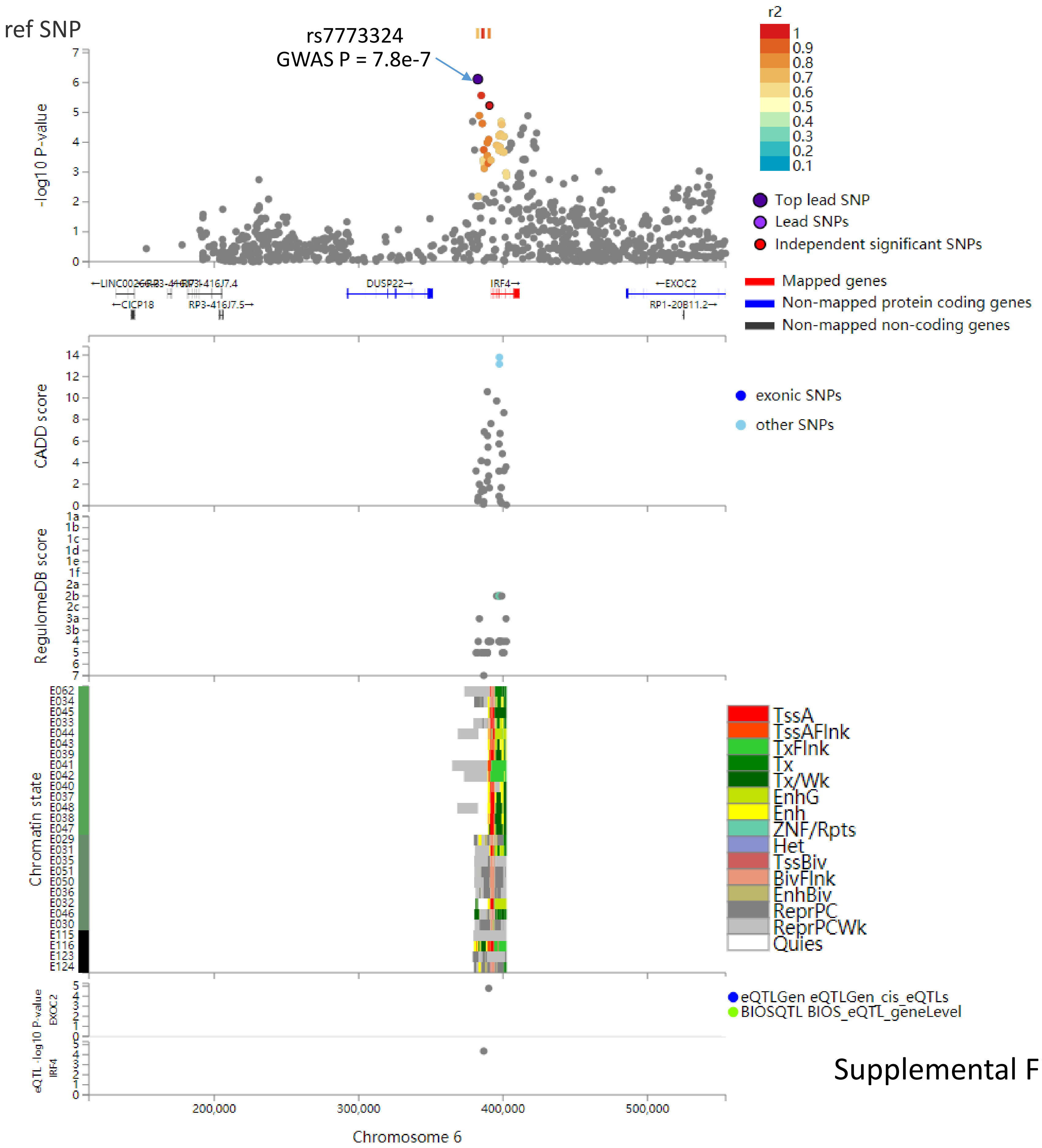


Supplemental Figure S17

**Supplemental Figure S18.** Multiple layers of evidence supporting the functional role of rs3125011 in NOTCH1. There were three distinct data used for assessing the variant functionality, including CADD score, regulomeDB, and chromatin state. This plot was generated by using the FUMA online tool (https://fuma.ctglab.nl/).

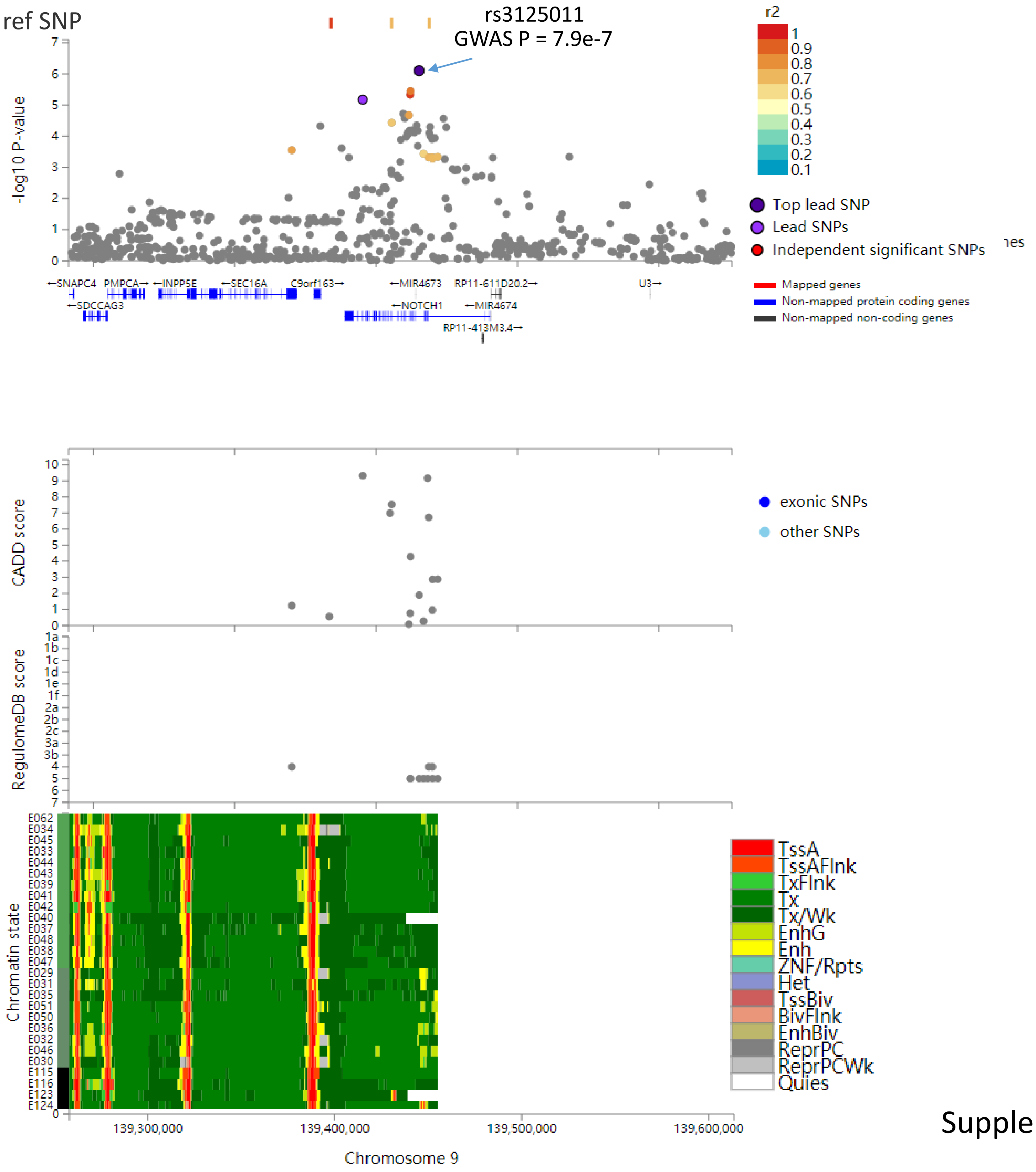


Supplemental Figure S18

**Supplemental Figure S19.** Multiple layers of evidence supporting the functional role of rs12916839 in FGF7. There were three distinct data used for assessing the variant functionality, including CADD score, regulomeDB, and chromatin state. This plot was generated by using the FUMA online tool (https://fuma.ctglab.nl/).

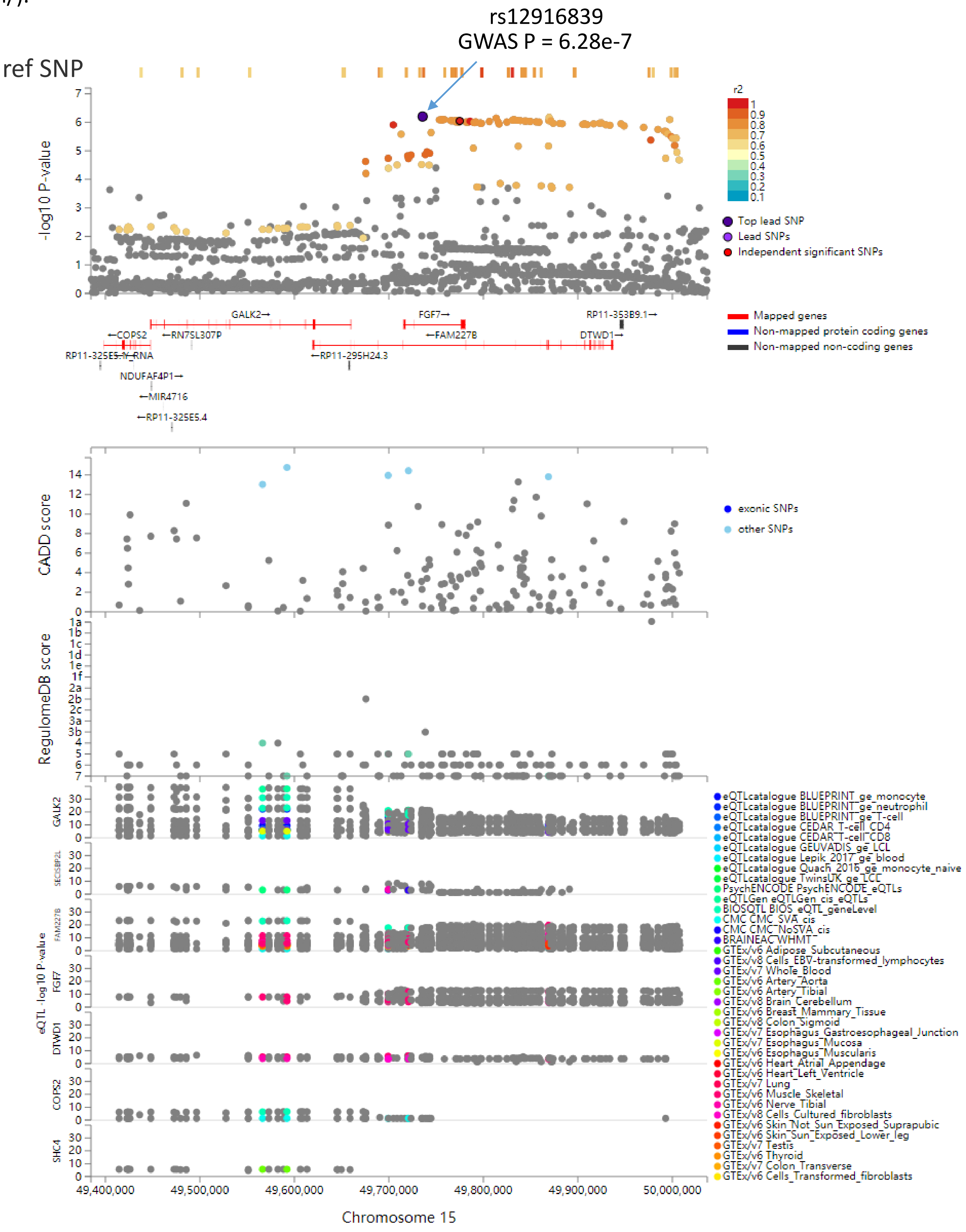


Supplemental Figure S19

**Supplemental Figure S20.** Violin plots showing rs12916839 influences the expression of FGF7 across multiple tissues. A)-B) Violin plots showing rs12916839 represents an eQTL quantitative trait locus for FGF7 across multiple GTEx tissues. C)-I) Violin plots showing rs12916839 represents a splicing quantitative trait locus for FGF7 across multiple GTEx tissues.

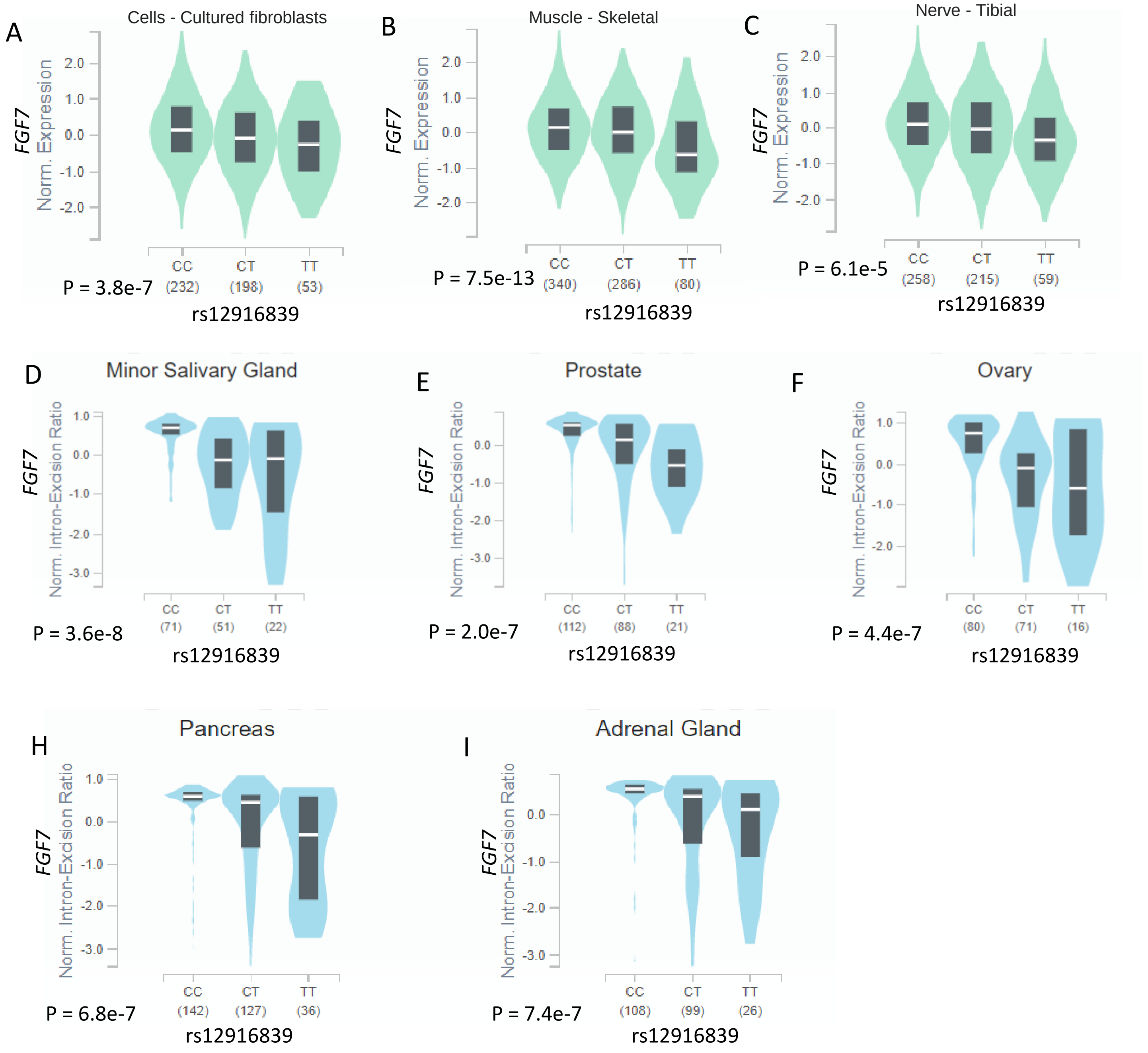


Supplemental Figure S20

**Supplemental Figure S21.** Violin plots showing rs12916839 influences the expression of FGF7 across multiple tissues. For A)-I), there were 9 tissues showing a significant sQTL for rs12916839.

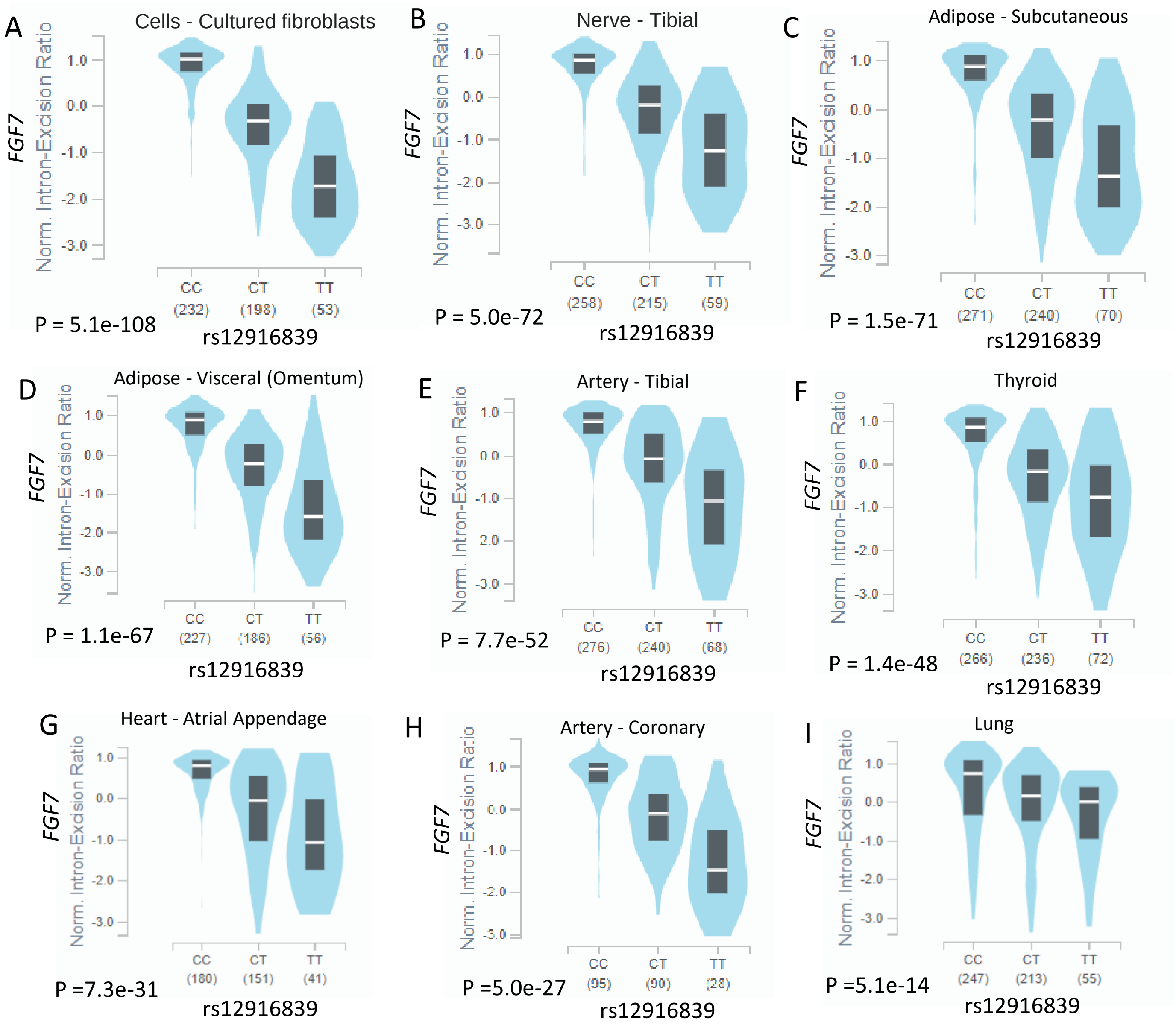


Supplemental Figure S21

**Supplemental Figure S22.** Violin plots showing rs12916839 represents a splicing quantitative trait locus for FGF7 across multiple GTEx tissues. For A)-L), there were 12 tissues showing a significant sQTL for rs12916839.

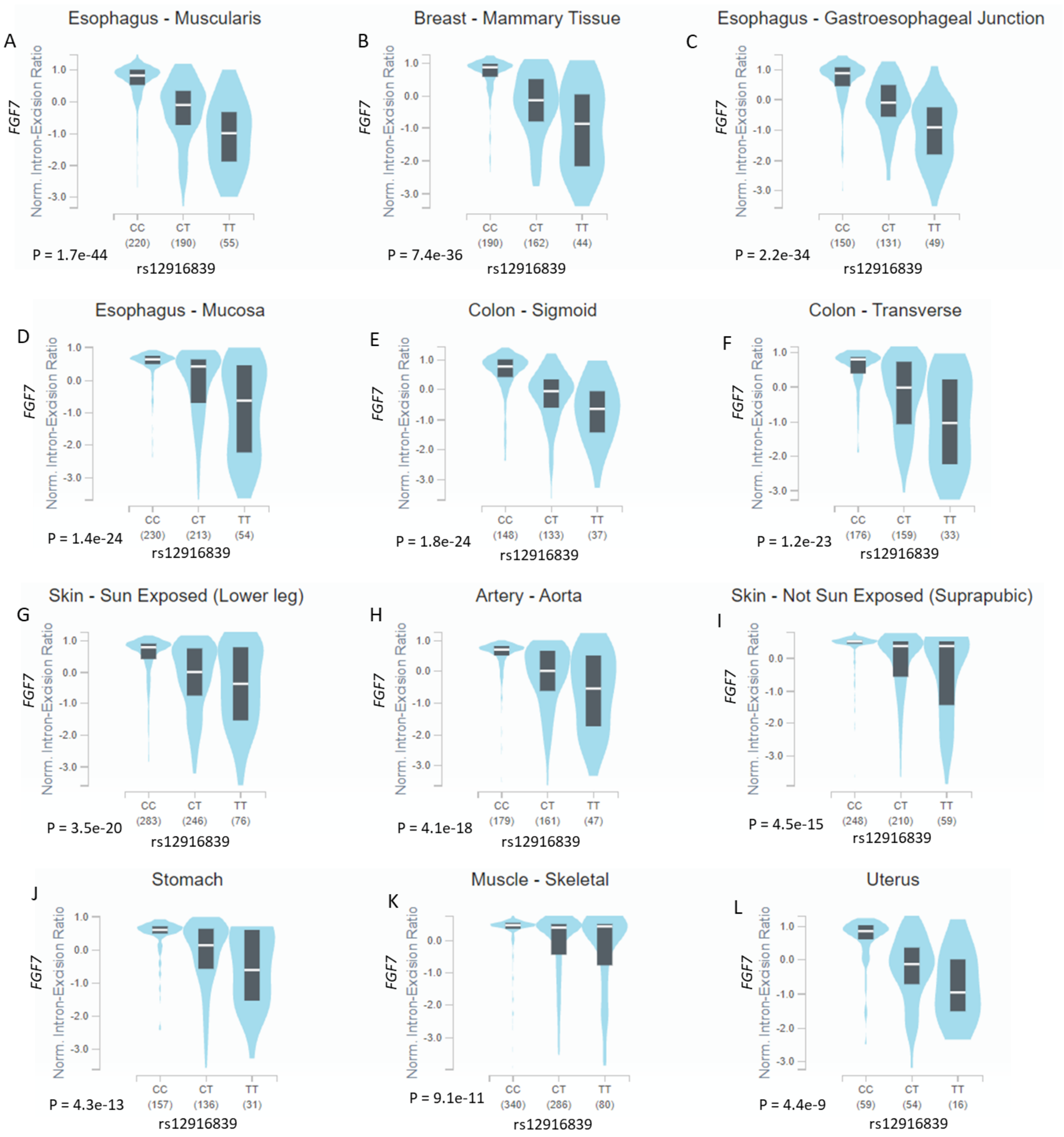


Supplemental Figure S22

**Supplemental Figure S23.** S-PrediXcan-based integrative genomics analysis identified risk genes associated with oral cancer in each tissue. A) Whole blood, B) Thyroid, C) Skin sun exposed lower leg, D) Skin not sun exposed suprapubic, E) Muscle skeletal, and F) Minor salivary gland.

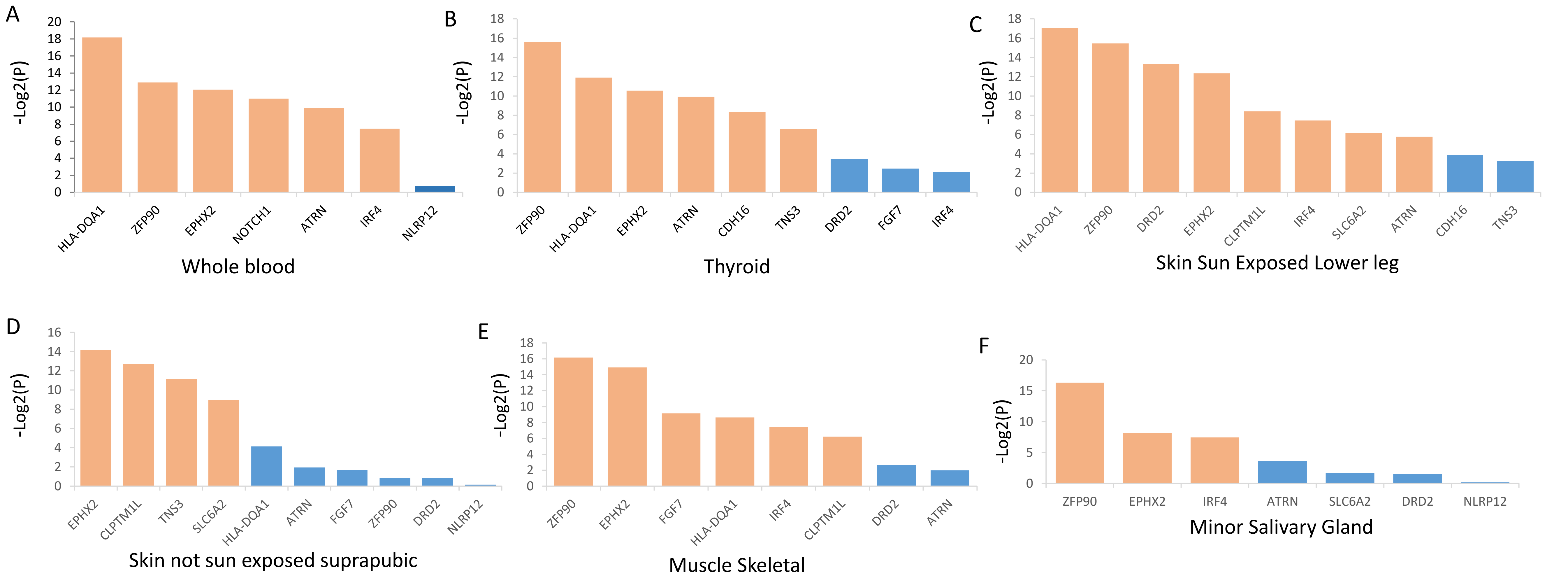


Supplemental Figure S23

**Supplemental Figure S24.** S-PrediXcan-based integrative genomics analysis identified risk genes associated with oral cancer in each tissue. A) Liver, B) Lung, C) Esophagus Mucosa, D) Adrenal Gland, E) Adipose Visceral Omentum, and F) Adipose Subcutaneous.

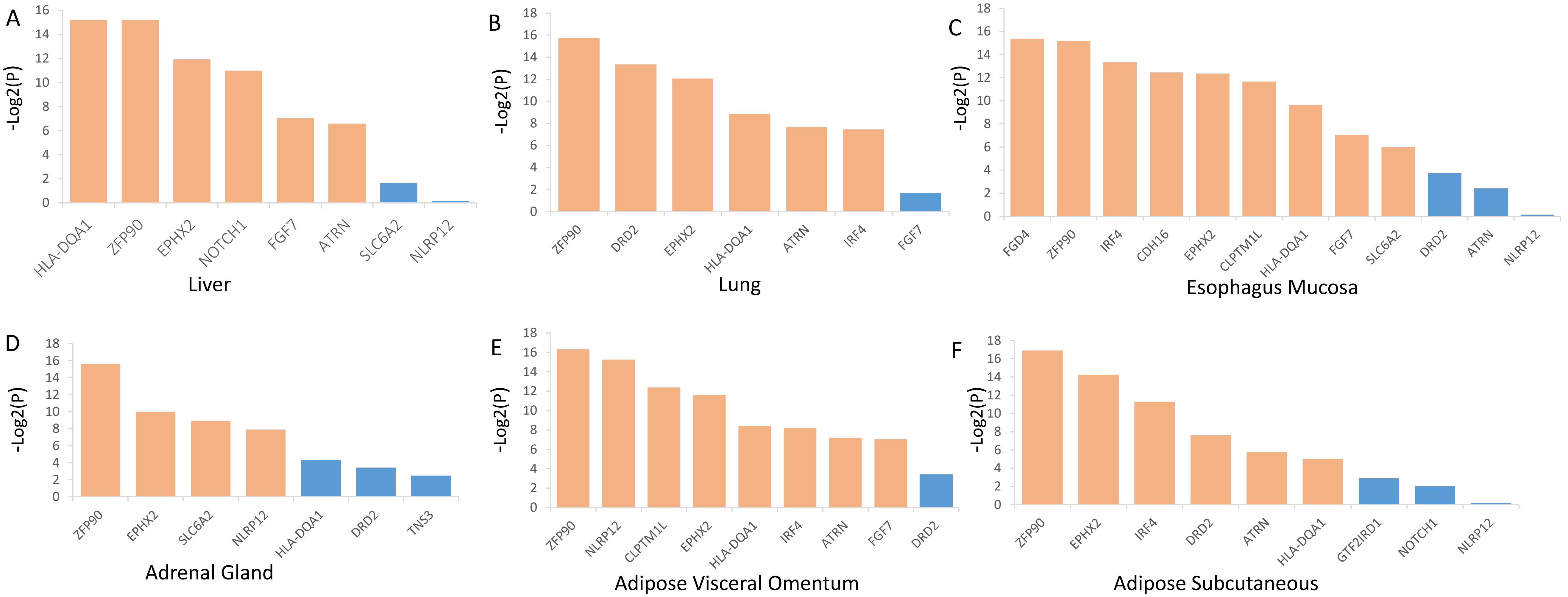


Supplemental Figure S24

**Supplemental Figure S25.** S-PrediXcan-based integrative genomics analysis identified risk genes associated with oral cancer in each tissue. A) Esophagus_Gastroesophageal_Junction, B) Esophagus Muscularis, C) Artery Aorta, D) Artery Coronary, E) Cells- EBV-transformed lymphocytes, and F) Cells-Cultured fibroblasts.

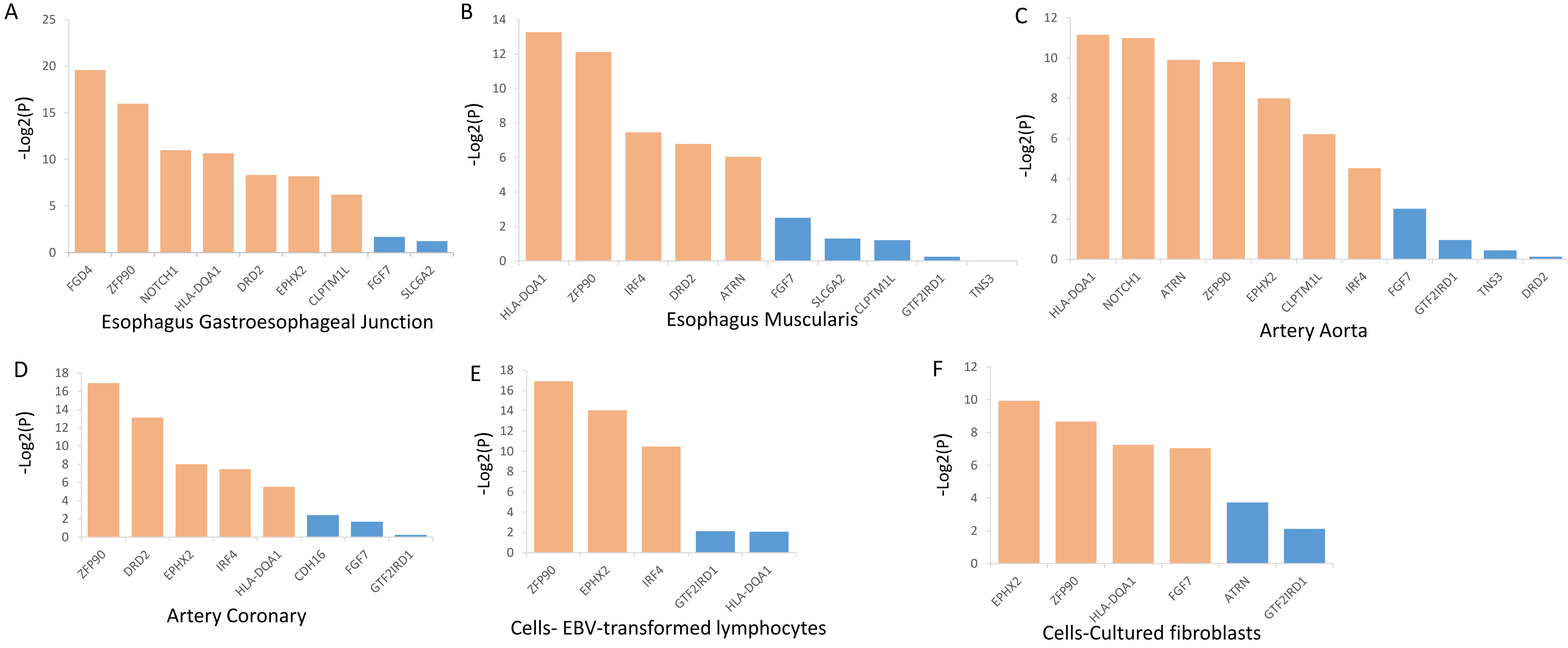


Supplemental Figure S25

**Supplemental Figure S26.** S-PrediXcan-based integrative genomics analysis identified risk genes associated with oral cancer in each tissue. A) Colon Transverse , B) Pancreas, C) Colon Sigmoid, D) Pituitary, E) Small intestine terminal ileum, and F) Stomach.

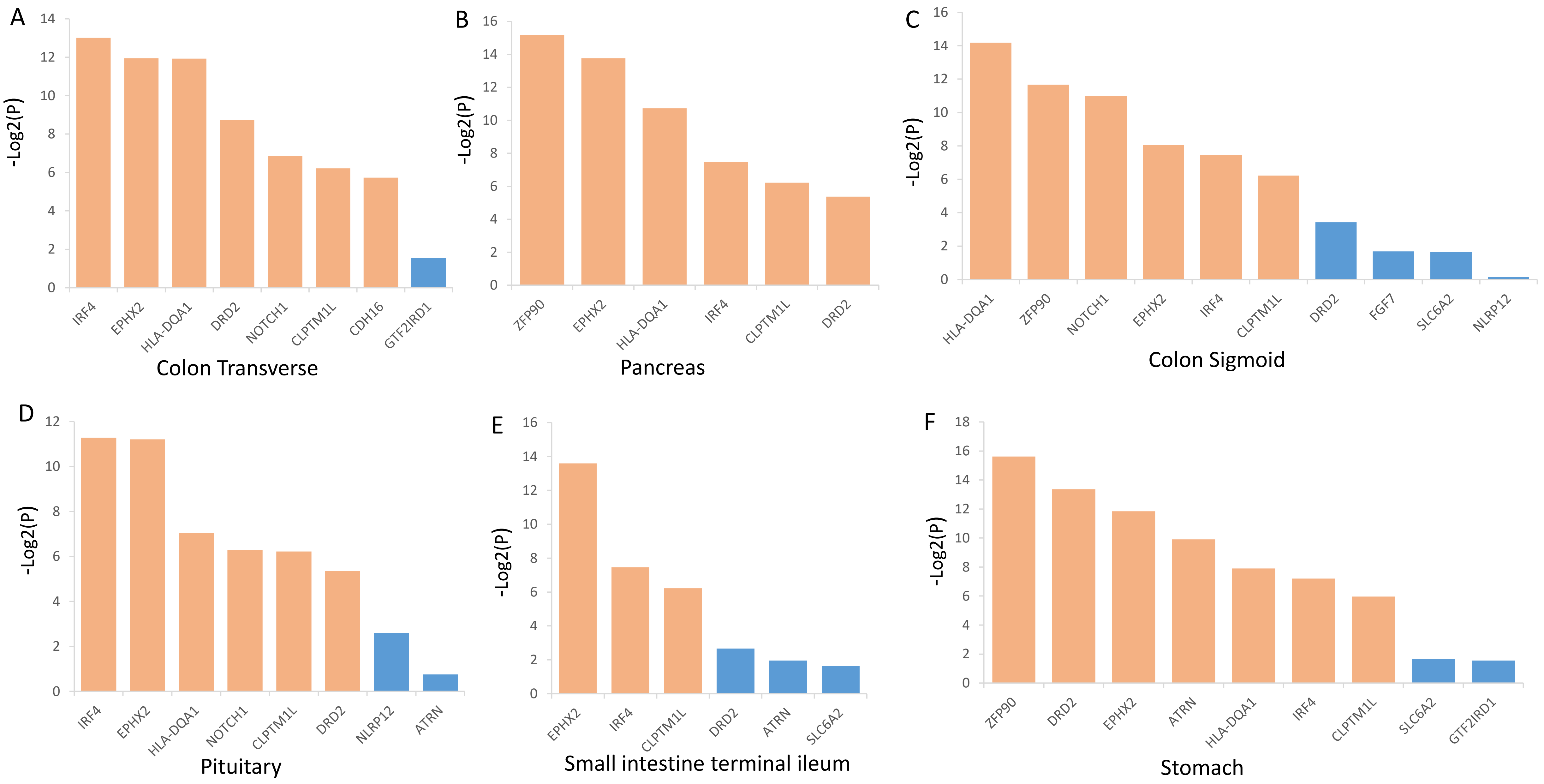


Supplemental Figure S26

**Supplemental Figure S27.** Bar plot shows the enrichment results of these 14 common risk genes identified from MAGMA and S-MultiXcan by using the DisGeNET algorithm in the Metascape.

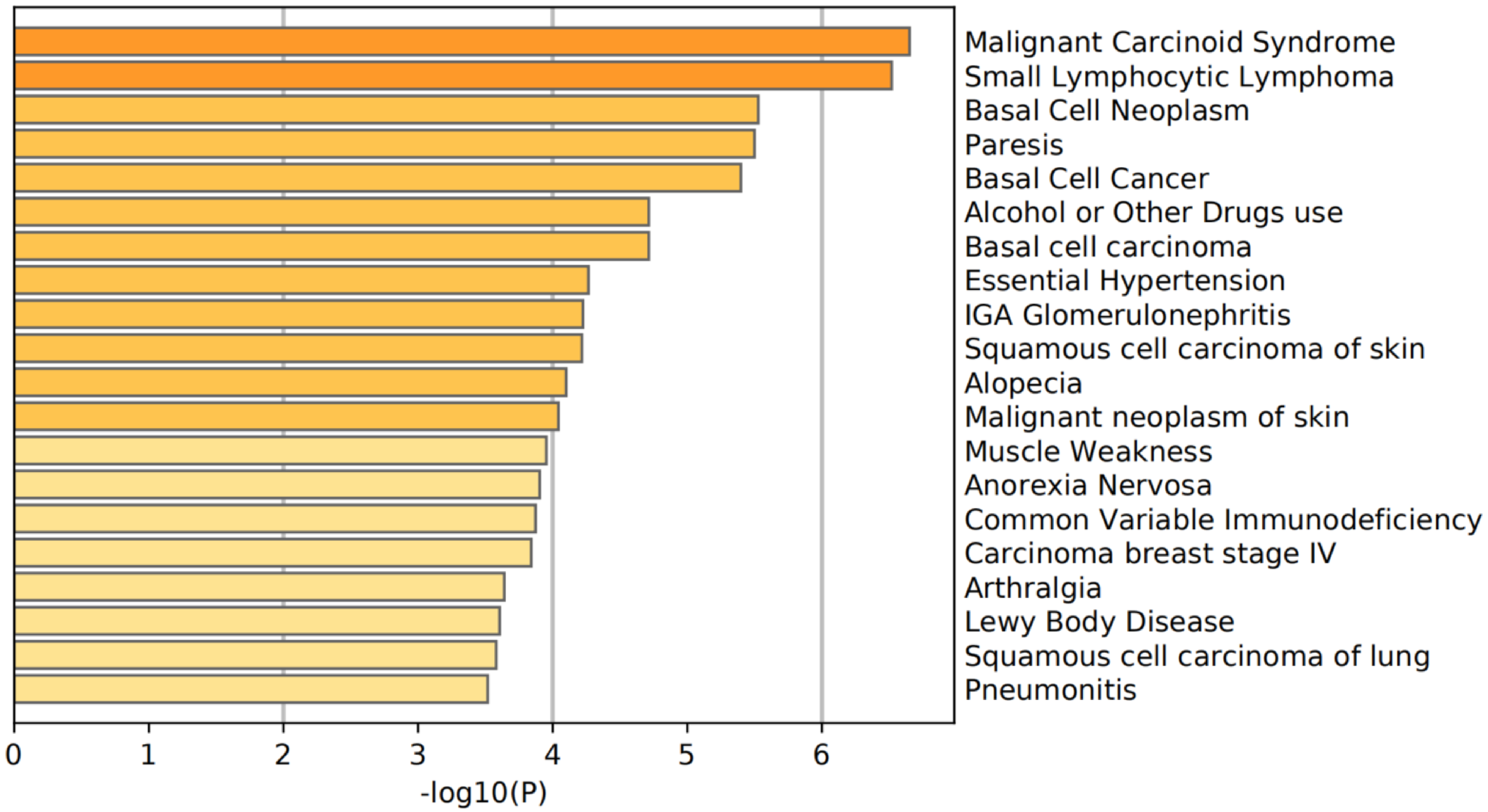


Supplemental Figure S27

**Supplemental Figure S28.** Drug-gene enrichment analysis shows 10 of 14 risk genes enriched in 10 potential druggable gene categories based on the DGIdb database.

| Druggable Gene Category | Matching Gene Count | Matching Gene(s) | Non-Matching Gene(s) |
|---|---|---|---|
| DRUGGABLE GENOME | 6 | NOTCH1, ATRN, DRD2, EPHX2, SLC6A2, FGF7 | IRF4, TNS3, HLA-DQA1, GTF2IRD1 |
| CLINICALLY ACTIONABLE | 4 | IRF4, NOTCH1, HLA-DQA1, FGF7 | TNS3, ATRN, DRD2, EPHX2, SLC6A2, GTF2IRD1 |
| CELL SURFACE | 2 | NOTCH1, SLC6A2 | IRF4, TNS3, ATRN, HLA-DQA1, DRD2, EPHX2, FGF7, GTF2IRD1 |
| DRUG RESISTANCE | 2 | DRD2, SLC6A2 | IRF4, TNS3, NOTCH1, ATRN, HLA-DQA1, EPHX2, FGF7, GTF2IRD1 |
| TRANSCRIPTION FACTOR | 2 | IRF4, GTF2IRD1 | TNS3, NOTCH1, ATRN, HLA-DQA1, DRD2, EPHX2, SLC6A2, FGF7 |
| TRANSPORTER | 2 | NOTCH1, SLC6A2 | IRF4, TNS3, ATRN, HLA-DQA1, DRD2, EPHX2, FGF7, GTF2IRD1 |
| ENZYME | 1 | EPHX2 | IRF4, TNS3, NOTCH1, ATRN, HLA-DQA1, DRD2, SLC6A2, FGF7, GTF2IRD1 |
| G PROTEIN COUPLED RECEPTOR | 1 | DRD2 | IRF4, TNS3, NOTCH1, ATRN, HLA-DQA1, EPHX2, SLC6A2, FGF7, GTF2IRD1 |
| GROWTH FACTOR | 1 | FGF7 | IRF4, TNS3, NOTCH1, ATRN, HLA-DQA1, DRD2, EPHX2, SLC6A2, GTF2IRD1 |
| PTEN FAMILY | 1 | TNS3 | IRF4, NOTCH1, ATRN, HLA-DQA1, DRD2, EPHX2, SLC6A2, FGF7, GTF2IRD1 |

Supplemental Figure S28

**Supplemental Figure S29.** Co-expression patterns of 14 genes between OCC patients and paracancerous controls based on the GSE139869 dataset.

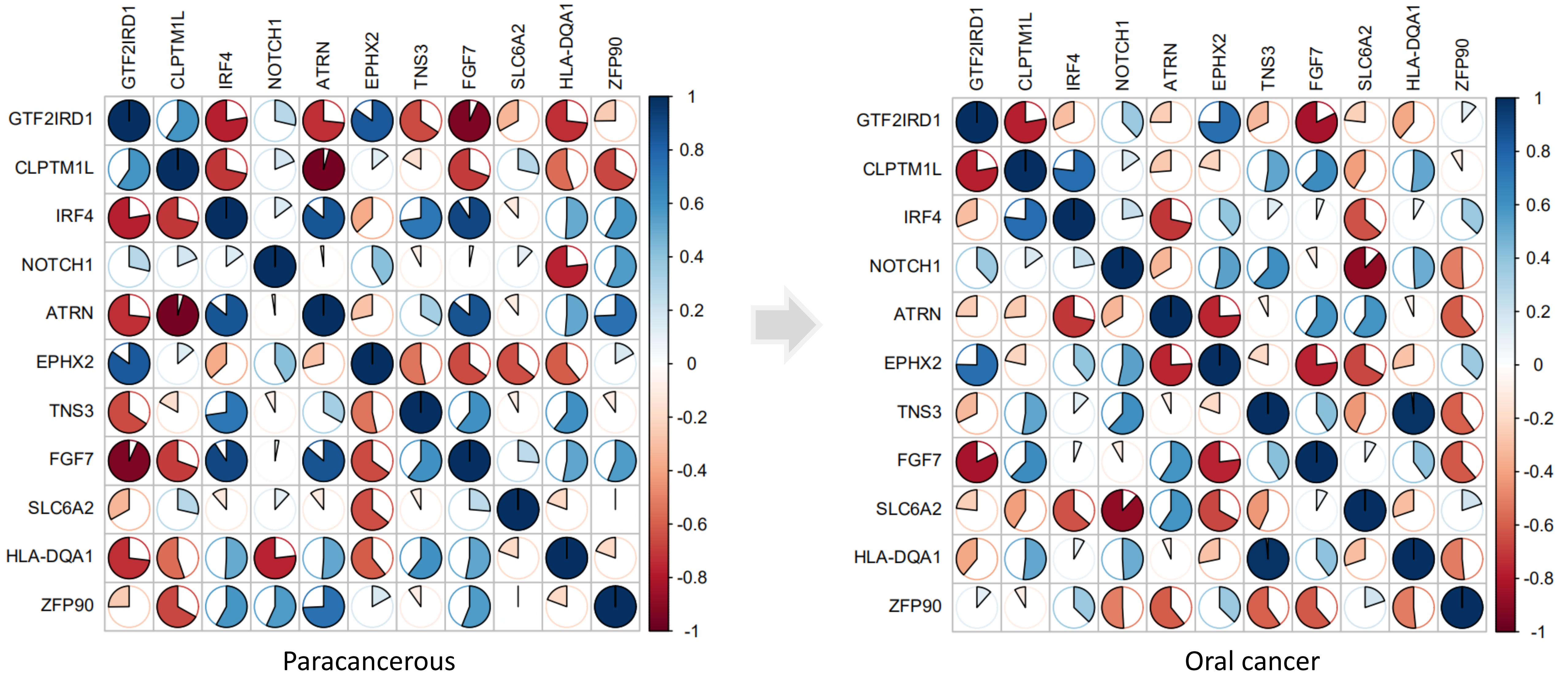


Supplemental Figure S29

**Supplemental Figure S30.** Co-expression patterns of 14 genes between OCC patients and paracancerous controls based on the GSE160042 dataset**.**

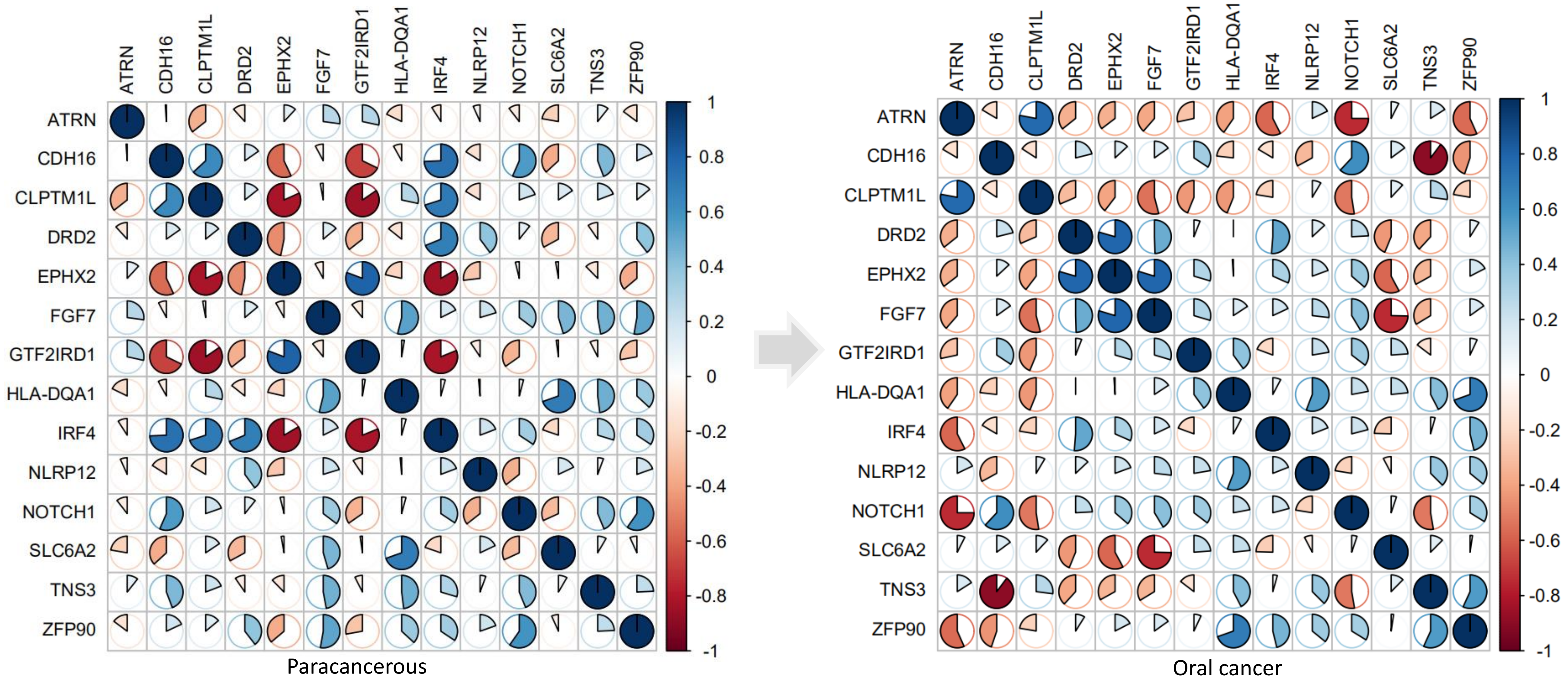


Supplemental Figure S30
