## Supplemental Tables for "Integrated multi-omics data analysis identifies a novel genetics-risk gene of *IRF4* associated with prognosis of oral cavity cancer"

**Supplemental Table S1. Significant genes identified by using MAGMA gene-based association analysis**

| **Gene** | **CHR** | **Z_score** | **P value** | **FDR** | **Reported studies on oral cancer** | **Cancer-related genes from the GWAS catalog** |
| --- | --- | --- | --- | --- | --- | --- |
| *IRF4* | 6 | 5.85 | 2.50E-09 | 4.59E-05 | Non-documented | Documented |
| *TNS3* | 7 | 4.68 | 1.44E-06 | 1.09E-02 | Non-documented | Documented |
| *NOTCH1* | 9 | 4.60 | 2.13E-06 | 1.09E-02 | Non-documented | Non-documented |
| *ZFP90* | 16 | 4.58 | 2.37E-06 | 1.09E-02 | Non-documented | Non-documented |
| *ATRN* | 20 | 4.51 | 3.28E-06 | 1.21E-02 | Non-documented | Non-documented |
| *HLA-DQA1* | 6 | 4.36 | 6.45E-06 | 1.98E-02 | Reported | Documented |
| *DRD2* | 11 | 4.11 | 2.00E-05 | 4.35E-02 | Non-documented | Non-documented |
| *EPHX2* | 8 | 4.10 | 2.03E-05 | 4.35E-02 | Non-documented | Documented |
| *CLPTM1L* | 5 | 4.09 | 2.13E-05 | 4.35E-02 | Reported | Documented |
| *SLC6A2* | 16 | 4.03 | 2.76E-05 | 4.54E-02 | Non-documented | Non-documented |
| *FGF7* | 15 | 4.03 | 2.78E-05 | 4.54E-02 | Non-documented | Documented |
| *GTF2IRD1* | 7 | 4.00 | 3.15E-05 | 4.54E-02 | Non-documented | Non-documented |
| *NLRP12* | 19 | 3.99 | 3.26E-05 | 4.54E-02 | Non-documented | Documented |
| *CDH16* | 16 | 3.98 | 3.46E-05 | 4.54E-02 | Non-documented | Non-documented |
| *SAPCD1* | 6 | 3.93 | 4.24E-05 | 5.19E-02 | Non-documented | Non-documented |
| *VWA7* | 6 | 3.90 | 4.72E-05 | 5.29E-02 | Non-documented | Non-documented |
| *GSTA5* | 6 | 3.88 | 5.18E-05 | 5.29E-02 | Non-documented | Non-documented |
| *FAM72B* | 1 | 3.88 | 5.18E-05 | 5.29E-02 | Non-documented | Non-documented |
| *FCGR1B* | 1 | 3.84 | 6.19E-05 | 5.72E-02 | Non-documented | Non-documented |
| *MSH5* | 6 | 3.84 | 6.23E-05 | 5.72E-02 | Non-documented | Documented |
| *ATP2A3* | 17 | 3.81 | 6.87E-05 | 6.01E-02 | Non-documented | Non-documented |
| *MROH9* | 1 | 3.79 | 7.59E-05 | 6.25E-02 | Non-documented | Non-documented |
| *PDP2* | 16 | 3.77 | 8.13E-05 | 6.25E-02 | Non-documented | Non-documented |
| *FAM227B* | 15 | 3.77 | 8.16E-05 | 6.25E-02 | Non-documented | Documented |
| *SIAH3* | 13 | 3.71 | 1.03E-04 | 7.11E-02 | Non-documented | Non-documented |
| *RRAD* | 16 | 3.71 | 1.04E-04 | 7.11E-02 | Non-documented | Non-documented |
| *SPSB4* | 3 | 3.71 | 1.05E-04 | 7.11E-02 | Non-documented | Non-documented |
| *LOC100996318* | 1 | 3.70 | 1.10E-04 | 7.21E-02 | Non-documented | Non-documented |
| *KLHL24* | 3 | 3.68 | 1.15E-04 | 7.30E-02 | Non-documented | Non-documented |
| *HLA-DQB1* | 6 | 3.67 | 1.19E-04 | 7.32E-02 | Reported | Documented |
| *VARS* | 6 | 3.66 | 1.25E-04 | 7.41E-02 | Non-documented | Non-documented |
| *NPY* | 7 | 3.64 | 1.34E-04 | 7.48E-02 | Non-documented | Non-documented |
| *42798* | 2 | 3.64 | 1.34E-04 | 7.48E-02 | Non-documented | Non-documented |
| *TNXB* | 6 | 3.62 | 1.45E-04 | 7.85E-02 | Non-documented | Documented |
| *XKR5* | 8 | 3.60 | 1.59E-04 | 8.33E-02 | Non-documented | Non-documented |
| *ADCY1* | 7 | 3.59 | 1.64E-04 | 8.38E-02 | Non-documented | Non-documented |
| *BCAT2* | 19 | 3.56 | 1.86E-04 | 9.04E-02 | Non-documented | Non-documented |
| *CDH3* | 16 | 3.56 | 1.87E-04 | 9.04E-02 | Non-documented | Documented |
| *NFE2L1* | 17 | 3.54 | 1.97E-04 | 9.17E-02 | Non-documented | Non-documented |
| *P2RX1* | 17 | 3.53 | 2.04E-04 | 9.17E-02 | Non-documented | Non-documented |
| *COPZ2* | 17 | 3.53 | 2.05E-04 | 9.17E-02 | Non-documented | Non-documented |
| *LOC101929008* | 7 | 3.51 | 2.26E-04 | 9.84E-02 | Non-documented | Non-documented |
| *FGF17* | 8 | 3.50 | 2.30E-04 | 9.84E-02 | Non-documented | Non-documented |
| *EHD3* | 2 | 3.45 | 2.86E-04 | 1.19E-01 | Non-documented | Non-documented |
| *SRCAP* | 16 | 3.43 | 2.98E-04 | 1.22E-01 | Non-documented | Non-documented |
| *CYP21A2* | 6 | 3.40 | 3.33E-04 | 1.33E-01 | Non-documented | Documented |
| *ACKR3* | 2 | 3.38 | 3.59E-04 | 1.40E-01 | Non-documented | Non-documented |
| *FAM189A1* | 15 | 3.37 | 3.83E-04 | 1.47E-01 | Non-documented | Non-documented |
| *TMEM265* | 16 | 3.35 | 4.00E-04 | 1.50E-01 | Non-documented | Non-documented |
| *CACHD1* | 1 | 3.34 | 4.22E-04 | 1.50E-01 | Non-documented | Non-documented |
| *PSG8* | 19 | 3.34 | 4.26E-04 | 1.50E-01 | Non-documented | Non-documented |
| *FAM96B* | 16 | 3.33 | 4.30E-04 | 1.50E-01 | Non-documented | Non-documented |
| *RNF150* | 4 | 3.33 | 4.37E-04 | 1.50E-01 | Non-documented | Non-documented |
| *CES2* | 16 | 3.32 | 4.45E-04 | 1.50E-01 | Non-documented | Non-documented |
| *PSG3* | 19 | 3.32 | 4.50E-04 | 1.50E-01 | Non-documented | Non-documented |
| *C4B* | 6 | 3.31 | 4.65E-04 | 1.52E-01 | Non-documented | Documented |
| *TMEM144* | 4 | 3.30 | 4.77E-04 | 1.54E-01 | Non-documented | Non-documented |
| *COL15A1* | 9 | 3.30 | 4.92E-04 | 1.54E-01 | Non-documented | Documented |
| *ZCCHC24* | 10 | 3.29 | 4.94E-04 | 1.54E-01 | Non-documented | Non-documented |
| *VTI1A* | 10 | 3.28 | 5.16E-04 | 1.58E-01 | Non-documented | Documented |
| *SLC5A10* | 17 | 3.26 | 5.57E-04 | 1.65E-01 | Non-documented | Non-documented |
| *RPP40* | 6 | 3.26 | 5.62E-04 | 1.65E-01 | Non-documented | Non-documented |
| *S100A2* | 1 | 3.26 | 5.65E-04 | 1.65E-01 | Non-documented | Non-documented |
| *ZNF512* | 2 | 3.25 | 5.86E-04 | 1.67E-01 | Reported | Non-documented |
| *GFRA4* | 20 | 3.24 | 5.92E-04 | 1.67E-01 | Non-documented | Non-documented |
| *HSPA1L* | 6 | 3.23 | 6.14E-04 | 1.68E-01 | Non-documented | Non-documented |
| *LIPC* | 15 | 3.23 | 6.14E-04 | 1.68E-01 | Non-documented | Documented |
| *FAM20C* | 7 | 3.21 | 6.57E-04 | 1.77E-01 | Non-documented | Non-documented |
| *KRT3* | 12 | 3.21 | 6.64E-04 | 1.77E-01 | Non-documented | Non-documented |
| *LDHAL6A* | 11 | 3.19 | 7.00E-04 | 1.80E-01 | Non-documented | Non-documented |
| *PPHLN1* | 12 | 3.18 | 7.43E-04 | 1.80E-01 | Non-documented | Non-documented |
| *CES3* | 16 | 3.17 | 7.53E-04 | 1.80E-01 | Non-documented | Non-documented |
| *CLCA4* | 1 | 3.17 | 7.58E-04 | 1.80E-01 | Non-documented | Non-documented |
| *CBX1* | 17 | 3.17 | 7.75E-04 | 1.80E-01 | Non-documented | Non-documented |
| *LCORL* | 4 | 3.16 | 7.85E-04 | 1.80E-01 | Non-documented | Documented |
| *JAK1* | 1 | 3.16 | 7.86E-04 | 1.80E-01 | Non-documented | Documented |
| *ZNF479* | 7 | 3.16 | 7.93E-04 | 1.80E-01 | Non-documented | Non-documented |
| *ZNF391* | 6 | 3.16 | 7.95E-04 | 1.80E-01 | Non-documented | Non-documented |
| *NCAPG* | 4 | 3.16 | 7.96E-04 | 1.80E-01 | Non-documented | Non-documented |
| *CCDC121* | 2 | 3.16 | 7.96E-04 | 1.80E-01 | Reported | Non-documented |
| *KCNJ12* | 17 | 3.16 | 8.02E-04 | 1.80E-01 | Non-documented | Non-documented |
| *LSM2* | 6 | 3.15 | 8.05E-04 | 1.80E-01 | Non-documented | Non-documented |
| *FABP7* | 6 | 3.14 | 8.53E-04 | 1.87E-01 | Non-documented | Non-documented |
| *C4A* | 6 | 3.14 | 8.55E-04 | 1.87E-01 | Non-documented | Documented |
| *HLA-DMA* | 6 | 3.13 | 8.68E-04 | 1.87E-01 | Non-documented | Non-documented |
| *LAMA1* | 18 | 3.13 | 8.75E-04 | 1.87E-01 | Non-documented | Non-documented |
| *TMCC1* | 3 | 3.12 | 8.91E-04 | 1.88E-01 | Non-documented | Non-documented |
| *LOC730183* | 16 | 3.12 | 9.02E-04 | 1.88E-01 | Non-documented | Non-documented |
| *NDUFAF7* | 2 | 3.12 | 9.18E-04 | 1.90E-01 | Non-documented | Non-documented |
| *NPM2* | 8 | 3.10 | 9.59E-04 | 1.94E-01 | Non-documented | Non-documented |
| *KDM2B* | 12 | 3.10 | 9.65E-04 | 1.94E-01 | Non-documented | Non-documented |
| *WBSCR28* | 7 | 3.10 | 9.70E-04 | 1.94E-01 | Non-documented | Non-documented |
| *C10orf62* | 10 | 3.10 | 9.83E-04 | 1.94E-01 | Non-documented | Non-documented |
| *S1PR1* | 1 | 3.08 | 1.03E-03 | 1.99E-01 | Non-documented | Documented |
| *MAP3K4* | 6 | 3.08 | 1.04E-03 | 1.99E-01 | Non-documented | Non-documented |
| *SMPDL3A* | 6 | 3.08 | 1.04E-03 | 1.99E-01 | Non-documented | Non-documented |
| *SECISBP2L* | 15 | 3.07 | 1.07E-03 | 1.99E-01 | Non-documented | Documented |
| *HCRT* | 17 | 3.07 | 1.08E-03 | 1.99E-01 | Non-documented | Non-documented |
| *RHPN1* | 8 | 3.07 | 1.08E-03 | 1.99E-01 | Non-documented | Non-documented |
| *C2orf16* | 2 | 3.07 | 1.08E-03 | 1.99E-01 | Reported | Non-documented |
| *DCAF16* | 4 | 3.06 | 1.11E-03 | 2.00E-01 | Non-documented | Non-documented |
| *ZCRB1* | 12 | 3.05 | 1.13E-03 | 2.00E-01 | Non-documented | Non-documented |
| *ASCL5* | 1 | 3.05 | 1.13E-03 | 2.00E-01 | Non-documented | Non-documented |
| *FBRS* | 16 | 3.05 | 1.13E-03 | 2.00E-01 | Non-documented | Non-documented |
| *ETNPPL* | 4 | 3.04 | 1.17E-03 | 2.04E-01 | Non-documented | Non-documented |
| *EPM2A* | 6 | 3.03 | 1.22E-03 | 2.12E-01 | Non-documented | Non-documented |
| *HLA-DMB* | 6 | 3.02 | 1.25E-03 | 2.14E-01 | Non-documented | Documented |
| *RNF4* | 4 | 3.02 | 1.26E-03 | 2.14E-01 | Non-documented | Documented |
| *HLA-B* | 6 | 3.02 | 1.27E-03 | 2.14E-01 | Non-documented | Documented |
| *S100A3* | 1 | 3.01 | 1.29E-03 | 2.14E-01 | Non-documented | Non-documented |
| *LDHC* | 11 | 3.01 | 1.30E-03 | 2.14E-01 | Non-documented | Non-documented |
| *BAG6* | 6 | 3.01 | 1.30E-03 | 2.14E-01 | Non-documented | Documented |
| *MPP5* | 14 | 2.99 | 1.38E-03 | 2.25E-01 | Non-documented | Non-documented |
| *RNF34* | 12 | 2.99 | 1.40E-03 | 2.25E-01 | Non-documented | Non-documented |
| *PFN2* | 3 | 2.98 | 1.43E-03 | 2.27E-01 | Non-documented | Non-documented |
| *MPHOSPH8* | 13 | 2.98 | 1.43E-03 | 2.27E-01 | Non-documented | Non-documented |
| *KCMF1* | 2 | 2.98 | 1.46E-03 | 2.27E-01 | Non-documented | Non-documented |
| *FOXE3* | 1 | 2.98 | 1.46E-03 | 2.27E-01 | Non-documented | Non-documented |
| *C6orf15* | 6 | 2.97 | 1.47E-03 | 2.27E-01 | Non-documented | Documented |
| *S100A4* | 1 | 2.97 | 1.49E-03 | 2.28E-01 | Non-documented | Non-documented |
| *SNX11* | 17 | 2.96 | 1.54E-03 | 2.34E-01 | Non-documented | Non-documented |
| *PSPC1* | 13 | 2.96 | 1.56E-03 | 2.34E-01 | Non-documented | Non-documented |
| *GHDC* | 17 | 2.95 | 1.57E-03 | 2.35E-01 | Non-documented | Non-documented |
| *HAND1* | 5 | 2.94 | 1.62E-03 | 2.38E-01 | Non-documented | Non-documented |
| *APOM* | 6 | 2.94 | 1.62E-03 | 2.38E-01 | Non-documented | Documented |
| *GPN1* | 2 | 2.94 | 1.64E-03 | 2.40E-01 | Reported | Non-documented |
| *CD320* | 19 | 2.93 | 1.70E-03 | 2.42E-01 | Non-documented | Non-documented |
| *NGEF* | 2 | 2.93 | 1.72E-03 | 2.42E-01 | Non-documented | Non-documented |
| *KIF25* | 6 | 2.93 | 1.72E-03 | 2.42E-01 | Non-documented | Non-documented |
| *S100A5* | 1 | 2.92 | 1.73E-03 | 2.42E-01 | Non-documented | Non-documented |
| *NEFL* | 8 | 2.92 | 1.74E-03 | 2.42E-01 | Non-documented | Non-documented |
| *IGFBP3* | 7 | 2.92 | 1.77E-03 | 2.42E-01 | Non-documented | Non-documented |
| *BCL2L15* | 1 | 2.92 | 1.77E-03 | 2.42E-01 | Non-documented | Documented |
| *CDYL2* | 16 | 2.92 | 1.77E-03 | 2.42E-01 | Non-documented | Documented |
| *SYT1* | 12 | 2.92 | 1.78E-03 | 2.42E-01 | Non-documented | Non-documented |
| *LOC731631* | 7 | 2.90 | 1.86E-03 | 2.50E-01 | Non-documented | Non-documented |
| *COX6A2* | 16 | 2.90 | 1.86E-03 | 2.50E-01 | Non-documented | Non-documented |
| *PHKG2* | 16 | 2.89 | 1.91E-03 | 2.51E-01 | Non-documented | Non-documented |
| *RNF13* | 3 | 2.89 | 1.91E-03 | 2.51E-01 | Non-documented | Non-documented |
| *TAMM41* | 3 | 2.89 | 1.91E-03 | 2.51E-01 | Non-documented | Non-documented |
| *RNASE8* | 14 | 2.89 | 1.94E-03 | 2.52E-01 | Non-documented | Non-documented |
| *HSPA1A* | 6 | 2.89 | 1.95E-03 | 2.52E-01 | Non-documented | Non-documented |
| *FKBPL* | 6 | 2.88 | 1.97E-03 | 2.52E-01 | Non-documented | Documented |
| *RPTOR* | 17 | 2.88 | 1.98E-03 | 2.52E-01 | Non-documented | Non-documented |
| *SELPLG* | 12 | 2.87 | 2.03E-03 | 2.56E-01 | Non-documented | Non-documented |
| *CCDC33* | 15 | 2.87 | 2.04E-03 | 2.56E-01 | Non-documented | Non-documented |
| *HOGA1* | 10 | 2.87 | 2.06E-03 | 2.56E-01 | Non-documented | Non-documented |
| *ABO* | 9 | 2.87 | 2.07E-03 | 2.56E-01 | Non-documented | Documented |
| *PTGFRN* | 1 | 2.86 | 2.09E-03 | 2.57E-01 | Non-documented | Documented |
| *ADAM33* | 20 | 2.86 | 2.13E-03 | 2.61E-01 | Non-documented | Non-documented |
| *PYDC2* | 3 | 2.85 | 2.17E-03 | 2.65E-01 | Non-documented | Non-documented |
| *OR5V1* | 6 | 2.85 | 2.21E-03 | 2.67E-01 | Non-documented | Non-documented |
| *SMC6* | 2 | 2.84 | 2.23E-03 | 2.67E-01 | Non-documented | Non-documented |
| *CDSN* | 6 | 2.84 | 2.23E-03 | 2.67E-01 | Non-documented | Documented |
| *WRAP73* | 1 | 2.84 | 2.25E-03 | 2.67E-01 | Non-documented | Non-documented |
| *KRT4* | 12 | 2.84 | 2.27E-03 | 2.67E-01 | Non-documented | Non-documented |
| *ZMYM5* | 13 | 2.84 | 2.29E-03 | 2.67E-01 | Non-documented | Non-documented |
| *C6orf47* | 6 | 2.83 | 2.31E-03 | 2.67E-01 | Non-documented | Non-documented |
| *FAM83G* | 17 | 2.83 | 2.31E-03 | 2.67E-01 | Non-documented | Non-documented |
| *FGD4* | 12 | 2.83 | 2.34E-03 | 2.69E-01 | Non-documented | Documented |
| *PRDX3* | 10 | 2.83 | 2.36E-03 | 2.70E-01 | Non-documented | Non-documented |
| *PPP2R2B* | 5 | 2.82 | 2.40E-03 | 2.70E-01 | Non-documented | Non-documented |
| *ANKRD2* | 10 | 2.82 | 2.41E-03 | 2.70E-01 | Non-documented | Non-documented |
| *ZNF184* | 6 | 2.82 | 2.42E-03 | 2.70E-01 | Non-documented | Documented |
| *SFXN4* | 10 | 2.81 | 2.46E-03 | 2.70E-01 | Non-documented | Non-documented |
| *DMTN* | 8 | 2.81 | 2.47E-03 | 2.70E-01 | Non-documented | Non-documented |
| *BTBD2* | 19 | 2.81 | 2.50E-03 | 2.70E-01 | Non-documented | Non-documented |
| *SHPRH* | 6 | 2.81 | 2.51E-03 | 2.70E-01 | Non-documented | Non-documented |
| *HSPB3* | 5 | 2.81 | 2.51E-03 | 2.70E-01 | Non-documented | Non-documented |
| *AKR1B1* | 7 | 2.80 | 2.52E-03 | 2.70E-01 | Non-documented | Non-documented |
| *KLHL30* | 2 | 2.80 | 2.54E-03 | 2.70E-01 | Non-documented | Non-documented |
| *FAM160B2* | 8 | 2.80 | 2.54E-03 | 2.70E-01 | Non-documented | Documented |
| *S100A6* | 1 | 2.80 | 2.54E-03 | 2.70E-01 | Non-documented | Non-documented |
| *ATF6B* | 6 | 2.80 | 2.56E-03 | 2.70E-01 | Non-documented | Documented |
| *FMOD* | 1 | 2.80 | 2.58E-03 | 2.70E-01 | Non-documented | Non-documented |
| *ATP6V1D* | 14 | 2.80 | 2.59E-03 | 2.70E-01 | Non-documented | Non-documented |
| *DEFA1* | 8 | 2.79 | 2.62E-03 | 2.70E-01 | Non-documented | Non-documented |
| *DNAL4* | 22 | 2.79 | 2.63E-03 | 2.70E-01 | Non-documented | Non-documented |
| *CCR8* | 3 | 2.79 | 2.63E-03 | 2.70E-01 | Non-documented | Non-documented |
| *TARSL2* | 15 | 2.79 | 2.67E-03 | 2.72E-01 | Non-documented | Non-documented |
| *SLC25A38* | 3 | 2.77 | 2.80E-03 | 2.81E-01 | Non-documented | Non-documented |
| *ZNF14* | 19 | 2.77 | 2.82E-03 | 2.81E-01 | Non-documented | Non-documented |
| *HSPA1B* | 6 | 2.77 | 2.83E-03 | 2.81E-01 | Non-documented | Non-documented |
| *ZNF345* | 19 | 2.77 | 2.85E-03 | 2.81E-01 | Non-documented | Non-documented |
| *HMX1* | 4 | 2.76 | 2.85E-03 | 2.81E-01 | Non-documented | Non-documented |
| *EFCAB8* | 20 | 2.76 | 2.85E-03 | 2.81E-01 | Non-documented | Non-documented |
| *SNX8* | 7 | 2.76 | 2.86E-03 | 2.81E-01 | Non-documented | Documented |
| *R3HDML* | 20 | 2.76 | 2.89E-03 | 2.81E-01 | Non-documented | Non-documented |
| *TERT* | 5 | 2.76 | 2.89E-03 | 2.81E-01 | Non-documented | Documented |
| *KLF7* | 2 | 2.76 | 2.92E-03 | 2.81E-01 | Non-documented | Non-documented |
| *NEU4* | 2 | 2.76 | 2.93E-03 | 2.81E-01 | Non-documented | Non-documented |
| *MAPRE1* | 20 | 2.76 | 2.93E-03 | 2.81E-01 | Non-documented | Non-documented |
| *BRD2* | 6 | 2.75 | 2.95E-03 | 2.81E-01 | Non-documented | Non-documented |
| *ZNF843* | 16 | 2.75 | 2.97E-03 | 2.81E-01 | Non-documented | Non-documented |
| *C6orf48* | 6 | 2.74 | 3.11E-03 | 2.93E-01 | Non-documented | Documented |
| *MBD3L1* | 19 | 2.73 | 3.14E-03 | 2.94E-01 | Non-documented | Non-documented |
| *GCSAML* | 1 | 2.73 | 3.17E-03 | 2.94E-01 | Non-documented | Non-documented |
| *HMX3* | 10 | 2.73 | 3.18E-03 | 2.94E-01 | Non-documented | Non-documented |
| *CADM2* | 3 | 2.73 | 3.19E-03 | 2.94E-01 | Non-documented | Non-documented |
| *SLC7A1* | 13 | 2.72 | 3.25E-03 | 2.99E-01 | Non-documented | Non-documented |
| *PRRC2A* | 6 | 2.71 | 3.32E-03 | 3.01E-01 | Non-documented | Non-documented |
| *CD3E* | 11 | 2.71 | 3.32E-03 | 3.01E-01 | Non-documented | Documented |
| *TGFBR2* | 3 | 2.71 | 3.33E-03 | 3.01E-01 | Non-documented | Documented |
| *MYT1* | 20 | 2.71 | 3.36E-03 | 3.01E-01 | Non-documented | Non-documented |
| *CYR61* | 1 | 2.71 | 3.37E-03 | 3.01E-01 | Non-documented | Non-documented |
| *CXCL12* | 10 | 2.71 | 3.37E-03 | 3.01E-01 | Non-documented | Non-documented |
| *PCDH7* | 4 | 2.71 | 3.39E-03 | 3.01E-01 | Non-documented | Non-documented |
| *GALNT14* | 2 | 2.70 | 3.42E-03 | 3.02E-01 | Reported | Non-documented |
| *BRE* | 2 | 2.70 | 3.46E-03 | 3.03E-01 | Non-documented | Documented |
| *FAM162A* | 3 | 2.70 | 3.46E-03 | 3.03E-01 | Non-documented | Non-documented |
| *SNRPA1* | 15 | 2.69 | 3.55E-03 | 3.09E-01 | Non-documented | Documented |
| *CDH1* | 16 | 2.69 | 3.60E-03 | 3.11E-01 | Non-documented | Documented |
| *DENND2A* | 7 | 2.69 | 3.61E-03 | 3.11E-01 | Non-documented | Non-documented |
| *TEKT4* | 2 | 2.69 | 3.62E-03 | 3.11E-01 | Non-documented | Non-documented |
| *FEZ2* | 2 | 2.68 | 3.70E-03 | 3.16E-01 | Non-documented | Non-documented |
| *FAM163B* | 9 | 2.67 | 3.74E-03 | 3.18E-01 | Non-documented | Non-documented |
| *EIF2S1* | 14 | 2.67 | 3.76E-03 | 3.19E-01 | Non-documented | Non-documented |
| *RSBN1* | 1 | 2.67 | 3.83E-03 | 3.23E-01 | Non-documented | Non-documented |
| *LDHB* | 12 | 2.66 | 3.86E-03 | 3.24E-01 | Non-documented | Non-documented |
| *TMEM183B* | 3 | 2.66 | 3.89E-03 | 3.24E-01 | Non-documented | Non-documented |
| *BIRC6* | 2 | 2.66 | 3.90E-03 | 3.24E-01 | Non-documented | Non-documented |
| *LTBP1* | 2 | 2.66 | 3.93E-03 | 3.25E-01 | Non-documented | Non-documented |
| *RPSA* | 3 | 2.66 | 3.96E-03 | 3.25E-01 | Non-documented | Non-documented |
| *CGRRF1* | 14 | 2.65 | 3.97E-03 | 3.25E-01 | Non-documented | Non-documented |
| *GEN1* | 2 | 2.65 | 4.02E-03 | 3.25E-01 | Non-documented | Non-documented |
| *KIAA1614* | 1 | 2.65 | 4.02E-03 | 3.25E-01 | Non-documented | Non-documented |
| *GAN* | 16 | 2.65 | 4.04E-03 | 3.25E-01 | Non-documented | Non-documented |
| *PTPN18* | 2 | 2.65 | 4.05E-03 | 3.25E-01 | Non-documented | Non-documented |
| *GIGYF2* | 2 | 2.65 | 4.05E-03 | 3.25E-01 | Non-documented | Non-documented |
| *ALDH9A1* | 1 | 2.64 | 4.10E-03 | 3.28E-01 | Non-documented | Non-documented |
| *WDR5B* | 3 | 2.64 | 4.12E-03 | 3.28E-01 | Non-documented | Non-documented |
| *ABCA4* | 1 | 2.64 | 4.16E-03 | 3.29E-01 | Non-documented | Non-documented |
| *VSNL1* | 2 | 2.64 | 4.20E-03 | 3.31E-01 | Non-documented | Documented |
| *FAM69B* | 9 | 2.63 | 4.27E-03 | 3.35E-01 | Non-documented | Non-documented |
| *ARHGEF10L* | 1 | 2.63 | 4.30E-03 | 3.35E-01 | Non-documented | Documented |
| *VIMP* | 15 | 2.63 | 4.30E-03 | 3.35E-01 | Non-documented | Non-documented |
| *GPANK1* | 6 | 2.62 | 4.34E-03 | 3.36E-01 | Non-documented | Non-documented |
| *PLA2G4A* | 1 | 2.61 | 4.47E-03 | 3.44E-01 | Non-documented | Non-documented |
| *GCKR* | 2 | 2.61 | 4.49E-03 | 3.44E-01 | Non-documented | Non-documented |
| *GAB4* | 22 | 2.61 | 4.50E-03 | 3.44E-01 | Non-documented | Non-documented |
| *MPP7* | 10 | 2.61 | 4.51E-03 | 3.44E-01 | Non-documented | Non-documented |
| *FBXO30* | 6 | 2.61 | 4.55E-03 | 3.44E-01 | Non-documented | Non-documented |
| *CFAP99* | 4 | 2.61 | 4.56E-03 | 3.44E-01 | Non-documented | Non-documented |
| *TSPAN17* | 5 | 2.61 | 4.56E-03 | 3.44E-01 | Non-documented | Non-documented |
| *CXCL14* | 5 | 2.60 | 4.61E-03 | 3.44E-01 | Non-documented | Non-documented |
| *IMP4* | 2 | 2.60 | 4.63E-03 | 3.44E-01 | Non-documented | Non-documented |
| *SH3PXD2B* | 5 | 2.60 | 4.64E-03 | 3.44E-01 | Non-documented | Non-documented |
| *ACP6* | 1 | 2.60 | 4.67E-03 | 3.44E-01 | Non-documented | Non-documented |
| *CA7* | 16 | 2.60 | 4.67E-03 | 3.44E-01 | Non-documented | Non-documented |
| *PRKD3* | 2 | 2.60 | 4.68E-03 | 3.44E-01 | Non-documented | Non-documented |
| *C10orf142* | 10 | 2.60 | 4.69E-03 | 3.44E-01 | Non-documented | Non-documented |
| *AIF1* | 6 | 2.60 | 4.72E-03 | 3.44E-01 | Reported | Documented |
| *AGPAT4* | 6 | 2.59 | 4.78E-03 | 3.45E-01 | Non-documented | Non-documented |
| *GABRB2* | 5 | 2.59 | 4.80E-03 | 3.45E-01 | Non-documented | Non-documented |
| *SYT7* | 11 | 2.59 | 4.80E-03 | 3.45E-01 | Non-documented | Non-documented |
| *RBKS* | 2 | 2.59 | 4.82E-03 | 3.45E-01 | Non-documented | Non-documented |
| *CECR1* | 22 | 2.59 | 4.83E-03 | 3.45E-01 | Non-documented | Non-documented |
| *CCDC12* | 3 | 2.59 | 4.86E-03 | 3.46E-01 | Non-documented | Non-documented |
| *ANO5* | 11 | 2.58 | 4.88E-03 | 3.46E-01 | Non-documented | Non-documented |
| *NUDT18* | 8 | 2.58 | 4.89E-03 | 3.46E-01 | Non-documented | Non-documented |
| *HMGB2* | 4 | 2.58 | 4.95E-03 | 3.48E-01 | Non-documented | Non-documented |
| *ECH1* | 19 | 2.57 | 5.02E-03 | 3.52E-01 | Non-documented | Non-documented |
| *CMKLR1* | 12 | 2.57 | 5.03E-03 | 3.52E-01 | Non-documented | Documented |
| *HNRNPL* | 19 | 2.57 | 5.14E-03 | 3.54E-01 | Non-documented | Non-documented |
| *SAAL1* | 11 | 2.57 | 5.15E-03 | 3.54E-01 | Non-documented | Non-documented |
| *EPB41L3* | 18 | 2.57 | 5.16E-03 | 3.54E-01 | Non-documented | Non-documented |
| *DXO* | 6 | 2.56 | 5.17E-03 | 3.54E-01 | Non-documented | Non-documented |
| *TPH1* | 11 | 2.56 | 5.20E-03 | 3.54E-01 | Non-documented | Non-documented |
| *CNOT4* | 7 | 2.56 | 5.21E-03 | 3.54E-01 | Non-documented | Non-documented |
| *PDP1* | 8 | 2.56 | 5.22E-03 | 3.54E-01 | Non-documented | Non-documented |
| *CHST12* | 7 | 2.56 | 5.24E-03 | 3.54E-01 | Non-documented | Non-documented |
| *RNF144B* | 6 | 2.56 | 5.26E-03 | 3.54E-01 | Non-documented | Non-documented |
| *GSTA1* | 6 | 2.56 | 5.29E-03 | 3.54E-01 | Non-documented | Non-documented |
| *CAPN12* | 19 | 2.56 | 5.30E-03 | 3.54E-01 | Non-documented | Non-documented |
| *INO80C* | 18 | 2.56 | 5.31E-03 | 3.54E-01 | Non-documented | Non-documented |
| *KANK3* | 19 | 2.55 | 5.31E-03 | 3.54E-01 | Non-documented | Non-documented |
| *ZNF329* | 19 | 2.55 | 5.37E-03 | 3.55E-01 | Non-documented | Non-documented |
| *NEU1* | 6 | 2.55 | 5.38E-03 | 3.55E-01 | Non-documented | Documented |
| *NDUFA7* | 19 | 2.54 | 5.47E-03 | 3.55E-01 | Non-documented | Non-documented |
| *MB* | 22 | 2.54 | 5.48E-03 | 3.55E-01 | Non-documented | Non-documented |
| *HPCAL1* | 2 | 2.54 | 5.48E-03 | 3.55E-01 | Non-documented | Non-documented |
| *EXPH5* | 11 | 2.54 | 5.53E-03 | 3.55E-01 | Non-documented | Non-documented |
| *RPS28* | 19 | 2.54 | 5.54E-03 | 3.55E-01 | Non-documented | Non-documented |
| *MPP6* | 7 | 2.54 | 5.54E-03 | 3.55E-01 | Non-documented | Non-documented |
| *LOC101927245* | 8 | 2.54 | 5.55E-03 | 3.55E-01 | Non-documented | Non-documented |
| *TPRG1L* | 1 | 2.54 | 5.57E-03 | 3.55E-01 | Non-documented | Non-documented |
| *DEFB1* | 8 | 2.54 | 5.58E-03 | 3.55E-01 | Non-documented | Non-documented |
| *LOC100128108* | 15 | 2.54 | 5.59E-03 | 3.55E-01 | Non-documented | Non-documented |
| *FAM132B* | 2 | 2.54 | 5.62E-03 | 3.55E-01 | Non-documented | Non-documented |
| *42801* | 2 | 2.53 | 5.63E-03 | 3.55E-01 | Non-documented | Non-documented |
| *KCNH4* | 17 | 2.53 | 5.65E-03 | 3.55E-01 | Non-documented | Non-documented |
| *ZMYND19* | 9 | 2.53 | 5.66E-03 | 3.55E-01 | Non-documented | Non-documented |
| *MYADM* | 19 | 2.53 | 5.66E-03 | 3.55E-01 | Non-documented | Non-documented |
| *PHTF1* | 1 | 2.53 | 5.72E-03 | 3.56E-01 | Non-documented | Documented |
| *SKIV2L* | 6 | 2.53 | 5.72E-03 | 3.56E-01 | Non-documented | Documented |
| *GABRR3* | 3 | 2.53 | 5.76E-03 | 3.58E-01 | Non-documented | Non-documented |
| *FTO* | 16 | 2.52 | 5.81E-03 | 3.59E-01 | Non-documented | Documented |
| *FNDC4* | 2 | 2.52 | 5.84E-03 | 3.60E-01 | Non-documented | Non-documented |
| *AGPAT2* | 9 | 2.52 | 5.89E-03 | 3.60E-01 | Non-documented | Non-documented |
| *CCDC115* | 2 | 2.52 | 5.89E-03 | 3.60E-01 | Non-documented | Non-documented |
| *XPR1* | 1 | 2.52 | 5.92E-03 | 3.60E-01 | Non-documented | Non-documented |
| *PRR14* | 16 | 2.52 | 5.93E-03 | 3.60E-01 | Non-documented | Non-documented |
| *DPH7* | 9 | 2.51 | 5.99E-03 | 3.60E-01 | Non-documented | Non-documented |
| *DYNC1I1* | 7 | 2.51 | 6.00E-03 | 3.60E-01 | Non-documented | Non-documented |
| *PPIF* | 10 | 2.51 | 6.00E-03 | 3.60E-01 | Non-documented | Non-documented |
| *UROD* | 1 | 2.51 | 6.00E-03 | 3.60E-01 | Non-documented | Non-documented |
| *EIF4E* | 4 | 2.51 | 6.01E-03 | 3.60E-01 | Non-documented | Non-documented |
| *KCNJ13* | 2 | 2.51 | 6.03E-03 | 3.60E-01 | Non-documented | Non-documented |
| *ROPN1L* | 5 | 2.51 | 6.10E-03 | 3.63E-01 | Non-documented | Documented |
| *SERPINA3* | 14 | 2.50 | 6.24E-03 | 3.69E-01 | Non-documented | Non-documented |
| *KIAA1456* | 8 | 2.50 | 6.25E-03 | 3.69E-01 | Non-documented | Documented |
| *SNX24* | 5 | 2.49 | 6.33E-03 | 3.71E-01 | Non-documented | Non-documented |
| *GRIA2* | 4 | 2.49 | 6.33E-03 | 3.71E-01 | Non-documented | Non-documented |
| *NSMF* | 9 | 2.49 | 6.34E-03 | 3.71E-01 | Non-documented | Non-documented |
| *WNT11* | 11 | 2.49 | 6.37E-03 | 3.72E-01 | Non-documented | Non-documented |
| *C2orf82* | 2 | 2.48 | 6.50E-03 | 3.78E-01 | Non-documented | Non-documented |
| *IFT172* | 2 | 2.48 | 6.56E-03 | 3.80E-01 | Non-documented | Non-documented |
| *MUC17* | 7 | 2.48 | 6.59E-03 | 3.81E-01 | Non-documented | Non-documented |
| *GOLGA3* | 12 | 2.48 | 6.62E-03 | 3.82E-01 | Non-documented | Non-documented |
| *NUDT1* | 7 | 2.47 | 6.68E-03 | 3.84E-01 | Non-documented | Non-documented |
| *ZSWIM5* | 1 | 2.47 | 6.73E-03 | 3.84E-01 | Non-documented | Non-documented |
| *CALN1* | 7 | 2.47 | 6.75E-03 | 3.84E-01 | Non-documented | Non-documented |
| *MAS1L* | 6 | 2.47 | 6.75E-03 | 3.84E-01 | Non-documented | Non-documented |
| *LDHA* | 11 | 2.47 | 6.78E-03 | 3.84E-01 | Non-documented | Non-documented |
| *TRIM56* | 7 | 2.47 | 6.79E-03 | 3.84E-01 | Non-documented | Non-documented |
| *CAMTA1* | 1 | 2.47 | 6.84E-03 | 3.85E-01 | Non-documented | Non-documented |
| *TAS2R10* | 12 | 2.46 | 6.90E-03 | 3.87E-01 | Non-documented | Non-documented |
| *SLMO1* | 18 | 2.46 | 6.91E-03 | 3.87E-01 | Non-documented | Non-documented |
| *PEX11G* | 19 | 2.46 | 6.97E-03 | 3.89E-01 | Non-documented | Non-documented |
| *TBCB* | 19 | 2.46 | 7.00E-03 | 3.89E-01 | Non-documented | Non-documented |
| *HAUS8* | 19 | 2.46 | 7.01E-03 | 3.89E-01 | Non-documented | Non-documented |
| *RPP21* | 6 | 2.45 | 7.05E-03 | 3.90E-01 | Non-documented | Non-documented |
| *ABHD17C* | 15 | 2.45 | 7.08E-03 | 3.90E-01 | Non-documented | Non-documented |
| *DISC1* | 1 | 2.45 | 7.11E-03 | 3.90E-01 | Non-documented | Non-documented |
| *GPA33* | 1 | 2.45 | 7.14E-03 | 3.90E-01 | Non-documented | Non-documented |
| *ACTN4* | 19 | 2.45 | 7.14E-03 | 3.90E-01 | Non-documented | Non-documented |
| *CLIC1* | 6 | 2.45 | 7.20E-03 | 3.92E-01 | Non-documented | Non-documented |
| *QKI* | 6 | 2.45 | 7.22E-03 | 3.92E-01 | Non-documented | Non-documented |
| *ASB16* | 17 | 2.44 | 7.25E-03 | 3.92E-01 | Non-documented | Non-documented |
| *TEKT3* | 17 | 2.44 | 7.29E-03 | 3.92E-01 | Non-documented | Non-documented |
| *FGD5* | 3 | 2.44 | 7.30E-03 | 3.92E-01 | Non-documented | Non-documented |
| *SLC12A9* | 7 | 2.44 | 7.30E-03 | 3.92E-01 | Non-documented | Non-documented |
| *DDAH2* | 6 | 2.44 | 7.43E-03 | 3.97E-01 | Non-documented | Non-documented |
| *SLC6A4* | 17 | 2.44 | 7.43E-03 | 3.97E-01 | Non-documented | Non-documented |
| *RCBTB2* | 13 | 2.43 | 7.45E-03 | 3.97E-01 | Non-documented | Non-documented |
| *DUSP13* | 10 | 2.43 | 7.64E-03 | 4.03E-01 | Non-documented | Non-documented |
| *NELFE* | 6 | 2.43 | 7.65E-03 | 4.03E-01 | Non-documented | Non-documented |
| *C2* | 6 | 2.42 | 7.66E-03 | 4.03E-01 | Non-documented | Documented |
| *PSORS1C1* | 6 | 2.42 | 7.67E-03 | 4.03E-01 | Non-documented | Documented |
| *HR* | 8 | 2.42 | 7.67E-03 | 4.03E-01 | Non-documented | Non-documented |
| *EHMT2* | 6 | 2.42 | 7.73E-03 | 4.04E-01 | Non-documented | Non-documented |
| *PNPLA7* | 9 | 2.42 | 7.76E-03 | 4.04E-01 | Non-documented | Non-documented |
| *ACO1* | 9 | 2.42 | 7.77E-03 | 4.04E-01 | Non-documented | Non-documented |
| *OR1S1* | 11 | 2.42 | 7.79E-03 | 4.04E-01 | Non-documented | Non-documented |
| *GRAP* | 17 | 2.42 | 7.81E-03 | 4.04E-01 | Non-documented | Non-documented |
| *ATAD3A* | 1 | 2.42 | 7.86E-03 | 4.05E-01 | Non-documented | Non-documented |
| *KRTCAP3* | 2 | 2.41 | 7.88E-03 | 4.05E-01 | Non-documented | Non-documented |
| *CSNK2B* | 6 | 2.41 | 7.92E-03 | 4.05E-01 | Non-documented | Non-documented |
| *ZMIZ2* | 7 | 2.41 | 7.92E-03 | 4.05E-01 | Non-documented | Non-documented |
| *CCDC61* | 19 | 2.41 | 8.02E-03 | 4.09E-01 | Non-documented | Non-documented |
| *REEP3* | 10 | 2.41 | 8.03E-03 | 4.09E-01 | Non-documented | Non-documented |
| *GML* | 8 | 2.41 | 8.07E-03 | 4.10E-01 | Non-documented | Non-documented |
| *FOXD2* | 1 | 2.40 | 8.11E-03 | 4.10E-01 | Non-documented | Non-documented |
| *SIK1* | 21 | 2.40 | 8.12E-03 | 4.10E-01 | Non-documented | Non-documented |
| *HECTD3* | 1 | 2.40 | 8.14E-03 | 4.10E-01 | Non-documented | Non-documented |
| *USP36* | 17 | 2.40 | 8.17E-03 | 4.10E-01 | Non-documented | Non-documented |
| *OVOL3* | 19 | 2.40 | 8.20E-03 | 4.10E-01 | Non-documented | Non-documented |
| *CDC5L* | 6 | 2.40 | 8.22E-03 | 4.10E-01 | Non-documented | Non-documented |
| *JAKMIP1* | 4 | 2.40 | 8.30E-03 | 4.10E-01 | Non-documented | Non-documented |
| *SERINC3* | 20 | 2.40 | 8.30E-03 | 4.10E-01 | Non-documented | Non-documented |
| *OR2G2* | 1 | 2.39 | 8.35E-03 | 4.10E-01 | Non-documented | Non-documented |
| *RPH3AL* | 17 | 2.39 | 8.36E-03 | 4.10E-01 | Non-documented | Non-documented |
| *TOP1MT* | 8 | 2.39 | 8.37E-03 | 4.10E-01 | Non-documented | Non-documented |
| *KCNQ1* | 11 | 2.39 | 8.40E-03 | 4.10E-01 | Non-documented | Non-documented |
| *PTPN22* | 1 | 2.39 | 8.42E-03 | 4.10E-01 | Non-documented | Documented |
| *OR2H1* | 6 | 2.39 | 8.43E-03 | 4.10E-01 | Non-documented | Non-documented |
| *CNTN2* | 1 | 2.39 | 8.43E-03 | 4.10E-01 | Non-documented | Non-documented |
| *POU2AF1* | 11 | 2.39 | 8.47E-03 | 4.10E-01 | Non-documented | Non-documented |
| *GTF3C2* | 2 | 2.39 | 8.48E-03 | 4.10E-01 | Non-documented | Non-documented |
| *C17orf53* | 17 | 2.39 | 8.51E-03 | 4.10E-01 | Non-documented | Non-documented |
| *ST6GALNAC1* | 17 | 2.38 | 8.56E-03 | 4.10E-01 | Non-documented | Non-documented |
| *STX11* | 6 | 2.38 | 8.62E-03 | 4.10E-01 | Non-documented | Non-documented |
| *TMUB2* | 17 | 2.38 | 8.62E-03 | 4.10E-01 | Non-documented | Non-documented |
| *EIF2B4* | 2 | 2.38 | 8.63E-03 | 4.10E-01 | Non-documented | Non-documented |
| *MIB1* | 18 | 2.38 | 8.65E-03 | 4.10E-01 | Non-documented | Non-documented |
| *NRBP1* | 2 | 2.38 | 8.69E-03 | 4.10E-01 | Non-documented | Non-documented |
| *CDKN2A* | 9 | 2.38 | 8.70E-03 | 4.10E-01 | Non-documented | Documented |
| *CDYL* | 6 | 2.38 | 8.71E-03 | 4.10E-01 | Non-documented | Non-documented |
| *LCE3C* | 1 | 2.38 | 8.72E-03 | 4.10E-01 | Non-documented | Non-documented |
| *EGFL8* | 6 | 2.38 | 8.74E-03 | 4.10E-01 | Non-documented | Documented |
| *WDR62* | 19 | 2.38 | 8.75E-03 | 4.10E-01 | Non-documented | Non-documented |
| *TCF4* | 18 | 2.38 | 8.76E-03 | 4.10E-01 | Non-documented | Documented |
| *PRR4* | 12 | 2.37 | 8.78E-03 | 4.10E-01 | Non-documented | Non-documented |
| *JMJD1C* | 10 | 2.37 | 8.85E-03 | 4.13E-01 | Non-documented | Non-documented |
| *CPD* | 17 | 2.37 | 8.88E-03 | 4.13E-01 | Non-documented | Non-documented |
| *HDAC5* | 17 | 2.37 | 8.95E-03 | 4.15E-01 | Non-documented | Non-documented |
| *C5orf47* | 5 | 2.37 | 8.98E-03 | 4.16E-01 | Non-documented | Non-documented |
| *GOLT1A* | 1 | 2.36 | 9.06E-03 | 4.18E-01 | Non-documented | Non-documented |
| *TTPAL* | 20 | 2.36 | 9.08E-03 | 4.18E-01 | Non-documented | Non-documented |
| *GLYCTK* | 3 | 2.36 | 9.12E-03 | 4.18E-01 | Non-documented | Non-documented |
| *NEFM* | 8 | 2.36 | 9.13E-03 | 4.18E-01 | Non-documented | Non-documented |
| *ARRDC1* | 9 | 2.36 | 9.17E-03 | 4.18E-01 | Non-documented | Non-documented |
| *OVOL2* | 20 | 2.36 | 9.18E-03 | 4.18E-01 | Non-documented | Non-documented |
| *CYP11B2* | 8 | 2.36 | 9.18E-03 | 4.18E-01 | Non-documented | Documented |
| *LOC101927628* | 15 | 2.36 | 9.22E-03 | 4.18E-01 | Non-documented | Non-documented |
| *NR2F2* | 15 | 2.36 | 9.25E-03 | 4.19E-01 | Non-documented | Non-documented |
| *POLR2I* | 19 | 2.35 | 9.33E-03 | 4.19E-01 | Non-documented | Non-documented |
| *C4BPA* | 1 | 2.35 | 9.34E-03 | 4.19E-01 | Non-documented | Non-documented |
| *PPT2* | 6 | 2.35 | 9.36E-03 | 4.19E-01 | Non-documented | Documented |
| *MKNK2* | 19 | 2.35 | 9.36E-03 | 4.19E-01 | Non-documented | Non-documented |
| *CXCR2* | 2 | 2.35 | 9.45E-03 | 4.21E-01 | Non-documented | Non-documented |
| *MME* | 3 | 2.35 | 9.46E-03 | 4.21E-01 | Non-documented | Non-documented |
| *ZNF829* | 19 | 2.35 | 9.49E-03 | 4.21E-01 | Non-documented | Non-documented |
| *STK19* | 6 | 2.35 | 9.49E-03 | 4.21E-01 | Non-documented | Documented |
| *MRAS* | 3 | 2.34 | 9.54E-03 | 4.21E-01 | Non-documented | Non-documented |
| *ERCC8* | 5 | 2.34 | 9.54E-03 | 4.21E-01 | Non-documented | Non-documented |
| *KPNA1* | 3 | 2.34 | 9.56E-03 | 4.21E-01 | Non-documented | Non-documented |
| *PLEKHA6* | 1 | 2.34 | 9.59E-03 | 4.21E-01 | Non-documented | Documented |
| *FAM71D* | 14 | 2.34 | 9.63E-03 | 4.22E-01 | Non-documented | Non-documented |
| *GLP1R* | 6 | 2.34 | 9.67E-03 | 4.23E-01 | Non-documented | Non-documented |
| *PPM1G* | 2 | 2.34 | 9.71E-03 | 4.24E-01 | Non-documented | Non-documented |
| *CST9* | 20 | 2.34 | 9.77E-03 | 4.25E-01 | Non-documented | Non-documented |
| *COMMD1* | 2 | 2.33 | 9.89E-03 | 4.29E-01 | Non-documented | Non-documented |
| *ZNF629* | 16 | 2.33 | 9.90E-03 | 4.29E-01 | Non-documented | Non-documented |
| *SHANK2* | 11 | 2.33 | 1.00E-02 | 4.32E-01 | Non-documented | Non-documented |
| *CDC27* | 17 | 2.33 | 1.00E-02 | 4.32E-01 | Non-documented | Non-documented |
| *VIPAS39* | 14 | 2.32 | 1.01E-02 | 4.34E-01 | Non-documented | Non-documented |
| *HNF4A* | 20 | 2.32 | 1.01E-02 | 4.34E-01 | Non-documented | Documented |
| *AGPAT1* | 6 | 2.32 | 1.02E-02 | 4.34E-01 | Non-documented | Documented |
| *CTNNA3* | 10 | 2.32 | 1.02E-02 | 4.34E-01 | Non-documented | Documented |
| *ITGAD* | 16 | 2.32 | 1.02E-02 | 4.34E-01 | Non-documented | Non-documented |
| *NOXRED1* | 14 | 2.32 | 1.02E-02 | 4.34E-01 | Non-documented | Non-documented |
| *CST3* | 20 | 2.31 | 1.03E-02 | 4.37E-01 | Non-documented | Non-documented |
| *CCDC124* | 19 | 2.31 | 1.04E-02 | 4.37E-01 | Non-documented | Non-documented |
| *NSMCE2* | 8 | 2.31 | 1.04E-02 | 4.37E-01 | Non-documented | Non-documented |
| *STOML3* | 13 | 2.31 | 1.04E-02 | 4.37E-01 | Non-documented | Non-documented |
| *OR11A1* | 6 | 2.31 | 1.04E-02 | 4.37E-01 | Non-documented | Non-documented |
| *RIMS1* | 6 | 2.31 | 1.05E-02 | 4.37E-01 | Non-documented | Documented |
| *SNX17* | 2 | 2.31 | 1.05E-02 | 4.37E-01 | Non-documented | Non-documented |
| *ZNF558* | 19 | 2.31 | 1.05E-02 | 4.37E-01 | Non-documented | Non-documented |
| *OR10C1* | 6 | 2.31 | 1.05E-02 | 4.37E-01 | Non-documented | Non-documented |
| *KIAA0232* | 4 | 2.31 | 1.06E-02 | 4.37E-01 | Non-documented | Non-documented |
| *C6orf25* | 6 | 2.30 | 1.06E-02 | 4.37E-01 | Non-documented | Non-documented |
| *ZNF513* | 2 | 2.30 | 1.06E-02 | 4.37E-01 | Non-documented | Non-documented |
| *THAP8* | 19 | 2.30 | 1.06E-02 | 4.37E-01 | Non-documented | Non-documented |
| *ATF6* | 1 | 2.30 | 1.07E-02 | 4.37E-01 | Non-documented | Non-documented |
| *DOK2* | 8 | 2.30 | 1.07E-02 | 4.37E-01 | Non-documented | Documented |
| *MAGEF1* | 3 | 2.30 | 1.07E-02 | 4.37E-01 | Non-documented | Non-documented |
| *ANKK1* | 11 | 2.30 | 1.07E-02 | 4.37E-01 | Non-documented | Non-documented |
| *GABRA6* | 5 | 2.30 | 1.07E-02 | 4.37E-01 | Non-documented | Non-documented |
| *ZBTB46* | 20 | 2.30 | 1.07E-02 | 4.37E-01 | Non-documented | Documented |
| *CFB* | 6 | 2.30 | 1.08E-02 | 4.38E-01 | Non-documented | Documented |
| *ATXN7L3* | 17 | 2.30 | 1.08E-02 | 4.38E-01 | Non-documented | Non-documented |
| *E2F2* | 1 | 2.30 | 1.08E-02 | 4.38E-01 | Non-documented | Non-documented |
| *GGCT* | 7 | 2.30 | 1.08E-02 | 4.38E-01 | Non-documented | Non-documented |
| *CTLA4* | 2 | 2.30 | 1.09E-02 | 4.38E-01 | Reported | Documented |
| *CYP11B1* | 8 | 2.29 | 1.10E-02 | 4.43E-01 | Non-documented | Non-documented |
| *LARGE* | 22 | 2.28 | 1.12E-02 | 4.48E-01 | Non-documented | Non-documented |
| *KCNQ5* | 6 | 2.28 | 1.12E-02 | 4.49E-01 | Non-documented | Non-documented |
| *KCTD15* | 19 | 2.28 | 1.12E-02 | 4.49E-01 | Non-documented | Non-documented |
| *GMFB* | 14 | 2.28 | 1.13E-02 | 4.52E-01 | Non-documented | Non-documented |
| *OR1S2* | 11 | 2.28 | 1.14E-02 | 4.53E-01 | Non-documented | Non-documented |
| *DAGLA* | 11 | 2.28 | 1.14E-02 | 4.53E-01 | Non-documented | Non-documented |
| *TMEM119* | 12 | 2.27 | 1.15E-02 | 4.54E-01 | Non-documented | Non-documented |
| *YAF2* | 12 | 2.27 | 1.15E-02 | 4.56E-01 | Non-documented | Non-documented |
| *CYTH1* | 17 | 2.27 | 1.16E-02 | 4.59E-01 | Non-documented | Non-documented |
| *GLOD4* | 17 | 2.27 | 1.17E-02 | 4.59E-01 | Non-documented | Documented |
| *MAGEL2* | 15 | 2.26 | 1.18E-02 | 4.62E-01 | Non-documented | Non-documented |
| *CD302* | 2 | 2.26 | 1.18E-02 | 4.62E-01 | Non-documented | Non-documented |
| *BET1* | 7 | 2.26 | 1.19E-02 | 4.62E-01 | Non-documented | Documented |
| *AKIRIN1* | 1 | 2.26 | 1.19E-02 | 4.62E-01 | Non-documented | Non-documented |
| *CACNA1S* | 1 | 2.26 | 1.19E-02 | 4.62E-01 | Non-documented | Non-documented |
| *LOC101927572* | 19 | 2.26 | 1.19E-02 | 4.62E-01 | Non-documented | Non-documented |
| *KCNA4* | 11 | 2.26 | 1.20E-02 | 4.65E-01 | Non-documented | Non-documented |
| *PGLYRP1* | 19 | 2.25 | 1.21E-02 | 4.68E-01 | Non-documented | Non-documented |
| *PLEK2* | 14 | 2.25 | 1.22E-02 | 4.70E-01 | Non-documented | Non-documented |
| *MRPL33* | 2 | 2.25 | 1.23E-02 | 4.70E-01 | Non-documented | Non-documented |
| *SLC2A14* | 12 | 2.25 | 1.23E-02 | 4.70E-01 | Non-documented | Non-documented |
| *HSD17B14* | 19 | 2.25 | 1.23E-02 | 4.70E-01 | Non-documented | Non-documented |
| *DTWD1* | 15 | 2.25 | 1.23E-02 | 4.70E-01 | Non-documented | Documented |
| *TUBB2A* | 6 | 2.25 | 1.23E-02 | 4.70E-01 | Non-documented | Non-documented |
| *NMNAT2* | 1 | 2.25 | 1.24E-02 | 4.70E-01 | Non-documented | Non-documented |
| *RNMTL1* | 17 | 2.25 | 1.24E-02 | 4.70E-01 | Non-documented | Documented |
| *SERPINA5* | 14 | 2.24 | 1.24E-02 | 4.71E-01 | Non-documented | Non-documented |
| *DUPD1* | 10 | 2.24 | 1.24E-02 | 4.71E-01 | Non-documented | Non-documented |
| *NBEAL2* | 3 | 2.24 | 1.25E-02 | 4.71E-01 | Non-documented | Non-documented |
| *CEBPZ* | 2 | 2.24 | 1.25E-02 | 4.71E-01 | Non-documented | Non-documented |
| *WDR82* | 3 | 2.24 | 1.26E-02 | 4.74E-01 | Non-documented | Non-documented |
| *NFIB* | 9 | 2.24 | 1.26E-02 | 4.74E-01 | Non-documented | Non-documented |
| *TMEM240* | 1 | 2.24 | 1.26E-02 | 4.74E-01 | Non-documented | Non-documented |
| *STRA6* | 15 | 2.23 | 1.27E-02 | 4.75E-01 | Non-documented | Non-documented |
| *CD82* | 11 | 2.23 | 1.28E-02 | 4.75E-01 | Non-documented | Non-documented |
| *ASB2* | 14 | 2.23 | 1.28E-02 | 4.75E-01 | Non-documented | Non-documented |
| *CCDC78* | 16 | 2.23 | 1.28E-02 | 4.75E-01 | Non-documented | Non-documented |
| *FERMT2* | 14 | 2.23 | 1.28E-02 | 4.76E-01 | Non-documented | Documented |
| *CRADD* | 12 | 2.23 | 1.29E-02 | 4.76E-01 | Non-documented | Non-documented |
| *ANPEP* | 15 | 2.23 | 1.29E-02 | 4.76E-01 | Non-documented | Non-documented |
| *MXD3* | 5 | 2.23 | 1.29E-02 | 4.76E-01 | Non-documented | Non-documented |
| *PHF7* | 3 | 2.23 | 1.30E-02 | 4.77E-01 | Non-documented | Non-documented |
| *FAM173A* | 16 | 2.23 | 1.30E-02 | 4.77E-01 | Non-documented | Non-documented |
| *KRT79* | 12 | 2.23 | 1.30E-02 | 4.77E-01 | Non-documented | Non-documented |
| *BARX2* | 11 | 2.22 | 1.30E-02 | 4.77E-01 | Non-documented | Documented |
| *DNAH1* | 3 | 2.22 | 1.31E-02 | 4.77E-01 | Non-documented | Non-documented |
| *TEAD1* | 11 | 2.22 | 1.31E-02 | 4.77E-01 | Non-documented | Non-documented |
| *ZFHX2* | 14 | 2.22 | 1.31E-02 | 4.77E-01 | Non-documented | Non-documented |
| *SEC61A2* | 10 | 2.22 | 1.31E-02 | 4.77E-01 | Non-documented | Non-documented |
| *LOC101928268* | 7 | 2.22 | 1.32E-02 | 4.77E-01 | Non-documented | Non-documented |
| *C18orf54* | 18 | 2.22 | 1.34E-02 | 4.81E-01 | Non-documented | Non-documented |
| *ZRANB3* | 2 | 2.22 | 1.34E-02 | 4.81E-01 | Non-documented | Non-documented |
| *ACAN* | 15 | 2.22 | 1.34E-02 | 4.81E-01 | Non-documented | Non-documented |
| *VSIG10L* | 19 | 2.22 | 1.34E-02 | 4.81E-01 | Non-documented | Non-documented |
| *FAM64A* | 17 | 2.21 | 1.35E-02 | 4.83E-01 | Non-documented | Non-documented |
| *TENM4* | 11 | 2.21 | 1.35E-02 | 4.83E-01 | Non-documented | Documented |
| *PARP8* | 5 | 2.21 | 1.35E-02 | 4.83E-01 | Non-documented | Non-documented |
| *PCSK6* | 15 | 2.21 | 1.35E-02 | 4.83E-01 | Non-documented | Non-documented |
| *SERGEF* | 11 | 2.21 | 1.36E-02 | 4.83E-01 | Non-documented | Non-documented |
| *PSG11* | 19 | 2.21 | 1.36E-02 | 4.83E-01 | Non-documented | Non-documented |
| *DACT1* | 14 | 2.21 | 1.36E-02 | 4.83E-01 | Non-documented | Documented |
| *RNF5* | 6 | 2.21 | 1.37E-02 | 4.84E-01 | Non-documented | Documented |
| *GAB2* | 11 | 2.21 | 1.37E-02 | 4.84E-01 | Non-documented | Documented |
| *NUDT5* | 10 | 2.20 | 1.37E-02 | 4.85E-01 | Non-documented | Non-documented |
| *ZSCAN18* | 19 | 2.20 | 1.40E-02 | 4.89E-01 | Non-documented | Non-documented |
| *GPHA2* | 11 | 2.20 | 1.40E-02 | 4.89E-01 | Non-documented | Non-documented |
| *IQCB1* | 3 | 2.20 | 1.40E-02 | 4.89E-01 | Non-documented | Non-documented |
| *ZNF496* | 1 | 2.20 | 1.41E-02 | 4.89E-01 | Non-documented | Non-documented |
| *BCL7C* | 16 | 2.20 | 1.41E-02 | 4.89E-01 | Non-documented | Non-documented |
| *LY6G5B* | 6 | 2.19 | 1.41E-02 | 4.89E-01 | Non-documented | Non-documented |
| *TMEM39B* | 1 | 2.19 | 1.42E-02 | 4.89E-01 | Non-documented | Non-documented |
| *MICAL2* | 11 | 2.19 | 1.42E-02 | 4.89E-01 | Non-documented | Non-documented |
| *RASSF7* | 11 | 2.19 | 1.42E-02 | 4.89E-01 | Non-documented | Non-documented |
| *CLIP3* | 19 | 2.19 | 1.42E-02 | 4.89E-01 | Non-documented | Non-documented |
| *CPNE3* | 8 | 2.19 | 1.42E-02 | 4.89E-01 | Non-documented | Non-documented |
| *SETBP1* | 18 | 2.19 | 1.42E-02 | 4.89E-01 | Non-documented | Documented |
| *ZBTB12* | 6 | 2.19 | 1.42E-02 | 4.89E-01 | Non-documented | Non-documented |
| *DYNC2LI1* | 2 | 2.19 | 1.43E-02 | 4.89E-01 | Non-documented | Non-documented |
| *TNRC6C* | 17 | 2.19 | 1.43E-02 | 4.89E-01 | Non-documented | Non-documented |
| *MATK* | 19 | 2.19 | 1.44E-02 | 4.91E-01 | Non-documented | Non-documented |
| *ABCD2* | 12 | 2.19 | 1.44E-02 | 4.91E-01 | Non-documented | Non-documented |
| *PPIC* | 5 | 2.19 | 1.44E-02 | 4.91E-01 | Non-documented | Non-documented |
| *HLA-DRB1* | 6 | 2.18 | 1.45E-02 | 4.91E-01 | Non-documented | Documented |
| *PHRF1* | 11 | 2.18 | 1.45E-02 | 4.91E-01 | Non-documented | Non-documented |
| *RNF40* | 16 | 2.18 | 1.46E-02 | 4.93E-01 | Non-documented | Non-documented |
| *KCNE3* | 11 | 2.18 | 1.46E-02 | 4.93E-01 | Non-documented | Non-documented |
| *THTPA* | 14 | 2.18 | 1.47E-02 | 4.93E-01 | Non-documented | Non-documented |
| *ADAMTSL2* | 9 | 2.18 | 1.47E-02 | 4.93E-01 | Non-documented | Non-documented |
| *SAG* | 2 | 2.18 | 1.47E-02 | 4.93E-01 | Non-documented | Non-documented |
| *ADAM10* | 15 | 2.18 | 1.47E-02 | 4.93E-01 | Non-documented | Non-documented |
| *FAM107B* | 10 | 2.18 | 1.48E-02 | 4.97E-01 | Non-documented | Non-documented |
| *RNASE7* | 14 | 2.17 | 1.49E-02 | 4.99E-01 | Non-documented | Non-documented |
| *TMPRSS12* | 12 | 2.17 | 1.50E-02 | 4.99E-01 | Non-documented | Non-documented |
| *AHSA1* | 14 | 2.17 | 1.50E-02 | 4.99E-01 | Non-documented | Non-documented |
| *UBD* | 6 | 2.17 | 1.50E-02 | 4.99E-01 | Non-documented | Non-documented |
| *SEMA3G* | 3 | 2.17 | 1.51E-02 | 5.01E-01 | Non-documented | Non-documented |
| *GHRHR* | 7 | 2.17 | 1.52E-02 | 5.01E-01 | Non-documented | Non-documented |
| *AP4B1* | 1 | 2.16 | 1.52E-02 | 5.01E-01 | Non-documented | Documented |
| *AGER* | 6 | 2.16 | 1.52E-02 | 5.01E-01 | Non-documented | Documented |
| *TSPAN33* | 7 | 2.16 | 1.52E-02 | 5.01E-01 | Non-documented | Non-documented |
| *C10orf32* | 10 | 2.16 | 1.52E-02 | 5.01E-01 | Non-documented | Documented |
| *SUPT7L* | 2 | 2.16 | 1.52E-02 | 5.01E-01 | Non-documented | Non-documented |
| *LGALS4* | 19 | 2.16 | 1.53E-02 | 5.02E-01 | Non-documented | Non-documented |
| *NPAS2* | 2 | 2.16 | 1.54E-02 | 5.06E-01 | Non-documented | Non-documented |
| *LOC100129083* | 19 | 2.16 | 1.55E-02 | 5.07E-01 | Non-documented | Non-documented |
| *METRN* | 16 | 2.16 | 1.55E-02 | 5.07E-01 | Non-documented | Non-documented |
| *RTCB* | 22 | 2.16 | 1.56E-02 | 5.07E-01 | Non-documented | Non-documented |
| *DCAF12* | 9 | 2.15 | 1.56E-02 | 5.08E-01 | Non-documented | Documented |
| *PSG5* | 19 | 2.15 | 1.57E-02 | 5.10E-01 | Non-documented | Non-documented |
| *ENTPD8* | 9 | 2.15 | 1.58E-02 | 5.10E-01 | Non-documented | Non-documented |
| *HLA-DOB* | 6 | 2.15 | 1.58E-02 | 5.10E-01 | Non-documented | Documented |
| *OR52N5* | 11 | 2.15 | 1.58E-02 | 5.10E-01 | Non-documented | Non-documented |
| *NOTCH2* | 1 | 2.15 | 1.58E-02 | 5.10E-01 | Non-documented | Documented |
| *MROH1* | 8 | 2.15 | 1.59E-02 | 5.13E-01 | Non-documented | Non-documented |
| *C16orf72* | 16 | 2.15 | 1.60E-02 | 5.13E-01 | Non-documented | Documented |
| *MTG2* | 20 | 2.14 | 1.60E-02 | 5.13E-01 | Non-documented | Non-documented |
| *SORCS3* | 10 | 2.14 | 1.61E-02 | 5.14E-01 | Non-documented | Non-documented |
| *PF4V1* | 4 | 2.14 | 1.62E-02 | 5.16E-01 | Non-documented | Non-documented |
| *BNC1* | 15 | 2.14 | 1.62E-02 | 5.18E-01 | Non-documented | Non-documented |
| *CDC42BPA* | 1 | 2.14 | 1.63E-02 | 5.19E-01 | Non-documented | Non-documented |
| *NEFH* | 22 | 2.14 | 1.64E-02 | 5.20E-01 | Non-documented | Non-documented |
| *ILDR1* | 3 | 2.14 | 1.64E-02 | 5.20E-01 | Non-documented | Non-documented |
| *EIF2AK4* | 15 | 2.13 | 1.64E-02 | 5.20E-01 | Non-documented | Non-documented |
| *SEZ6L* | 22 | 2.13 | 1.65E-02 | 5.20E-01 | Non-documented | Non-documented |
| *TNN* | 1 | 2.13 | 1.65E-02 | 5.20E-01 | Non-documented | Non-documented |
| *LIN7A* | 12 | 2.13 | 1.65E-02 | 5.20E-01 | Non-documented | Non-documented |
| *OR12D3* | 6 | 2.13 | 1.65E-02 | 5.20E-01 | Non-documented | Non-documented |
| *ZBBX* | 3 | 2.13 | 1.66E-02 | 5.21E-01 | Non-documented | Documented |
| *PSMD6* | 3 | 2.13 | 1.66E-02 | 5.21E-01 | Non-documented | Non-documented |
| *SOX18* | 20 | 2.13 | 1.66E-02 | 5.21E-01 | Non-documented | Non-documented |
| *SFXN1* | 5 | 2.13 | 1.67E-02 | 5.22E-01 | Non-documented | Non-documented |
| *RB1* | 13 | 2.13 | 1.67E-02 | 5.22E-01 | Non-documented | Non-documented |
| *RXFP1* | 4 | 2.13 | 1.68E-02 | 5.22E-01 | Non-documented | Documented |
| *STXBP4* | 17 | 2.12 | 1.68E-02 | 5.24E-01 | Non-documented | Documented |
| *EBF1* | 5 | 2.12 | 1.70E-02 | 5.27E-01 | Non-documented | Documented |
| *COX20* | 1 | 2.12 | 1.70E-02 | 5.27E-01 | Non-documented | Non-documented |
| *PRMT8* | 12 | 2.12 | 1.70E-02 | 5.27E-01 | Non-documented | Non-documented |
| *BNIP2* | 15 | 2.12 | 1.71E-02 | 5.29E-01 | Non-documented | Non-documented |
| *TCERG1L* | 10 | 2.12 | 1.72E-02 | 5.29E-01 | Non-documented | Documented |
| *SAMD15* | 14 | 2.12 | 1.72E-02 | 5.29E-01 | Non-documented | Non-documented |
| *TFRC* | 3 | 2.11 | 1.73E-02 | 5.29E-01 | Non-documented | Documented |
| *HAGHL* | 16 | 2.11 | 1.73E-02 | 5.29E-01 | Non-documented | Non-documented |
| *EREG* | 4 | 2.11 | 1.73E-02 | 5.29E-01 | Non-documented | Non-documented |
| *LOC728392* | 17 | 2.11 | 1.73E-02 | 5.29E-01 | Non-documented | Non-documented |
| *CNNM1* | 10 | 2.11 | 1.74E-02 | 5.32E-01 | Non-documented | Documented |
| *CTRB1* | 16 | 2.11 | 1.75E-02 | 5.32E-01 | Non-documented | Documented |
| *ZNF514* | 2 | 2.11 | 1.75E-02 | 5.32E-01 | Non-documented | Non-documented |
| *C1D* | 2 | 2.11 | 1.75E-02 | 5.32E-01 | Non-documented | Documented |
| *FTSJ2* | 7 | 2.11 | 1.76E-02 | 5.33E-01 | Non-documented | Non-documented |
| *FAM214A* | 15 | 2.11 | 1.76E-02 | 5.33E-01 | Non-documented | Non-documented |
| *BICD1* | 12 | 2.10 | 1.77E-02 | 5.35E-01 | Non-documented | Documented |
| *SLC44A4* | 6 | 2.10 | 1.78E-02 | 5.35E-01 | Non-documented | Documented |
| *SLC25A45* | 11 | 2.10 | 1.78E-02 | 5.35E-01 | Non-documented | Non-documented |
| *OR10Q1* | 11 | 2.10 | 1.78E-02 | 5.35E-01 | Non-documented | Non-documented |
| *PRAME* | 22 | 2.10 | 1.78E-02 | 5.36E-01 | Non-documented | Non-documented |
| *SLC44A5* | 1 | 2.10 | 1.79E-02 | 5.36E-01 | Non-documented | Non-documented |
| *WASH1* | 9 | 2.10 | 1.79E-02 | 5.36E-01 | Non-documented | Non-documented |
| *DCN* | 12 | 2.10 | 1.79E-02 | 5.36E-01 | Non-documented | Non-documented |
| *ZDHHC3* | 3 | 2.10 | 1.80E-02 | 5.36E-01 | Non-documented | Non-documented |
| *HMX2* | 10 | 2.09 | 1.82E-02 | 5.42E-01 | Non-documented | Non-documented |
| *RANBP6* | 9 | 2.09 | 1.82E-02 | 5.42E-01 | Non-documented | Non-documented |
| *SLC35E4* | 22 | 2.09 | 1.83E-02 | 5.42E-01 | Non-documented | Non-documented |
| *NMBR* | 6 | 2.09 | 1.84E-02 | 5.46E-01 | Non-documented | Non-documented |
| *WBSCR17* | 7 | 2.09 | 1.84E-02 | 5.46E-01 | Non-documented | Documented |
| *CDKN3* | 14 | 2.09 | 1.85E-02 | 5.46E-01 | Non-documented | Documented |
| *NYAP2* | 2 | 2.08 | 1.86E-02 | 5.47E-01 | Non-documented | Documented |
| *MIS12* | 17 | 2.08 | 1.86E-02 | 5.47E-01 | Non-documented | Non-documented |
| *PRRX1* | 1 | 2.08 | 1.86E-02 | 5.47E-01 | Non-documented | Non-documented |
| *C1QTNF9B-AS1* | 13 | 2.08 | 1.87E-02 | 5.49E-01 | Non-documented | Non-documented |
| *C1QTNF9B* | 13 | 2.08 | 1.87E-02 | 5.49E-01 | Non-documented | Non-documented |
| *HNRNPC* | 14 | 2.08 | 1.88E-02 | 5.49E-01 | Non-documented | Non-documented |
| *STARD6* | 18 | 2.08 | 1.88E-02 | 5.49E-01 | Non-documented | Non-documented |
| *DPF2* | 11 | 2.08 | 1.89E-02 | 5.49E-01 | Non-documented | Non-documented |
| *SPDYE4* | 17 | 2.08 | 1.89E-02 | 5.49E-01 | Non-documented | Non-documented |
| *C14orf105* | 14 | 2.08 | 1.90E-02 | 5.49E-01 | Non-documented | Non-documented |
| *USP15* | 12 | 2.08 | 1.90E-02 | 5.49E-01 | Non-documented | Non-documented |
| *SMNDC1* | 10 | 2.08 | 1.90E-02 | 5.49E-01 | Non-documented | Non-documented |
| *CASZ1* | 1 | 2.08 | 1.90E-02 | 5.49E-01 | Non-documented | Non-documented |
| *TMEM81* | 1 | 2.08 | 1.90E-02 | 5.49E-01 | Non-documented | Non-documented |
| *ASAP3* | 1 | 2.07 | 1.91E-02 | 5.49E-01 | Non-documented | Non-documented |
| *LGI1* | 10 | 2.07 | 1.91E-02 | 5.49E-01 | Non-documented | Non-documented |
| *C11orf85* | 11 | 2.07 | 1.91E-02 | 5.49E-01 | Non-documented | Non-documented |
| *ZNF485* | 10 | 2.07 | 1.91E-02 | 5.49E-01 | Non-documented | Documented |
| *GNAL* | 18 | 2.07 | 1.91E-02 | 5.49E-01 | Non-documented | Non-documented |
| *TMEM130* | 7 | 2.07 | 1.93E-02 | 5.52E-01 | Non-documented | Non-documented |
| *GOSR1* | 17 | 2.07 | 1.93E-02 | 5.52E-01 | Non-documented | Non-documented |
| *THOC5* | 22 | 2.07 | 1.94E-02 | 5.52E-01 | Non-documented | Non-documented |
| *RAX2* | 19 | 2.07 | 1.94E-02 | 5.52E-01 | Non-documented | Non-documented |
| *TM2D3* | 15 | 2.07 | 1.94E-02 | 5.52E-01 | Non-documented | Non-documented |
| *AGO1* | 1 | 2.06 | 1.95E-02 | 5.52E-01 | Non-documented | Non-documented |
| *MAN2A2* | 15 | 2.06 | 1.95E-02 | 5.52E-01 | Non-documented | Non-documented |
| *ADGRG1* | 16 | 2.06 | 1.95E-02 | 5.52E-01 | Non-documented | Non-documented |
| *PEG3* | 19 | 2.06 | 1.96E-02 | 5.52E-01 | Non-documented | Non-documented |
| *ZIM2* | 19 | 2.06 | 1.96E-02 | 5.52E-01 | Non-documented | Non-documented |
| *INSIG2* | 2 | 2.06 | 1.96E-02 | 5.52E-01 | Non-documented | Non-documented |
| *PCNXL3* | 11 | 2.06 | 1.98E-02 | 5.55E-01 | Non-documented | Non-documented |
| *TIMP2* | 17 | 2.06 | 1.98E-02 | 5.55E-01 | Non-documented | Non-documented |
| *ETFB* | 19 | 2.06 | 1.98E-02 | 5.55E-01 | Non-documented | Non-documented |
| *HNRNPU* | 1 | 2.06 | 1.99E-02 | 5.55E-01 | Non-documented | Non-documented |
| *MAL* | 2 | 2.06 | 1.99E-02 | 5.55E-01 | Non-documented | Non-documented |
| *KLF10* | 8 | 2.06 | 1.99E-02 | 5.55E-01 | Non-documented | Non-documented |
| *CASP1* | 11 | 2.05 | 2.00E-02 | 5.55E-01 | Non-documented | Non-documented |
| *GTF2A2* | 15 | 2.05 | 2.00E-02 | 5.55E-01 | Non-documented | Non-documented |
| *PRRT1* | 6 | 2.05 | 2.00E-02 | 5.55E-01 | Non-documented | Documented |
| *FXYD2* | 11 | 2.05 | 2.00E-02 | 5.55E-01 | Non-documented | Non-documented |
| *TRIP12* | 2 | 2.05 | 2.00E-02 | 5.55E-01 | Non-documented | Non-documented |
| *CMTM6* | 3 | 2.05 | 2.01E-02 | 5.55E-01 | Non-documented | Non-documented |
| *AGO4* | 1 | 2.05 | 2.01E-02 | 5.55E-01 | Non-documented | Non-documented |
| *TBCA* | 5 | 2.05 | 2.01E-02 | 5.55E-01 | Non-documented | Non-documented |
| *HIST1H2AH* | 6 | 2.05 | 2.02E-02 | 5.55E-01 | Non-documented | Non-documented |
| *SIGLEC1* | 20 | 2.05 | 2.03E-02 | 5.58E-01 | Non-documented | Non-documented |
| *GOT1* | 10 | 2.05 | 2.04E-02 | 5.61E-01 | Non-documented | Non-documented |
| *KCNF1* | 2 | 2.04 | 2.04E-02 | 5.61E-01 | Non-documented | Non-documented |
| *TCN2* | 22 | 2.04 | 2.05E-02 | 5.61E-01 | Non-documented | Non-documented |
| *EXOC2* | 6 | 2.04 | 2.06E-02 | 5.61E-01 | Non-documented | Documented |
| *MEX3B* | 15 | 2.04 | 2.06E-02 | 5.61E-01 | Non-documented | Documented |
| *DMAP1* | 1 | 2.04 | 2.06E-02 | 5.61E-01 | Non-documented | Non-documented |
| *RPP30* | 10 | 2.04 | 2.06E-02 | 5.61E-01 | Non-documented | Non-documented |
| *CEP295NL* | 17 | 2.04 | 2.07E-02 | 5.62E-01 | Non-documented | Non-documented |
| *POLA2* | 11 | 2.04 | 2.08E-02 | 5.63E-01 | Non-documented | Non-documented |
| *RAB3C* | 5 | 2.04 | 2.08E-02 | 5.64E-01 | Non-documented | Documented |
| *LETMD1* | 12 | 2.04 | 2.09E-02 | 5.64E-01 | Non-documented | Non-documented |
| *DCP1B* | 12 | 2.04 | 2.09E-02 | 5.64E-01 | Non-documented | Documented |
| *STAMBP* | 2 | 2.03 | 2.10E-02 | 5.64E-01 | Non-documented | Non-documented |
| *BRD1* | 22 | 2.03 | 2.10E-02 | 5.64E-01 | Non-documented | Non-documented |
| *DMBT1* | 10 | 2.03 | 2.10E-02 | 5.64E-01 | Non-documented | Non-documented |
| *YEATS2* | 3 | 2.03 | 2.11E-02 | 5.64E-01 | Non-documented | Non-documented |
| *SLC35D2* | 9 | 2.03 | 2.11E-02 | 5.64E-01 | Non-documented | Documented |
| *TYW3* | 1 | 2.03 | 2.12E-02 | 5.64E-01 | Non-documented | Non-documented |
| *MICB* | 6 | 2.03 | 2.12E-02 | 5.64E-01 | Reported | Documented |
| *MTHFSD* | 16 | 2.03 | 2.12E-02 | 5.64E-01 | Non-documented | Non-documented |
| *EPHB4* | 7 | 2.03 | 2.12E-02 | 5.64E-01 | Non-documented | Non-documented |
| *RAVER2* | 1 | 2.03 | 2.12E-02 | 5.64E-01 | Non-documented | Non-documented |
| *BCAR1* | 16 | 2.03 | 2.12E-02 | 5.64E-01 | Non-documented | Documented |
| *MKI67* | 10 | 2.03 | 2.13E-02 | 5.64E-01 | Non-documented | Non-documented |
| *TAS2R7* | 12 | 2.03 | 2.13E-02 | 5.64E-01 | Non-documented | Non-documented |
| *TIGD3* | 11 | 2.03 | 2.13E-02 | 5.64E-01 | Non-documented | Non-documented |
| *GABRA2* | 4 | 2.03 | 2.14E-02 | 5.65E-01 | Non-documented | Non-documented |
| *NOD1* | 7 | 2.03 | 2.14E-02 | 5.65E-01 | Non-documented | Non-documented |
| *ACADSB* | 10 | 2.02 | 2.15E-02 | 5.65E-01 | Non-documented | Non-documented |
| *EIF4E1B* | 5 | 2.02 | 2.16E-02 | 5.65E-01 | Non-documented | Non-documented |
| *KIAA2026* | 9 | 2.02 | 2.16E-02 | 5.65E-01 | Non-documented | Non-documented |
| *HAVCR1* | 5 | 2.02 | 2.16E-02 | 5.65E-01 | Non-documented | Non-documented |
| *EIF5AL1* | 10 | 2.02 | 2.16E-02 | 5.65E-01 | Non-documented | Non-documented |
| *WBSCR27* | 7 | 2.02 | 2.16E-02 | 5.65E-01 | Non-documented | Non-documented |
| *SLC47A1* | 17 | 2.02 | 2.16E-02 | 5.65E-01 | Non-documented | Non-documented |
| *GUCA1C* | 3 | 2.02 | 2.17E-02 | 5.65E-01 | Non-documented | Non-documented |
| *DEFA1B* | 8 | 2.02 | 2.17E-02 | 5.65E-01 | Non-documented | Non-documented |
| *NRP2* | 2 | 2.02 | 2.18E-02 | 5.68E-01 | Non-documented | Non-documented |
| *ACHE* | 7 | 2.02 | 2.19E-02 | 5.68E-01 | Non-documented | Non-documented |
| *WNT10A* | 2 | 2.02 | 2.19E-02 | 5.68E-01 | Non-documented | Non-documented |
| *FAM124B* | 2 | 2.02 | 2.19E-02 | 5.68E-01 | Non-documented | Documented |
| *TRIM39-RPP21* | 6 | 2.01 | 2.20E-02 | 5.68E-01 | Non-documented | Documented |
| *SPIRE1* | 18 | 2.01 | 2.20E-02 | 5.68E-01 | Non-documented | Non-documented |
| *RCN1* | 11 | 2.01 | 2.20E-02 | 5.68E-01 | Non-documented | Non-documented |
| *TAP2* | 6 | 2.01 | 2.21E-02 | 5.68E-01 | Non-documented | Non-documented |
| *TMED8* | 14 | 2.01 | 2.21E-02 | 5.68E-01 | Non-documented | Non-documented |
| *NME9* | 3 | 2.01 | 2.21E-02 | 5.68E-01 | Non-documented | Non-documented |
| *SLC16A6* | 17 | 2.01 | 2.22E-02 | 5.69E-01 | Non-documented | Non-documented |
| *CAPRIN2* | 12 | 2.01 | 2.22E-02 | 5.69E-01 | Non-documented | Non-documented |
| *SNX6* | 14 | 2.01 | 2.23E-02 | 5.69E-01 | Non-documented | Non-documented |
| *FMO5* | 1 | 2.01 | 2.24E-02 | 5.69E-01 | Non-documented | Non-documented |
| *PDE6C* | 10 | 2.01 | 2.24E-02 | 5.69E-01 | Non-documented | Non-documented |
| *NOXA1* | 9 | 2.01 | 2.25E-02 | 5.69E-01 | Non-documented | Non-documented |
| *NARS2* | 11 | 2.00 | 2.26E-02 | 5.69E-01 | Non-documented | Non-documented |
| *RFK* | 9 | 2.00 | 2.26E-02 | 5.69E-01 | Non-documented | Non-documented |
| *CSRNP2* | 12 | 2.00 | 2.27E-02 | 5.69E-01 | Non-documented | Non-documented |
| *ACTBL2* | 5 | 2.00 | 2.27E-02 | 5.69E-01 | Non-documented | Documented |
| *SNRPE* | 1 | 2.00 | 2.27E-02 | 5.69E-01 | Non-documented | Non-documented |
| *WDR37* | 10 | 2.00 | 2.27E-02 | 5.69E-01 | Non-documented | Non-documented |
| *MON2* | 12 | 2.00 | 2.27E-02 | 5.69E-01 | Non-documented | Non-documented |
| *UBE2V2* | 8 | 2.00 | 2.28E-02 | 5.69E-01 | Non-documented | Non-documented |
| *SHB* | 9 | 2.00 | 2.28E-02 | 5.69E-01 | Non-documented | Non-documented |
| *FZD1* | 7 | 2.00 | 2.28E-02 | 5.69E-01 | Non-documented | Non-documented |
| *LOC199882* | 1 | 2.00 | 2.28E-02 | 5.69E-01 | Non-documented | Non-documented |
| *ATXN7* | 3 | 2.00 | 2.29E-02 | 5.69E-01 | Non-documented | Non-documented |
| *MRGPRX1* | 11 | 2.00 | 2.29E-02 | 5.69E-01 | Non-documented | Non-documented |
| *LMX1B* | 9 | 2.00 | 2.29E-02 | 5.69E-01 | Non-documented | Documented |
| *KLHDC7A* | 1 | 2.00 | 2.29E-02 | 5.69E-01 | Non-documented | Documented |
| *EPS8L2* | 11 | 2.00 | 2.29E-02 | 5.69E-01 | Non-documented | Non-documented |
| *ACOX2* | 3 | 2.00 | 2.30E-02 | 5.69E-01 | Non-documented | Non-documented |
| *NOVA2* | 19 | 2.00 | 2.30E-02 | 5.69E-01 | Non-documented | Non-documented |
| *42800* | 5 | 2.00 | 2.30E-02 | 5.69E-01 | Non-documented | Non-documented |
| *GPHN* | 14 | 2.00 | 2.30E-02 | 5.69E-01 | Non-documented | Non-documented |
| *RASL11B* | 4 | 1.99 | 2.30E-02 | 5.69E-01 | Non-documented | Non-documented |
| *HIGD1B* | 17 | 1.99 | 2.30E-02 | 5.69E-01 | Non-documented | Non-documented |
| *KLRG2* | 7 | 1.99 | 2.30E-02 | 5.69E-01 | Non-documented | Non-documented |
| *ADCY5* | 3 | 1.99 | 2.31E-02 | 5.70E-01 | Non-documented | Non-documented |
| *CYP17A1* | 10 | 1.99 | 2.32E-02 | 5.70E-01 | Non-documented | Documented |
| *PPM1M* | 3 | 1.99 | 2.32E-02 | 5.70E-01 | Non-documented | Non-documented |
| *NEDD4L* | 18 | 1.99 | 2.32E-02 | 5.70E-01 | Non-documented | Non-documented |
| *POU2F3* | 11 | 1.99 | 2.33E-02 | 5.70E-01 | Non-documented | Non-documented |
| *CDC42EP2* | 11 | 1.99 | 2.33E-02 | 5.70E-01 | Non-documented | Non-documented |
| *TM6SF1* | 15 | 1.99 | 2.33E-02 | 5.71E-01 | Non-documented | Non-documented |
| *LPAR6* | 13 | 1.99 | 2.34E-02 | 5.72E-01 | Non-documented | Non-documented |
| *MAP3K11* | 11 | 1.99 | 2.35E-02 | 5.72E-01 | Non-documented | Non-documented |
| *HDGFRP3* | 15 | 1.99 | 2.35E-02 | 5.72E-01 | Non-documented | Non-documented |
| *GJC1* | 17 | 1.99 | 2.35E-02 | 5.72E-01 | Non-documented | Non-documented |
| *ST6GALNAC2* | 17 | 1.99 | 2.36E-02 | 5.72E-01 | Non-documented | Non-documented |
| *PLD6* | 17 | 1.99 | 2.36E-02 | 5.72E-01 | Non-documented | Non-documented |
| *TMEM106C* | 12 | 1.98 | 2.36E-02 | 5.72E-01 | Non-documented | Non-documented |
| *NPHP4* | 1 | 1.98 | 2.37E-02 | 5.72E-01 | Non-documented | Non-documented |
| *PTAFR* | 1 | 1.98 | 2.38E-02 | 5.72E-01 | Non-documented | Non-documented |
| *IDI1* | 10 | 1.98 | 2.38E-02 | 5.72E-01 | Non-documented | Non-documented |
| *ARHGEF26* | 3 | 1.98 | 2.39E-02 | 5.72E-01 | Non-documented | Non-documented |
| *BCL2L14* | 12 | 1.98 | 2.39E-02 | 5.72E-01 | Non-documented | Non-documented |
| *SCCPDH* | 1 | 1.98 | 2.39E-02 | 5.72E-01 | Non-documented | Non-documented |
| *ERI3* | 1 | 1.98 | 2.40E-02 | 5.72E-01 | Non-documented | Non-documented |
| *SLC13A3* | 20 | 1.98 | 2.40E-02 | 5.72E-01 | Non-documented | Non-documented |
| *NISCH* | 3 | 1.98 | 2.40E-02 | 5.72E-01 | Non-documented | Non-documented |
| *SUSD2* | 22 | 1.98 | 2.41E-02 | 5.72E-01 | Non-documented | Non-documented |
| *GNA13* | 17 | 1.98 | 2.41E-02 | 5.72E-01 | Non-documented | Non-documented |
| *IGSF5* | 21 | 1.98 | 2.41E-02 | 5.72E-01 | Non-documented | Non-documented |
| *SLCO2B1* | 11 | 1.98 | 2.41E-02 | 5.72E-01 | Non-documented | Non-documented |
| *DAB2IP* | 9 | 1.97 | 2.42E-02 | 5.72E-01 | Non-documented | Documented |
| *CHDH* | 3 | 1.97 | 2.42E-02 | 5.72E-01 | Non-documented | Non-documented |
| *RINL* | 19 | 1.97 | 2.42E-02 | 5.72E-01 | Non-documented | Non-documented |
| *KPNA6* | 1 | 1.97 | 2.42E-02 | 5.72E-01 | Non-documented | Non-documented |
| *SRP14* | 15 | 1.97 | 2.42E-02 | 5.72E-01 | Non-documented | Non-documented |
| *GXYLT1* | 12 | 1.97 | 2.42E-02 | 5.72E-01 | Non-documented | Non-documented |
| *CST1* | 20 | 1.97 | 2.42E-02 | 5.72E-01 | Non-documented | Non-documented |
| *STAB1* | 3 | 1.97 | 2.43E-02 | 5.72E-01 | Non-documented | Non-documented |
| *ACTG2* | 2 | 1.97 | 2.44E-02 | 5.75E-01 | Non-documented | Non-documented |
| *VCL* | 10 | 1.97 | 2.45E-02 | 5.75E-01 | Non-documented | Non-documented |
| *PPP2R5B* | 11 | 1.97 | 2.45E-02 | 5.75E-01 | Non-documented | Non-documented |
| *RND3* | 2 | 1.97 | 2.45E-02 | 5.75E-01 | Non-documented | Documented |
| *LSM8* | 7 | 1.97 | 2.46E-02 | 5.75E-01 | Non-documented | Non-documented |
| *ABCG5* | 2 | 1.97 | 2.46E-02 | 5.75E-01 | Non-documented | Non-documented |
| *QDPR* | 4 | 1.97 | 2.46E-02 | 5.75E-01 | Non-documented | Non-documented |
| *DMXL1* | 5 | 1.97 | 2.46E-02 | 5.75E-01 | Non-documented | Non-documented |
| *EGR4* | 2 | 1.97 | 2.47E-02 | 5.75E-01 | Non-documented | Non-documented |
| *NAMPT* | 7 | 1.97 | 2.47E-02 | 5.75E-01 | Non-documented | Non-documented |
| *RAB3IL1* | 11 | 1.96 | 2.48E-02 | 5.76E-01 | Non-documented | Non-documented |
| *INADL* | 1 | 1.96 | 2.48E-02 | 5.77E-01 | Non-documented | Non-documented |
| *ANK1* | 8 | 1.96 | 2.49E-02 | 5.77E-01 | Non-documented | Non-documented |
| *CCDC106* | 19 | 1.96 | 2.49E-02 | 5.77E-01 | Non-documented | Non-documented |
| *TNNC1* | 3 | 1.96 | 2.52E-02 | 5.82E-01 | Non-documented | Non-documented |
| *BMP2* | 20 | 1.96 | 2.53E-02 | 5.84E-01 | Non-documented | Documented |
| *CLDN4* | 7 | 1.95 | 2.53E-02 | 5.84E-01 | Non-documented | Non-documented |
| *OR9Q2* | 11 | 1.95 | 2.54E-02 | 5.85E-01 | Non-documented | Non-documented |
| *TAS2R9* | 12 | 1.95 | 2.54E-02 | 5.85E-01 | Non-documented | Non-documented |
| *MRPS5* | 2 | 1.95 | 2.55E-02 | 5.85E-01 | Non-documented | Non-documented |
| *UST* | 6 | 1.95 | 2.55E-02 | 5.85E-01 | Non-documented | Non-documented |
| *UFSP1* | 7 | 1.95 | 2.55E-02 | 5.85E-01 | Non-documented | Non-documented |
| *OR2C3* | 1 | 1.95 | 2.55E-02 | 5.85E-01 | Non-documented | Documented |
| *SS18L1* | 20 | 1.95 | 2.56E-02 | 5.85E-01 | Non-documented | Non-documented |
| *GOLGA2* | 9 | 1.95 | 2.56E-02 | 5.85E-01 | Non-documented | Non-documented |
| *NPAS3* | 14 | 1.95 | 2.57E-02 | 5.85E-01 | Non-documented | Documented |
| *DHX16* | 6 | 1.95 | 2.57E-02 | 5.85E-01 | Non-documented | Non-documented |
| *PITX1* | 5 | 1.95 | 2.57E-02 | 5.85E-01 | Non-documented | Documented |
| *STYX* | 14 | 1.95 | 2.57E-02 | 5.85E-01 | Non-documented | Documented |
| *PROM2* | 2 | 1.95 | 2.57E-02 | 5.85E-01 | Non-documented | Non-documented |
| *BHLHA9* | 17 | 1.95 | 2.58E-02 | 5.86E-01 | Non-documented | Non-documented |
| *PRKAG1* | 12 | 1.95 | 2.59E-02 | 5.86E-01 | Non-documented | Non-documented |
| *ACTRT2* | 1 | 1.94 | 2.60E-02 | 5.86E-01 | Non-documented | Documented |
| *C11orf96* | 11 | 1.94 | 2.60E-02 | 5.86E-01 | Non-documented | Non-documented |
| *CCDC58* | 3 | 1.94 | 2.60E-02 | 5.86E-01 | Non-documented | Non-documented |
| *SNX10* | 7 | 1.94 | 2.61E-02 | 5.86E-01 | Non-documented | Non-documented |
| *CLSPN* | 1 | 1.94 | 2.62E-02 | 5.86E-01 | Non-documented | Non-documented |
| *PRICKLE1* | 12 | 1.94 | 2.62E-02 | 5.86E-01 | Non-documented | Documented |
| *RPL26L1* | 5 | 1.94 | 2.62E-02 | 5.86E-01 | Non-documented | Non-documented |
| *EHMT1* | 9 | 1.94 | 2.62E-02 | 5.86E-01 | Non-documented | Non-documented |
| *KCNK7* | 11 | 1.94 | 2.62E-02 | 5.86E-01 | Non-documented | Non-documented |
| *FAM175A* | 4 | 1.94 | 2.62E-02 | 5.86E-01 | Reported | Non-documented |
| *LOC149373* | 1 | 1.94 | 2.62E-02 | 5.86E-01 | Non-documented | Non-documented |
| *OTOGL* | 12 | 1.94 | 2.63E-02 | 5.86E-01 | Non-documented | Non-documented |
| *CYP2U1* | 4 | 1.94 | 2.64E-02 | 5.86E-01 | Non-documented | Non-documented |
| *MAD1L1* | 7 | 1.94 | 2.64E-02 | 5.86E-01 | Non-documented | Documented |
| *MFSD6L* | 17 | 1.94 | 2.64E-02 | 5.86E-01 | Non-documented | Non-documented |
| *CNTN3* | 3 | 1.94 | 2.64E-02 | 5.86E-01 | Non-documented | Non-documented |
| *TRAF3IP2* | 6 | 1.94 | 2.64E-02 | 5.86E-01 | Non-documented | Non-documented |
| *BEST1* | 11 | 1.94 | 2.65E-02 | 5.86E-01 | Non-documented | Non-documented |
| *DAOA* | 13 | 1.93 | 2.65E-02 | 5.87E-01 | Non-documented | Non-documented |
| *MOS* | 8 | 1.93 | 2.66E-02 | 5.87E-01 | Non-documented | Non-documented |
| *KMT2D* | 12 | 1.93 | 2.66E-02 | 5.87E-01 | Non-documented | Non-documented |
| *MRPS18C* | 4 | 1.93 | 2.66E-02 | 5.87E-01 | Non-documented | Non-documented |
| *PPP1R18* | 6 | 1.93 | 2.67E-02 | 5.87E-01 | Non-documented | Documented |
| *LARP4* | 12 | 1.93 | 2.67E-02 | 5.87E-01 | Non-documented | Non-documented |
| *CMTM3* | 16 | 1.93 | 2.67E-02 | 5.87E-01 | Non-documented | Non-documented |
| *NRM* | 6 | 1.93 | 2.67E-02 | 5.87E-01 | Non-documented | Non-documented |
| *LLGL2* | 17 | 1.93 | 2.67E-02 | 5.87E-01 | Non-documented | Non-documented |
| *MBD3* | 19 | 1.93 | 2.68E-02 | 5.87E-01 | Non-documented | Non-documented |
| *C1S* | 12 | 1.93 | 2.69E-02 | 5.88E-01 | Non-documented | Documented |
| *TFEB* | 6 | 1.93 | 2.69E-02 | 5.88E-01 | Non-documented | Documented |
| *PRDM5* | 4 | 1.93 | 2.70E-02 | 5.88E-01 | Non-documented | Non-documented |
| *MACROD2* | 20 | 1.93 | 2.70E-02 | 5.88E-01 | Non-documented | Non-documented |
| *HIST1H2BK* | 6 | 1.92 | 2.72E-02 | 5.92E-01 | Non-documented | Non-documented |
| *KIF28P* | 1 | 1.92 | 2.74E-02 | 5.96E-01 | Non-documented | Non-documented |
| *CPO* | 2 | 1.92 | 2.74E-02 | 5.96E-01 | Non-documented | Non-documented |
| *SYNE4* | 19 | 1.92 | 2.75E-02 | 5.96E-01 | Non-documented | Non-documented |
| *TCEA2* | 20 | 1.92 | 2.76E-02 | 5.98E-01 | Non-documented | Non-documented |
| *MRPL41* | 9 | 1.92 | 2.77E-02 | 5.98E-01 | Non-documented | Non-documented |
| *C4orf46* | 4 | 1.92 | 2.77E-02 | 5.98E-01 | Non-documented | Non-documented |
| *ABHD6* | 3 | 1.91 | 2.78E-02 | 5.98E-01 | Non-documented | Non-documented |
| *ZNF490* | 19 | 1.91 | 2.78E-02 | 5.98E-01 | Non-documented | Non-documented |
| *SNRPF* | 12 | 1.91 | 2.78E-02 | 5.98E-01 | Non-documented | Non-documented |
| *SCAP* | 3 | 1.91 | 2.79E-02 | 5.98E-01 | Non-documented | Non-documented |
| *HLA-A* | 6 | 1.91 | 2.79E-02 | 5.98E-01 | Non-documented | Documented |
| *HMP19* | 5 | 1.91 | 2.79E-02 | 5.98E-01 | Non-documented | Documented |
| *OR6C6* | 12 | 1.91 | 2.79E-02 | 5.98E-01 | Non-documented | Non-documented |
| *HLA-DRA* | 6 | 1.91 | 2.80E-02 | 5.98E-01 | Non-documented | Documented |
| *RLTPR* | 16 | 1.91 | 2.80E-02 | 5.98E-01 | Non-documented | Non-documented |
| *MAP4K1* | 19 | 1.91 | 2.80E-02 | 5.98E-01 | Non-documented | Non-documented |
| *ADCY6* | 12 | 1.91 | 2.81E-02 | 5.98E-01 | Non-documented | Non-documented |
| *RHBDL2* | 1 | 1.91 | 2.81E-02 | 5.98E-01 | Non-documented | Non-documented |
| *KIFAP3* | 1 | 1.91 | 2.82E-02 | 5.98E-01 | Non-documented | Non-documented |
| *MMRN1* | 4 | 1.91 | 2.82E-02 | 5.98E-01 | Non-documented | Non-documented |
| *ZGLP1* | 19 | 1.91 | 2.82E-02 | 5.98E-01 | Non-documented | Non-documented |
| *CDHR5* | 11 | 1.91 | 2.83E-02 | 5.98E-01 | Non-documented | Non-documented |
| *LPPR1* | 9 | 1.91 | 2.84E-02 | 5.98E-01 | Non-documented | Non-documented |
| *KIAA1211L* | 2 | 1.90 | 2.84E-02 | 5.98E-01 | Non-documented | Non-documented |
| *LY6G5C* | 6 | 1.90 | 2.84E-02 | 5.98E-01 | Non-documented | Non-documented |
| *EFCAB9* | 5 | 1.90 | 2.84E-02 | 5.98E-01 | Non-documented | Non-documented |
| *ADRA1B* | 5 | 1.90 | 2.84E-02 | 5.98E-01 | Non-documented | Non-documented |
| *PES1* | 22 | 1.90 | 2.85E-02 | 5.98E-01 | Non-documented | Non-documented |
| *TRIM39* | 6 | 1.90 | 2.85E-02 | 5.98E-01 | Non-documented | Non-documented |
| *CARD16* | 11 | 1.90 | 2.85E-02 | 5.98E-01 | Non-documented | Non-documented |
| *MESP2* | 15 | 1.90 | 2.86E-02 | 5.98E-01 | Non-documented | Non-documented |
| *PRR31* | 9 | 1.90 | 2.86E-02 | 5.98E-01 | Non-documented | Non-documented |
| *GGACT* | 13 | 1.90 | 2.86E-02 | 5.98E-01 | Non-documented | Non-documented |
| *SDE2* | 1 | 1.90 | 2.86E-02 | 5.98E-01 | Non-documented | Non-documented |
| *SETD2* | 3 | 1.90 | 2.87E-02 | 6.00E-01 | Non-documented | Non-documented |
| *SRRT* | 7 | 1.90 | 2.88E-02 | 6.01E-01 | Non-documented | Non-documented |
| *CMTM2* | 16 | 1.90 | 2.88E-02 | 6.01E-01 | Non-documented | Non-documented |
| *PTGDS* | 9 | 1.90 | 2.88E-02 | 6.01E-01 | Non-documented | Non-documented |
| *SIGLEC10* | 19 | 1.90 | 2.90E-02 | 6.03E-01 | Non-documented | Non-documented |
| *TRIP6* | 7 | 1.89 | 2.91E-02 | 6.03E-01 | Non-documented | Non-documented |
| *UBA52* | 19 | 1.89 | 2.91E-02 | 6.03E-01 | Non-documented | Non-documented |
| *SLC9A2* | 2 | 1.89 | 2.91E-02 | 6.03E-01 | Non-documented | Non-documented |
| *PBX4* | 19 | 1.89 | 2.91E-02 | 6.03E-01 | Non-documented | Non-documented |
| *DERL2* | 17 | 1.89 | 2.91E-02 | 6.03E-01 | Non-documented | Non-documented |
| *DPPA3* | 12 | 1.89 | 2.93E-02 | 6.05E-01 | Non-documented | Non-documented |
| *UBE3A* | 15 | 1.89 | 2.93E-02 | 6.05E-01 | Non-documented | Non-documented |
| *CDH23* | 10 | 1.89 | 2.93E-02 | 6.05E-01 | Non-documented | Non-documented |
| *HDDC3* | 15 | 1.89 | 2.97E-02 | 6.11E-01 | Non-documented | Non-documented |
| *ANKRD1* | 10 | 1.88 | 2.97E-02 | 6.11E-01 | Non-documented | Non-documented |
| *ESPNL* | 2 | 1.88 | 2.97E-02 | 6.11E-01 | Non-documented | Non-documented |
| *TPTE2* | 13 | 1.88 | 2.98E-02 | 6.12E-01 | Non-documented | Non-documented |
| *CLK1* | 2 | 1.88 | 2.99E-02 | 6.13E-01 | Non-documented | Documented |
| *CAMK1D* | 10 | 1.88 | 2.99E-02 | 6.13E-01 | Non-documented | Non-documented |
| *DDX58* | 9 | 1.88 | 3.00E-02 | 6.14E-01 | Non-documented | Non-documented |
| *PRELID1* | 5 | 1.88 | 3.00E-02 | 6.14E-01 | Non-documented | Non-documented |
| *FAM196A* | 10 | 1.88 | 3.01E-02 | 6.14E-01 | Non-documented | Non-documented |
| *INSR* | 19 | 1.88 | 3.02E-02 | 6.16E-01 | Non-documented | Non-documented |
| *TALDO1* | 11 | 1.88 | 3.03E-02 | 6.16E-01 | Non-documented | Non-documented |
| *MPZL2* | 11 | 1.88 | 3.03E-02 | 6.16E-01 | Non-documented | Documented |
| *MGRN1* | 16 | 1.88 | 3.03E-02 | 6.16E-01 | Non-documented | Non-documented |
| *KCNIP3* | 2 | 1.88 | 3.03E-02 | 6.16E-01 | Non-documented | Non-documented |
| *NOX5* | 15 | 1.87 | 3.04E-02 | 6.17E-01 | Non-documented | Non-documented |
| *NPTXR* | 22 | 1.87 | 3.05E-02 | 6.18E-01 | Non-documented | Non-documented |
| *RBL1* | 20 | 1.87 | 3.07E-02 | 6.22E-01 | Non-documented | Non-documented |
| *SF3B5* | 6 | 1.87 | 3.08E-02 | 6.22E-01 | Non-documented | Non-documented |
| *OR4M2* | 15 | 1.87 | 3.08E-02 | 6.22E-01 | Non-documented | Non-documented |
| *OR4N4* | 15 | 1.87 | 3.08E-02 | 6.22E-01 | Non-documented | Non-documented |
| *TFEC* | 7 | 1.87 | 3.09E-02 | 6.22E-01 | Non-documented | Non-documented |
| *MCM4* | 8 | 1.86 | 3.12E-02 | 6.27E-01 | Non-documented | Non-documented |
| *LCNL1* | 9 | 1.86 | 3.13E-02 | 6.28E-01 | Non-documented | Non-documented |
| *GRAMD4* | 22 | 1.86 | 3.13E-02 | 6.28E-01 | Non-documented | Non-documented |
| *SCT* | 11 | 1.86 | 3.13E-02 | 6.28E-01 | Non-documented | Non-documented |
| *MPDZ* | 9 | 1.86 | 3.13E-02 | 6.28E-01 | Non-documented | Non-documented |
| *ANKRD9* | 14 | 1.86 | 3.14E-02 | 6.28E-01 | Non-documented | Non-documented |
| *HIST1H4I* | 6 | 1.86 | 3.15E-02 | 6.29E-01 | Non-documented | Non-documented |
| *ZBED4* | 22 | 1.86 | 3.15E-02 | 6.29E-01 | Non-documented | Non-documented |
| *BVES* | 6 | 1.86 | 3.15E-02 | 6.29E-01 | Non-documented | Non-documented |
| *WRB* | 21 | 1.86 | 3.16E-02 | 6.29E-01 | Non-documented | Non-documented |
| *SENP6* | 6 | 1.86 | 3.16E-02 | 6.30E-01 | Non-documented | Non-documented |
| *PLK2* | 5 | 1.86 | 3.17E-02 | 6.30E-01 | Non-documented | Documented |
| *TLR5* | 1 | 1.86 | 3.17E-02 | 6.30E-01 | Non-documented | Non-documented |
| *MGST3* | 1 | 1.86 | 3.18E-02 | 6.30E-01 | Non-documented | Non-documented |
| *TAS2R8* | 12 | 1.86 | 3.18E-02 | 6.30E-01 | Non-documented | Non-documented |
| *GEMIN4* | 17 | 1.85 | 3.18E-02 | 6.30E-01 | Non-documented | Documented |
| *MRPL54* | 19 | 1.85 | 3.19E-02 | 6.30E-01 | Non-documented | Non-documented |
| *FAM76B* | 11 | 1.85 | 3.19E-02 | 6.30E-01 | Non-documented | Non-documented |
| *CYP4F8* | 19 | 1.85 | 3.20E-02 | 6.30E-01 | Non-documented | Non-documented |
| *GPBP1* | 5 | 1.85 | 3.20E-02 | 6.30E-01 | Non-documented | Non-documented |
| *CES4A* | 16 | 1.85 | 3.21E-02 | 6.30E-01 | Non-documented | Non-documented |
| *TMPRSS11D* | 4 | 1.85 | 3.21E-02 | 6.30E-01 | Non-documented | Non-documented |
| *MZF1* | 19 | 1.85 | 3.22E-02 | 6.30E-01 | Non-documented | Non-documented |
| *DAW1* | 2 | 1.85 | 3.22E-02 | 6.30E-01 | Non-documented | Non-documented |
| *SIGLEC11* | 19 | 1.85 | 3.22E-02 | 6.30E-01 | Non-documented | Documented |
| *CTXN3* | 5 | 1.85 | 3.22E-02 | 6.30E-01 | Non-documented | Non-documented |
| *TMEM100* | 17 | 1.85 | 3.23E-02 | 6.30E-01 | Non-documented | Non-documented |
| *RASGEF1A* | 10 | 1.85 | 3.23E-02 | 6.30E-01 | Non-documented | Non-documented |
| *ADGRE3* | 19 | 1.85 | 3.23E-02 | 6.30E-01 | Non-documented | Non-documented |
| *FBXO4* | 5 | 1.85 | 3.24E-02 | 6.31E-01 | Non-documented | Non-documented |
| *SELENBP1* | 1 | 1.85 | 3.24E-02 | 6.31E-01 | Non-documented | Non-documented |
| *ZNF581* | 19 | 1.85 | 3.24E-02 | 6.31E-01 | Non-documented | Non-documented |
| *PRKCG* | 19 | 1.85 | 3.25E-02 | 6.31E-01 | Non-documented | Non-documented |
| *CHEK1* | 11 | 1.85 | 3.25E-02 | 6.31E-01 | Non-documented | Documented |
| *SPESP1* | 15 | 1.84 | 3.25E-02 | 6.31E-01 | Non-documented | Non-documented |
| *TNNT2* | 1 | 1.84 | 3.26E-02 | 6.32E-01 | Non-documented | Non-documented |
| *PPP1R13B* | 14 | 1.84 | 3.26E-02 | 6.32E-01 | Non-documented | Non-documented |
| *GABRG3* | 15 | 1.84 | 3.26E-02 | 6.32E-01 | Non-documented | Documented |
| *EAPP* | 14 | 1.84 | 3.27E-02 | 6.32E-01 | Non-documented | Non-documented |
| *HSD17B4* | 5 | 1.84 | 3.29E-02 | 6.33E-01 | Non-documented | Non-documented |
| *CXCL6* | 4 | 1.84 | 3.29E-02 | 6.33E-01 | Non-documented | Non-documented |
| *GNAI3* | 1 | 1.84 | 3.29E-02 | 6.33E-01 | Non-documented | Non-documented |
| *YTHDC1* | 4 | 1.84 | 3.30E-02 | 6.33E-01 | Non-documented | Non-documented |
| *OR1L1* | 9 | 1.84 | 3.30E-02 | 6.33E-01 | Non-documented | Non-documented |
| *BEGAIN* | 14 | 1.84 | 3.30E-02 | 6.33E-01 | Non-documented | Non-documented |
| *ELOVL7* | 5 | 1.84 | 3.30E-02 | 6.33E-01 | Non-documented | Non-documented |
| *SIX3* | 2 | 1.83 | 3.33E-02 | 6.37E-01 | Non-documented | Documented |
| *BZW1* | 2 | 1.83 | 3.33E-02 | 6.37E-01 | Non-documented | Non-documented |
| *ZNF385B* | 2 | 1.83 | 3.34E-02 | 6.37E-01 | Non-documented | Non-documented |
| *LIN7C* | 11 | 1.83 | 3.35E-02 | 6.37E-01 | Non-documented | Documented |
| *BTG2* | 1 | 1.83 | 3.35E-02 | 6.37E-01 | Non-documented | Non-documented |
| *PLIN2* | 9 | 1.83 | 3.35E-02 | 6.37E-01 | Non-documented | Non-documented |
| *ISCU* | 12 | 1.83 | 3.35E-02 | 6.37E-01 | Non-documented | Non-documented |
| *U2AF2* | 19 | 1.83 | 3.36E-02 | 6.37E-01 | Non-documented | Non-documented |
| *LOC101929097* | 19 | 1.83 | 3.36E-02 | 6.37E-01 | Non-documented | Non-documented |
| *SCN10A* | 3 | 1.83 | 3.36E-02 | 6.37E-01 | Non-documented | Non-documented |
| *MALT1* | 18 | 1.83 | 3.37E-02 | 6.37E-01 | Non-documented | Non-documented |
| *BAP1* | 3 | 1.83 | 3.38E-02 | 6.37E-01 | Non-documented | Non-documented |
| *HELQ* | 4 | 1.83 | 3.38E-02 | 6.37E-01 | Non-documented | Documented |
| *ZNF580* | 19 | 1.83 | 3.39E-02 | 6.37E-01 | Non-documented | Non-documented |
| *ARHGEF40* | 14 | 1.83 | 3.39E-02 | 6.37E-01 | Non-documented | Documented |
| *EAF2* | 3 | 1.83 | 3.39E-02 | 6.37E-01 | Non-documented | Non-documented |
| *SLC44A1* | 9 | 1.83 | 3.39E-02 | 6.37E-01 | Non-documented | Documented |
| *TTC6* | 14 | 1.83 | 3.40E-02 | 6.37E-01 | Non-documented | Non-documented |
| *CACFD1* | 9 | 1.83 | 3.40E-02 | 6.37E-01 | Non-documented | Non-documented |
| *SHFM1* | 7 | 1.83 | 3.40E-02 | 6.37E-01 | Non-documented | Non-documented |
| *OR10G3* | 14 | 1.82 | 3.41E-02 | 6.37E-01 | Non-documented | Non-documented |
| *LOC101927764* | 2 | 1.82 | 3.41E-02 | 6.37E-01 | Non-documented | Non-documented |
| *LMNTD2* | 11 | 1.82 | 3.41E-02 | 6.37E-01 | Non-documented | Non-documented |
| *THOC7* | 3 | 1.82 | 3.42E-02 | 6.37E-01 | Non-documented | Non-documented |
| *KRTAP8-1* | 21 | 1.82 | 3.42E-02 | 6.37E-01 | Non-documented | Non-documented |
| *KRT76* | 12 | 1.82 | 3.42E-02 | 6.37E-01 | Non-documented | Non-documented |
| *COPS8* | 2 | 1.82 | 3.42E-02 | 6.37E-01 | Non-documented | Documented |
| *CEP57* | 11 | 1.82 | 3.42E-02 | 6.37E-01 | Non-documented | Non-documented |
| *ARHGAP21* | 10 | 1.82 | 3.43E-02 | 6.37E-01 | Non-documented | Non-documented |
| *GNPNAT1* | 14 | 1.82 | 3.43E-02 | 6.37E-01 | Non-documented | Documented |
| *UGT8* | 4 | 1.82 | 3.43E-02 | 6.37E-01 | Non-documented | Non-documented |
| *VRK3* | 19 | 1.82 | 3.43E-02 | 6.37E-01 | Non-documented | Non-documented |
| *CCDC28B* | 1 | 1.82 | 3.44E-02 | 6.37E-01 | Non-documented | Non-documented |
| *IL36G* | 2 | 1.82 | 3.44E-02 | 6.37E-01 | Non-documented | Non-documented |
| *MTURN* | 7 | 1.82 | 3.44E-02 | 6.37E-01 | Non-documented | Non-documented |
| *ALKBH6* | 19 | 1.82 | 3.45E-02 | 6.37E-01 | Non-documented | Non-documented |
| *C7orf65* | 7 | 1.82 | 3.45E-02 | 6.37E-01 | Non-documented | Non-documented |
| *PHOX2B* | 4 | 1.82 | 3.45E-02 | 6.37E-01 | Non-documented | Non-documented |
| *ATAD3B* | 1 | 1.82 | 3.45E-02 | 6.37E-01 | Non-documented | Non-documented |
| *CDKN1C* | 11 | 1.82 | 3.46E-02 | 6.37E-01 | Non-documented | Non-documented |
| *CAPN14* | 2 | 1.82 | 3.46E-02 | 6.37E-01 | Non-documented | Non-documented |
| *TXLNA* | 1 | 1.82 | 3.47E-02 | 6.37E-01 | Non-documented | Non-documented |
| *RBBP5* | 1 | 1.82 | 3.47E-02 | 6.38E-01 | Non-documented | Non-documented |
| *PSG4* | 19 | 1.81 | 3.49E-02 | 6.39E-01 | Non-documented | Non-documented |
| *SCNN1D* | 1 | 1.81 | 3.49E-02 | 6.39E-01 | Non-documented | Non-documented |
| *ZNF311* | 6 | 1.81 | 3.49E-02 | 6.39E-01 | Non-documented | Documented |
| *PTPN23* | 3 | 1.81 | 3.50E-02 | 6.40E-01 | Non-documented | Non-documented |
| *RUSC2* | 9 | 1.81 | 3.50E-02 | 6.40E-01 | Non-documented | Non-documented |
| *CAPNS1* | 19 | 1.81 | 3.51E-02 | 6.40E-01 | Non-documented | Non-documented |
| *MYH7* | 14 | 1.81 | 3.51E-02 | 6.40E-01 | Non-documented | Non-documented |
| *VPS9D1* | 16 | 1.81 | 3.54E-02 | 6.41E-01 | Non-documented | Non-documented |
| *IFI16* | 1 | 1.81 | 3.54E-02 | 6.41E-01 | Non-documented | Non-documented |
| *CNIH1* | 14 | 1.81 | 3.54E-02 | 6.41E-01 | Non-documented | Non-documented |
| *OVCH2* | 11 | 1.81 | 3.55E-02 | 6.41E-01 | Non-documented | Non-documented |
| *ZNF790* | 19 | 1.81 | 3.55E-02 | 6.41E-01 | Non-documented | Non-documented |
| *LY75-CD302* | 2 | 1.81 | 3.55E-02 | 6.41E-01 | Non-documented | Non-documented |
| *FAAH* | 1 | 1.81 | 3.55E-02 | 6.41E-01 | Non-documented | Non-documented |
| *ARL1* | 12 | 1.81 | 3.55E-02 | 6.41E-01 | Non-documented | Documented |
| *ACOT11* | 1 | 1.81 | 3.55E-02 | 6.41E-01 | Non-documented | Documented |
| *CYP4F3* | 19 | 1.81 | 3.55E-02 | 6.41E-01 | Non-documented | Non-documented |
| *RPL3* | 22 | 1.80 | 3.56E-02 | 6.41E-01 | Non-documented | Non-documented |
| *NPTN* | 15 | 1.80 | 3.56E-02 | 6.42E-01 | Non-documented | Non-documented |
| *ZSWIM6* | 5 | 1.80 | 3.56E-02 | 6.42E-01 | Non-documented | Non-documented |
| *ZNF784* | 19 | 1.80 | 3.58E-02 | 6.43E-01 | Non-documented | Non-documented |
| *GLI2* | 2 | 1.80 | 3.59E-02 | 6.43E-01 | Non-documented | Documented |
| *RIOK3* | 18 | 1.80 | 3.59E-02 | 6.43E-01 | Non-documented | Non-documented |
| *TAP1* | 6 | 1.80 | 3.60E-02 | 6.43E-01 | Non-documented | Non-documented |
| *KXD1* | 19 | 1.80 | 3.60E-02 | 6.43E-01 | Non-documented | Non-documented |
| *NARFL* | 16 | 1.80 | 3.60E-02 | 6.43E-01 | Non-documented | Non-documented |
| *RAB24* | 5 | 1.80 | 3.60E-02 | 6.43E-01 | Non-documented | Non-documented |
| *ZNF350* | 19 | 1.80 | 3.61E-02 | 6.43E-01 | Non-documented | Non-documented |
| *NIPSNAP3A* | 9 | 1.80 | 3.61E-02 | 6.43E-01 | Non-documented | Non-documented |
| *AIPL1* | 17 | 1.80 | 3.61E-02 | 6.43E-01 | Non-documented | Non-documented |
| *LNPEP* | 5 | 1.80 | 3.62E-02 | 6.43E-01 | Non-documented | Non-documented |
| *AKR1B10* | 7 | 1.80 | 3.62E-02 | 6.43E-01 | Non-documented | Documented |
| *CYC1* | 8 | 1.79 | 3.63E-02 | 6.43E-01 | Non-documented | Non-documented |
| *LY6G6C* | 6 | 1.79 | 3.64E-02 | 6.43E-01 | Non-documented | Non-documented |
| *EPN1* | 19 | 1.79 | 3.64E-02 | 6.43E-01 | Non-documented | Non-documented |
| *GTSCR1* | 18 | 1.79 | 3.65E-02 | 6.43E-01 | Non-documented | Non-documented |
| *TEX30* | 13 | 1.79 | 3.65E-02 | 6.43E-01 | Non-documented | Non-documented |
| *MATN3* | 2 | 1.79 | 3.65E-02 | 6.43E-01 | Non-documented | Non-documented |
| *DSCAML1* | 11 | 1.79 | 3.65E-02 | 6.43E-01 | Non-documented | Non-documented |
| *OR52J3* | 11 | 1.79 | 3.66E-02 | 6.43E-01 | Non-documented | Non-documented |
| *SESN2* | 1 | 1.79 | 3.66E-02 | 6.43E-01 | Non-documented | Non-documented |
| *RBBP8* | 18 | 1.79 | 3.66E-02 | 6.43E-01 | Non-documented | Non-documented |
| *IQCC* | 1 | 1.79 | 3.66E-02 | 6.43E-01 | Non-documented | Non-documented |
| *PSMB8* | 6 | 1.79 | 3.67E-02 | 6.43E-01 | Non-documented | Non-documented |
| *DLG5* | 10 | 1.79 | 3.67E-02 | 6.43E-01 | Non-documented | Non-documented |
| *SLC22A18AS* | 11 | 1.79 | 3.67E-02 | 6.43E-01 | Non-documented | Non-documented |
| *LOC101929185* | 17 | 1.79 | 3.68E-02 | 6.43E-01 | Non-documented | Non-documented |
| *DCDC2B* | 1 | 1.79 | 3.68E-02 | 6.43E-01 | Non-documented | Non-documented |
| *PPP1R36* | 14 | 1.79 | 3.68E-02 | 6.43E-01 | Non-documented | Non-documented |
| *ICAM5* | 19 | 1.79 | 3.68E-02 | 6.44E-01 | Non-documented | Non-documented |
| *CKMT1A* | 15 | 1.79 | 3.69E-02 | 6.44E-01 | Non-documented | Non-documented |
| *TP53INP1* | 8 | 1.79 | 3.69E-02 | 6.44E-01 | Non-documented | Non-documented |
| *HSPH1* | 13 | 1.79 | 3.69E-02 | 6.44E-01 | Non-documented | Non-documented |
| *VEZF1* | 17 | 1.79 | 3.71E-02 | 6.44E-01 | Non-documented | Non-documented |
| *ANKS6* | 9 | 1.79 | 3.71E-02 | 6.44E-01 | Non-documented | Non-documented |
| *B4GALT1* | 9 | 1.79 | 3.71E-02 | 6.44E-01 | Non-documented | Non-documented |
| *LIPJ* | 10 | 1.79 | 3.71E-02 | 6.44E-01 | Non-documented | Non-documented |
| *FAM173B* | 5 | 1.78 | 3.71E-02 | 6.44E-01 | Non-documented | Non-documented |
| *SLC23A2* | 20 | 1.78 | 3.72E-02 | 6.45E-01 | Non-documented | Non-documented |
| *XRCC5* | 2 | 1.78 | 3.73E-02 | 6.45E-01 | Non-documented | Non-documented |
| *ZNF333* | 19 | 1.78 | 3.74E-02 | 6.45E-01 | Non-documented | Non-documented |
| *SWI5* | 9 | 1.78 | 3.74E-02 | 6.45E-01 | Non-documented | Non-documented |
| *MAP6D1* | 3 | 1.78 | 3.74E-02 | 6.45E-01 | Non-documented | Non-documented |
| *OR52E2* | 11 | 1.78 | 3.74E-02 | 6.45E-01 | Non-documented | Non-documented |
| *ERC2* | 3 | 1.78 | 3.76E-02 | 6.45E-01 | Non-documented | Documented |
| *PRKAB2* | 1 | 1.78 | 3.76E-02 | 6.45E-01 | Non-documented | Non-documented |
| *IQCJ-SCHIP1* | 3 | 1.78 | 3.76E-02 | 6.45E-01 | Non-documented | Non-documented |
| *EHBP1L1* | 11 | 1.78 | 3.76E-02 | 6.45E-01 | Non-documented | Non-documented |
| *MAGI3* | 1 | 1.78 | 3.76E-02 | 6.45E-01 | Non-documented | Documented |
| *ZNF555* | 19 | 1.78 | 3.76E-02 | 6.45E-01 | Non-documented | Non-documented |
| *ZNF276* | 16 | 1.78 | 3.76E-02 | 6.45E-01 | Non-documented | Documented |
| *PLEKHG7* | 12 | 1.78 | 3.77E-02 | 6.45E-01 | Non-documented | Non-documented |
| *CMTM1* | 16 | 1.78 | 3.77E-02 | 6.45E-01 | Non-documented | Non-documented |
| *UBTF* | 17 | 1.78 | 3.78E-02 | 6.45E-01 | Non-documented | Non-documented |
| *ICAM4* | 19 | 1.78 | 3.78E-02 | 6.46E-01 | Non-documented | Non-documented |
| *EIF3B* | 7 | 1.78 | 3.79E-02 | 6.46E-01 | Non-documented | Non-documented |
| *SHARPIN* | 8 | 1.78 | 3.79E-02 | 6.46E-01 | Non-documented | Non-documented |
| *RGS19* | 20 | 1.77 | 3.80E-02 | 6.47E-01 | Non-documented | Non-documented |
| *UBE2J2* | 1 | 1.77 | 3.80E-02 | 6.47E-01 | Non-documented | Non-documented |
| *PRKDC* | 8 | 1.77 | 3.81E-02 | 6.47E-01 | Non-documented | Non-documented |
| *C19orf60* | 19 | 1.77 | 3.81E-02 | 6.47E-01 | Non-documented | Non-documented |
| *PKIG* | 20 | 1.77 | 3.81E-02 | 6.47E-01 | Non-documented | Documented |
| *LST1* | 6 | 1.77 | 3.81E-02 | 6.47E-01 | Non-documented | Non-documented |
| *C20orf203* | 20 | 1.77 | 3.82E-02 | 6.48E-01 | Non-documented | Non-documented |
| *TECPR2* | 14 | 1.77 | 3.84E-02 | 6.50E-01 | Non-documented | Non-documented |
| *LPCAT3* | 12 | 1.77 | 3.85E-02 | 6.50E-01 | Non-documented | Documented |
| *POPDC3* | 6 | 1.77 | 3.85E-02 | 6.51E-01 | Non-documented | Non-documented |
| *CRLF1* | 19 | 1.77 | 3.86E-02 | 6.51E-01 | Non-documented | Non-documented |
| *RPS14* | 5 | 1.77 | 3.86E-02 | 6.51E-01 | Non-documented | Non-documented |
| *PCNP* | 3 | 1.77 | 3.86E-02 | 6.51E-01 | Non-documented | Non-documented |
| *UFC1* | 1 | 1.77 | 3.88E-02 | 6.52E-01 | Non-documented | Non-documented |
| *PIKFYVE* | 2 | 1.76 | 3.88E-02 | 6.53E-01 | Non-documented | Non-documented |
| *LRRC4B* | 19 | 1.76 | 3.89E-02 | 6.53E-01 | Non-documented | Non-documented |
| *FAM91A1* | 8 | 1.76 | 3.89E-02 | 6.53E-01 | Non-documented | Documented |
| *FAM188A* | 10 | 1.76 | 3.90E-02 | 6.53E-01 | Non-documented | Non-documented |
| *ANKLE2* | 12 | 1.76 | 3.91E-02 | 6.53E-01 | Non-documented | Non-documented |
| *SPIC* | 12 | 1.76 | 3.91E-02 | 6.53E-01 | Non-documented | Documented |
| *ARG1* | 6 | 1.76 | 3.92E-02 | 6.53E-01 | Non-documented | Non-documented |
| *IGLON5* | 19 | 1.76 | 3.92E-02 | 6.53E-01 | Non-documented | Non-documented |
| *NCR3* | 6 | 1.76 | 3.92E-02 | 6.53E-01 | Non-documented | Non-documented |
| *EIF2B3* | 1 | 1.76 | 3.93E-02 | 6.53E-01 | Non-documented | Non-documented |
| *RAB4A* | 1 | 1.76 | 3.93E-02 | 6.53E-01 | Non-documented | Non-documented |
| *ZNF628* | 19 | 1.76 | 3.93E-02 | 6.53E-01 | Non-documented | Non-documented |
| *RAB5C* | 17 | 1.76 | 3.93E-02 | 6.53E-01 | Non-documented | Non-documented |
| *NEDD9* | 6 | 1.76 | 3.93E-02 | 6.53E-01 | Non-documented | Documented |
| *VWA3B* | 2 | 1.76 | 3.94E-02 | 6.54E-01 | Non-documented | Non-documented |
| *GPR135* | 14 | 1.76 | 3.95E-02 | 6.54E-01 | Non-documented | Non-documented |
| *TOMM5* | 9 | 1.76 | 3.96E-02 | 6.55E-01 | Non-documented | Non-documented |
| *RHOF* | 12 | 1.76 | 3.96E-02 | 6.55E-01 | Non-documented | Non-documented |
| *SLC25A2* | 5 | 1.76 | 3.96E-02 | 6.55E-01 | Non-documented | Non-documented |
| *FDX1L* | 19 | 1.76 | 3.96E-02 | 6.55E-01 | Non-documented | Non-documented |
| *SCN9A* | 2 | 1.75 | 3.98E-02 | 6.57E-01 | Non-documented | Non-documented |
| *C1orf87* | 1 | 1.75 | 3.99E-02 | 6.57E-01 | Non-documented | Non-documented |
| *MEF2C* | 5 | 1.75 | 3.99E-02 | 6.57E-01 | Non-documented | Non-documented |
| *CASP5* | 11 | 1.75 | 3.99E-02 | 6.57E-01 | Non-documented | Non-documented |
| *INF2* | 14 | 1.75 | 3.99E-02 | 6.57E-01 | Non-documented | Non-documented |
| *PARD6A* | 16 | 1.75 | 4.01E-02 | 6.59E-01 | Non-documented | Non-documented |
| *METTL7A* | 12 | 1.75 | 4.01E-02 | 6.59E-01 | Non-documented | Non-documented |
| *C16orf93* | 16 | 1.75 | 4.02E-02 | 6.60E-01 | Non-documented | Non-documented |
| *ACD* | 16 | 1.75 | 4.03E-02 | 6.60E-01 | Non-documented | Non-documented |
| *ADAM30* | 1 | 1.75 | 4.04E-02 | 6.60E-01 | Non-documented | Non-documented |
| *DEFA6* | 8 | 1.75 | 4.04E-02 | 6.60E-01 | Non-documented | Non-documented |
| *SAMD4A* | 14 | 1.75 | 4.05E-02 | 6.60E-01 | Non-documented | Non-documented |
| *PPP2R2C* | 4 | 1.74 | 4.05E-02 | 6.60E-01 | Non-documented | Documented |
| *PNPLA5* | 22 | 1.74 | 4.05E-02 | 6.60E-01 | Non-documented | Non-documented |
| *PRKD1* | 14 | 1.74 | 4.06E-02 | 6.60E-01 | Non-documented | Non-documented |
| *ITGA8* | 10 | 1.74 | 4.07E-02 | 6.60E-01 | Non-documented | Non-documented |
| *DRD4* | 11 | 1.74 | 4.07E-02 | 6.60E-01 | Non-documented | Non-documented |
| *CABIN1* | 22 | 1.74 | 4.07E-02 | 6.60E-01 | Non-documented | Non-documented |
| *SEL1L2* | 20 | 1.74 | 4.07E-02 | 6.60E-01 | Non-documented | Non-documented |
| *CLHC1* | 2 | 1.74 | 4.07E-02 | 6.60E-01 | Non-documented | Non-documented |
| *GALNS* | 16 | 1.74 | 4.08E-02 | 6.60E-01 | Non-documented | Non-documented |
| *SLC24A2* | 9 | 1.74 | 4.08E-02 | 6.60E-01 | Non-documented | Non-documented |
| *ABCA13* | 7 | 1.74 | 4.10E-02 | 6.60E-01 | Non-documented | Non-documented |
| *NDRG1* | 8 | 1.74 | 4.10E-02 | 6.60E-01 | Non-documented | Non-documented |
| *BPIFC* | 22 | 1.74 | 4.10E-02 | 6.60E-01 | Non-documented | Non-documented |
| *MORC1* | 3 | 1.74 | 4.11E-02 | 6.60E-01 | Non-documented | Documented |
| *DEFA4* | 8 | 1.74 | 4.11E-02 | 6.60E-01 | Non-documented | Non-documented |
| *UMPS* | 3 | 1.74 | 4.11E-02 | 6.60E-01 | Non-documented | Non-documented |
| *ARHGAP44* | 17 | 1.74 | 4.11E-02 | 6.60E-01 | Non-documented | Non-documented |
| *APLP1* | 19 | 1.74 | 4.11E-02 | 6.60E-01 | Non-documented | Non-documented |
| *SLC22A18* | 11 | 1.74 | 4.12E-02 | 6.60E-01 | Non-documented | Non-documented |
| *SLC9A1* | 1 | 1.74 | 4.13E-02 | 6.60E-01 | Non-documented | Non-documented |
| *C20orf141* | 20 | 1.74 | 4.13E-02 | 6.60E-01 | Non-documented | Non-documented |
| *SH3PXD2A* | 10 | 1.74 | 4.13E-02 | 6.60E-01 | Non-documented | Documented |
| *PKDCC* | 2 | 1.73 | 4.14E-02 | 6.60E-01 | Non-documented | Non-documented |
| *PAH* | 12 | 1.73 | 4.14E-02 | 6.60E-01 | Non-documented | Non-documented |
| *WWP2* | 16 | 1.73 | 4.15E-02 | 6.60E-01 | Non-documented | Non-documented |
| *LY6G6D* | 6 | 1.73 | 4.15E-02 | 6.60E-01 | Non-documented | Non-documented |
| *C9orf170* | 9 | 1.73 | 4.15E-02 | 6.60E-01 | Non-documented | Non-documented |
| *TMEM234* | 1 | 1.73 | 4.15E-02 | 6.60E-01 | Non-documented | Non-documented |
| *CLDN3* | 7 | 1.73 | 4.16E-02 | 6.60E-01 | Non-documented | Non-documented |
| *RYR1* | 19 | 1.73 | 4.16E-02 | 6.60E-01 | Non-documented | Non-documented |
| *MAF1* | 8 | 1.73 | 4.16E-02 | 6.60E-01 | Non-documented | Non-documented |
| *TRIM9* | 14 | 1.73 | 4.17E-02 | 6.60E-01 | Non-documented | Non-documented |
| *TMEM241* | 18 | 1.73 | 4.17E-02 | 6.60E-01 | Non-documented | Non-documented |
| *SYNPO2* | 4 | 1.73 | 4.17E-02 | 6.60E-01 | Non-documented | Documented |
| *SIGLEC8* | 19 | 1.73 | 4.17E-02 | 6.60E-01 | Non-documented | Non-documented |
| *GATA3* | 10 | 1.73 | 4.17E-02 | 6.60E-01 | Non-documented | Documented |
| *FRA10AC1* | 10 | 1.73 | 4.17E-02 | 6.60E-01 | Non-documented | Non-documented |
| *FOXB2* | 9 | 1.73 | 4.17E-02 | 6.60E-01 | Non-documented | Non-documented |
| *KIRREL2* | 19 | 1.73 | 4.18E-02 | 6.60E-01 | Non-documented | Non-documented |
| *KIF6* | 6 | 1.73 | 4.18E-02 | 6.60E-01 | Non-documented | Non-documented |
| *APRT* | 16 | 1.73 | 4.19E-02 | 6.61E-01 | Non-documented | Non-documented |
| *CRISPLD1* | 8 | 1.73 | 4.20E-02 | 6.61E-01 | Non-documented | Non-documented |
| *SNX15* | 11 | 1.73 | 4.21E-02 | 6.61E-01 | Non-documented | Non-documented |
| *TTC32* | 2 | 1.73 | 4.21E-02 | 6.61E-01 | Non-documented | Non-documented |
| *CYB561A3* | 11 | 1.73 | 4.22E-02 | 6.61E-01 | Non-documented | Non-documented |
| *PTPRN2* | 7 | 1.73 | 4.22E-02 | 6.61E-01 | Non-documented | Documented |
| *NTSR1* | 20 | 1.73 | 4.22E-02 | 6.61E-01 | Non-documented | Non-documented |
| *LYSMD4* | 15 | 1.73 | 4.22E-02 | 6.61E-01 | Non-documented | Non-documented |
| *SLC26A8* | 6 | 1.73 | 4.22E-02 | 6.61E-01 | Non-documented | Non-documented |
| *ZNF689* | 16 | 1.73 | 4.22E-02 | 6.61E-01 | Non-documented | Non-documented |
| *ASAH2B* | 10 | 1.73 | 4.23E-02 | 6.61E-01 | Non-documented | Non-documented |
| *RRAS2* | 11 | 1.72 | 4.23E-02 | 6.61E-01 | Non-documented | Non-documented |
| *TMEM210* | 9 | 1.72 | 4.25E-02 | 6.61E-01 | Non-documented | Non-documented |
| *COPS2* | 15 | 1.72 | 4.25E-02 | 6.61E-01 | Non-documented | Documented |
| *RASSF2* | 20 | 1.72 | 4.25E-02 | 6.61E-01 | Non-documented | Non-documented |
| *LRRC26* | 9 | 1.72 | 4.25E-02 | 6.61E-01 | Non-documented | Non-documented |
| *CBLN1* | 16 | 1.72 | 4.26E-02 | 6.61E-01 | Non-documented | Non-documented |
| *UBE2R2* | 9 | 1.72 | 4.26E-02 | 6.61E-01 | Non-documented | Non-documented |
| *SLC2A6* | 9 | 1.72 | 4.26E-02 | 6.61E-01 | Non-documented | Non-documented |
| *NOS3* | 7 | 1.72 | 4.26E-02 | 6.61E-01 | Non-documented | Non-documented |
| *SSU72* | 1 | 1.72 | 4.27E-02 | 6.61E-01 | Non-documented | Non-documented |
| *PPP3R2* | 9 | 1.72 | 4.27E-02 | 6.61E-01 | Non-documented | Non-documented |
| *PSMC6* | 14 | 1.72 | 4.27E-02 | 6.61E-01 | Non-documented | Documented |
| *FRMD6* | 14 | 1.72 | 4.28E-02 | 6.61E-01 | Non-documented | Documented |
| *G6PC3* | 17 | 1.72 | 4.28E-02 | 6.61E-01 | Non-documented | Non-documented |
| *ELP6* | 3 | 1.72 | 4.28E-02 | 6.61E-01 | Non-documented | Non-documented |
| *FOXF1* | 16 | 1.72 | 4.29E-02 | 6.61E-01 | Non-documented | Documented |
| *ZNF614* | 19 | 1.72 | 4.29E-02 | 6.61E-01 | Non-documented | Non-documented |
| *SLC15A2* | 3 | 1.72 | 4.31E-02 | 6.63E-01 | Non-documented | Non-documented |
| *LAMC2* | 1 | 1.72 | 4.31E-02 | 6.63E-01 | Non-documented | Non-documented |
| *GPAA1* | 8 | 1.72 | 4.32E-02 | 6.64E-01 | Non-documented | Non-documented |
| *ZNF32* | 10 | 1.71 | 4.32E-02 | 6.64E-01 | Non-documented | Non-documented |
| *APCDD1* | 18 | 1.71 | 4.33E-02 | 6.64E-01 | Non-documented | Documented |
| *EIF3I* | 1 | 1.71 | 4.33E-02 | 6.64E-01 | Non-documented | Non-documented |
| *RABGAP1* | 9 | 1.71 | 4.33E-02 | 6.64E-01 | Non-documented | Non-documented |
| *ADAM21* | 14 | 1.71 | 4.34E-02 | 6.64E-01 | Non-documented | Documented |
| *KIAA0753* | 17 | 1.71 | 4.36E-02 | 6.66E-01 | Non-documented | Documented |
| *LARS2* | 3 | 1.71 | 4.36E-02 | 6.66E-01 | Non-documented | Non-documented |
| *KDELC1* | 13 | 1.71 | 4.37E-02 | 6.68E-01 | Non-documented | Non-documented |
| *PDDC1* | 11 | 1.71 | 4.38E-02 | 6.68E-01 | Non-documented | Non-documented |
| *TAF3* | 10 | 1.71 | 4.38E-02 | 6.68E-01 | Non-documented | Non-documented |
| *DPEP1* | 16 | 1.71 | 4.39E-02 | 6.68E-01 | Non-documented | Non-documented |
| *RHBDL3* | 17 | 1.71 | 4.39E-02 | 6.68E-01 | Non-documented | Non-documented |
| *PTGIR* | 19 | 1.71 | 4.40E-02 | 6.69E-01 | Non-documented | Non-documented |
| *SLC12A3* | 16 | 1.71 | 4.40E-02 | 6.69E-01 | Non-documented | Non-documented |
| *TAL1* | 1 | 1.71 | 4.41E-02 | 6.69E-01 | Non-documented | Non-documented |
| *N6AMT2* | 13 | 1.70 | 4.43E-02 | 6.72E-01 | Non-documented | Documented |
| *UNC45A* | 15 | 1.70 | 4.43E-02 | 6.72E-01 | Non-documented | Non-documented |
| *RBM20* | 10 | 1.70 | 4.44E-02 | 6.72E-01 | Non-documented | Non-documented |
| *ZBTB20* | 3 | 1.70 | 4.45E-02 | 6.72E-01 | Non-documented | Documented |
| *CHKA* | 11 | 1.70 | 4.45E-02 | 6.72E-01 | Non-documented | Non-documented |
| *SPATS1* | 6 | 1.70 | 4.45E-02 | 6.72E-01 | Non-documented | Non-documented |
| *SLC4A3* | 2 | 1.70 | 4.45E-02 | 6.72E-01 | Non-documented | Non-documented |
| *CD72* | 9 | 1.70 | 4.46E-02 | 6.72E-01 | Non-documented | Non-documented |
| *RBMXL2* | 11 | 1.70 | 4.46E-02 | 6.72E-01 | Non-documented | Non-documented |
| *UCHL1* | 4 | 1.70 | 4.47E-02 | 6.72E-01 | Non-documented | Non-documented |
| *DCUN1D5* | 11 | 1.70 | 4.47E-02 | 6.72E-01 | Non-documented | Non-documented |
| *HGH1* | 8 | 1.70 | 4.47E-02 | 6.72E-01 | Non-documented | Non-documented |
| *IGDCC4* | 15 | 1.70 | 4.48E-02 | 6.72E-01 | Non-documented | Non-documented |
| *DUSP27* | 1 | 1.70 | 4.48E-02 | 6.72E-01 | Non-documented | Non-documented |
| *ACRV1* | 11 | 1.70 | 4.48E-02 | 6.72E-01 | Non-documented | Documented |
| *GRAMD3* | 5 | 1.70 | 4.48E-02 | 6.72E-01 | Non-documented | Non-documented |
| *CRYZ* | 1 | 1.69 | 4.51E-02 | 6.74E-01 | Non-documented | Non-documented |
| *IDH1* | 2 | 1.69 | 4.51E-02 | 6.74E-01 | Non-documented | Non-documented |
| *ENKD1* | 16 | 1.69 | 4.51E-02 | 6.74E-01 | Non-documented | Non-documented |
| *ZSCAN5B* | 19 | 1.69 | 4.51E-02 | 6.74E-01 | Non-documented | Non-documented |
| *KLHL42* | 12 | 1.69 | 4.52E-02 | 6.75E-01 | Non-documented | Non-documented |
| *CDK15* | 2 | 1.69 | 4.53E-02 | 6.75E-01 | Non-documented | Non-documented |
| *OR2B3* | 6 | 1.69 | 4.54E-02 | 6.76E-01 | Non-documented | Non-documented |
| *ARL2* | 11 | 1.69 | 4.54E-02 | 6.76E-01 | Non-documented | Non-documented |
| *C1orf216* | 1 | 1.69 | 4.55E-02 | 6.77E-01 | Non-documented | Non-documented |
| *MRPL46* | 15 | 1.69 | 4.56E-02 | 6.77E-01 | Non-documented | Non-documented |
| *C1QL1* | 17 | 1.69 | 4.57E-02 | 6.77E-01 | Non-documented | Non-documented |
| *EIF3K* | 19 | 1.69 | 4.57E-02 | 6.77E-01 | Non-documented | Non-documented |
| *FMO4* | 1 | 1.69 | 4.57E-02 | 6.77E-01 | Non-documented | Non-documented |
| *ELAC2* | 17 | 1.69 | 4.58E-02 | 6.77E-01 | Non-documented | Documented |
| *ZBED3* | 5 | 1.69 | 4.58E-02 | 6.77E-01 | Non-documented | Non-documented |
| *PON3* | 7 | 1.69 | 4.59E-02 | 6.77E-01 | Non-documented | Non-documented |
| *MYRF* | 11 | 1.69 | 4.59E-02 | 6.77E-01 | Non-documented | Documented |
| *C20orf78* | 20 | 1.69 | 4.59E-02 | 6.77E-01 | Non-documented | Non-documented |
| *ASNS* | 7 | 1.69 | 4.59E-02 | 6.77E-01 | Non-documented | Non-documented |
| *GLYAT* | 11 | 1.69 | 4.59E-02 | 6.77E-01 | Non-documented | Non-documented |
| *ZBTB26* | 9 | 1.69 | 4.59E-02 | 6.77E-01 | Non-documented | Non-documented |
| *C15orf53* | 15 | 1.69 | 4.60E-02 | 6.77E-01 | Non-documented | Non-documented |
| *C19orf40* | 19 | 1.68 | 4.60E-02 | 6.77E-01 | Non-documented | Non-documented |
| *TRIM67* | 1 | 1.68 | 4.60E-02 | 6.77E-01 | Non-documented | Non-documented |
| *SPATA5* | 4 | 1.68 | 4.61E-02 | 6.77E-01 | Non-documented | Non-documented |
| *TIAM1* | 21 | 1.68 | 4.61E-02 | 6.77E-01 | Non-documented | Non-documented |
| *MTR* | 1 | 1.68 | 4.62E-02 | 6.77E-01 | Non-documented | Non-documented |
| *MUC16* | 19 | 1.68 | 4.62E-02 | 6.77E-01 | Non-documented | Documented |
| *CCDC67* | 11 | 1.68 | 4.63E-02 | 6.78E-01 | Non-documented | Non-documented |
| *CEMIP* | 15 | 1.68 | 4.66E-02 | 6.80E-01 | Non-documented | Non-documented |
| *ECHDC3* | 10 | 1.68 | 4.66E-02 | 6.80E-01 | Non-documented | Documented |
| *MDM4* | 1 | 1.68 | 4.66E-02 | 6.80E-01 | Non-documented | Documented |
| *DDR2* | 1 | 1.68 | 4.66E-02 | 6.80E-01 | Non-documented | Non-documented |
| *RMDN1* | 8 | 1.68 | 4.66E-02 | 6.80E-01 | Non-documented | Non-documented |
| *METTL22* | 16 | 1.68 | 4.68E-02 | 6.82E-01 | Non-documented | Non-documented |
| *RP1* | 8 | 1.68 | 4.68E-02 | 6.82E-01 | Non-documented | Documented |
| *HAPLN3* | 15 | 1.68 | 4.69E-02 | 6.82E-01 | Non-documented | Documented |
| *CLDND2* | 19 | 1.68 | 4.69E-02 | 6.82E-01 | Non-documented | Non-documented |
| *SLC26A11* | 17 | 1.68 | 4.70E-02 | 6.82E-01 | Non-documented | Non-documented |
| *RNF186* | 1 | 1.67 | 4.70E-02 | 6.82E-01 | Non-documented | Non-documented |
| *TMEM138* | 11 | 1.67 | 4.71E-02 | 6.82E-01 | Non-documented | Non-documented |
| *NAALADL1* | 11 | 1.67 | 4.71E-02 | 6.82E-01 | Non-documented | Non-documented |
| *NAE1* | 16 | 1.67 | 4.71E-02 | 6.82E-01 | Non-documented | Non-documented |
| *NR2F1* | 5 | 1.67 | 4.72E-02 | 6.82E-01 | Non-documented | Non-documented |
| *KIR2DL1* | 19 | 1.67 | 4.72E-02 | 6.82E-01 | Non-documented | Non-documented |
| *TSEN54* | 17 | 1.67 | 4.72E-02 | 6.82E-01 | Non-documented | Non-documented |
| *PPP1R17* | 7 | 1.67 | 4.73E-02 | 6.82E-01 | Non-documented | Non-documented |
| *TMEM239* | 20 | 1.67 | 4.73E-02 | 6.82E-01 | Non-documented | Non-documented |
| *CDC37L1* | 9 | 1.67 | 4.74E-02 | 6.83E-01 | Non-documented | Non-documented |
| *ANO7P1* | 1 | 1.67 | 4.75E-02 | 6.83E-01 | Non-documented | Non-documented |
| *SPHAR* | 1 | 1.67 | 4.75E-02 | 6.84E-01 | Non-documented | Non-documented |
| *FBXO36* | 2 | 1.67 | 4.76E-02 | 6.84E-01 | Non-documented | Documented |
| *PDCD1* | 2 | 1.67 | 4.77E-02 | 6.85E-01 | Non-documented | Non-documented |
| *ALG12* | 22 | 1.67 | 4.78E-02 | 6.86E-01 | Non-documented | Non-documented |
| *XPO4* | 13 | 1.67 | 4.79E-02 | 6.86E-01 | Non-documented | Documented |
| *GNG8* | 19 | 1.67 | 4.79E-02 | 6.86E-01 | Non-documented | Non-documented |
| *TP63* | 3 | 1.66 | 4.80E-02 | 6.86E-01 | Non-documented | Documented |
| *SLC1A1* | 9 | 1.66 | 4.80E-02 | 6.86E-01 | Non-documented | Documented |
| *CCKBR* | 11 | 1.66 | 4.80E-02 | 6.86E-01 | Non-documented | Non-documented |
| *TESK1* | 9 | 1.66 | 4.81E-02 | 6.86E-01 | Non-documented | Non-documented |
| *NIPSNAP1* | 22 | 1.66 | 4.81E-02 | 6.86E-01 | Non-documented | Non-documented |
| *QPCT* | 2 | 1.66 | 4.81E-02 | 6.86E-01 | Non-documented | Non-documented |
| *KRTAP19-8* | 21 | 1.66 | 4.82E-02 | 6.86E-01 | Non-documented | Non-documented |
| *AS3MT* | 10 | 1.66 | 4.82E-02 | 6.86E-01 | Non-documented | Documented |
| *RGMA* | 15 | 1.66 | 4.83E-02 | 6.86E-01 | Non-documented | Non-documented |
| *ISM2* | 14 | 1.66 | 4.83E-02 | 6.86E-01 | Non-documented | Non-documented |
| *KCNV1* | 8 | 1.66 | 4.83E-02 | 6.86E-01 | Non-documented | Non-documented |
| *AGGF1* | 5 | 1.66 | 4.83E-02 | 6.86E-01 | Non-documented | Non-documented |
| *PPP1R14A* | 19 | 1.66 | 4.84E-02 | 6.87E-01 | Non-documented | Non-documented |
| *EXOSC4* | 8 | 1.66 | 4.84E-02 | 6.87E-01 | Non-documented | Non-documented |
| *MRPL3* | 3 | 1.66 | 4.86E-02 | 6.88E-01 | Non-documented | Non-documented |
| *ZNF224* | 19 | 1.66 | 4.87E-02 | 6.90E-01 | Non-documented | Non-documented |
| *GALK2* | 15 | 1.66 | 4.88E-02 | 6.90E-01 | Non-documented | Documented |
| *UBALD1* | 16 | 1.66 | 4.89E-02 | 6.92E-01 | Non-documented | Non-documented |
| *TMBIM1* | 2 | 1.65 | 4.90E-02 | 6.92E-01 | Non-documented | Documented |
| *CALM3* | 19 | 1.65 | 4.91E-02 | 6.92E-01 | Non-documented | Non-documented |
| *OR52N4* | 11 | 1.65 | 4.91E-02 | 6.92E-01 | Non-documented | Non-documented |
| *TDRP* | 8 | 1.65 | 4.91E-02 | 6.92E-01 | Non-documented | Non-documented |
| *CEBPD* | 8 | 1.65 | 4.91E-02 | 6.92E-01 | Non-documented | Non-documented |
| *RFPL3* | 22 | 1.65 | 4.92E-02 | 6.92E-01 | Non-documented | Non-documented |
| *SUZ12* | 17 | 1.65 | 4.92E-02 | 6.92E-01 | Non-documented | Non-documented |
| *TMEM186* | 16 | 1.65 | 4.93E-02 | 6.92E-01 | Non-documented | Non-documented |
| *CELSR1* | 22 | 1.65 | 4.94E-02 | 6.92E-01 | Non-documented | Non-documented |
| *FRMPD1* | 9 | 1.65 | 4.95E-02 | 6.92E-01 | Non-documented | Non-documented |
| *42990* | 16 | 1.65 | 4.95E-02 | 6.92E-01 | Non-documented | Non-documented |
| *NOS1* | 12 | 1.65 | 4.96E-02 | 6.92E-01 | Non-documented | Documented |
| *ADH7* | 4 | 1.65 | 4.96E-02 | 6.92E-01 | Reported | Non-documented |
| *CRELD2* | 22 | 1.65 | 4.97E-02 | 6.92E-01 | Non-documented | Non-documented |
| *GLYR1* | 16 | 1.65 | 4.97E-02 | 6.92E-01 | Non-documented | Documented |
| *ZBTB6* | 9 | 1.65 | 4.98E-02 | 6.92E-01 | Non-documented | Non-documented |
| *BATF2* | 11 | 1.65 | 4.98E-02 | 6.92E-01 | Non-documented | Non-documented |
| *TKFC* | 11 | 1.65 | 4.99E-02 | 6.92E-01 | Non-documented | Non-documented |
| *UBN1* | 16 | 1.65 | 4.99E-02 | 6.92E-01 | Non-documented | Non-documented |
| *LOC100287387* | 2 | 1.65 | 4.99E-02 | 6.92E-01 | Non-documented | Non-documented |
| *MYLK2* | 20 | 1.65 | 4.99E-02 | 6.92E-01 | Non-documented | Non-documented |
| *CXCL1* | 4 | 1.65 | 4.99E-02 | 6.92E-01 | Non-documented | Non-documented |
| *USP21* | 1 | 1.65 | 5.00E-02 | 6.92E-01 | Non-documented | Non-documented |
| *ZNF2* | 2 | 1.64 | 5.00E-02 | 6.92E-01 | Non-documented | Non-documented |

**Supplemental Table S2. Significant pathways (FDR< 0.1) enriched by 1,324 genes identified from MAGMA**

| **Gene Set** | **Description** | **Size** | **Ratio** | **P Value** | **FDR** |
| --- | --- | --- | --- | --- | --- |
| hsa05150 | Staphylococcus aureus infection | 56 | 3.89 | 8.04E-06 | 2.62E-03 |
| hsa04612 | Antigen processing and presentation | 77 | 3.03 | 9.18E-05 | 1.50E-02 |
| hsa05332 | Graft-versus-host disease | 41 | 3.80 | 2.01E-04 | 2.18E-02 |
| hsa04940 | Type I diabetes mellitus | 43 | 3.62 | 3.05E-04 | 2.49E-02 |
| hsa05330 | Allograft rejection | 38 | 3.69 | 5.28E-04 | 3.44E-02 |
| hsa04514 | Cell adhesion molecules (CAMs) | 144 | 2.16 | 8.40E-04 | 4.56E-02 |
| hsa04371 | Apelin signaling pathway | 137 | 2.16 | 1.14E-03 | 5.33E-02 |
| hsa05320 | Autoimmune thyroid disease | 53 | 2.94 | 1.74E-03 | 7.03E-02 |
| hsa04934 | Cushing syndrome | 154 | 2.02 | 1.94E-03 | 7.03E-02 |
| hsa05310 | Asthma | 31 | 3.51 | 2.95E-03 | 9.22E-02 |
| hsa05166 | Human T-cell leukemia virus 1 infection | 255 | 1.71 | 3.59E-03 | 9.22E-02 |
| hsa04727 | GABAergic synapse | 88 | 2.30 | 3.84E-03 | 9.22E-02 |
| hsa04145 | Phagosome | 152 | 1.95 | 3.85E-03 | 9.22E-02 |
| hsa05416 | Viral myocarditis | 59 | 2.64 | 3.97E-03 | 9.22E-02 |
| hsa04211 | Longevity regulating pathway | 89 | 2.27 | 4.24E-03 | 9.22E-02 |
| hsa00790 | Folate biosynthesis | 26 | 3.59 | 5.21E-03 | 9.32E-02 |
| hsa03040 | Spliceosome | 134 | 1.97 | 5.24E-03 | 9.32E-02 |
| hsa05145 | Toxoplasmosis | 113 | 2.07 | 5.58E-03 | 9.32E-02 |
| hsa04658 | Th1 and Th2 cell differentiation | 92 | 2.20 | 5.65E-03 | 9.32E-02 |
| hsa04213 | Longevity regulating pathway | 62 | 2.51 | 5.72E-03 | 9.32E-02 |

**Supplemental Table S3. Significant GO-terms of cellular components enriched by 1,324 genes**

| **Gene Set** | **Description** | **Size** | **Ratio** | **P Value** |
| --- | --- | --- | --- | --- |
| GO:0042611 | MHC protein complex | 21 | 4.19 | 0.0022 |
| GO:0097060 | synaptic membrane | 430 | 1.43 | 0.0112 |
| GO:1905348 | endonuclease complex | 29 | 3.04 | 0.012 |
| GO:0098552 | side of membrane | 481 | 1.37 | 0.018 |

**Supplemental Table S4. Significant GO-terms of cellular components enriched by 1,324 genes**

| **Gene Set** | **Description** | **Size** | **Ratio** | **P Value** |
| --- | --- | --- | --- | --- |
| GO:0004497 | monooxygenase activity | 95 | 2.16 | 0.005 |
| GO:0052689 | carboxylic ester hydrolase activity | 136 | 1.83 | 0.011 |
| GO:0016917 | GABA receptor activity | 22 | 3.33 | 0.015 |
| GO:0019208 | phosphatase regulator activity | 88 | 2.00 | 0.016 |
| GO:0008519 | ammonium transmembrane transporter activity | 32 | 2.74 | 0.020 |
| GO:0001085 | RNA polymerase II transcription factor binding | 140 | 1.67 | 0.029 |
| GO:0019787 | ubiquitin-like protein transferase activity | 417 | 1.33 | 0.042 |
| GO:0051861 | glycolipid binding | 20 | 2.93 | 0.044 |
| GO:0000149 | SNARE binding | 102 | 1.72 | 0.045 |
| GO:0015077 | monovalent inorganic cation transmembrane transporter activity | 382 | 1.34 | 0.046 |
| GO:0046906 | tetrapyrrole binding | 138 | 1.59 | 0.050 |

**Supplemental Table S5. Common significant genes identified by both S-MultiXcan and MAGMA tool**

| **Gene name** | **S-MultiXcan P value** | **MAGMA P value** | **Reported studies on oral cancer** | **Cancer-related genes from the GWAS catalog** |
| --- | --- | --- | --- | --- |
| *ATRN* | 5.90E-06 | 3.28E-06 | Non-documented | Non-documented |
| *VWA7* | 1.64E-05 | 4.72E-05 | Non-documented | Non-documented |
| *FGD4* | 3.79E-05 | 2.34E-03 | Non-documented | Documented |
| *TERT* | 6.61E-05 | 2.89E-03 | Non-documented | Documented |
| *HLA-DRB1* | 8.51E-05 | 1.45E-02 | Non-documented | Documented |
| *CDH3* | 8.84E-05 | 1.87E-04 | Non-documented | Documented |
| *FAM227B* | 9.88E-05 | 8.16E-05 | Non-documented | Documented |
| *IRF4* | 1.06E-04 | 2.50E-09 | Non-documented | Documented |
| *HLA-DQA1* | 1.19E-04 | 6.45E-06 | Reported | Documented |
| *TP63* | 1.73E-04 | 4.80E-02 | Non-documented | Documented |
| *LY6G6C* | 1.95E-04 | 3.64E-02 | Non-documented | Non-documented |
| *C4A* | 2.12E-04 | 8.55E-04 | Non-documented | Documented |
| *TAMM41* | 2.27E-04 | 1.91E-03 | Non-documented | Non-documented |
| *CLPTM1L* | 2.66E-04 | 2.13E-05 | Reported | Documented |
| *C4B* | 2.76E-04 | 4.65E-04 | Non-documented | Documented |
| *ZFP90* | 2.93E-04 | 2.37E-06 | Non-documented | Non-documented |
| *CYP21A2* | 3.16E-04 | 3.33E-04 | Non-documented | Documented |
| *SLC6A2* | 3.21E-04 | 2.76E-05 | Non-documented | Non-documented |
| *CELSR1* | 3.27E-04 | 4.94E-02 | Non-documented | Non-documented |
| *NOTCH1* | 3.35E-04 | 2.13E-06 | Non-documented | Non-documented |
| *NLRP12* | 5.02E-04 | 3.26E-05 | Non-documented | Documented |
| *BAG6* | 5.57E-04 | 1.30E-03 | Non-documented | Documented |
| *MTG2* | 5.60E-04 | 1.60E-02 | Non-documented | Non-documented |
| *C2* | 6.62E-04 | 7.66E-03 | Non-documented | Documented |
| *CEBPZ* | 6.74E-04 | 1.25E-02 | Non-documented | Non-documented |
| *TNXB* | 6.87E-04 | 1.45E-04 | Non-documented | Documented |
| *SECISBP2L* | 6.91E-04 | 1.07E-03 | Non-documented | Documented |
| *GPANK1* | 7.33E-04 | 4.34E-03 | Non-documented | Non-documented |
| *MYH7* | 7.40E-04 | 3.51E-02 | Non-documented | Non-documented |
| *ACP6* | 7.41E-04 | 4.67E-03 | Non-documented | Non-documented |
| *QPCT* | 7.92E-04 | 4.81E-02 | Non-documented | Non-documented |
| *KLHL24* | 8.65E-04 | 1.15E-04 | Non-documented | Non-documented |
| *SNRPA1* | 8.80E-04 | 3.55E-03 | Non-documented | Documented |
| *CLCA4* | 9.22E-04 | 7.58E-04 | Non-documented | Non-documented |
| *EPHX2* | 9.37E-04 | 2.03E-05 | Non-documented | Documented |
| *DTWD1* | 1.05E-03 | 1.23E-02 | Non-documented | Documented |
| *TMCC1* | 1.08E-03 | 8.91E-04 | Non-documented | Non-documented |
| *GSTA1* | 1.11E-03 | 5.29E-03 | Non-documented | Non-documented |
| *NAE1* | 1.13E-03 | 4.71E-02 | Non-documented | Non-documented |
| *OVOL3* | 1.13E-03 | 8.20E-03 | Non-documented | Non-documented |
| *TP53INP1* | 1.24E-03 | 3.69E-02 | Non-documented | Non-documented |
| *FGF7* | 1.37E-03 | 2.78E-05 | Non-documented | Documented |
| *ZNF32* | 1.48E-03 | 4.32E-02 | Non-documented | Non-documented |
| *TPTE2* | 1.51E-03 | 2.98E-02 | Non-documented | Non-documented |
| *NPY* | 1.57E-03 | 1.34E-04 | Non-documented | Non-documented |
| *CAPN14* | 1.60E-03 | 3.46E-02 | Non-documented | Non-documented |
| *RNF144B* | 1.66E-03 | 5.26E-03 | Non-documented | Non-documented |
| *ZCRB1* | 1.67E-03 | 1.13E-03 | Non-documented | Non-documented |
| *FBRS* | 1.68E-03 | 1.13E-03 | Non-documented | Non-documented |
| *NCAPG* | 1.98E-03 | 7.96E-04 | Non-documented | Non-documented |
| *FAM96B* | 2.01E-03 | 4.30E-04 | Non-documented | Non-documented |
| *CES3* | 2.13E-03 | 7.53E-04 | Non-documented | Non-documented |
| *FABP7* | 2.17E-03 | 8.53E-04 | Non-documented | Non-documented |
| *HLA-DMB* | 2.42E-03 | 1.25E-03 | Non-documented | Documented |
| *RNASE8* | 2.42E-03 | 1.94E-03 | Non-documented | Non-documented |
| *MSH5* | 2.44E-03 | 6.23E-05 | Non-documented | Documented |
| *MIB1* | 2.48E-03 | 8.65E-03 | Non-documented | Non-documented |
| *WDR5B* | 2.51E-03 | 4.12E-03 | Non-documented | Non-documented |
| *ZNF512* | 2.55E-03 | 5.86E-04 | Reported | Non-documented |
| *SRCAP* | 2.75E-03 | 2.98E-04 | Non-documented | Non-documented |
| *CES2* | 2.95E-03 | 4.45E-04 | Non-documented | Non-documented |
| *IGFBP3* | 2.99E-03 | 1.77E-03 | Non-documented | Non-documented |
| *CD320* | 3.00E-03 | 1.70E-03 | Non-documented | Non-documented |
| *UBD* | 3.01E-03 | 1.50E-02 | Non-documented | Non-documented |
| *HMGB2* | 3.03E-03 | 4.95E-03 | Non-documented | Non-documented |
| *HLA-DQB1* | 3.08E-03 | 1.19E-04 | Reported | Documented |
| *DCAF16* | 3.13E-03 | 1.11E-03 | Non-documented | Non-documented |
| *TCF4* | 3.15E-03 | 8.76E-03 | Non-documented | Documented |
| *ZCCHC24* | 3.18E-03 | 4.94E-04 | Non-documented | Non-documented |
| *GOT1* | 3.20E-03 | 2.04E-02 | Non-documented | Non-documented |
| *RRAD* | 3.23E-03 | 1.04E-04 | Non-documented | Non-documented |
| *EHD3* | 3.24E-03 | 2.86E-04 | Non-documented | Non-documented |
| *NPM2* | 3.30E-03 | 9.59E-04 | Non-documented | Non-documented |
| *PRRC2A* | 3.35E-03 | 3.32E-03 | Non-documented | Non-documented |
| *MUC17* | 3.41E-03 | 6.59E-03 | Non-documented | Non-documented |
| *KCMF1* | 3.60E-03 | 1.46E-03 | Non-documented | Non-documented |
| *NAMPT* | 3.68E-03 | 2.47E-02 | Non-documented | Non-documented |
| *RNF13* | 3.77E-03 | 1.91E-03 | Non-documented | Non-documented |
| *MYRF* | 3.79E-03 | 4.59E-02 | Non-documented | Documented |
| *BARX2* | 3.91E-03 | 1.30E-02 | Non-documented | Documented |
| *ASCL5* | 4.17E-03 | 1.13E-03 | Non-documented | Non-documented |
| *HLA-DMA* | 4.18E-03 | 8.68E-04 | Non-documented | Non-documented |
| *ACKR3* | 4.23E-03 | 3.59E-04 | Non-documented | Non-documented |
| *RAB24* | 4.32E-03 | 3.60E-02 | Non-documented | Non-documented |
| *SAPCD1* | 4.44E-03 | 4.24E-05 | Non-documented | Non-documented |
| *TNS3* | 4.45E-03 | 1.44E-06 | Non-documented | Documented |
| *PRKD3* | 4.89E-03 | 4.68E-03 | Non-documented | Non-documented |
| *SHARPIN* | 4.91E-03 | 3.79E-02 | Non-documented | Non-documented |
| *COPZ2* | 4.95E-03 | 2.05E-04 | Non-documented | Non-documented |
| *PDP1* | 5.06E-03 | 5.22E-03 | Non-documented | Non-documented |
| *LGI1* | 5.06E-03 | 1.91E-02 | Non-documented | Non-documented |
| *C6orf48* | 5.19E-03 | 3.11E-03 | Non-documented | Documented |
| *CYR61* | 5.34E-03 | 3.37E-03 | Non-documented | Non-documented |
| *CDC5L* | 5.36E-03 | 8.22E-03 | Non-documented | Non-documented |
| *GALNT14* | 5.41E-03 | 3.42E-03 | Reported | Non-documented |
| *MRAS* | 5.42E-03 | 9.54E-03 | Non-documented | Non-documented |
| *PSORS1C1* | 5.70E-03 | 7.67E-03 | Non-documented | Documented |
| *FMOD* | 5.72E-03 | 2.58E-03 | Non-documented | Non-documented |
| *CCDC121* | 6.08E-03 | 7.96E-04 | Reported | Non-documented |
| *MPP5* | 6.11E-03 | 1.38E-03 | Non-documented | Non-documented |
| *DRD2* | 6.12E-03 | 2.00E-05 | Non-documented | Non-documented |
| *YAF2* | 6.20E-03 | 1.15E-02 | Non-documented | Non-documented |
| *PTPN18* | 6.38E-03 | 4.05E-03 | Non-documented | Non-documented |
| *CDC27* | 6.53E-03 | 1.00E-02 | Non-documented | Non-documented |
| *CCDC58* | 6.60E-03 | 2.60E-02 | Non-documented | Non-documented |
| *WBSCR27* | 6.70E-03 | 2.16E-02 | Non-documented | Non-documented |
| *P2RX1* | 7.06E-03 | 2.04E-04 | Non-documented | Non-documented |
| *CD302* | 7.18E-03 | 1.18E-02 | Non-documented | Non-documented |
| *DEFB1* | 7.23E-03 | 5.58E-03 | Non-documented | Non-documented |
| *RASSF7* | 7.26E-03 | 1.42E-02 | Non-documented | Non-documented |
| *ANKRD9* | 7.41E-03 | 3.14E-02 | Non-documented | Non-documented |
| *LCORL* | 7.49E-03 | 7.85E-04 | Non-documented | Documented |
| *SLC44A4* | 7.51E-03 | 1.78E-02 | Non-documented | Documented |
| *ZNF391* | 7.61E-03 | 7.95E-04 | Non-documented | Non-documented |
| *ZBTB12* | 7.65E-03 | 1.42E-02 | Non-documented | Non-documented |
| *JAK1* | 7.67E-03 | 7.86E-04 | Non-documented | Documented |
| *APOM* | 7.81E-03 | 1.62E-03 | Non-documented | Documented |
| *CLIC1* | 7.84E-03 | 7.20E-03 | Non-documented | Non-documented |
| *SNX11* | 7.88E-03 | 1.54E-03 | Non-documented | Non-documented |
| *CRISPLD1* | 7.88E-03 | 4.20E-02 | Non-documented | Non-documented |
| *ZNF496* | 7.97E-03 | 1.41E-02 | Non-documented | Non-documented |
| *CCDC115* | 8.00E-03 | 5.89E-03 | Non-documented | Non-documented |
| *EIF2S1* | 8.08E-03 | 3.76E-03 | Non-documented | Non-documented |
| *PDP2* | 8.25E-03 | 8.13E-05 | Non-documented | Non-documented |
| *MATN3* | 8.33E-03 | 3.65E-02 | Non-documented | Non-documented |
| *CXCR2* | 8.53E-03 | 9.45E-03 | Non-documented | Non-documented |
| *ARHGEF10L* | 8.62E-03 | 4.30E-03 | Non-documented | Documented |
| *VIPAS39* | 8.66E-03 | 1.01E-02 | Non-documented | Non-documented |
| *S100A5* | 8.67E-03 | 1.73E-03 | Non-documented | Non-documented |
| *ALDH9A1* | 8.68E-03 | 4.10E-03 | Non-documented | Non-documented |
| *ABO* | 8.73E-03 | 2.07E-03 | Non-documented | Documented |
| *GPHN* | 8.79E-03 | 2.30E-02 | Non-documented | Non-documented |
| *TOMM5* | 8.81E-03 | 3.96E-02 | Non-documented | Non-documented |
| *S100A2* | 8.84E-03 | 5.65E-04 | Non-documented | Non-documented |
| *RBBP5* | 8.85E-03 | 3.47E-02 | Non-documented | Non-documented |
| *MAD1L1* | 8.89E-03 | 2.64E-02 | Non-documented | Documented |
| *NEFL* | 9.03E-03 | 1.74E-03 | Non-documented | Non-documented |
| *S100A3* | 9.26E-03 | 1.29E-03 | Non-documented | Non-documented |
| *ATP6V1D* | 9.35E-03 | 2.59E-03 | Non-documented | Non-documented |
| *DCP1B* | 9.54E-03 | 2.09E-02 | Non-documented | Documented |
| *CTXN3* | 9.56E-03 | 3.22E-02 | Non-documented | Non-documented |
| *CACNA1S* | 9.57E-03 | 1.19E-02 | Non-documented | Non-documented |
| *PPHLN1* | 9.75E-03 | 7.43E-04 | Non-documented | Non-documented |
| *DAGLA* | 9.76E-03 | 1.14E-02 | Non-documented | Non-documented |
| *FAM173A* | 9.86E-03 | 1.30E-02 | Non-documented | Non-documented |
| *DISC1* | 9.90E-03 | 7.11E-03 | Non-documented | Non-documented |
| *PPIF* | 9.91E-03 | 6.00E-03 | Non-documented | Non-documented |
| *MAP3K4* | 9.91E-03 | 1.04E-03 | Non-documented | Non-documented |
| *NDUFA7* | 1.00E-02 | 5.47E-03 | Non-documented | Non-documented |
| *MAPRE1* | 1.01E-02 | 2.93E-03 | Non-documented | Non-documented |
| *VARS* | 1.02E-02 | 1.25E-04 | Non-documented | Non-documented |
| *DUSP13* | 1.03E-02 | 7.64E-03 | Non-documented | Non-documented |
| *PPM1G* | 1.03E-02 | 9.71E-03 | Non-documented | Non-documented |
| *CXCL12* | 1.05E-02 | 3.37E-03 | Non-documented | Non-documented |
| *BRD1* | 1.05E-02 | 2.10E-02 | Non-documented | Non-documented |
| *ECHDC3* | 1.05E-02 | 4.66E-02 | Non-documented | Documented |
| *MPP6* | 1.07E-02 | 5.54E-03 | Non-documented | Non-documented |
| *DMXL1* | 1.07E-02 | 2.46E-02 | Non-documented | Non-documented |
| *JMJD1C* | 1.09E-02 | 8.85E-03 | Non-documented | Non-documented |
| *NRM* | 1.10E-02 | 2.67E-02 | Non-documented | Non-documented |
| *CDH1* | 1.12E-02 | 3.60E-03 | Non-documented | Documented |
| *CEP57* | 1.13E-02 | 3.42E-02 | Non-documented | Non-documented |
| *CTLA4* | 1.16E-02 | 1.09E-02 | Reported | Documented |
| *COMMD1* | 1.17E-02 | 9.89E-03 | Non-documented | Non-documented |
| *CCDC12* | 1.18E-02 | 4.86E-03 | Non-documented | Non-documented |
| *SLC25A45* | 1.18E-02 | 1.78E-02 | Non-documented | Non-documented |
| *MPHOSPH8* | 1.19E-02 | 1.43E-03 | Non-documented | Non-documented |
| *FAM69B* | 1.19E-02 | 4.27E-03 | Non-documented | Non-documented |
| *HNRNPL* | 1.19E-02 | 5.14E-03 | Non-documented | Non-documented |
| *ERCC8* | 1.22E-02 | 9.54E-03 | Non-documented | Non-documented |
| *ZNF629* | 1.22E-02 | 9.90E-03 | Non-documented | Non-documented |
| *ZNF184* | 1.23E-02 | 2.42E-03 | Non-documented | Documented |
| *BRD2* | 1.24E-02 | 2.95E-03 | Non-documented | Non-documented |
| *HSPB3* | 1.24E-02 | 2.51E-03 | Non-documented | Non-documented |
| *NOD1* | 1.25E-02 | 2.14E-02 | Non-documented | Non-documented |
| *LCE3C* | 1.26E-02 | 8.72E-03 | Non-documented | Non-documented |
| *SLC25A38* | 1.26E-02 | 2.80E-03 | Non-documented | Non-documented |
| *PHF7* | 1.27E-02 | 1.30E-02 | Non-documented | Non-documented |
| *ASNS* | 1.27E-02 | 4.59E-02 | Non-documented | Non-documented |
| *CALN1* | 1.28E-02 | 6.75E-03 | Non-documented | Non-documented |
| *CGRRF1* | 1.29E-02 | 3.97E-03 | Non-documented | Non-documented |
| *CAMTA1* | 1.29E-02 | 6.84E-03 | Non-documented | Non-documented |
| *S100A4* | 1.29E-02 | 1.49E-03 | Non-documented | Non-documented |
| *TMUB2* | 1.30E-02 | 8.62E-03 | Non-documented | Non-documented |
| *LDHA* | 1.32E-02 | 6.78E-03 | Non-documented | Non-documented |
| *PPP2R2B* | 1.33E-02 | 2.40E-03 | Non-documented | Non-documented |
| *DPH7* | 1.33E-02 | 5.99E-03 | Non-documented | Non-documented |
| *LDHAL6A* | 1.35E-02 | 7.00E-04 | Non-documented | Non-documented |
| *FGF17* | 1.35E-02 | 2.30E-04 | Non-documented | Non-documented |
| *PPP1R36* | 1.41E-02 | 3.68E-02 | Non-documented | Non-documented |
| *NOVA2* | 1.41E-02 | 2.30E-02 | Non-documented | Non-documented |
| *ZMYM5* | 1.44E-02 | 2.29E-03 | Non-documented | Non-documented |
| *NUDT18* | 1.47E-02 | 4.89E-03 | Non-documented | Non-documented |
| *GML* | 1.48E-02 | 8.07E-03 | Non-documented | Non-documented |
| *NIPSNAP1* | 1.48E-02 | 4.81E-02 | Non-documented | Non-documented |
| *SAAL1* | 1.50E-02 | 5.15E-03 | Non-documented | Non-documented |
| *PDE6C* | 1.52E-02 | 2.24E-02 | Non-documented | Non-documented |
| *ARL1* | 1.54E-02 | 3.55E-02 | Non-documented | Documented |
| *FAM20C* | 1.54E-02 | 6.57E-04 | Non-documented | Non-documented |
| *EAPP* | 1.59E-02 | 3.27E-02 | Non-documented | Non-documented |
| *MYADM* | 1.60E-02 | 5.66E-03 | Non-documented | Non-documented |
| *ANO5* | 1.60E-02 | 4.88E-03 | Non-documented | Non-documented |
| *TPH1* | 1.61E-02 | 5.20E-03 | Non-documented | Non-documented |
| *TMEM239* | 1.62E-02 | 4.73E-02 | Non-documented | Non-documented |
| *HLA-B* | 1.62E-02 | 1.27E-03 | Non-documented | Documented |
| *GEN1* | 1.63E-02 | 4.02E-03 | Non-documented | Non-documented |
| *FERMT2* | 1.64E-02 | 1.28E-02 | Non-documented | Documented |
| *TBCA* | 1.66E-02 | 2.01E-02 | Non-documented | Non-documented |
| *MEX3B* | 1.68E-02 | 2.06E-02 | Non-documented | Documented |
| *ZNF333* | 1.68E-02 | 3.74E-02 | Non-documented | Non-documented |
| *GPN1* | 1.68E-02 | 1.64E-03 | Reported | Non-documented |
| *BTBD2* | 1.69E-02 | 2.50E-03 | Non-documented | Non-documented |
| *SOX18* | 1.70E-02 | 1.66E-02 | Non-documented | Non-documented |
| *MRPL54* | 1.70E-02 | 3.19E-02 | Non-documented | Non-documented |
| *SLC44A5* | 1.72E-02 | 1.79E-02 | Non-documented | Non-documented |
| *VSIG10L* | 1.73E-02 | 1.34E-02 | Non-documented | Non-documented |
| *FNDC4* | 1.73E-02 | 5.84E-03 | Non-documented | Non-documented |
| *KCNQ5* | 1.74E-02 | 1.12E-02 | Non-documented | Non-documented |
| *SELPLG* | 1.76E-02 | 2.03E-03 | Non-documented | Non-documented |
| *THOC5* | 1.78E-02 | 1.94E-02 | Non-documented | Non-documented |
| *EHMT2* | 1.79E-02 | 7.73E-03 | Non-documented | Non-documented |
| *PIKFYVE* | 1.81E-02 | 3.88E-02 | Non-documented | Non-documented |
| *HAUS8* | 1.82E-02 | 7.01E-03 | Non-documented | Non-documented |
| *MAP6D1* | 1.83E-02 | 3.74E-02 | Non-documented | Non-documented |
| *GCKR* | 1.84E-02 | 4.49E-03 | Non-documented | Non-documented |
| *RANBP6* | 1.85E-02 | 1.82E-02 | Non-documented | Non-documented |
| *KANK3* | 1.86E-02 | 5.31E-03 | Non-documented | Non-documented |
| *SIGLEC10* | 1.86E-02 | 2.90E-02 | Non-documented | Non-documented |
| *FAM76B* | 1.87E-02 | 3.19E-02 | Non-documented | Non-documented |
| *MAF1* | 1.88E-02 | 4.16E-02 | Non-documented | Non-documented |
| *NYAP2* | 1.90E-02 | 1.86E-02 | Non-documented | Documented |
| *CDKN3* | 1.92E-02 | 1.85E-02 | Non-documented | Documented |
| *ZNF514* | 1.92E-02 | 1.75E-02 | Non-documented | Non-documented |
| *VSNL1* | 1.92E-02 | 4.20E-03 | Non-documented | Documented |
| *SMPDL3A* | 1.93E-02 | 1.04E-03 | Non-documented | Non-documented |
| *FOXE3* | 1.93E-02 | 1.46E-03 | Non-documented | Non-documented |
| *REEP3* | 1.94E-02 | 8.03E-03 | Non-documented | Non-documented |
| *RNF34* | 1.96E-02 | 1.40E-03 | Non-documented | Non-documented |
| *CBX1* | 1.97E-02 | 7.75E-04 | Non-documented | Non-documented |
| *PHTF1* | 2.00E-02 | 5.72E-03 | Non-documented | Documented |
| *GPHA2* | 2.01E-02 | 1.40E-02 | Non-documented | Non-documented |
| *GFRA4* | 2.02E-02 | 5.92E-04 | Non-documented | Non-documented |
| *KRT3* | 2.02E-02 | 6.64E-04 | Non-documented | Non-documented |
| *KPNA6* | 2.03E-02 | 2.42E-02 | Non-documented | Non-documented |
| *SERINC3* | 2.06E-02 | 8.30E-03 | Non-documented | Non-documented |
| *RNF4* | 2.06E-02 | 1.26E-03 | Non-documented | Documented |
| *CMTM3* | 2.07E-02 | 2.67E-02 | Non-documented | Non-documented |
| *NARS2* | 2.08E-02 | 2.26E-02 | Non-documented | Non-documented |
| *PTPN22* | 2.08E-02 | 8.42E-03 | Non-documented | Documented |
| *CLDN4* | 2.09E-02 | 2.53E-02 | Non-documented | Non-documented |
| *GLYCTK* | 2.09E-02 | 9.12E-03 | Non-documented | Non-documented |
| *HIST1H2AH* | 2.14E-02 | 2.02E-02 | Non-documented | Non-documented |
| *PGLYRP1* | 2.16E-02 | 1.21E-02 | Non-documented | Non-documented |
| *TMEM106C* | 2.17E-02 | 2.36E-02 | Non-documented | Non-documented |
| *MRPL33* | 2.18E-02 | 1.23E-02 | Non-documented | Non-documented |
| *SERGEF* | 2.19E-02 | 1.36E-02 | Non-documented | Non-documented |
| *TPRG1L* | 2.19E-02 | 5.57E-03 | Non-documented | Non-documented |
| *ZSCAN18* | 2.23E-02 | 1.40E-02 | Non-documented | Non-documented |
| *VRK3* | 2.23E-02 | 3.43E-02 | Non-documented | Non-documented |
| *USP15* | 2.23E-02 | 1.90E-02 | Non-documented | Non-documented |
| *KRTAP8-1* | 2.24E-02 | 3.42E-02 | Non-documented | Non-documented |
| *MRPS18C* | 2.24E-02 | 2.66E-02 | Non-documented | Non-documented |
| *LARP4* | 2.25E-02 | 2.67E-02 | Non-documented | Non-documented |
| *ZNF689* | 2.25E-02 | 4.22E-02 | Non-documented | Non-documented |
| *ARHGEF26* | 2.25E-02 | 2.39E-02 | Non-documented | Non-documented |
| *GCSAML* | 2.25E-02 | 3.17E-03 | Non-documented | Non-documented |
| *EFCAB8* | 2.26E-02 | 2.85E-03 | Non-documented | Non-documented |
| *CRADD* | 2.26E-02 | 1.29E-02 | Non-documented | Non-documented |
| *GNG8* | 2.27E-02 | 4.79E-02 | Non-documented | Non-documented |
| *OTOGL* | 2.27E-02 | 2.63E-02 | Non-documented | Non-documented |
| *ELP6* | 2.28E-02 | 4.28E-02 | Non-documented | Non-documented |
| *ANKRD2* | 2.29E-02 | 2.41E-03 | Non-documented | Non-documented |
| *TMEM81* | 2.30E-02 | 1.90E-02 | Non-documented | Non-documented |
| *NFE2L1* | 2.32E-02 | 1.97E-04 | Non-documented | Non-documented |
| *BCL2L15* | 2.33E-02 | 1.77E-03 | Non-documented | Documented |
| *ZNF843* | 2.34E-02 | 2.97E-03 | Non-documented | Non-documented |
| *PKIG* | 2.34E-02 | 3.81E-02 | Non-documented | Documented |
| *SHPRH* | 2.36E-02 | 2.51E-03 | Non-documented | Non-documented |
| *XPR1* | 2.36E-02 | 5.92E-03 | Non-documented | Non-documented |
| *RPP40* | 2.43E-02 | 5.62E-04 | Non-documented | Non-documented |
| *IMP4* | 2.45E-02 | 4.63E-03 | Non-documented | Non-documented |
| *HDDC3* | 2.46E-02 | 2.97E-02 | Non-documented | Non-documented |
| *INSR* | 2.46E-02 | 3.02E-02 | Non-documented | Non-documented |
| *CNNM1* | 2.46E-02 | 1.74E-02 | Non-documented | Documented |
| *QDPR* | 2.49E-02 | 2.46E-02 | Non-documented | Non-documented |
| *AHSA1* | 2.49E-02 | 1.50E-02 | Non-documented | Non-documented |
| *RNF40* | 2.50E-02 | 1.46E-02 | Non-documented | Non-documented |
| *IQCC* | 2.51E-02 | 3.66E-02 | Non-documented | Non-documented |
| *CACFD1* | 2.51E-02 | 3.40E-02 | Non-documented | Non-documented |
| *FAM71D* | 2.52E-02 | 9.63E-03 | Non-documented | Non-documented |
| *ZMIZ2* | 2.58E-02 | 7.92E-03 | Non-documented | Non-documented |
| *PSPC1* | 2.58E-02 | 1.56E-03 | Non-documented | Non-documented |
| *ADGRE3* | 2.59E-02 | 3.23E-02 | Non-documented | Non-documented |
| *C16orf72* | 2.64E-02 | 1.60E-02 | Non-documented | Documented |
| *RHPN1* | 2.66E-02 | 1.08E-03 | Non-documented | Non-documented |
| *PON3* | 2.67E-02 | 4.59E-02 | Non-documented | Non-documented |
| *KCNF1* | 2.67E-02 | 2.04E-02 | Non-documented | Non-documented |
| *FOXD2* | 2.69E-02 | 8.11E-03 | Non-documented | Non-documented |
| *PRR14* | 2.70E-02 | 5.93E-03 | Non-documented | Non-documented |
| *ZNF329* | 2.70E-02 | 5.37E-03 | Non-documented | Non-documented |
| *DENND2A* | 2.72E-02 | 3.61E-03 | Non-documented | Non-documented |
| *FAM64A* | 2.72E-02 | 1.35E-02 | Non-documented | Non-documented |
| *RBBP8* | 2.73E-02 | 3.66E-02 | Non-documented | Non-documented |
| *EPM2A* | 2.74E-02 | 1.22E-03 | Non-documented | Non-documented |
| *CFAP99* | 2.74E-02 | 4.56E-03 | Non-documented | Non-documented |
| *BVES* | 2.74E-02 | 3.15E-02 | Non-documented | Non-documented |
| *SLC9A2* | 2.75E-02 | 2.91E-02 | Non-documented | Non-documented |
| *AP4B1* | 2.78E-02 | 1.52E-02 | Non-documented | Documented |
| *DNAL4* | 2.80E-02 | 2.63E-03 | Non-documented | Non-documented |
| *PPT2* | 2.81E-02 | 9.36E-03 | Non-documented | Documented |
| *CPD* | 2.84E-02 | 8.88E-03 | Non-documented | Non-documented |
| *RBMXL2* | 2.87E-02 | 4.46E-02 | Non-documented | Non-documented |
| *GALK2* | 2.87E-02 | 4.88E-02 | Non-documented | Documented |
| *PLIN2* | 2.88E-02 | 3.35E-02 | Non-documented | Non-documented |
| *NDUFAF7* | 2.88E-02 | 9.18E-04 | Non-documented | Non-documented |
| *KPNA1* | 2.91E-02 | 9.56E-03 | Non-documented | Non-documented |
| *NBEAL2* | 2.91E-02 | 1.25E-02 | Non-documented | Non-documented |
| *NSMCE2* | 2.92E-02 | 1.04E-02 | Non-documented | Non-documented |
| *BIRC6* | 2.92E-02 | 3.90E-03 | Non-documented | Non-documented |
| *CDH16* | 2.93E-02 | 3.46E-05 | Non-documented | Non-documented |
| *DUPD1* | 2.94E-02 | 1.24E-02 | Non-documented | Non-documented |
| *LPCAT3* | 2.97E-02 | 3.85E-02 | Non-documented | Documented |
| *MATK* | 2.97E-02 | 1.44E-02 | Non-documented | Non-documented |
| *WDR62* | 2.98E-02 | 8.75E-03 | Non-documented | Non-documented |
| *TALDO1* | 2.98E-02 | 3.03E-02 | Non-documented | Non-documented |
| *E2F2* | 2.98E-02 | 1.08E-02 | Non-documented | Non-documented |
| *EIF4E* | 3.00E-02 | 6.01E-03 | Non-documented | Non-documented |
| *SAMD15* | 3.01E-02 | 1.72E-02 | Non-documented | Non-documented |
| *EIF3I* | 3.01E-02 | 4.33E-02 | Non-documented | Non-documented |
| *UROD* | 3.02E-02 | 6.00E-03 | Non-documented | Non-documented |
| *C6orf15* | 3.02E-02 | 1.47E-03 | Non-documented | Documented |
| *FAM162A* | 3.03E-02 | 3.46E-03 | Non-documented | Non-documented |
| *SCCPDH* | 3.04E-02 | 2.39E-02 | Non-documented | Non-documented |
| *FKBPL* | 3.06E-02 | 1.97E-03 | Non-documented | Documented |
| *TMEM234* | 3.06E-02 | 4.15E-02 | Non-documented | Non-documented |
| *UBN1* | 3.08E-02 | 4.99E-02 | Non-documented | Non-documented |
| *SRRT* | 3.09E-02 | 2.88E-02 | Non-documented | Non-documented |
| *UBA52* | 3.10E-02 | 2.91E-02 | Non-documented | Non-documented |
| *AGO1* | 3.11E-02 | 1.95E-02 | Non-documented | Non-documented |
| *FAM173B* | 3.11E-02 | 3.71E-02 | Non-documented | Non-documented |
| *ZNF14* | 3.12E-02 | 2.82E-03 | Non-documented | Non-documented |
| *CD82* | 3.16E-02 | 1.28E-02 | Non-documented | Non-documented |
| *TFEC* | 3.16E-02 | 3.09E-02 | Non-documented | Non-documented |
| *OR11A1* | 3.18E-02 | 1.04E-02 | Non-documented | Non-documented |
| *SPATS1* | 3.20E-02 | 4.45E-02 | Non-documented | Non-documented |
| *TOP1MT* | 3.21E-02 | 8.37E-03 | Non-documented | Non-documented |
| *CSNK2B* | 3.22E-02 | 7.92E-03 | Non-documented | Non-documented |
| *TYW3* | 3.29E-02 | 2.12E-02 | Non-documented | Non-documented |
| *ZIM2* | 3.30E-02 | 1.96E-02 | Non-documented | Non-documented |
| *GIGYF2* | 3.31E-02 | 4.05E-03 | Non-documented | Non-documented |
| *LYSMD4* | 3.33E-02 | 4.22E-02 | Non-documented | Non-documented |
| *NIPSNAP3A* | 3.37E-02 | 3.61E-02 | Non-documented | Non-documented |
| *SLC44A1* | 3.39E-02 | 3.39E-02 | Non-documented | Documented |
| *EPN1* | 3.39E-02 | 3.64E-02 | Non-documented | Non-documented |
| *MAS1L* | 3.40E-02 | 6.75E-03 | Non-documented | Non-documented |
| *ABHD6* | 3.40E-02 | 2.78E-02 | Non-documented | Non-documented |
| *C6orf47* | 3.41E-02 | 2.31E-03 | Non-documented | Non-documented |
| *FCGR1B* | 3.43E-02 | 6.19E-05 | Non-documented | Non-documented |
| *CCDC33* | 3.44E-02 | 2.04E-03 | Non-documented | Non-documented |
| *ECH1* | 3.44E-02 | 5.02E-03 | Non-documented | Non-documented |
| *EIF2B4* | 3.44E-02 | 8.63E-03 | Non-documented | Non-documented |
| *GAB2* | 3.45E-02 | 1.37E-02 | Non-documented | Documented |
| *PRKD1* | 3.45E-02 | 4.06E-02 | Non-documented | Non-documented |
| *EIF2B3* | 3.47E-02 | 3.93E-02 | Non-documented | Non-documented |
| *TEKT3* | 3.48E-02 | 7.29E-03 | Non-documented | Non-documented |
| *C17orf53* | 3.50E-02 | 8.51E-03 | Non-documented | Non-documented |
| *TMED8* | 3.56E-02 | 2.21E-02 | Non-documented | Non-documented |
| *IDH1* | 3.56E-02 | 4.51E-02 | Non-documented | Non-documented |
| *SENP6* | 3.58E-02 | 3.16E-02 | Non-documented | Non-documented |
| *TTPAL* | 3.59E-02 | 9.08E-03 | Non-documented | Non-documented |
| *ZBTB20* | 3.60E-02 | 4.45E-02 | Non-documented | Documented |
| *FBXO30* | 3.61E-02 | 4.55E-03 | Non-documented | Non-documented |
| *PTGIR* | 3.61E-02 | 4.40E-02 | Non-documented | Non-documented |
| *C2orf16* | 3.62E-02 | 1.08E-03 | Reported | Non-documented |
| *ANKS6* | 3.62E-02 | 3.71E-02 | Non-documented | Non-documented |
| *CNTN3* | 3.63E-02 | 2.64E-02 | Non-documented | Non-documented |
| *PRR31* | 3.64E-02 | 2.86E-02 | Non-documented | Non-documented |
| *NEU1* | 3.65E-02 | 5.38E-03 | Non-documented | Documented |
| *MBD3* | 3.65E-02 | 2.68E-02 | Non-documented | Non-documented |
| *PHKG2* | 3.65E-02 | 1.91E-03 | Non-documented | Non-documented |
| *TMEM240* | 3.66E-02 | 1.26E-02 | Non-documented | Non-documented |
| *FAAH* | 3.72E-02 | 3.55E-02 | Non-documented | Non-documented |
| *MRPS5* | 3.72E-02 | 2.55E-02 | Non-documented | Non-documented |
| *RIMS1* | 3.76E-02 | 1.05E-02 | Non-documented | Documented |
| *RAB5C* | 3.76E-02 | 3.93E-02 | Non-documented | Non-documented |
| *CTRB1* | 3.77E-02 | 1.75E-02 | Non-documented | Documented |
| *ZNF558* | 3.78E-02 | 1.05E-02 | Non-documented | Non-documented |
| *MAP3K11* | 3.79E-02 | 2.35E-02 | Non-documented | Non-documented |
| *HSPA1L* | 3.80E-02 | 6.14E-04 | Non-documented | Non-documented |
| *GOSR1* | 3.81E-02 | 1.93E-02 | Non-documented | Non-documented |
| *MAL* | 3.86E-02 | 1.99E-02 | Non-documented | Non-documented |
| *COPS2* | 3.87E-02 | 4.25E-02 | Non-documented | Documented |
| *BCAR1* | 3.87E-02 | 2.12E-02 | Non-documented | Documented |
| *KCNA4* | 3.87E-02 | 1.20E-02 | Non-documented | Non-documented |
| *KCNE3* | 3.88E-02 | 1.46E-02 | Non-documented | Non-documented |
| *CNOT4* | 3.88E-02 | 5.21E-03 | Non-documented | Non-documented |
| *CDC42EP2* | 3.95E-02 | 2.33E-02 | Non-documented | Non-documented |
| *ENTPD8* | 3.97E-02 | 1.58E-02 | Non-documented | Non-documented |
| *GABRB2* | 3.97E-02 | 4.80E-03 | Non-documented | Non-documented |
| *CTNNA3* | 3.99E-02 | 1.02E-02 | Non-documented | Documented |
| *C10orf142* | 3.99E-02 | 4.69E-03 | Non-documented | Non-documented |
| *ADAM21* | 3.99E-02 | 4.34E-02 | Non-documented | Documented |
| *ACHE* | 4.00E-02 | 2.19E-02 | Non-documented | Non-documented |
| *CDH23* | 4.00E-02 | 2.93E-02 | Non-documented | Non-documented |
| *C10orf62* | 4.01E-02 | 9.83E-04 | Non-documented | Non-documented |
| *RBKS* | 4.02E-02 | 4.82E-03 | Non-documented | Non-documented |
| *EGFL8* | 4.02E-02 | 8.74E-03 | Non-documented | Documented |
| *GLYR1* | 4.03E-02 | 4.97E-02 | Non-documented | Documented |
| *EPHB4* | 4.04E-02 | 2.12E-02 | Non-documented | Non-documented |
| *SLC12A9* | 4.05E-02 | 7.30E-03 | Non-documented | Non-documented |
| *NAALADL1* | 4.05E-02 | 4.71E-02 | Non-documented | Non-documented |
| *ACO1* | 4.09E-02 | 7.77E-03 | Non-documented | Non-documented |
| *ARRDC1* | 4.09E-02 | 9.17E-03 | Non-documented | Non-documented |
| *RASGEF1A* | 4.09E-02 | 3.23E-02 | Non-documented | Non-documented |
| *HLA-DRA* | 4.16E-02 | 2.80E-02 | Non-documented | Documented |
| *MGST3* | 4.19E-02 | 3.18E-02 | Non-documented | Non-documented |
| *AKIRIN1* | 4.20E-02 | 1.19E-02 | Non-documented | Non-documented |
| *PTPN23* | 4.24E-02 | 3.50E-02 | Non-documented | Non-documented |
| *EHMT1* | 4.24E-02 | 2.62E-02 | Non-documented | Non-documented |
| *DCN* | 4.25E-02 | 1.79E-02 | Non-documented | Non-documented |
| *RINL* | 4.27E-02 | 2.42E-02 | Non-documented | Non-documented |
| *IGSF5* | 4.29E-02 | 2.41E-02 | Non-documented | Non-documented |
| *GPR135* | 4.30E-02 | 3.95E-02 | Non-documented | Non-documented |
| *KRTCAP3* | 4.30E-02 | 7.88E-03 | Non-documented | Non-documented |
| *KCNJ13* | 4.30E-02 | 6.03E-03 | Non-documented | Non-documented |
| *EAF2* | 4.31E-02 | 3.39E-02 | Non-documented | Non-documented |
| *ATP2A3* | 4.32E-02 | 6.87E-05 | Non-documented | Non-documented |
| *SLC13A3* | 4.32E-02 | 2.40E-02 | Non-documented | Non-documented |
| *AGPAT1* | 4.33E-02 | 1.02E-02 | Non-documented | Documented |
| *LIPJ* | 4.33E-02 | 3.71E-02 | Non-documented | Non-documented |
| *SLC22A18* | 4.34E-02 | 4.12E-02 | Non-documented | Non-documented |
| *RNF5* | 4.35E-02 | 1.37E-02 | Non-documented | Documented |
| *TFEB* | 4.36E-02 | 2.69E-02 | Non-documented | Documented |
| *CYP11B1* | 4.40E-02 | 1.10E-02 | Non-documented | Non-documented |
| *ZNF276* | 4.41E-02 | 3.76E-02 | Non-documented | Documented |
| *BATF2* | 4.42E-02 | 4.98E-02 | Non-documented | Non-documented |
| *LPAR6* | 4.42E-02 | 2.34E-02 | Non-documented | Non-documented |
| *PEG3* | 4.42E-02 | 1.96E-02 | Non-documented | Non-documented |
| *DCDC2B* | 4.43E-02 | 3.68E-02 | Non-documented | Non-documented |
| *ETNPPL* | 4.44E-02 | 1.17E-03 | Non-documented | Non-documented |
| *CST3* | 4.44E-02 | 1.03E-02 | Non-documented | Non-documented |
| *GTF2A2* | 4.44E-02 | 2.00E-02 | Non-documented | Non-documented |
| *ALG12* | 4.46E-02 | 4.78E-02 | Non-documented | Non-documented |
| *CFB* | 4.49E-02 | 1.08E-02 | Non-documented | Documented |
| *SIX3* | 4.50E-02 | 3.33E-02 | Non-documented | Documented |
| *C20orf141* | 4.51E-02 | 4.13E-02 | Non-documented | Non-documented |
| *TRIM9* | 4.54E-02 | 4.17E-02 | Non-documented | Non-documented |
| *TRIM56* | 4.55E-02 | 6.79E-03 | Non-documented | Non-documented |
| *IL36G* | 4.56E-02 | 3.44E-02 | Non-documented | Non-documented |
| *SLC35D2* | 4.59E-02 | 2.11E-02 | Non-documented | Documented |
| *KRT79* | 4.64E-02 | 1.30E-02 | Non-documented | Non-documented |
| *MAGI3* | 4.66E-02 | 3.76E-02 | Non-documented | Documented |
| *LETMD1* | 4.69E-02 | 2.09E-02 | Non-documented | Non-documented |
| *ASB16* | 4.69E-02 | 7.25E-03 | Non-documented | Non-documented |
| *NRBP1* | 4.71E-02 | 8.69E-03 | Non-documented | Non-documented |
| *FAM83G* | 4.71E-02 | 2.31E-03 | Non-documented | Non-documented |
| *PDDC1* | 4.73E-02 | 4.38E-02 | Non-documented | Non-documented |
| *KCNIP3* | 4.79E-02 | 3.03E-02 | Non-documented | Non-documented |
| *FXYD2* | 4.80E-02 | 2.00E-02 | Non-documented | Non-documented |
| *RPSA* | 4.85E-02 | 3.96E-03 | Non-documented | Non-documented |
| *CXCL14* | 4.85E-02 | 4.61E-03 | Non-documented | Non-documented |
| *PPP1R18* | 4.87E-02 | 2.67E-02 | Non-documented | Documented |
| *SERPINA5* | 4.89E-02 | 1.24E-02 | Non-documented | Non-documented |
| *SIGLEC8* | 4.89E-02 | 4.17E-02 | Non-documented | Non-documented |
| *CYP17A1* | 4.90E-02 | 2.32E-02 | Non-documented | Documented |
| *THOC7* | 4.91E-02 | 3.42E-02 | Non-documented | Non-documented |
| *TGFBR2* | 4.92E-02 | 3.33E-03 | Non-documented | Documented |
| *SORCS3* | 4.93E-02 | 1.61E-02 | Non-documented | Non-documented |
| *NOXRED1* | 4.95E-02 | 1.02E-02 | Non-documented | Non-documented |
| *COX6A2* | 4.98E-02 | 1.86E-03 | Non-documented | Non-documented |
| *NCR3* | 4.99E-02 | 3.92E-02 | Non-documented | Non-documented |
| *YEATS2* | 4.99E-02 | 2.11E-02 | Non-documented | Non-documented |

**Supplemental Table S6. Significantly enriched KEGG pathways by 472 common genes from MAGMA and S-MultiXcan**

| **Gene Set** | **Description** | **Size** | **Ratio** | **P Value** | **FDR** |
| --- | --- | --- | --- | --- | --- |
| hsa05150 | Staphylococcus aureus infection | 56 | 8.38 | 5.09E-08 | 1.66E-05 |
| hsa05330 | Allograft rejection | 38 | 7.86 | 2.35E-05 | 3.03E-03 |
| hsa05320 | Autoimmune thyroid disease | 53 | 6.44 | 2.79E-05 | 3.03E-03 |
| hsa05332 | Graft-versus-host disease | 41 | 7.29 | 3.94E-05 | 3.20E-03 |
| hsa04940 | Type I diabetes mellitus | 43 | 6.95 | 5.43E-05 | 3.20E-03 |
| hsa05416 | Viral myocarditis | 59 | 5.79 | 6.19E-05 | 3.20E-03 |
| hsa05310 | Asthma | 31 | 8.26 | 6.87E-05 | 3.20E-03 |
| hsa04672 | Intestinal immune network for IgA production | 49 | 6.10 | 1.29E-04 | 4.80E-03 |
| hsa04514 | Cell adhesion molecules (CAMs) | 144 | 3.56 | 1.32E-04 | 4.80E-03 |
| hsa05323 | Rheumatoid arthritis | 90 | 4.27 | 2.40E-04 | 7.81E-03 |
| hsa04612 | Antigen processing and presentation | 77 | 4.43 | 4.11E-04 | 1.22E-02 |
| hsa04659 | Th17 cell differentiation | 107 | 3.59 | 8.66E-04 | 2.35E-02 |
| hsa05322 | Systemic lupus erythematosus | 133 | 3.21 | 1.08E-03 | 2.72E-02 |
| hsa05145 | Toxoplasmosis | 113 | 3.40 | 1.28E-03 | 2.93E-02 |
| hsa04658 | Th1 and Th2 cell differentiation | 92 | 3.71 | 1.35E-03 | 2.93E-02 |
| hsa05140 | Leishmaniasis | 74 | 4.04 | 1.65E-03 | 3.36E-02 |
| hsa05166 | Human T-cell leukemia virus 1 infection | 255 | 2.34 | 2.57E-03 | 4.93E-02 |
| hsa04145 | Phagosome | 152 | 2.81 | 2.94E-03 | 5.32E-02 |
| hsa05321 | Inflammatory bowel disease (IBD) | 65 | 3.94 | 3.99E-03 | 6.85E-02 |

**Supplemental Table S7. Significantly enriched Reactome pathways by 472 common genes from MAGMA and S-MultiXcan**

| Gene Set | Description | Size | Ratio | P Value |
| --- | --- | --- | --- | --- |
| R-HSA-174577 | Activation of C3 and C5 | 8 | 19.40 | 2.78E-05 |
| R-HSA-202430 | Translocation of ZAP-70 to Immunological synapse | 19 | 10.21 | 9.47E-05 |
| R-HSA-202427 | Phosphorylation of CD3 and TCR zeta chains | 22 | 8.82 | 2.01E-04 |
| R-HSA-1483191 | Synthesis of PC | 28 | 6.93 | 6.62E-04 |
| R-HSA-72731 | Recycling of eIF2:GDP | 8 | 14.55 | 8.61E-04 |
| R-HSA-194002 | Glucocorticoid biosynthesis | 9 | 12.93 | 1.27E-03 |
| R-HSA-877300 | Interferon gamma signaling | 92 | 3.37 | 2.52E-03 |
| R-HSA-166663 | Initial triggering of complement | 23 | 6.75 | 2.60E-03 |
| R-HSA-389948 | PD-1 signaling | 23 | 6.75 | 2.60E-03 |
| R-HSA-5668914 | Diseases of metabolism | 105 | 2.96 | 5.67E-03 |
| R-HSA-1483206 | Glycerophospholipid biosynthesis | 129 | 2.71 | 6.10E-03 |
| R-HSA-2691230 | Signaling by NOTCH1 HD Domain Mutants in Cancer | 15 | 7.76 | 6.12E-03 |
| R-HSA-2691232 | Constitutive Signaling by NOTCH1 HD Domain Mutants | 15 | 7.76 | 6.12E-03 |
| R-HSA-426048 | Arachidonate production from DAG | 5 | 15.52 | 6.29E-03 |
| R-HSA-70268 | Pyruvate metabolism | 31 | 5.01 | 7.85E-03 |
| R-HSA-5633007 | Regulation of TP53 Activity | 160 | 2.43 | 8.42E-03 |
| R-HSA-202433 | Generation of second messenger molecules | 33 | 4.70 | 9.81E-03 |

**Supplemental Table S8. Drug enrichment analysis based on the GLAD4U database of 472 common genes**

| **Gene Set** | **Size** | **Ratio** | **P Value** | **FDR** |
| --- | --- | --- | --- | --- |
| Mumps vaccines | 46 | 8.02 | 5.73E-06 | 1.05E-02 |
| oxcarbazepine | 16 | 14.41 | 1.67E-05 | 1.54E-02 |
| trichloroethylene | 18 | 12.81 | 3.16E-05 | 1.94E-02 |
| Rubella vaccines | 60 | 6.15 | 4.32E-05 | 1.98E-02 |
| desflurane | 5 | 27.66 | 9.79E-05 | 3.00E-02 |
| lumiracoxib | 5 | 27.66 | 9.79E-05 | 3.00E-02 |
| clavulanate | 7 | 19.76 | 3.32E-04 | 7.16E-02 |
| cosyntropin | 7 | 19.76 | 3.32E-04 | 7.16E-02 |
| Measles vaccines | 44 | 6.29 | 3.51E-04 | 7.16E-02 |
| abatacept | 21 | 8.78 | 9.72E-04 | 1.79E-01 |
| metyrapone | 11 | 12.57 | 1.47E-03 | 2.45E-01 |
| thyroglobulin | 40 | 5.76 | 1.64E-03 | 2.52E-01 |
| human serum albumin | 103 | 3.58 | 1.79E-03 | 2.53E-01 |
| physostigmine | 12 | 11.53 | 1.92E-03 | 2.53E-01 |
| MI-63 | 66 | 4.19 | 3.03E-03 | 3.71E-01 |
| amphetamine | 67 | 4.13 | 3.27E-03 | 3.76E-01 |
| serum albumin | 93 | 3.47 | 4.08E-03 | 4.41E-01 |
| clonazepam | 33 | 5.59 | 5.42E-03 | 5.53E-01 |
| Beta-lactamase inhibitors | 18 | 7.68 | 6.48E-03 | 5.55E-01 |
| xenon | 18 | 7.68 | 6.48E-03 | 5.55E-01 |
| benzyl alcohol | 6 | 15.37 | 6.64E-03 | 5.55E-01 |
| canakinumab | 6 | 15.37 | 6.64E-03 | 5.55E-01 |
| Hepatitis vaccines | 36 | 5.12 | 7.41E-03 | 5.80E-01 |
| beryllium | 19 | 7.28 | 7.57E-03 | 5.80E-01 |
| abacavir | 7 | 13.17 | 9.17E-03 | 6.24E-01 |
| cortisone acetate | 7 | 13.17 | 9.17E-03 | 6.24E-01 |
| plerixafor | 7 | 13.17 | 9.17E-03 | 6.24E-01 |
| Combinations of adrenergics | 21 | 6.59 | 1.01E-02 | 6.61E-01 |
| Tests for allergic diseases | 41 | 4.50 | 1.17E-02 | 7.14E-01 |
| imidapril | 8 | 11.53 | 1.20E-02 | 7.14E-01 |
| vardenafil | 8 | 11.53 | 1.20E-02 | 7.14E-01 |
| lamotrigine | 42 | 4.39 | 1.27E-02 | 7.15E-01 |
| arsenite | 89 | 3.11 | 1.28E-02 | 7.15E-01 |
| ziprasidone | 43 | 4.29 | 1.38E-02 | 7.46E-01 |
| Other antithyroid preparations | 9 | 10.25 | 1.53E-02 | 7.56E-01 |
| carbimazole | 9 | 10.25 | 1.53E-02 | 7.56E-01 |
| milnacipran | 9 | 10.25 | 1.53E-02 | 7.56E-01 |
| lipase | 151 | 2.44 | 1.73E-02 | 7.56E-01 |
| quetiapine | 46 | 4.01 | 1.73E-02 | 7.56E-01 |
| Other immunosuppressants | 123 | 2.62 | 1.77E-02 | 7.56E-01 |
| heroin | 47 | 3.92 | 1.86E-02 | 7.56E-01 |
| Other anticestodals | 246 | 2.06 | 1.87E-02 | 7.56E-01 |
| potassium | 280 | 1.98 | 1.95E-02 | 7.62E-01 |
| ivermectin | 11 | 8.38 | 2.27E-02 | 8.09E-01 |
| Amino acids and derivatives | 222 | 2.08 | 2.33E-02 | 8.09E-01 |
| calcium | 831 | 1.50 | 2.38E-02 | 8.09E-01 |
| Benzimidazole derivatives | 12 | 7.68 | 2.68E-02 | 8.21E-01 |
| phenylacetic acid | 12 | 7.68 | 2.68E-02 | 8.21E-01 |
| d-sorbitol | 53 | 3.48 | 2.77E-02 | 8.21E-01 |
| aloe | 15 | 6.15 | 4.09E-02 | 9.31E-01 |
| betaine | 60 | 3.07 | 4.10E-02 | 9.31E-01 |

**Supplemental Table S9. Significantly enriched GO-terms by 14 identified risk genes for oral cancer using the DisGeNET algorithm in the Metascape tool**

| **GO** | **Description** | **Count** | **%** | **Log10(P)** | **Log10(q)** |
| --- | --- | --- | --- | --- | --- |
| C0024586 | Malignant Carcinoid Syndrome | 3 | 21 | -6.7 | -2.3 |
| C0855095 | Small Lymphocytic Lymphoma | 4 | 29 | -6.5 | -2.3 |
| C0206710 | Basal Cell Neoplasm | 3 | 21 | -5.5 | -1.6 |
| C0030552 | Paresis | 4 | 29 | -5.5 | -1.6 |
| C0751676 | Basal Cell Cancer | 3 | 21 | -5.4 | -1.6 |
| C0007117 | Basal cell carcinoma | 3 | 21 | -4.7 | -1.1 |
| C0237123 | Alcohol or Other Drugs use | 3 | 21 | -4.7 | -1.1 |
| C0085580 | Essential Hypertension | 4 | 29 | -4.3 | -0.71 |
| C0017661 | IGA Glomerulonephritis | 4 | 29 | -4.2 | -0.71 |
| C0553723 | Squamous cell carcinoma of skin | 4 | 29 | -4.2 | -0.71 |
| C0002170 | Alopecia | 4 | 29 | -4.1 | -0.63 |
| C0007114 | Malignant neoplasm of skin | 4 | 29 | -4 | -0.61 |
| C0151786 | Muscle Weakness | 4 | 29 | -4 | -0.56 |
| C0003125 | Anorexia Nervosa | 3 | 21 | -3.9 | -0.54 |
| C0009447 | Common Variable Immunodeficiency | 3 | 21 | -3.9 | -0.54 |
| C0278488 | Carcinoma breast stage IV | 4 | 29 | -3.8 | -0.54 |
| C0003862 | Arthralgia | 3 | 21 | -3.6 | -0.36 |
| C0752347 | Lewy Body Disease | 3 | 21 | -3.6 | -0.35 |
| C0149782 | Squamous cell carcinoma of lung | 4 | 29 | -3.6 | -0.35 |
| C3714636 | Pneumonitis | 4 | 29 | -3.5 | -0.31 |
